# Refining the generation, interpretation, and application of multi-organ, multi-omics biological aging clocks

**DOI:** 10.1101/2025.02.06.25321803

**Authors:** Junhao Wen

## Abstract

Multi-organ biological aging clocks derived from clinical phenotypes and neuroimaging have emerged as valuable tools for studying human aging and disease^1,2,3,4^. Plasma proteomics provides an additional molecular dimension to enrich these clocks^5^. Building on previous work^1,3^, I developed 11 multi-organ proteome-based biological age gaps (ProtBAGs) using 2448 plasma proteins from 43,498 participants in the UK Biobank. I highlighted key methodological and clinical considerations for developing and using ProtBAG, including age bias correction^6^, and investigated the impact of training data sample size, protein organ specificity, and the underlying pathologies of the training data on model generalizability and clinical interpretability. I then integrated the 11 ProtBAGs with our previously developed 9 multi-organ phenotype-based biological age gaps (PhenoBAG^1^) to investigate their genetic underpinnings, causal associations with 525 disease endpoints (DE) from FinnGen and PGC, and their clinical potential in predicting 14 disease categories and mortality. Genetic analyses revealed overlap between ProtBAGs and PhenoBAGs via shared loci, genetic correlations, and colocalization signals. A three-layer causal network linked ProtBAG, PhenoBAG, and DE, exemplified by the pathway of obesity→renal PhenoBAG→renal ProtBAG to holistically understand human aging and disease. Combining features across multiple organs improved predictions for disease categories and mortality. These findings provide a framework for integrating multi-organ and multi-omics biological aging clocks in biomedicine.

## Main

Multi-organ biological aging clocks, derived from *in vivo* medical image (e.g., neuroimaging) and clinical phenotypes, are increasingly being explored in clinical research and computational neuroscience as tools to understand human aging, disease, and mortality^1,2,4,7^. These clocks provide a comprehensive view of biological age, reflecting the functional and structural changes across different organs. While significant advancements have been made in leveraging phenotypic data for such models, there remains a growing interest in incorporating molecular-level data, such as plasma proteomics^5^, epigenetics^8^, and metabolomics^9^, to enrich the landscape of the multi-organ biological aging clocks. Plasma proteomics from different platforms (e.g., Olink^10^ and SomaScan^11^) offers the unique ability to identify and quantify proteins and post-translational modifications with high sensitivity, potentially uncovering new insights into organ-specific aging and its relationship with health and disease^12^.

Despite its promise, deriving proteome-based biological age biomarkers presents several challenges and unresolved questions. One common practice observed in neuroimaging-derived brain age is to correct the age bias in an age prediction model, which may be critical for associations between the biological age gap (BAG) and disease outcomes^13,14,6,15^. That is, brain age tends to be overestimated for younger individuals and underestimated for older individuals, while predictions are most accurate for those whose ages are closer to the mean of the training dataset (**Fig. 1b**). Furthermore, the lack of organ specificity of plasma proteins (analogous to pleiotropy in genetics), where a protein is over-expressed in multiple organ tissues may complicate model development, leading to overfitting and reduced interpretability. Previous studies identified similar overfitting issues and addressed them by employing data-driven feature selection methods to mitigate the problem^5,16^. Furthermore, key factors that influence model performance and generalizability, such as the type of omics data, sample size, and underlying pathology of the training population, as well as the balance between the tightness of model fit and the clinical power of BAG, have not been systematically evaluated. These challenges highlight the need for systematic and reproducible evaluations of proteome-derived BAGs (i.e., ProtBAG)^17^. Addressing these gaps is essential to unlocking the full potential of plasma proteomics in aging research and its clinical applications.

**Figure 1:**
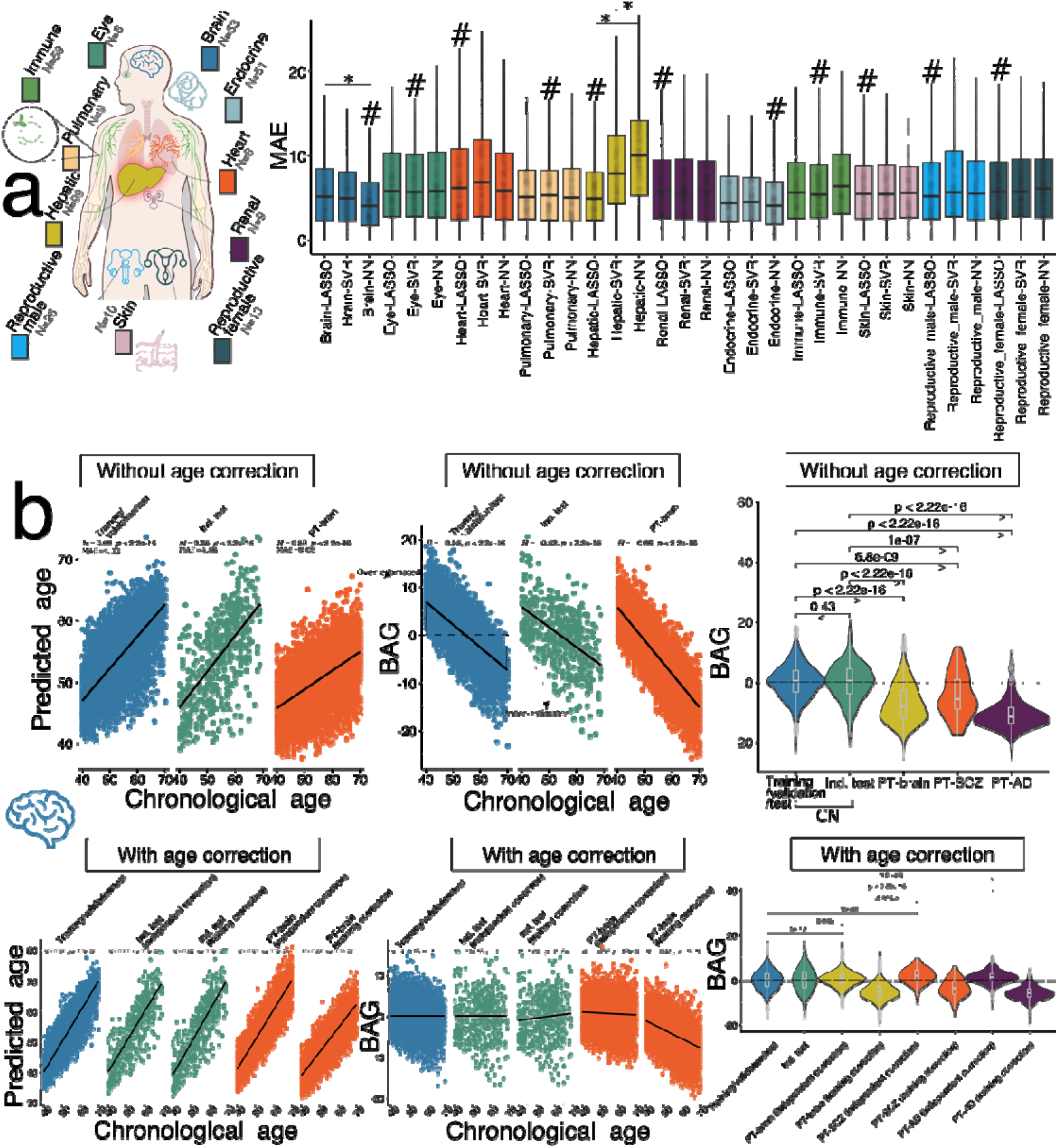
Three AI/ML models to derive the 11 multi-organ ProtBAGs. **a**) Age prediction performance quantified by the mean absolute error (MAE for the independent test data) across 3 AI models and 11 organ systems using Olink plasma proteomics from UKBB. The Human Protein Atlas project determined the organ-specific proteins (i.e., enriched genes for at least four-fold higher mRNA level in the tissue of interest than other tissues; https://www.proteinatlas.org/humanproteome/tissue). The # symbol denotes the model achieving the lowest MAE; the * symbol indicates statistical significance (P-value<0.05) using a two-sample t-test between two models. The dots present the model performance for the 50 repetitions. **b**) When the model was trained in healthy control and applied to other populations, age bias correction should be explicitly applied. Without implementing age bias correction, we showed that group comparisons between the healthy control (CN: training/validation and independent test) group and the patient (PT) groups [PT-brain for participants with brain disorders classified under ICD codes G and F, PT-SCZ for those with schizophrenia (F20 and F21), and PT-AD for those with Alzheimer’s disease (G30)] could lead to biased conclusions. Two approaches for age bias correction were considered: *i*) using the parameters trained in the CN group to correct the independent test and PT populations (i.e., “training correction”), and *ii*) performing independent corrections within the independent test and PT populations (i.e., “independent correction”). Abbreviations: Ind. test: independent test; BAG: biological age gap.

Phenome-wide BAGs (PhenoBAG) and ProtBAG represent two essential aspects of human aging and disease causal pathways, connecting genetics→transcriptomics→proteomics (ProtBAG)→endophenotypes (PhenoBAG)→disease endpoint (DE). Our prior studies^1,3^ have examined the genetic architecture of 12 multi-organ PhenoBAG in 9 organ systems through genome-wide association studies (GWAS) and post-GWAS validations, such as genetic correlation^18^, polygenic risk scores^19^, and causal inference^20^. A comprehensive framework to explore the overlap and distinctions between ProtBAG and PhenoBAG is currently lacking. Addressing this gap requires connecting genetics, ProtBAG, PhenoBAG, and DE. Such an integrative approach is essential for developing a holistic understanding of the causal pathways for potential therapeutic development.

Multi-organ and multi-omics approaches^21,3,1,22,23,7,24,5,25,26,27,28^ are gaining prominence in modeling human aging and disease, driven by the hypothesis that integrating insights across multiple spatial and temporal scales better captures underlying disease-related neurobiological processes, thus enhancing diagnostic and prognostic biomarker discovery. For instance, Zhao et al.^29^ demonstrated improved cognitive prediction by integrating brain and heart MRI features with PRS. Similarly, our prior work on AI/ML-derived brain disease subtypes showed enhanced systemic disease prediction when combining these brain imaging-derived biomarkers with PRS^23,27^. However, the potential of multi-omics and multi-organ BAGs as complementary biomarkers for disease and mortality remains unexplored.

This study used 2448 Olink plasma proteins from 43,498 UK Biobank participants (**UKBB** and **Supplementary eTable 1**) to develop 11 organ-specific ProtBAGs (**Method 1**). I systematically compared the 11 ProtBAGs with 9 PhenoBAGs derived from our previous studies^1,3^ (**Method 2-3**). I evaluated the influence of key methodological components (**Method 4**) on model performance and clinical interpretation using the 11 ProtBAGs. Subsequently, I examined their genetic architecture and causal relationships with 525 DEs from FinnGen^30^ and PGC^31^ (**Method 5**). Finally, I assessed the potential of ProtBAGs, PhenoBAGs, and their PRSs for predicting disease categories and mortality (**Method 6**). All results and pre-trained AI/ML models are publicly disseminated at the MEDICINE portal: https://labs-laboratory.com/medicine/.

## Results

### Age prediction performance of the 11 ProtBAGs derived from three AI/ML models

To rigorously evaluate the performance of biological age prediction models, I partitioned the 5089 healthy control (CN, without any pathologies) participants into the CN training/validation/test (*N*=4589) and independent test (ind. test; *N*=500) datasets. **Extended Data Fig. 1** details this study’s population selection and overall workflow. The CN training/validation/test datasets were used for model selection and nested cross-validation when applicable, while the independent test dataset provided an unbiased assessment of the model for overfitting and generalizability to unseen data. Notably, the cross-validation procedure used in this study differs slightly from two recent proteome-based aging clocks^5,16^ in two key ways: *i*) I explicitly implemented nested cross-validation for hyperparameter tuning of the model, and *ii*) model training was conducted exclusively on 5,089 CN participants to assess generalizability to unseen independent test data.

When fitting the organ-specific proteins (**Method 3**) to the three AI/ML models [i.e., lasso regression, support vector regressor (SVR), and neural network (NN)], I observed marginal variability in model performance, with no single model consistently outperforming the others (**Fig. 1a**). For instance, lasso outperformed NN and SVR for the hepatic ProtBAG (P-value < 2.27×10^−6^), though a two-sample t-test may be permissive^32^ in a complex cross-validation setting. On the other hand, the brain ProtBAG derived from NN obtained a lower MAE than lasso and SVR models (P-value < 2.31×10^−3^). Across different organ systems, the best model performance, before applying the age bias correction^6^, was achieved for the brain ProtBAG via NN (ind. test MAE=4.86; Pearson’s *r*=0.65); the highest MAE was achieved for the hepatic ProtBAG via NN (MAE=10.19; *r*=0.61). Notably, I found instances where MAE and *r* coefficient were not aligned – a lower MAE (reflecting the magnitude of errors) did not always correspond to a higher *r* (indicating the strength and direction of predictions), as these metrics capture different aspects of the model performance and can serve as a potential bias-variance tradeoff and the nonlinear dynamics of proteomics aging^33^. For example, the hepatic ProtBAG predicted using the NN exhibited a high (*r*=0.61) despite a substantial MAE (MAE=10.19), while the eye ProtBAG using the same model achieved a lower MAE (MAE=6.78) but a much weaker correlation (*r*=0.13). **Supplementary eTable 2** presents detailed statistics for the age prediction tasks before and after the age bias correction^6^. **Extended Data Fig. 2** shows the Pearson’s *r* coefficient between predicted and chronological age. **Supplementary eNote 1** presents the detailed tissue-enriched proteins in each organ to train the 11 multi-organ ProtBAGs in the primary results (**Fig. 1a** and **Method 3c**).

### Key considerations for aging clock generation and interpretation

Ikram^34^ and Ferrucci et al^35^ recently discussed the use and misuse of biological aging in biomedicine. This study provided additional critical considerations regarding methodology and clinical interpretation in deriving the 11 multi-organ ProtBAGs (**Method 4**). In this section, I used the brain ProtBAG as a case study to examine the impact of key factors on model performance and clinical interpretability.

#### When training models in healthy controls and applying them to diseased populations, correction for age bias should be performed

The age bias correction was commonly and explicitly practiced in the brain imaging-derived age prediction model^6^, leading to a lower MAE and a higher *r* coefficient (**Fig. 1b**). The first consideration in biological age research is reporting metrics before applying age bias correction^6^. Reporting uncorrected metrics ensures consistency in comparing model performance across studies, preventing potential confusion or misapplication from comparing model performance across studies. Additionally, age bias correction is essential for deriving age-independent aging clocks, which focus on capturing biological aging rather than merely reflecting chronological age. In the analysis comparing brain ProtBAG between the healthy control (CN) and patient (PT-brain) groups with participants for all brain disorders, I found that, without age bias correction^14^, the PT-brain group exhibited a lower brain ProtBAG than the CN group (P-value=2.22 ×10^−16^). However, after applying age bias correction, I observed a reversed and more clinically plausible trend, with the PT group showing a higher brain ProtBAG than the CN group (P-value=0.045). Similar biases were observed when analyzing single disease entities such as schizophrenia and Alzheimer’s disease (AD) (**Fig. 1b**). Additionally, different age bias correction strategies (e.g., directly applying the parameters trained on the CN training/validation/test data vs. independently deriving parameters from the PT data) should be considered when applying the model to the PT data. This is because the parameters trained on the CN training/validation/test data could generalize to CN independent test data, but may not generalize well to the PT data due to potential domain shifts resulting from differences in age, pathology, and other factors (**Fig. 1b**). This was also demonstrated in the work of Oh et al.^5^, who argued that age gaps were calculated separately for each cohort to account for cohort-specific differences using the locally weighted regression (LOWESS residual-based) method. I also compared the neuroimaging-based approach to the LOWESS method used in Oh et al.^5^, assuming proteomics aging clocks’ nolinear trajectory and implicilty addressing this bias during modeling. Both approaches were effective in reducing this bias to some extent (**Supplementary eFigure 1a-d**), although the LOWESS method still indicated a negative mean brain ProtBAG close to 0 (i.e., −0.179 and −0.086 for schizophrenia and AD) when the parameters were learned from the target populations. While including chronological age as a covariate in clinical associations (e.g., mortality) is a standard practice to control for confounding effects related to the variable of interest (i.e., mortality), this does not resolve the issue of age-dependent variance in the aging clock itself.

#### Model overfitting can be mitigated by increasing the sample size of the training dataset, and introducing participants with mixed-pathologies from the UK Biobank

Argentieri et al.^16^ reported an MAE of 2.24 years and an *r* of 0.94 in their holdout test data using UKBB data. My approach differs from Argentieri et al. in several ways. First, I used only 4,589 CN participants for training, whereas Argentieri et al. included a much larger training sample (i.e., 31,808 participants from the general population, including mixed-pathology participants). Although my primary results using tissue-enriched proteins (**Method 3c**) in the 4,589 CN participants did not exhibit clear signs of overfitting or poor generalizability to independent test data (**Fig. 1a**), this was not the case when less organ-specific proteins were included (**Fig. 2b**). Therefore, I investigated whether increasing the sample size could help alleviate the observed overfitting and improve generalizability.

**Figure 2:**
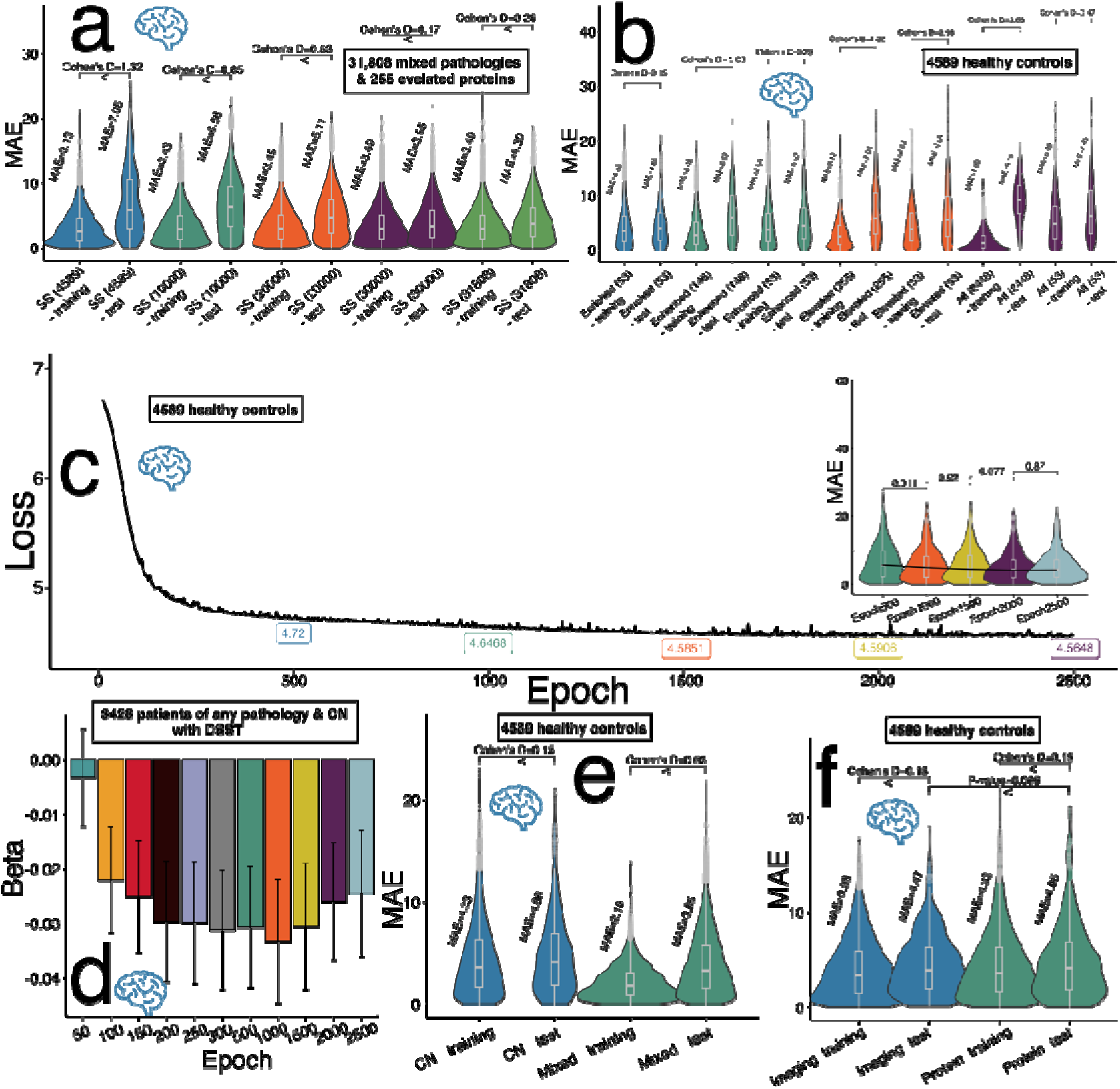
The impact of key components on model performance and generalizability via the brain ProtBAG. **a**) The issue of poor generalizability observed in the 255 brain tissue-elevated proteins was alleviated by increasing the training sample size. I expanded the training population to include a mixed cohort encompassing individuals with ICD-based disease diagnoses^16,5^, rather than restricting it to the CN population – a common practice in the neuroimaging-based brain PhenoBAG. **b**) Different levels of protein organ specificity serve as a means of feature selection, which refers to protein-coding genes with elevated expression levels in a specific tissue or organ, categorized as *i*) tissue-enriched genes, *ii*) tissue-enhanced genes, and *iii*) tissue-elevated genes (https://www.proteinatlas.org/humanproteome/brain/human+brain). Training the model on the 4589 healthy controls revealed that using proteins with lower organ specificity (i.e., incorporating a broader set of proteins as features) led to poor generalizability. **c**) The loss of the validation dataset for training the NN to predict the chronological age at representative epochs until the 2500^th^ epoch. The MAE of the age prediction task at epochs 500, 1000, 1500, 2000, and 2500 is presented. **d**) A more “tightly-fitted” model (Epoch = 2500) did not yield greater statistical power in predicting cognition (i.e., digital symbol substitution test) compared to a “moderately-fitted” model (Epoch = 1000), as indicated by the U-shaped relationship. The *β* coefficient from the linear regression model associating the brain ProtBAG with the cognitive score was evaluated at different epochs. While Epoch 1000 exhibited a trend toward a larger effect size than Epoch 2500, the permutation test yielded a P-value of 0.30 (*N*=10,000 times). **e**) The brain ProtBAG model trained on a mixed population (comprising both CN and PT) demonstrated a lower MAE compared to the model trained exclusively on the CN population (sample size=4589). **f**) The brain PhenoBAG and ProtBAG models achieved comparable performance using brain imaging and plasma protein features, respectively. Of note, training refers to the CN training/validation/test results during the nested CV, and test refers to the independent test dataset.

As increasing the sample size inevitably introduces diverse pathologies, I conducted comparative analyses to assess the impact of training sample size (SS) on model generalizability using the 255 brain tissue-elevated proteins (**Method 3c**) as input features. I selected different SS values (4,589, 10,000, 20,000, 30,000, and 31,808) from the general population (with mixed pathologies) to train the model and evaluated its performance on unseen independent test data. As shown in **Fig. 2a**, a larger SS enhanced the model’s generalizability to independent test data, as indicated by smaller Cohen’s D values (i.e., SS=30,000). I also reproduced this pattern using the complete set of 2,448 proteins, following the approach outlined by Argentieri et al.^16^ (**Supplementary eFigure 1e**). However, incorporating participants with mixed pathologies into the training data also introduces challenges related to clinical interpretation and potential model overfitting, particularly when the sample size is small to moderate, as discussed below.

#### Biologically-driven feature selection based on protein organ specificity alleviates model overfitting in 4589 healthy controls samples

Previous ProtBAG studies have demonstrated that feature selection algorithms can help mitigate model overfitting when applying AI/ML models to unseen test data. For example, Oh et al.^5^ utilized L1 regularization in aggregated lasso models to address overfitting. Similarly, Argentieri et al.^16^ applied the Boruta feature selection algorithm, revealing that the most relevant 204 proteins achieved comparable performance to models trained on the complete set of 2,897 proteins.

Using the 4589 healthy controls as training data, I demonstrated that the generalizability of AI/ML models to independent test data diminished further when using less organ-specific proteins (e.g., tissue-elevated proteins) compared to a smaller subset of highly organ-specific proteins (e.g., tissue-enriched proteins). **Method 3c** details the definition of different levels of organ specificity; more organ-specific proteins resulted in fewer features. In my experiments, I found that restricting the model to brain tissue-enriched proteins (*N*=53) resulted in better model generalizability from the training/validation/test dataset to the independent test dataset (Cohen’s D=0.15) than the other two scenarios. That is, this discrepancy was larger when models included 146 tissue-enhanced proteins (P-value<2.22×10^−16^; Cohen’s D=1.24), 255 tissue-elevated proteins (P-value<2.22×10^−16^; Cohen’s D=1.46), and all the 2448 proteins (P-value<2.22×10^−16^; Cohen’s D=3.52). This pattern persisted even after randomly down-sampling 53 proteins from the brain tissue-enhanced, tissue-elevated, and tissue-nonspecific categories, although the magnitude of Cohen’s D was reduced (**Fig. 2b**).

#### A tightly-fitted model does not provide higher statistical power to predict cognition than a moderately-fitted model

I underscore that the primary objective of developing ProtBAG, or any biological aging clock, is not to achieve a highly tightly-fitted model (e.g., a lower MAE), as this can come at the cost of overfitting and reduced power for cross-domain prediction (**Fig. 2c**). Instead, the focus should be on ensuring that the ProtBAGs demonstrate strong statistical associations with cross-domain clinical variables, such as cognitive scores and mortality.

When assessing the association between the brain ProtBAG and the digital symbol substitution test (DSST) score using a linear regression model, the model at Epoch 2500 (|*β|*=0.024±0.011; P-value=0.03) demonstrated a smaller *β* coefficient compared to the model at Epoch 1000 (|*β|*=0.033±0.011;P-value=0.003); the association at Epoch 1000 was ten times more significant than at Epoch 2500 (with the same sample size), albeit the *β* coefficient did not reach statistical significance based on the permutation test (P-value = 0.30 for 10,000 permutations) (**Fig. 2d**). The observed U-shaped relationship between epochs and the *β* coefficient reinforces my argument that the primary goal of an aging clock model is not solely to optimize model fit (e.g., minimizing MAE or maximizing *r*), but rather to predict cross-domain clinical outcomes, such as cognition and mortality (i.e., a similar U-shaped relationship for age at death in **Supplementary eFigure 2**). Detailed statistics for all 8 cognitive scores and age at death are presented in **Supplementary eTable 3**.

#### The underlying pathology of the training sample is important for clinical interpretation and model performance

In this study, the AI/ML models were trained exclusively on a healthy population, aligning with the approach used in brain neuroimaging-based BAG models, where training on a healthy population establishes a normative reference for brain aging. This framework enables deviations in the brain PhenoBAG to be linked to pathological factors when applied to external populations with disease, facilitating clinical interpretability. In prior proteome-based aging clocks, Oh et al.^5^ adopted this training approach, while Argentieri et al.^16^ trained the model on a cohort of over 30,000 participants with mixed pathologies.

I conducted a comparative experiment with varied training populations to examine how disease diagnosis influences model performance and generalizability. Models trained on the CN population (*N*=4589) showed slightly less overfitting, while mixed-population models achieved lower MAE with moderate overfitting (**Fig. 2e**). This may be due to increased heterogeneity/variability and extreme features tied to pathology, which risk capturing noise over generalizable signals. Another crucial consideration is that a model trained on mixed-pathology populations, with the availability to increase sample size for training power, may limit the clinical interpretability of the resulting aging clocks within the training sample, as well as their applicability to external datasets. This is potentially because different pathologies can lead to distinct protein perturbations, complicating the generalizability and interpretation of the model.

#### Neuroimaging-derived brain PhenoBAG and brain ProtBAG achieved comparable predictive performance

I compared the brain PhenoBAG (ind. test MAE=4.47), generated from 119 MRI-derived brain imaging features^3^, with the brain ProtBAG (ind. test MAE=4.86), constructed using 53 brain tissue-enriched proteins, and found their performance comparable (P-value=0.088) (**Fig. 2f**).

#### Which factor dominates the poor model generalizability to independent test data?

I identified three key factors influencing the model’s generalizability to an independent test dataset and they may guide the clinical interpretability: *i*) sample size, *ii*) the underlying pathology of the training sample, and *iii*) organ specificity. These factors appear to be interdependent, raising the question of which one plays the dominant role in driving overfitting.

I performed additional ablation analyses to identify the key factor influencing the model’s generalizability to unseen data. First, I re-assessed the impact of organ specificity (tissue-enriched, -enhanced, -elevated vs. all 2,448 proteins) using the larger sample size (*N*=31,808), as the one used in Argentieri et al.^16^, which includes both mixed pathologies and healthy participants. I found that a larger sample size reduced overfitting (**Supplementary eFigure 3a**) caused by organ specificity observed in the CN population (**Fig. 2a**) and potential overfitting from mixed pathologies (**Fig. 2e**). Additionally, the observed overfitting related to organ specificity may be partly explained by the relatively lower collinearity among organ-enriched proteins compared to other protein categories (**Supplementary eFigure 3b**). Secondly, I investigated whether organ specificity or the number of features (proteins) contributes to the overfitting issues observed in **Fig. 2a**. To this end, I compared model performance using the 53 brain tissue-enriched proteins and 53 randomly selected non-organ-specific proteins (excluding any proteins classified as tissue-enriched). This comparison was conducted across two cohorts: 4589 CN participants (**Fig. 2a** and **Supplementary eFigure 4a**) and 31,308 participants with mixed pathologies (**Supplementary eFigure 4b**). I found that when training the model on 4589 CN participants, organ specificity significantly contributes to the overfitting phenomenon. However, increasing the sample size to over 30,000 participants with mixed pathologies helps mitigate this overfitting issue.

In summary, I identified three key factors contributing to overfitting, with sample size having the most significant impact. However, training an age prediction model on a large-scale population with mixed pathologies, such as the general UKBB population, also presents certain challenges, as discussed above. I also investigated the influence of sexes (male vs. female) in predicting the brain ProtBAG, as well as a discussion on sex difference in the literature^1,3,36,16^ (**Supplementary eNote 2** and **eFigure 5**). Given that the experiments using organ-specific proteins to derive 11 ProtBAGs in 4589 CN participants did not show prominent overfitting (**Supplementary eTable 2**) and facilitated clinical interpretation, I used the 11 ProtBAGs derived from the 4589 CN participants for downstream genetic and predictive analyses.

#### Protein importance to derive the brain ProtBAG

Using the brain ProtBAG as an example, I identified the most influential proteins contributing to the aging clock through shapley additive explanations (SHAP) analysis^37^, which explains the contribution of each feature (i.e., a brain tissue-enriched protein) to a model’s prediction for a specific individual or data point in the training dataset.

I identified that among the 53 brain tissue-enriched proteins used as input features, only a subset of proteins contributed substantially to the age prediction. The top 9 most influential proteins based on mean absolute SHAP values were: MOG, BCAN, GFAP, PTPRZ1, SEPTIN8, MEPE, CNP, C1QL2, and NPTXR (**Extended Data Fig. 3 a-b**). For example, for the MOG (Myelin Oligodendrocyte Glycoprotein) protein, I observed a high mean absolute SHAP value of 1.8, indicating a strong contribution to the predicted brain age. Moreover, the directionality of the SHAP values suggests that higher MOG levels are associated with an older predicted brain age, potentially reflecting age-related demyelination processes. In contrast, the BCAN (brevican) protein, the second most influential protein, showed an inverse relationship, where higher protein levels were linked to a younger predicted brain age, possibly reflecting its role in maintaining extracellular matrix integrity in the aging brain. Protein-protein interaction network analysis (**Extended Data Fig. 3d**) and subsequent protein-set enrichment analysis^38^ revealed significant involvement of perineuronal nets (**Extended Data Fig. 3e**), which specialized extracellular matrix structures that envelop parvalbumin-expressing GABAergic interneurons in the central nervous system. Finally, I compared the all-cause mortality prediction performance of the Cox model using the brain ProtBAG versus the top 9 most influential individual proteins as predictors, and found that the brain ProtBAG demonstrated superior predictive power (**Extended Data Fig. 3f**). **Supplementary eFigure 6** presents the SHAP anlaysis results for all the 11 ProtBAGs.

### The genetic overlap between ProtBAG and PhenoBAG

I conducted GWAS for the 11 ProtBAGs to identify shared genomic loci and regions with the 9 PhenoBAGs from our previous study (**Method 5a**).

For the 20 GWASs using European ancestry populations, I identified 129 (P-value<5×10^−8^/11) and 308 (P-value<5×10^−8^/9) genomic locus-BAG pairs for the 11 ProtBAGs and 9 PhenoBAGs, respectively. I denoted the genomic loci using their top lead SNPs defined by FUMA (**Supplementary eNote 3**) considering linkage disequilibrium (LD); the genomic loci are presented in **Supplementary eTable 4**. I visually present the shared genomic loci annotated by cytogenetic regions based on the GRCh37 cytoband (**Fig. 3a**). Manhattan and QQ plots, as well as the genomic inflation factor (*λ*) of the 11 ProtBAG and 9 PhenoBAG GWASs, are presented in the MEDICINE portal (e.g., hepatic ProtBAG: https://labs-laboratory.com/medicine/hepatic_protbag). The LDSC intercept (LDSC*_b_*=1.02 [0.99, 1.03]) of the 11 ProtBAG GWASs was close to 1, indicating no severe population stratification observed. **Extended Data Fig. 4** presents the trumpet plots of the effective allele frequency vs. the *β* coefficients of the 11 ProtBAG GWASs.

**Figure 3:**
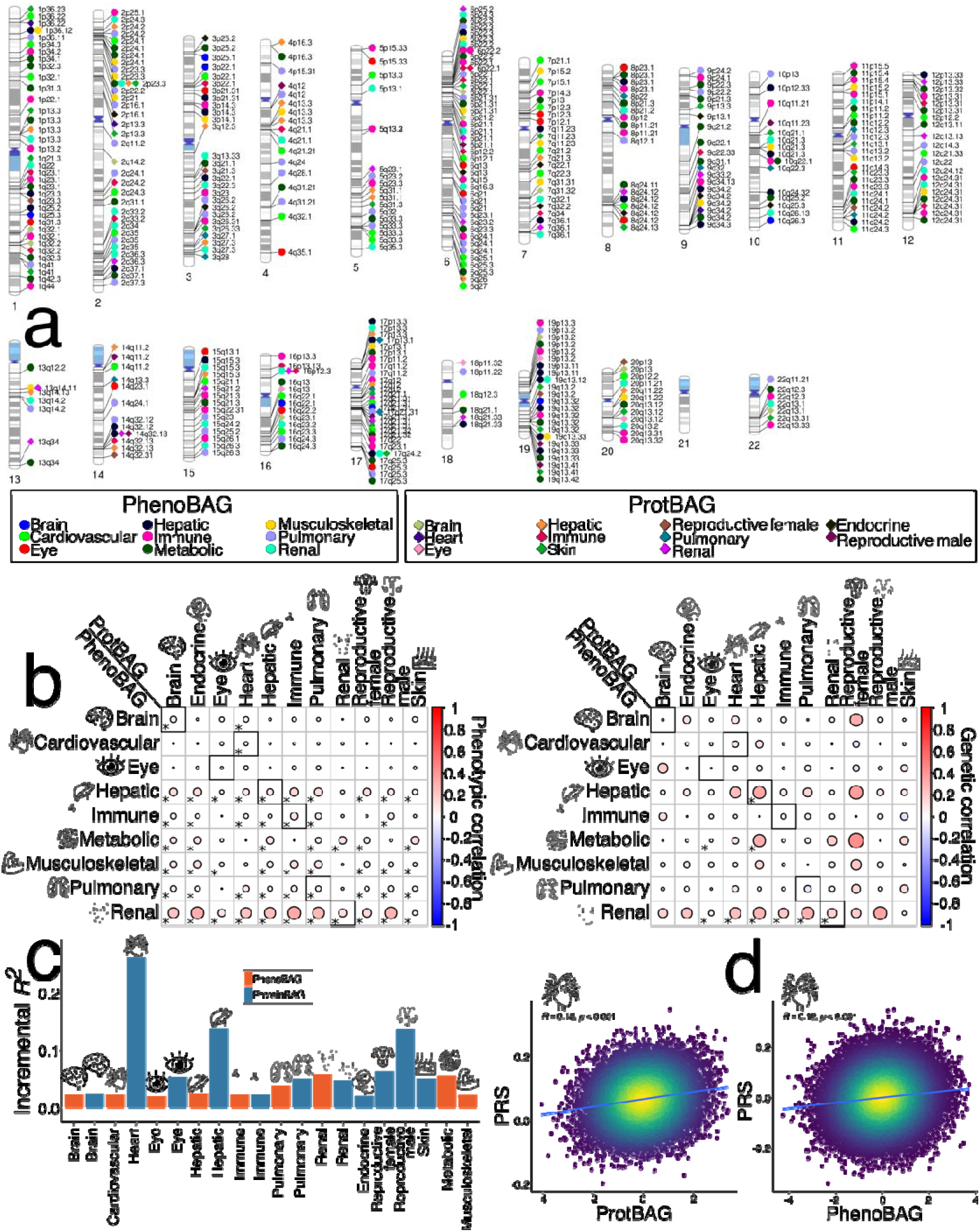
Genetic overlap between PhenoBAG and ProtBAG and the prediction power of their polygenic risk score. **a**) Cytogenetic regions where the genomic region was jointly linked to PhenoBAG and ProtBAG. Bonferroni correction was applied to denote significant genomic loci associated with PhenoBAG (P-value<5×10^−8^/9) and ProtBAG (P-value<5×10^−8^/11). **b**) Phenotypic (*p_c_*) and genetic (*g_c_*) associations were evaluated between each pair of the 9 PhenoBAGs and 11 ProtBAGs. Statistically significant associations after Bonferroni correction (0.05/9/11) are marked with an asterisk (*), and within-organ associations (e.g., between the brain PhenoBAG and ProtBAG) are highlighted with black squares. **c**) The bar plot shows the incremental *R*^2^ (i.e., the *R*^2^ of the alternative model minus that of the null model) for the polygenic risk score (PRS) of each PhenoBAG and ProtBAG. The PRS was calculated using the split2 target GWAS data, with split1 GWAS data serving as the training set for the PRScs model. **d**) The scatter plot shows the relationship between the heart ProtBAG, cardiovascular PhenoBAG, and their corresponding PRS, including the P-value and Pearson’s *r*. Notably, the relationship between PRS and PhenoBAG/ProtBAG is likely not linear (although a linear model was fitted), as PRS inherently accounts for only a small proportion of the variance in the phenotypes of interest. GWAS results are publicly disseminated at https://labs-laboratory.com/medicine/.

I then computed the pairwise genetic correlation (*g_c_*) and phenotypic correlation (*p_c_*) between the 11 ProtBAGs and 9 PhenoBAGs (**Method 5b**). I observed strong associations between the renal PhenoBAG with multiple ProtBAG at both genetic and phenotypic levels, including the immune ProtBAG (*g_c_*=0.21; *p_c_*=0.33) and pulmonary ProtBAG (*g_c_*=0.30; *p_c_*=0.28). Additionally, within-organ associations were not consistently observed; for instance, the eye exhibited neither significant nor phenotypic correlations between the eye PhenoBAG and ProtBAG (**Fig. 3b**). **Supplementary eTable 5a** presents detailed statistics on genetic and phenotypic correlations. **Supplementary eNote 4** and **eTable 5b** present the phenotypic correlation and genetic correlation between the 11 ProtBAGs and the 2448 plasma proteins. A interactive webpage is developed (https://labs-laboratory.com/medicine/protbag_protein_interaction) to browse significant ProtBAG-protein pairs that remain both genetically and phenotypically significant after Bonferroni correction.

### The polygenic risk score of ProtBAG is more predictive than PhenoBAG

I conducted split-sample GWAS to develop the PRS model, using split1 GWAS for training and split2 GWAS for testing, ensuring the two splits had similar age and sex distributions. I evaluated the predictive power of the PRS for the 11 ProtBAG and 9 PhenoBAG by measuring the incremental *R*^2^ gained when predicting the BAG with the PRS as a feature on top of age and sex (**Method 5c**).

All the PRSs demonstrated significant associations with the BAGs (P-value<4.58×10^−81^). The 11 ProtBAG-PRSs showed larger predictive power (incremental *R*^2^ ranging from 2.03% to 26.3%) than the 9 PhenoBAG-PRSs (incremental *R*^2^ ranging from 2.01% to 5.91%) when predicting the BAGs (**Fig. 3c**). For instance, the heart ProtBAG exhibited a higher Pearson’s correlation coefficient with ProtBAG-PRS (*r*=0.18) compared to the heart PhenoBAG and PhenoBAG-PRS (*r*=0.12) (**Fig. 3d**). **Supplementary eTable 6** presents detailed statistics of the PRS analyses.

### The causal relationship between the 11 ProtBAGs, 9 PhenoBAGs, and 525 DEs

I employed two computational genomics methods to explore the causal relationships among the 11 ProtBAGs, 9 PhenoBAGs, and 525 DEs: *i*) Bayesian colocalization (**Method 5d**) and *ii*) Mendelian randomization (**Method 5e**).

Guided by the strong genetic correlation between the hepatic ProtBAG, hepatic PhenoBAG (*g_c_*=0.32), and renal PhenoBAG (*g_c_*=0.29), I investigated the shared causal variants between two traits via Approximate Bayes Factor colocalization^39^ analyses. I demonstrated one genomic locus where the hepatic ProtBAG shared a potential causal variant with both the hepatic PhenoBAG and renal PhenoBAG (**Fig. 4a**). The shared causal variant (rs7212936 at 17p13.3) showed a PP.H4.ABF (Approximate Bayes Factor)=0.99, which examines the posterior probability (PP) to evaluate the hypothesis of a single shared causal variant associated with both traits within this genomic locus. This causal SNV was mapped to the *SERPINF2* gene and has been previously linked in the GWAS Catalog to traits such as serum albumin levels and urate measurements. Additionally, other variants within this locus have been associated with various traits, including blood protein levels and waist-to-hip ratio. These associations, initially identified in the GWAS Catalog, were further validated using the GWAS Atlas platform (**Supplementary eFigure 7**). I mapped this causal SNP to its corresponding gene based on its physical location and evaluated its tissue-specific gene expression profiles using the GTEx^40^ database. Additionally, I analyzed single-cell type enrichment through data curated by the HPA^41^ platform, examined RNA expression across cancer types using the TCGA^42^ database, investigated protein-protein interactions via the STRING^38^ database, and conducted biological pathway enrichment analysis using the Gene Ontology (GO^43^) database. (**Supplementary eFigure 8**).

**Figure 4:**
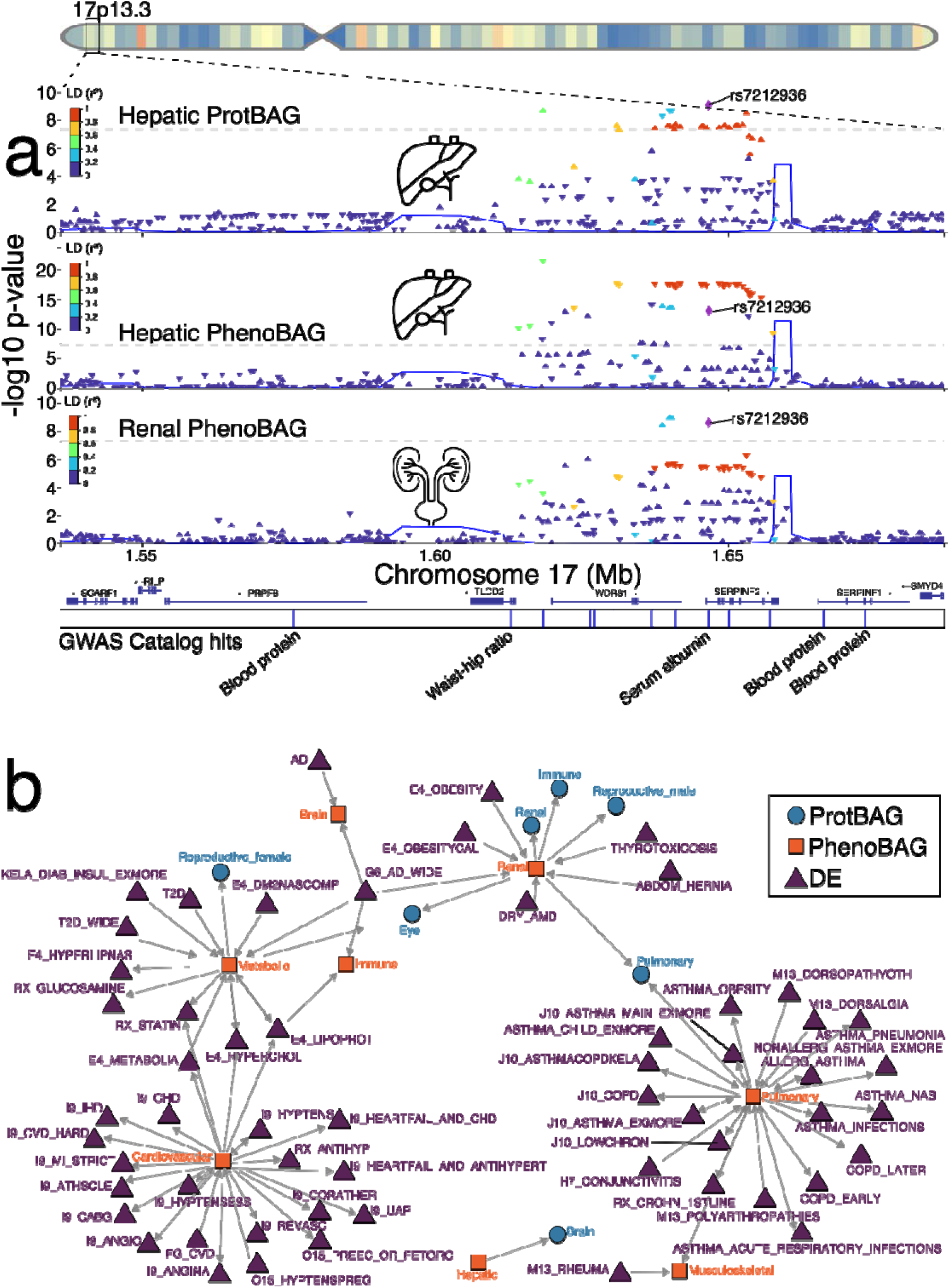
Casual relationship between ProtBAG, PhenoBAG, and disease endpoints. **a**) Genetic colocalization was evidenced at one locus (17p13.3) between the hepatic ProtBAG, hepatic PhenoBAG, and renal PhenoBAG. The signed PP.H4.ABF (>0.8) denotes the posterior probability (PP) of hypothesis H4, which suggests that both traits share the same causal SNP (rs7212936). Representative GWAS hits are annotated based on previous studies available on the NHGRI-EBI GWAS Catalog. **b**) I constructed a three-layer (ProtBAG-PhenoBAG-DE) causal network by employing bi-directional two-sample Mendelian randomization, following a rigorous quality control procedure to select exposure and instrumental variables (number of IVs>7), corrected for multiple comparisons (based on either the number of exposure or outcome variables whichever is larger), and performed sensitivity analyses (e.g., horizontal pleiotropy and removing overlap populations) to scrutinize the robustness of my results. Four causal networks were analyzed: i) *ProtBAG2PhenoBAG*, ii) *PhenoBAG2ProtBAG*, iii) *PhenoBAG2DE*, and iv) *DE2PhenoBAG*. Notably, the ProtBAG GWASs (*N*>40,000) were underpowered compared to the PhenoGWASs (*N*>11,000 for body PhenoBAG), providing no evidence of established causality from ProtBAG to PhenoBAG; Instrumental variables were selected via clumping for these genome-wide significant SNPs considering LD. The arrows indicate the direction of the established causal relationship from the exposure variable to the outcome variable. The interactive network visualization is also available at https://labs-laboratory.com/medicine/protbag_mr. Abbreviations: DE: disease endpoint; LD: linkage disequilibrium. It is crucial to approach the interpretation of these potential causal relationships with caution despite my efforts in conducting multiple sensitivity checks to assess any potential violations of underlying assumptions.

Using bi-directional, two-sample Mendelian randomization analyses, I subsequently established a three-layer causal network that linked ProtBAG, PhenoBAG, and DE (**Fig. 4b**). The *ProtBAG2PhenoBAG* network did not show any significant causal signals (P-value<0.05/10 exposure variables). The *PhenoBAG2ProtBAG* network found 9 causal relationships, including from the renal PhenoBAG to the renal ProtBAG [P-value=4.11×10^−3^<0.05/11; OR (95% CI)=1.18 (1.05, 1.31); number of IVs=46] and from the hepatic PhenoBAG to the brain ProtBAG [P-value=3.44×10^−3^; OR (95% CI)=1.12 (1.04, 1.21); number of IVs=41]. The *PhenoBAG2DE* network found 41 causal relationships, including from the cardiovascular PhenoBAG to hypertension [FinnGen code: I9_HYPTENS; P-value=3.00×10^−7^<0.05/455; OR (95% CI)=1.73 (1.37, 2.17); number of IVs=37] and from the pulmonary PhenoBAG to chronic obstructive pulmonary disease [FinnGen code: J10_COPD; P-value=1.48×10^−19^; OR (95% CI)=1.79 (1.58, 2.03); number of IVs=58]. Finally, for the *DE2PhenoBAG* network, I found 40 causal relationships, including from AD (PGC) to the brain PhenoBAG [P-value=5.00×10^−5^<0.05/179; OR (95% CI)=1.06 (1.03, 1.09); number of IVs=20]. This was further strengthened by the causal link from AD (FinnGen code: G6_AD_WIDE) to the brain PhenoBAG [P-value=3.10×10^−5^; OR (95% CI)=1.10 (1.06, 1.14); number of IVs=8], as well as other PhenoBAG (e.g., immune and renal PhenoBAGs) (**Fig. 4b**). I highlighted a causal pathway connecting three layers: obesity→renal PhenoBAG→renal ProtBAG. Obesity (FinnGen code: E4_OBESITY) demonstrated a positive causal relationship with the renal PhenoBAG [P-value=2.18×10^−8^; OR (95% CI)=1.11 (1.07, 1.15); number of IVs=19], which subsequently exerted a causal effect on the renal ProtBAG [P-value=4.11×10^−3^; OR (95% CI)=1.18 (1.05, 1.31); number of IVs=46], among other ProtBAGs (i.e., eye, immune, male reproductive, and pulmonary) (**Fig. 4b**).

Mendelian randomization relies on stringent assumptions that can sometimes be violated. I conducted comprehensive sensitivity analyses for the significant signals identified to scrutinize this. **Extended Data Fig. 5** provides the results of these analyses for the abovementioned causal pathway, with a detailed discussion available in **Supplementary eNote 5**. Detailed statistics for all five estimators are presented in **Supplementary eTable 7**, and the results of the sensitivity analyses are presented in **Supplementary eDataset 1**.

### The clinical promise of the 11 ProtBAGs, 9 PhenoBAGs, and 20 PRSs

I demonstrated the clinical promise of the 11 ProtBAGs, 11 ProtBAG-PRSs, 9 PhenoBAGs, and 9 PhenoBAG-PRSs in predicting various clinical outcomes through binary classification and survival analysis: *i*) the classification of 14 systemic disease categories and *ii*) the risk of mortality (**Method 6a-b**).

I assessed the prediction ability of support vector machines (SVM) at the individual level to classify the 14 disease categories (**Method 6a**). The highest performance was observed for the respiratory disease category (ICD-codes: J; balanced accuracy (BA)=0.62). The PRS and ProtBAG individually exhibited lower predictive accuracy for disease categories than PhenoBAG. Furthermore, combining all three feature sets failed to outperform the PhenoBAG alone (**Fig. 5a**). Adding age and sex enhanced the classification accuracy (**Supplementary eFigure 9**). Furthermore, I used the circulatory system disease categories (ICD code: I) as an example (**Fig. 5b**) and demonstrated that adding cross-organ features can improve classification performance. The full evaluation metrics of the cross-validated results are presented in **Supplementary eTable 8**.

**Figure 5:**
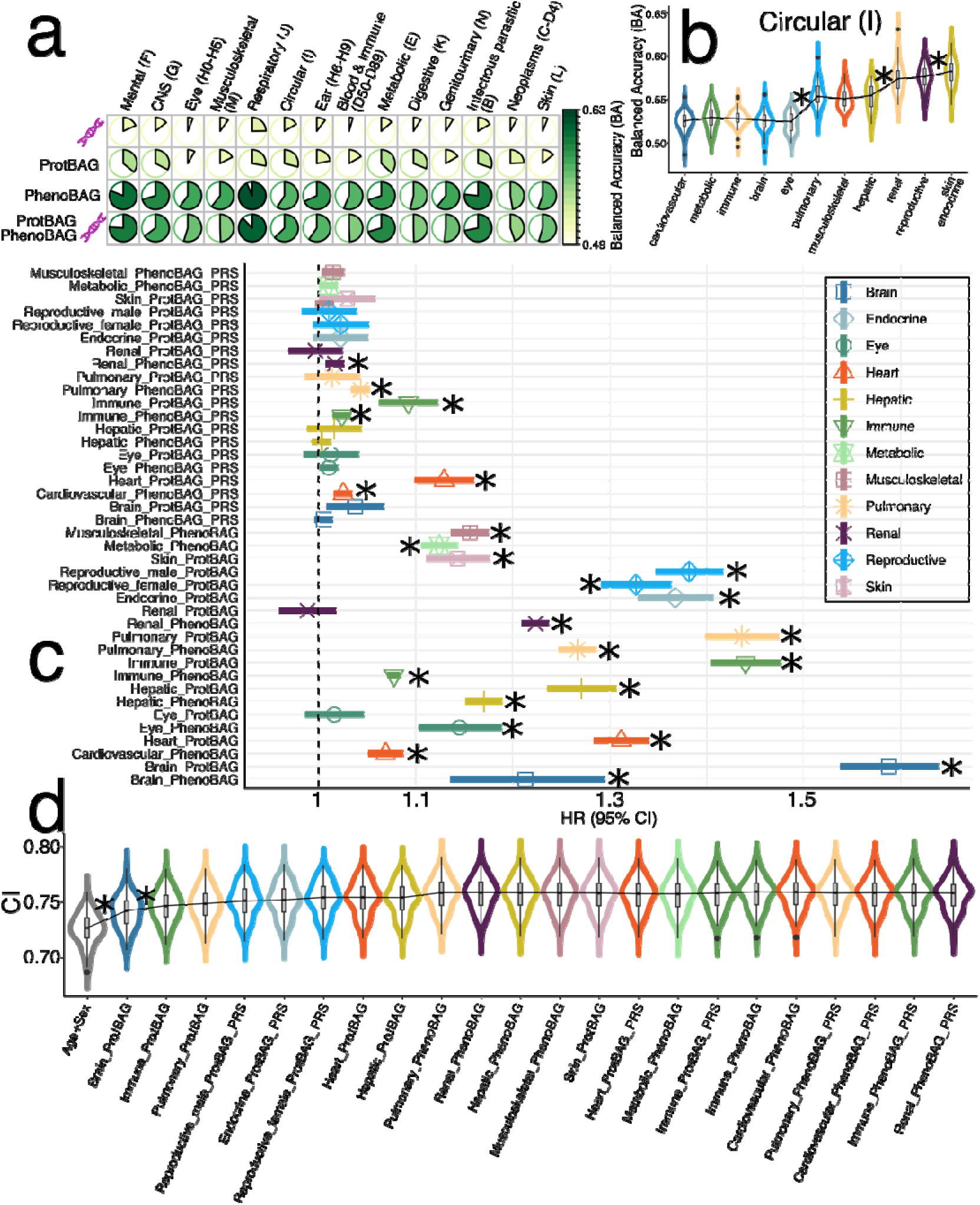
ProtBAG, PhenoBAG, and their PRS predict systemic disease categories and mortality. **a**) The classification balanced accuracy (BA) for 14 ICD-based disease categories was evaluated using PRS, ProtBAG, and PhenoBAG as features within a support vector machine (SVM) framework employing a nested cross-validation (CV) approach (training/validation/test datasets). Balanced accuracy results from the CV are presented, with additional metrics provided in the Supplement. Overall, PhenoBAG demonstrated greater predictive power than other omics data, and simply combining ProtBAG, PhenoBAG, and PRS did not enhance classification performance. The brain and eye PhenoBAG were excluded because merging them with the populations of other features resulted in a very small sample size (*N*<1000). **b**) The cumulative inclusion of organ-specific features enhanced classification performance in predicting circulatory system diseases (ICD code: I). The * symbol indicates statistical significance (<0.05) from a two-sample t-test comparing CV test accuracy between two SVM models; however, a standard t-test is liberal^32^ and should be interpreted cautiously. **c**) ProtBAG, PhenoBAG, and their PRS show significant associations with the risk of mortality. Age and sex were included as covariates in the Cox proportional hazard model. The symbol * indicates significant results that survived the Bonferroni correction (<0.05/9/11). It is important to note that the population sample sizes for ProtBAG and PhenoBAG differ, making their HRs not directly comparable. **d**) The significant ProtBAG, PhenoBAG, and PRS were cumulatively included as features for mortality risk prediction. The * symbol indicates statistical significance (<0.05) from a two-sample t-test comparing results between two Cox models. The populations across omics layers were kept consistent for a fair comparison in panels **a**, **b**, and **d**. However, in panel **c**, I used omics-specific populations since the analysis focused on assessing the predictive power of individual features in a survival analysis. HR: hazard ratio; CI: concordance index.

I also used the 40 BAGs to predict mortality risk using UKBB data (**Method 6b**). The analysis revealed that 24 BAGs or PRSs, including ProtBAGs, PhenoBAGs, and their PRSs, showed significant associations (P-value<0.05/9/11) with mortality. The brain ProtBAG showed the highest mortality risks [HR (95% CI)=1.58 (1.54, 1.63); P-value=7.09×10^−176^], followed by the immune ProtBAG [HR (95% CI)=1.44 (1.40, 1.48); P-value=3.07×10^−181^], and pulmonary ProtBAG [HR (95% CI)=1.43 (1.40, 1.47); P-value=1.98×10^−156^]. Among the 9 PhenoBAGs, the renal PhenoBAG [HR (95% CI)=1.22 (1.21, 1.24); P-value=1.85×10^−252^] and brain PhenoBAG [HR (95% CI)=1.21 (1.14, 1.30); P-value=8.63×10^−9^] showed the highest risks. For the 20 PRSs, the highest mortality risk was achieved with the heart ProtBAG-PRS [HR (95% CI)=1.13 (1.10, 1.16); P-value=1.99×10^−18^ (**Fig. 5c**). Given the population differences among ProtBAGs, PhenoBAGs, and PRSs, comparing hazard ratios (HR) directly is not advisable, as variations in baseline hazard could affect the interpretation. I conducted a cumulative prediction analysis based on the substantial associations identified in the 22 significant BAGs (excluding the brain and eye PhenoBAGs due to their limited sample sizes). This analysis demonstrated that combining these features provided additional predictive power beyond age and sex, achieving an average concordance index of 0.76 ± 0.014 (**Fig. 5d**). The brain and immune ProtBAGs contributed most significantly to this improvement. Comprehensive statistics, including HRs, P-values, and sample sizes, are available in **Supplementary eTable 9**. Finally, I also investigated whether the 11 multi-organ ProtBAGs offer additional statistical power compared to the conventional, non-organ-specific ProtBAG trained on the full UKBB Olink protein set (>2000 proteins and 31808 mixed-pathology participants), as in Argentieri et al^16^. The additional variance explained by the 11 ProtBAGs, beyond age (the largest contributor), sex, and the conventional ProtBAG, was minimal for the 8 cognitive scores (0.16%<incremental *R²*<0.50%; P-value < 10^−10^) and age at death (incremental *R²*=0.64%; P-value < 10^−10^) (**Supplementary eTable 10**). Multi-organ aging clocks are initially motivated by the assumption that different organs may age at distinct rates, offering opportunities to study cross-organ interactions and organ-specific aging dynamics^2^. While their added predictive value over conventional clocks may be modest in some contexts, as shown in my experiments, even small improvements can yield biologically meaningful insights, especially in understanding systemic aging and informing therapeutic strategies^44^.

## Discussion

This study systematically benchmarks the age prediction performance across 11 multi-organ ProtBAGs, revealing insights into the factors influencing model performance and generalizability to unseen data. Inspired by common practices in brain age research^6^, I introduced critical methodological considerations to enhance rigor and clinical interpretability in multi-organ aging research. Subsequently, I comprehensively compared the genetic overlap between the 11 multi-organ ProtBAGs and the 9 PhenoBAGs. By constructing a three-layer causal network, I connected genetics, proteomics, imaging/phenotypic endophenotypes, and disease outcomes, providing an integrative framework for understanding these complex interactions. Finally, I delivered compelling evidence of the clinical potential of the ProtBAGs, PhenoBAGs, and their PRSs in predicting disease categories and mortality, positioning these biomarkers as powerful tools for translational medicine.

### Reproducible and systematic evaluation of ProtBAG generation

I addressed several critical considerations for developing and applying ProtBAG. First, I emphasized the importance of age bias correction, a technique that enhances the clinical relevance of ProtBAG models. In neuroimaging-based brain age research, age bias correction has been explicitly investigated^13,14,6,15^. I provided specific scenarios using proteomics data to emphasize the importance of practicing this in ProtBAG. This is strongly recommended when age prediction models are trained solely on healthy control populations and then applied to other clinical cohorts. Interestingly, the neuroimaging-based age bias correction^6^ for brain age is statistically and mathematically similar to the concept of residual-based aging clocks in epigenetics (i.e., relative aging acceleration in Teschendorff and Horvath^45^), as well as a previous proteomics-based aging clock study^5^. I applied the LOWESS approach from Oh et al.^5^ to correct for age bias and found that it alleviated this bias to some extent (**Supplementary eFigure 1**). In Argentieri et al.^16^, the authors did not apply any methods to correct for age bias but included age as a covariate in their downstream analyses when evaluating associations with clinical outcomes (e.g., disease diagnosis). They concluded that age bias correction had no significant impact on downstream associations. To verify this, I conducted a comparative analysis by linking DSST and age at death to both the brain ProtBAG with and without age bias correction, while including age and sex as covariates for both approaches. I found that age bias correction did not alter the direction of the association; the brain ProtBAG with correction (*β*=-0.026±0.011) yielded a slightly larger *β* coefficient for DSST compared to the uncorrected version (−0.012±0.013; permutation P-value=0.70), while results for age at death were comparable (**Supplementary eTable 11**). I also performed an additional sensitivity analysis by comparing the GWAS signals of the brain ProtBAG obtained from two distinct training datasets: 4589 CN participants (my approach and this from Oh et al.^5^) and 31,808 participants with mixed pathologies (as used in the training approach by Argentieri et al.^16^). The findings demonstrated a significant overlap in genetic correlations between these two training methodologies (*g_c_*=0.97), as well as the two different approaches for age bias correction (*g_c_*=0.98) (**Supplementary eFigure 10**).

My findings also demonstrated the significance of biologically-driven feature selection in alleviating overfitting. Focusing on organ-specific proteins, such as brain tissue-enriched proteins, I achieved better generalizability to unseen data than models using broader, less specific protein sets. Methodologically-driven feature selection algorithms, such as the Boruta algorithm used by Argentieri et al.^16^, offer valuable tools in refining predictive models by identifying a subset of proteins most relevant to biological aging. However, several critical considerations must be addressed. First, complex feature selection should be incorporated within the (nested) cross-validation framework to prevent potential “data leakage,” as highlighted in prior research on AD classification^46^. Second, integrating feature selection within cross-validation can complicate the application of trained models to unseen data, as the features selected may vary across different folds.

Moreover, increasing the training sample size reduced overfitting, emphasizing the importance of large and diverse training populations for enhancing model performance. However, mixed-pathology populations may obscure clinical interpretation, and increased data heterogeneity remains a critical area for further investigation^47^. In addition, I noted that a tighter model fit, reflected in lower MAE, does not necessarily equate to stronger clinical associations, as shown in my analysis of cognitive prediction using the brain ProtBAG. This observation aligns with findings from a previous study that reported similar results using neuroimaging-derived brain age models^48^. Additionally, evaluation metrics such as MAE, Pearson’s *r*, and others may reflect different aspects of the model, and should be considered together for a comprehensive assessment of model performance. A deeper understanding of age prediction models is crucial for accurate interpretation. Practitioners using these aging clocks should have a clear grasp of the model choices to interpret the results correctly, rather than treating them as a ‘black box’.

### The genetic overlap and associations between the 11 ProtBAGs, 9 PhenoBAGs, and 525 DEs

My findings underscore the substantial genetic overlap between ProtBAGs and PhenoBAGs, offering perspectives on the shared and distinct genetic architectures underlying proteomics-driven and phenotypic aging profiles. The identification of hundreds of significant genomic loci linked to these BAGs, along with strong cross-omics and cross-organ genetic correlations, emphasizes the interconnected nature of systemic and organ-specific processes in aging^1,2,5,4^. Notably, the observed associations, such as those between the renal PhenoBAG and immune and pulmonary ProtBAGs, suggest the existence of genetic networks that transcend traditional organ boundaries. Our previous research^3,1^ explored the genetic overlap across organs among the 9 PhenoBAGs. Building on that foundation, the current study expands this scope by integrating 11 ProtBAGs with cross-omics data spanning multiple organs, offering a comprehensive multi-scale framework for understanding human aging and disease.

The superior predictive performance of ProtBAG-PRSs compared to PhenoBAG-PRSs underscores the potential of proteomics-based approaches to advance precision medicine in genetic aging research^10,49,11,50^. The observed differences suggest that ProtBAG may capture distinct genetic signals with stronger biological relevance. This supports the growing recognition of proteomics as a critical component in aging studies, offering deeper insights into novel biomarkers and pathways that may remain elusive through traditional phenotypic analyses. Since proteomics is more closely linked to the underlying genetics and etiology of aging, it offers a valuable molecular layer for studying human aging.

Causal inference analyses provided further insights into the intricate relationships between BAGs and DEs. The colocalization signal of a shared causal variant in the hepatic and renal BAGs exemplifies how integrating proteomic and phenotypic dimensions can uncover biologically relevant loci with translational potential. Similarly, the causal pathway linking obesity, renal PhenoBAGs, and renal ProtBAGs highlights the systemic impact of metabolic factors on organ-specific aging processes. Renal aging clocks can causally link to other organ-specific aging clocks and diseases due to systemic aging processes. The kidneys play a crucial role in metabolic regulation, detoxification, and maintaining homeostasis, meaning their decline can influence cardiovascular, hepatic, and even neurobiological aging or AD. For example, reduced renal function is associated with vascular aging, increased inflammation, and metabolic dysregulation, which can accelerate aging in the heart and brain. Additionally, shared molecular mechanisms, such as oxidative stress, mitochondrial dysfunction, and epigenetic modifications, may drive parallel aging trajectories across multiple organs.

In summary, I demonstrated the value of integrative analyses for BAGs for uncovering the genetic and causal underpinnings of aging across multiple scales. Expanding sample sizes and incorporating diverse ancestries will be critical to enhancing the generalizability of these findings. In addition, exploring the functional consequences of shared loci and causal pathways may provide actionable insights for therapeutic interventions targeting age-related conditions^51^. Finally, it is also crucial to understand the impact of gene-environment interactions on aging clocks. A recent study has linked proteome-based aging clocks to the exposome, including various environmental factors^52^.

### The prediction power of the 11 ProtBAGs, 9 PhenoBAGs, and their PRSs

The observed differences in predictive power for systemic disease categories between PhenoBAG, ProtBAG, and PRS can be attributed to the nature of the data and how they relate to disease categories versus mortality outcomes. For disease category prediction, PhenoBAG, which incorporates phenotypic traits directly linked to specific diseases, is likely more predictive because these traits often represent the clinical manifestation of disease, offering immediate and tangible insights into disease risk. Clinical features such as biomarkers, imaging data, and medical history are more directly associated with disease effects, which makes phenotypic data more informative for predicting disease outcomes. In contrast, PRS, based on genetic predisposition, and ProtBAGs, which rely on proteomic data, may not effectively capture disease-specific features. In particular, the current study focused exclusively on common genetic variants, excluding rare ones typically associated with larger effect sizes^53^. These omics layers provide broader insights into genetic risk and molecular pathways, but their relationships to specific disease categories may be more complex and indirect, making them less predictive for disease classification. Similarly, a recent study showed that multi-omics data and biomarkers can be effectively integrated to outperform PRS in disease predictions^54^.

For mortality prediction, however, ProtBAG and PhenoBAGs show strong predictive power. This is likely because a complex interplay of molecular and clinical factors influences mortality. ProtBAG, which captures proteomic profiles, offers a more direct measure of the molecular processes that underlie aging and disease, such as inflammation, cellular stress, and metabolic dysfunction. These processes are key contributors to mortality, especially in aging populations^16,55^. PhenoBAG, incorporating clinical traits, also reflects the cumulative effects of health deterioration and is strongly correlated with mortality outcomes^13^. PRS, while valuable for predicting genetic susceptibility, may not fully capture the dynamic and multifactorial nature of mortality risk, which involves genetic predisposition, lifestyle factors, physiological markers, and environmental factors^56^.

Interestingly, combining multi-omics BAGs did not significantly improve disease prediction, suggesting that integrating multiple omic layers does not necessarily lead to enhanced performance for disease categories. This may be because disease prediction requires biomarkers specifically relevant to each disease or the broad category, and the multi-omics approach may still lack the necessary disease-specific biomarkers^57^. However, when predicting mortality, the multi-organ BAGs and PRS improved prediction, highlighting the importance of integrating different biological layers across multiple organs. Mortality is a more complex outcome that involves systemic processes across the entire body, making multi-organ and multi-omic approaches more effective. This suggests that combining various molecular layers across organs/omics for comprehensive risk prediction is crucial for capturing the full spectrum of biological processes that influence aging and mortality.

### Outlook

This study investigates several pivotal aspects of biological age research. Future research should expand on this foundation by integrating epigenetic, transcriptomic, and metabolomic^58^ data. This will enrich the causal pathways from genetics to disease outcomes, provide a more comprehensive perspective on human aging and disease^59,60^, and aid in the development of future anti-aging and disease-targeted therapeutics.

## Methods

### Method 1: The MULTI consortium

The MULTI consortium is an ongoing initiative to integrate and consolidate multi-organ data (e.g., brain and heart MRI and eye OCT) with multi-omics data, including imaging, genetics, and proteomics. Building on existing consortia and studies, MULTI aims to curate and harmonize the data to model human aging and disease across the lifespan. This study used individual-level and summary-level multi-omics data from UKBB, FinnGen, and PGC to derive the multi-omics and multi-organ BAGs. **Supplementary eTable 1** details the sample characteristics.

#### UK Biobank

UKBB^61^ is a population-based research initiative comprising around 500,000 individuals from the United Kingdom between 2006 and 2010. Ethical approval for the UKBB study has been secured, and information about the ethics committee can be found here: https://www.ukbiobank.ac.uk/learn-more-about-uk-biobank/governance/ethics-advisory-committee. This study used brain MRI, eye OCT, and clinical phenotypes (e.g., physiological and physical biomarkers) to derive the 9 PhenoBAGs; our previous studies^1,2^ detailed the generation and the phenotypes used for each organ-specific PhenoBAGs. The 11 ProtBAGs were derived from 2448 plasma proteomics data derived from the Olink platform. Imputed genotype data covering the populations of ProtBAG and PhenoBAG were used for all genetic analyses.

#### FinnGen

The FinnGen^30^ study is a large-scale genomics initiative that has analyzed over 500,000 Finnish biobank samples and correlated genetic variation with health data to understand disease mechanisms and predispositions. The project is a collaboration between research organizations and biobanks within Finland and international industry partners. For the benefit of research, FinnGen generously made their GWAS findings accessible to the wider scientific community (https://www.finngen.fi/en/access_results). This research utilized the publicly released GWAS summary statistics (version R9), which became available on May 11, 2022, after harmonization by the consortium. No individual data were used in the current study.

FinnGen published the R9 version of GWAS summary statistics via REGENIE software (v2.2.4)^62^, covering 2272 DEs, including 2269 binary traits and 3 quantitative traits. The GWAS model encompassed covariates like age, sex, the initial 10 genetic principal components, and the genotyping batch. Genotype imputation was referenced on the population-specific SISu v4.0 panel. I included GWAS summary statistics for 521 FinnGen DEs in my analyses.

#### Psychiatric Genomics Consortium

PGC^31^ is an international collaboration of researchers studying the genetic basis of psychiatric disorders. PGC aims to identify and understand the genetic factors contributing to various psychiatric disorders such as schizophrenia, bipolar disorder, major depressive disorder, and others. The GWAS summary statistics were acquired from the PGC website (https://pgc.unc.edu/for-researchers/download-results/), underwent quality checks, and were harmonized to ensure seamless integration into my analysis. No individual data were used from PGC. Each study detailed its specific GWAS models and methodologies, and the consortium consolidated the release of GWAS summary statistics derived from individual studies. In the current study, I included summary data for 4 brain diseases for which allele frequencies were present.

### Method 2: Phenotype analyses to derive the 9 PhenoBAGs

We derived the 9 PhenoBAGs in our previous study^1,2^, and we also present the final included phenotypes for the 9 human organs in **Supplementary eTable 1**. In summary, we selected brain MRI, physical and physiological measures indicative of key organ systems’ function, structure, or general health, including the brain (e.g., brain volume), cardiovascular (e.g., pulse rate), pulmonary (e.g., peak expiratory flow), musculoskeletal (e.g., BMI), immune (e.g., leukocytes), renal (e.g., glomerular filtration), hepatic (e.g., albumin), and metabolic systems (e.g., lipid). Data were processed to ensure reliability: averages were calculated for bilateral measures (e.g., handgrip strength), repeated tests (e.g., blood pressure), or the best performance from multiple attempts (e.g., lung function via spirometry). The eye PhenoBAG was subsequently derived in our follow-up study using eye OCT data^1^.

To derive the 9 PhenoBAGs, we used a linear support vector regressor (SVR) and fit the organ-specific phenotypes as features with a 20-fold cross-validation procedure. Optimization of the SVR’s hyperparameters (box constraint, kernel function, and ε) did not substantially improve performance. Critically, the SVR models were trained exclusively on healthy individuals, defined as those without self-reported or healthcare-documented lifetime chronic medical conditions. This approach supports the clinical interpretation of the trained models when applied to disease groups, with deviations in these PhenoBAGs presumed to reflect specific pathological factors.

### Method 3: Proteomics analyses to derive the 11 ProtBAGs

#### (a) Additional quality checks

I downloaded the original data (Category code: 1838), which were analyzed and made available to the community by the UKB-PPP^63^. The initial quality check was detailed in the original work^64^; I performed additional quality check steps as below. I focused on the first instance of the proteomics data (“instance”=0). Subsequently, I merged the Olink files containing coding information, batch numbers, assay details, and limit of detection (LOD) data (Category ID: 1839) to match the ID of the proteomics dataset. I eliminated Normalized Protein eXpression (NPX) values below the protein-specific LOD. Furthermore, I restricted my analysis to proteins with sample sizes exceeding 10,000. This resulted in 2448 proteins in 43,498 participants.

#### (b) Missing protein NPX imputation

I observed a substantial missing rate for the 2448 proteins (1229 proteins with > 10% missing values), which made it challenging to employ downstream AI/ML models for age prediction because many of these models do not directly handle missing features. I used the AutoComplete^65^ deep learning algorithm to impute the missing proteins to overcome this. In the original paper, the authors have thoroughly evaluated the impact of the missing rate on the imputation accuracy. Here, I followed the same approach proposed in the paper, assessed the impact of the probability of an individual’s being masked during training, and found that this impact is minimal for the imputation accuracy (**Supplementary eFigure 11**). I observed a mean *R*^2^ value of 0.45 between the imputed values and the ground truth for the 2448 proteins, showing improved model performance compared to the original study on cardiometabolic and psychiatric phenotypes (0.14<*R*^2^<0.30).

#### (c) Organ-specific profiles of the 2448 plasma proteins

I used the Human Protein Atlas (HPA) project (https://www.proteinatlas.org/humanproteome/tissue) to profile the over-expression of a specific protein at both RNA-seq and protein levels. The HPA highlights the expression profiles of genes in human tissues, primarily at the mRNA level, and complements this with protein-level localization using antibody-based methods such as immunohistochemistry and immunofluorescence. Protein expression data from 44 normal human tissue types were obtained through antibody-based protein profiling using conventional and multiplex immunohistochemistry. Accompanying the resource are annotated protein expression levels and all images of immunohistochemically stained tissues. Protein data encompass 15,302 genes (76%) with available antibodies. Additionally, mRNA expression data were generated through RNA sequencing (RNA-seq) of 40 different normal tissue types. In my primary analyses to derive the 11 organ-specific ProtBAGs, I considered defining whether a protein is over-expressed in a particular organ/tissue using the following criterion: tissue-enriched genes are characterized by mRNA expression levels at least four times higher in the tissue or organ of interest compared to all other tissues. This approach aligns with the definition employed by Oh et al.^5^, which relied solely on data from the Genotype-Tissue Expression (GTEx) project. In contrast, the Human Protein Atlas (HPA) integrates resources from multiple consortia, extending beyond GTEx data. A more recent study^66^ employed a protein-centric, mass spectrometry–based strategy to systematically profile tissue-enriched proteins across 18 organs and directly connect them to their abundance and behavior in human plasma.

An important yet unexplored question is the relative lack of organ specificity in plasma proteins circulating throughout the human body compared to clinical phenotypes, such as brain MRI features. Many proteins are frequently over-expressed across multiple tissues or organs, akin to the pleiotropic effects observed in genetics. This observation is biologically plausible, as proteomics is more closely linked to underlying genetic mechanisms, whereas clinical phenotypes are more directly associated with disease endpoints. Additionally, Argentieri et al.^16^ demonstrated through feature selection that 204 out of 2,897 proteins from the UKBB Olink platform could accurately predict chronological age. However, the impact of protein organ-specificity definitions on model overfitting remains an unresolved question. In this study, I explored this issue by systematically relaxing the organ-specific profiles of proteins under three distinct scenarios. Using the brain ProtBAG as an example, I assessed how varying the number of proteins included in the training of AI/ML models influences performance and overfitting phenomena.

- **Tissue-enriched genes/proteins**: At least four-fold higher mRNA level in the tissue of interest than in other tissues (*N*=53 proteins).
- **Tissue-enhanced genes/proteins**: At least four-fold higher mRNA level in the tissue of interest compared to the average level in all other tissues (*N*=146 proteins).
- **Tissue-elevated genes/proteins**: tissue-enriched genes (including group-enriched genes) and tissue-enhanced genes (*N*=255 proteins).

#### (d) Three AI/ML models

I systematically benchmarked age prediction performance using 4 AI/ML models on multi-modal brain MRI features in our previous study^3^. Using the same methodology, I assessed the performance of models in deriving the 11 ProtBAGs using two linear approaches (lasso regression and SVR) and one non-linear method (neural networks). For the linear models, hyperparameter selection (e.g., the *C* parameter for SVR) was conducted through nested, repeated hold-out cross-validation^17^ with 50 repetitions (80% training/validation and 20% testing) for the outer loop and 10-fold cross-validation for the inner loop for hyperparameter selection. Nested cross-validation was not applied to the neural network due to the impracticality of exhaustively exploring hyperparameter combinations.

#### (e) Population selections

To rigorously train the AI/ML models, I split the CN data (*N*=5089) into the following datasets:

- **CN independent test dataset**: 500 participants were randomly drawn from the CN population;
- **CN training/validation dataset**: 80% of the remaining 4589 CN were used for the inner loop 10-fold CV for hyperparameter selection;
- **CN cross-validation test dataset**: 20% of the remaining 4589 CN were used for the outer loop 50 repetitions;
- **PT dataset**: 38,409 participants that have at least one ICD-10-based diagnosis. Model evaluation metrics included mean absolute error (MAE) and Pearson’s *r*.

Importantly, consistent with our prior studies, only healthy control participants were included in the training/validation dataset, while individuals with any disease diagnosis were reserved for the independent test dataset. **Supplementary eFigure 12** outlines my modeling considerations and the nested cross-validation procedure; **Supplementary eTable 1** provides the basic demographic information, including age and sex.

### Method 4: Influence of key components in deriving the brain ProtBAG

I systematically evaluated key factors influencing model performance using the brain ProtBAG as a case study. These factors included *i*) the choice of AI/ML models (i.e., SVR, lasso, and neural networks), *ii*) the impact of age bias correction on downstream clinical applications, such as group differences between CN and PT groups, *iii*) the effect of protein organ specificity on model overfitting, comparing enriched, enhanced, and elevated gene categories, *iv*) the influence of model fitting tightness on cross-domain prediction, particularly associations with cognitive outcomes at various epochs (i.e., 500, 1000, 1500, 2000, and 2500 epochs), and *v*) the impact of feature type on model performance, comparing brain imaging-derived features with brain over-expressed plasma proteins. These analyses provide practical guidance for using plasma proteins to develop ProtBAGs while enhancing clinical interpretability and methodological rigor. I also evaluated the feature importance of the 11 ProtBAGs using the SHAP method on the training data.

### Method 5: Genetic analyses

I used the imputed genotype data for all genetic analyses. My quality check pipeline focused on European ancestry in UKBB (6,477,810 SNPs passing quality checks), and the quality-checked genetic data were merged with respective organ-specific populations for GWAS. I summarize my genetic quality check steps. First, I skipped the step for family relationship inference^67^ because the linear mixed model via fastGWA^68^ inherently addresses population stratification, encompassing additional cryptic population stratification factors. I then removed duplicated variants from all 22 autosomal chromosomes. Individuals whose genetically identified sex did not match their self-acknowledged sex were removed. Other excluding criteria were: *i*) individuals with more than 3% of missing genotypes; *ii*) variants with minor allele frequency (MAF; dosage mode) of less than 1%; *iii*) variants with larger than 3% missing genotyping rate; *iv*) variants that failed the Hardy-Weinberg test at 1×10^−10^. To further adjust for population stratification,^69^ I derived the first 40 genetic principle components using the FlashPCA software^70^. Details of the genetic quality check protocol are described elsewhere^71,3,1,72,73^.

#### (a) GWAS

I applied a linear mixed model regression to the European ancestry populations using fastGWA^68^ implemented in GCTA^74^.

##### PhenoBAG GWAS

In our initial investigation, I conducted GWAS for the 9 PhenoBAGs using a linear model in PLINK, with fastGWA employed as a sensitivity analysis. For consistency with the ProtBAG GWASs in this study, I used the fastGWA summary statistics for the 9 PhenoBAGs in all post-GWAS analyses. The fastGWA GWAS accounted for key confounders, including age, dataset status (training/validation/test or independent test), age-squared, sex, interactions of age with sex, and the first 40 genetic principal components. For the brain BAG GWAS specifically, additional covariates for total intracranial volume and brain position in the scanner were included. A genome-wide significance threshold (5 × 10 /9), was applied.

##### ProtBAG GWAS

I used fastGWA to perform the 11 ProtBAGs, adjusting age, dataset status (training/validation/test or independent test), age-squared, sex, interactions of age with sex, systolic/diastolic blood pressure, BMI, waist circumstance, standing height, weight, and the first 40 genetic principal components. I applied a genome-wide significance threshold (5 × 10 /11) to annotate the significant independent genomic loci.

##### Annotation of genomic loci

For all GWASs, genomic loci were annotated using FUMA^75^. For genomic loci annotation, FUMA initially identified lead SNPs (correlation *r*^2^ ≤ 0.1, distance < 250 kilobases) and assigned them to non-overlapping genomic loci. The lead SNP with the lowest P-value (i.e., the top lead SNP) represented the genomic locus. Further details on the definitions of top lead SNP, lead SNP, independent significant SNP, and candidate SNP can be found in **Supplementary eNote 3**. For visualization purposes in **Fig. 3**, I have mapped the top lead SNP of each locus to the cytogenetic regions based on the GRCh37 cytoband.

#### (b) Genetic correlation

I estimated the genetic correlation (*g_c_*) between each PhenoBAG-ProtBAG pair using the LDSC software. I employed precomputed LD scores from the 1000 Genomes of European ancestry, maintaining default settings for other parameters in LDSC. It’s worth noting that LDSC corrects for sample overlap, ensuring an unbiased genetic correlation estimate^76^. I also computed the pairwise Pearson’s *r* correlation coefficient to understand whether the genetic correlation largely mirrors the phenotypic correlation (*p_c_*). Statistical significance was determined using Bonferroni correction (0.05/9/11).

#### (c) PRS calculation

PRS was computed using split-sample sensitivity GWASs (split1 and split2) for the PhenoBAG and ProtBAG GWASs. The PRS weights were established using split1/discovery GWAS data as the base/training set, while the split2/replication GWAS summary statistics served as the target/testing data. Both base and target data underwent rigorous quality control procedures involving several steps: *i*) excluding duplicated and ambiguous SNPs in the base data; *ii*) excluding high heterozygosity samples in the target data; and *v*) eliminating duplicated, mismatching, and ambiguous SNPs in the target data.

After completing the QC procedures, PRS for the split2 group was calculated using the PRS-CS^77^ method. PRS-CS applies a continuous shrinkage prior, which adjusts the SNP effect sizes based on their LD structure. SNPs with weaker evidence are “shrunk” toward zero, while those with stronger evidence retain larger effect sizes. This avoids overfitting and improves prediction performance. No clumping was performed because the method takes LD into account. The shrinkage parameter was not set, and the algorithm learned it via a fully Bayesian approach.

#### (d) Bayesian colocalization

I used the R package (*coloc*) to investigate the genetic colocalization signals between two traits (i.e., hepatic ProtBAG vs. hepatic PhenoBAG, and hepatic ProtBAG vs. renal PhenoBAG) at each genomic locus. I employed the Fully Bayesian colocalization analysis using Bayes Factors (*coloc.abf*). This method examines the posterior probability (PP.H4.ABF: Approximate Bayes Factor) to evaluate hypothesis *H4*, which suggests the presence of a single shared causal variant associated with both traits within a specific genomic locus. To determine the significance of the *H4* hypothesis, I set a threshold of PP.H4.ABF>0.8^39^. All other parameters (e.g., the prior probability of p_12_) were set as default. For each pair of traits, the genomic locus (*N*>100 SNPs) was defined by default from FUMA for one trait, and then the *coloc* package extracted and harmonized the GWAS summary statistics within this locus for the other trait.

#### (e) Two-sample bidirectional Mendelian randomization

I constructed a multi-layer causal network linking ProtBAG, PhenoBAG, and DE using a bi-directional Mendelian randomization approach. In total, 4 bi-directional causal networks were established: i) *ProtBAG2PhenoBAG*, ii) *PhenoBAG2ProtBAG*, iii) *PhenoBAG2DE*, and iv) *DE2PhenoBAG*. These networks used summary statistics from our ProtBAG and PhenoBAG GWAS in the UKBB, the FinnGen^30^, and the PGC^31^ study for the 525 DEs. For example, the *ProtBAG2PhenoBAG* causal network employed the 11 ProtBAG as exposure variables and the 9 PhenoBAGs as outcome variables. The systematic quality-checking procedures to ensure unbiased exposure/outcome variable and instrumental variable (IVs) selection are detailed below.

I used a two-sample Mendelian randomization approach implemented in the *TwoSampleMR* package^78^ to infer the causal relationships within these networks. I employed five distinct Mendelian randomization methods, presenting the results of the inverse variance weighted (IVW) method in the main text and the outcomes of the other four methods (Egger, weighted median, simple mode, and weighted mode estimators) in the supplement. The STROBE-MR Statement^79^ guided my analyses to increase transparency and reproducibility, encompassing the selection of exposure and outcome variables, reporting statistics, and implementing sensitivity checks to identify potential violations of underlying assumptions. First, I performed an unbiased quality check on the GWAS summary statistics. Notably, the absence of population overlapping bias^80^ was confirmed, given that FinnGen and UKBB participants largely represent populations of European ancestry without explicit overlap. PGC GWAS summary data were ensured to exclude UKBB participants. For the *ProtBAG2PhenoBAG* and *PhenoBAG2ProtBAG* networks from UKBB, I reran the ProtBAG GWAS and ensured no overlapping populations with PhenoBAG. Furthermore, all consortia’s GWAS summary statistics were based on or lifted to GRCh37. Subsequently, I selected the effective exposure variables by assessing the statistical power of the exposure GWAS summary statistics in terms of instrumental variables (IVs), ensuring that the number of IVs exceeded 7 before harmonizing the data. Crucially, the function “*clump_data*” was applied to the exposure GWAS data, considering LD. The function “*harmonise_data*” was then used to harmonize the GWAS summary statistics of the exposure and outcome variables. Bonferroni correction was applied to all tested traits based on the number of effective ProtBAGs, PhenoBAGs, or DEs, whichever was larger.

Finally, I conducted multiple sensitivity analyses. First, I conducted a heterogeneity test to scrutinize potential violations in the IV’s assumptions. To assess horizontal pleiotropy, which indicates the IV’s exclusivity assumption^81^, I utilized a funnel plot, single-SNP Mendelian randomization methods, and the Egger estimator. Furthermore, I performed a leave-one-out analysis, systematically excluding one instrument (SNP/IV) at a time, to gauge the sensitivity of the results to individual SNPs.

### Method 6: Prediction analyses for 14 systemic disease categories and the risk of mortality

I investigated the clinical promise of the 11 ProtBAGs, 9 PhenoBAGs, and their PRSs in two sets of prediction analyses: i) classification tasks for predicting 14 systemic disease categories based on the ICD-10 code (**Supplementary eTable 8**) and *ii*) survival analysis for the risk of all-cause mortality.

#### (a) Support vector machines to classify patients of disease categories vs. controls

I applied SVM with ProtBAG, PhenoBAG, and their PRS, implementing a nested cross-validation procedure^17^ to optimize the hyperparameter *C* and predict individual-level outcomes. Unlike previous studies^57^, I did not set aside an independent test dataset due to the relatively small sample size of the control population without any disease diagnoses (*N*=1651); patients for each disease category were defined by the ICD-10 code (Field-ID: 41270). Brain and eye PhenoBAGs were excluded from the analysis due to insufficient sample sizes after integrating all features. **Figure 5a-b** reports the balanced accuracy (BA) obtained from the nested test data. The nested cross-validation procedure involved an outer loop repeated 50 times, with 80% of the data randomly allocated for training/validation and 20% for testing. Within the inner loop, the training/validation data underwent a 10-fold split for model optimization. **Supplementary eTable 8** provides detailed metrics, including balanced accuracy, sensitivity, specificity, negative predictive value, positive predictive value, and sample sizes for the training/validation/test datasets.

#### (b) Survival analysis for mortality risk

I employed a Cox proportional hazard model while adjusting for covariates(i.e., age and sex) to test the associations of the 11 ProtBAGs, 9 PhenoBAGs, and their PRS with all-cause mortality. The covariates were included as additional right-side variables in the model. The hazard ratio (HR), exp(β*_R_*), was calculated and reported as the effect size measure that indicates the influence of each biomarker on the risk of mortality. To train the model, the “time” variable was determined by calculating the difference between the date of death (Field ID: 40000) for cases (or the censoring date for non-cases) and the date attending the assessment center (Field ID: 53). Participants who passed away after enrolling in the study were classified as cases.

## Data Availability

The GWAS summary statistics and pre-trained AI models from this study are publicly accessible via the MEDICINE Knowledge Portal (https://labs-laboratory.com/medicine/) and Synapse (https://www.synapse.org/Synapse:syn64923248/wiki/630992). My study used data generated by the human protein atlas (HPA: https://www.proteinatlas.org). GWAS summary data for the DEs were downloaded from the official websites of FinnGen (R9: https://www.finngen.fi/en/access_results) and PGC (https://pgc.unc.edu/for-researchers/download-results/). Individual data from UKBB can be requested with proper registration at https://www.ukbiobank.ac.uk/. Certain sensitive data (e.g., allele frequency information) supporting the findings are also available from the author upon request. The MR sensitivity analysis results (**Supplementary eDataset 1**), the 5 main figures, and the scripts to reproduce the methodological evaluations are publicly available on Synapse (https://zenodo.org/records/15211957^82^).

## Code Availability

The software and resources used in this study are all publicly available:

- MLNI^83^ (v0.1.2): https://github.com/anbai106/mlni, ProtBAG generation and classification for disease categories;
- AutoComplete (git version de78189): https://github.com/sriramlab/AutoComplete, Proteomics imputation;
- FUMA (v1.5.0): https://fuma.ctglab.nl/, Gene mapping, genomic locus annotation;
- GCTA (v1.94.1): https://yanglab.westlake.edu.cn/software/gcta/#Overview, fastGWA;
- LDSC (git version aa33296): https://github.com/bulik/ldsc, genetic correlation
- TwoSampleMR (v0.5.6): https://mrcieu.github.io/TwoSampleMR/index.html, Mendelian randomization;
- PRScs (release date: Aug 10, 2023): https://github.com/getian107/PRScs, PRS calculation;
- Lifelines (v0.27.8): https://lifelines.readthedocs.io/en/latest/, Survival analysis;
- coloc (v5): https://github.com/chr1swallace/coloc; Bayesian colocalization.

The pre-trained AI models are accessible to the public through the MEDICINE portal (https://labs-laboratory.com/medicine/). Users can directly apply these models to external datasets, provided the data has been preprocessed (e.g., missing value imputation) using my MLNI package (https://anbai106.github.io/mlni/; Version: 0.1.2).

## Acknowledgments

The MULTI consortium (J.W; UK Biobank Application Number: 647044) aims to integrate multi-organ imaging and multi-omics data to advance our understanding of human aging and disease mechanisms. This research has been conducted using data from UK Biobank, a major biomedical database, under Application Number 647044, 35148, and 60698. I want to express our sincere gratitude to the UK Biobank team for their invaluable contribution to advancing clinical research in our field (https://www.ukbiobank.ac.uk/). I also acknowledge the UKB-PPP consortium (https://registry.opendata.aws/ukbppp/; Category code: 1838) to share the returned data with the community. I want to acknowledge the participants and investigators of the FinnGen study and the PGC consortium, and I thank FinnGen (https://www.finngen.fi/en) and PGC (https://pgc.unc.edu/) for their generosity in sharing the GWAS summary statistics with the scientific community. I would like to express my gratitude to Dr. Christos Davatzikos, Dr. Andrew Zalesky, and Dr. Ye Ella Tian for their invaluable support and collaboration on my research. Their contributions were crucial in helping me write this manuscript, which originally began as a proposal for a Commentary on the aging clocks in the literature, but ultimately evolved into a full set of analyses. I acknowledge the leadership of the Brain Imaging Genetics (BIG) workgroup, led by Dr. Tavia Evans, Dr. Natalia Vilor-Tejedor, and Dr. Junhao Wen, within the International Society to Advance Alzheimer’s Research and Treatment (ISTAART) community, for advocating brain imaging genetics in Alzheimer’s and aging research. I acknowledge the contribution of Mr. Zhiyuan (Peter) Song to develop and maintain the MEDICINE knowledge portal.

## Dxtended Data Figures

**Extended Data Figure 1:**
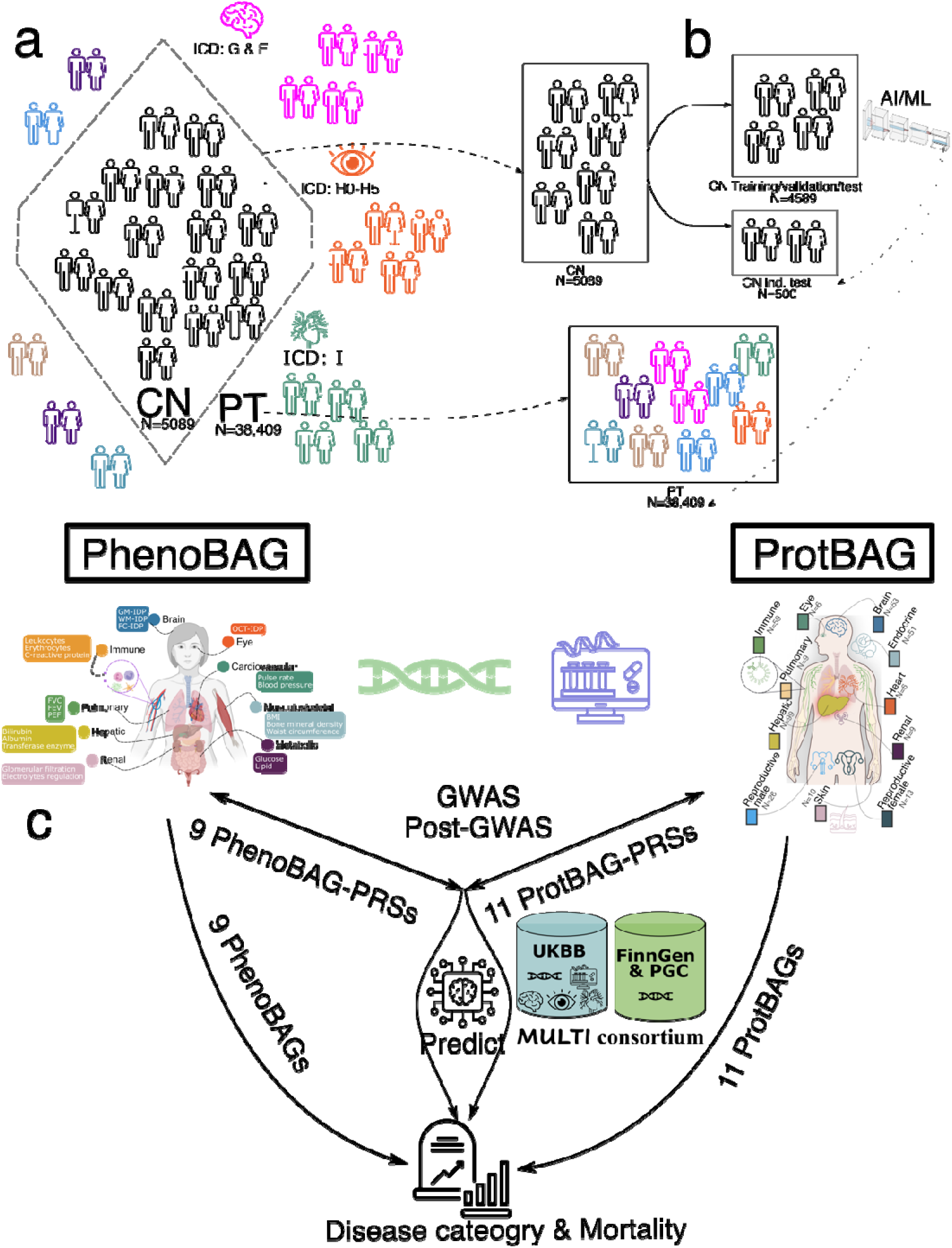
Schematic diagram of the definition of populations to derive ProtBAG and overall analytic workflow of the study. **a**) I first split the entire proteomics population in the UK Biobank into 5089 healthy control (CN) and 38,409 patient (PT) populations based on the ICD-10 code and other clinical history information. **b**) To derive the 11 ProtBAGs, I trained three AI/ML models using only the CN training/validation/test population (*N*=4589) with a (nested) cross-validation procedure to select the optimal model. The CN independent test (ind. test; *N*=500) and the PT population (*N*=38,409) were used as independent datasets. **c**) The analytical workflow of this study involved deriving 11 ProtBAGs, integrating them with 9 PhenoBAGs, and conducting GWAS and post-GWAS analyses. The ProtBAGs, PhenoBAGs, and their PRSs were then evaluated for their predictive power across 14 systemic disease categories and mortality outcomes.

**Extended Data Figure 2:**
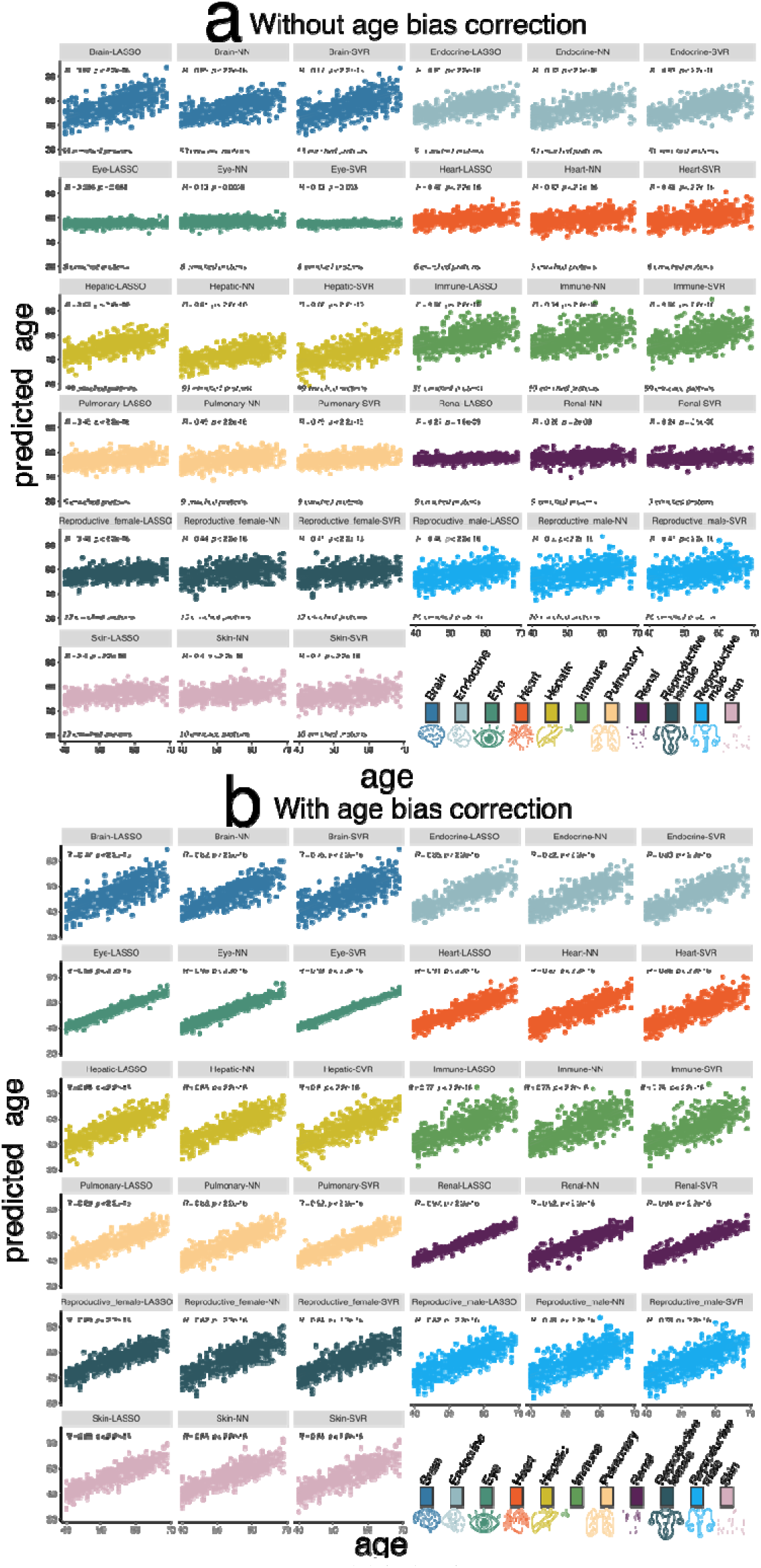
The scatter plot between AI/ML-predicted biological age and chronological age before and after age bias correction. **a**) The scatter plot between the AI/ML-derived biological age and chronological age without applying the age bias correction. **b**) The scatter plot between the AI/ML-derived biological age and chronological age after the age bias correction is applied.

**Extended Data Figure 3:**
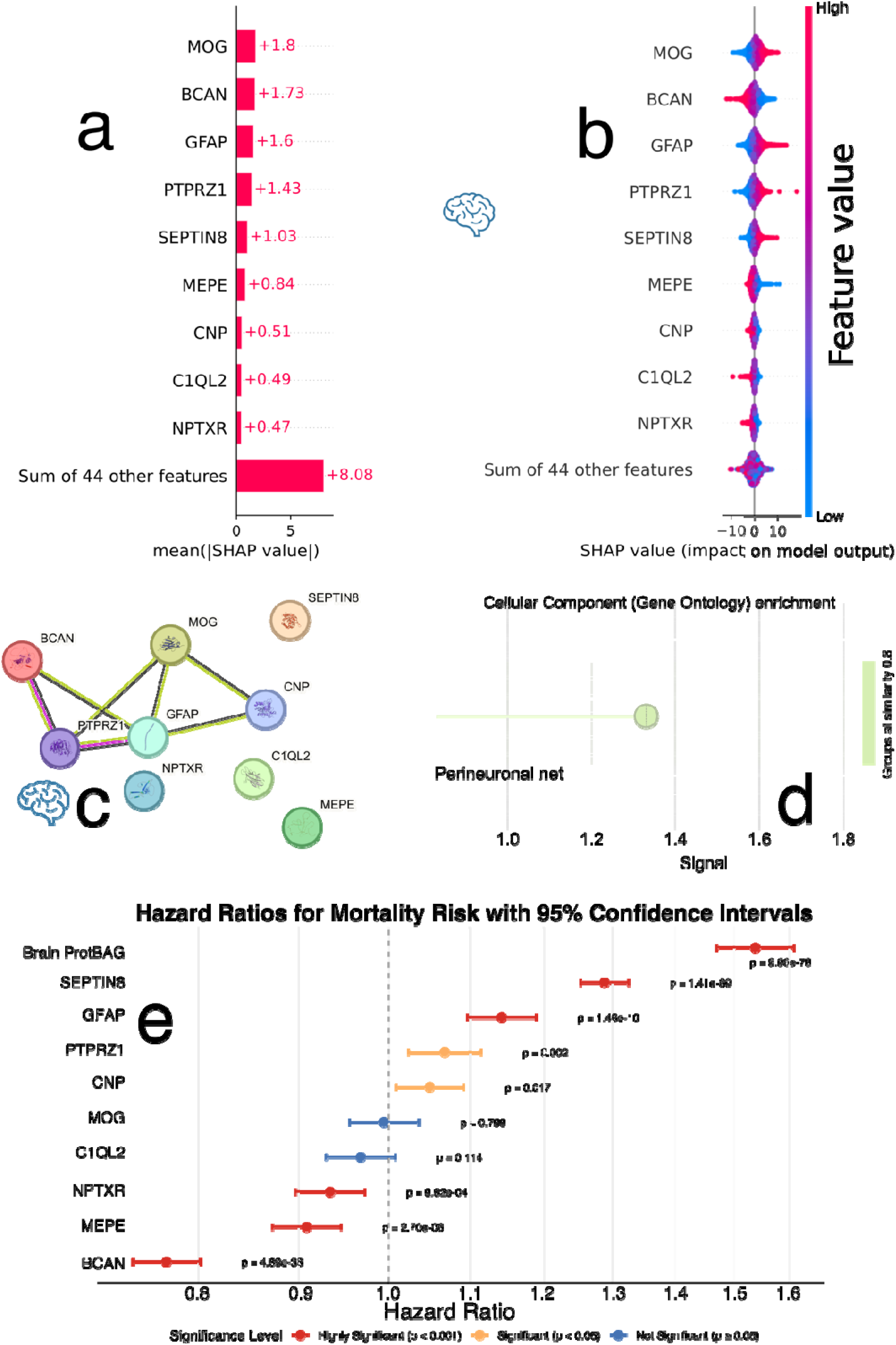
Feature importance of the organ-enriched proteins for the brain ProtBAG. **a**) SHAP analysis of key brain tissue-enriched proteins for the brain ProtBAG. The left panel presents a bar plot of mean absolute SHAP values, indicating each protein’s overall contribution to the model’s predictions. **b**) The right panel shows a bee swarm plot, illustrating the distribution of SHAP values across individual training samples. Positive SHAP values indicate that the increasing protein abundance contributes to predicting an older biological age, while negative values suggest a contribution toward a younger predicted age. Protein abundance values are color-coded: red for high values and blue for low. **c)** STRING conducts protein-protein interaction (PPI) analysis by combining various sources of interaction evidence, such as experimental data, computational predictions, and text mining. In addition to interactions, STRING can carry out functional enrichment analysis to identify Gene Ontology (GO) terms (e.g., biological processes, molecular functions), KEGG pathways (e.g., metabolic and signaling pathways), and protein domains (e.g., shared structural motifs). Protein-protein interaction network of the 9 most influential brain ProtBAG-related proteins. **d**) Functional protein-set enrichment analysis of the brain ProtBAG-related proteins. Results and figures are generated using STRING v12.0 (https://string-db.org/). **e**) All-cause mortality prediction (**Method 6b**) using the brain ProtBAG compared to the predictions power using the 9 most influential brain ProtBAG-related proteins.

**Extended Data Figure 4:**
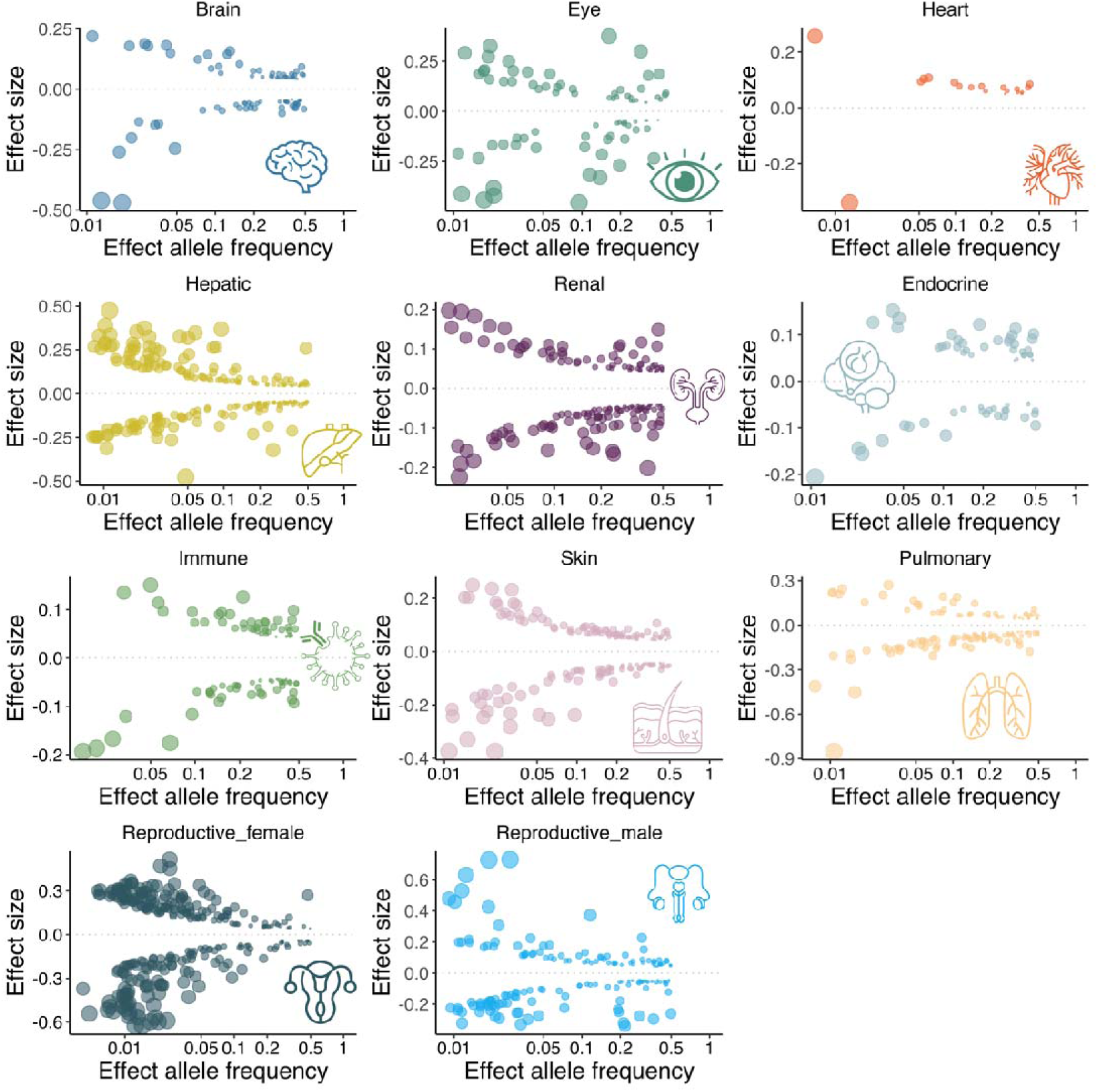
Trumpet plots of the effect allele frequency vs. the *β* coefficient of the 11 ProtBAG GWASs. The trumpet plots display the inverse relationship between the alternative (effect) allele frequency and the effect size (*β* coefficient) for the 11 ProtBAGs. I present the independent significant SNPs defined in FUMA. The dot size corresponds to the effect size, while the transparency of the dot is proportional to its statistical significance.

**Extended Data Figure 5:**
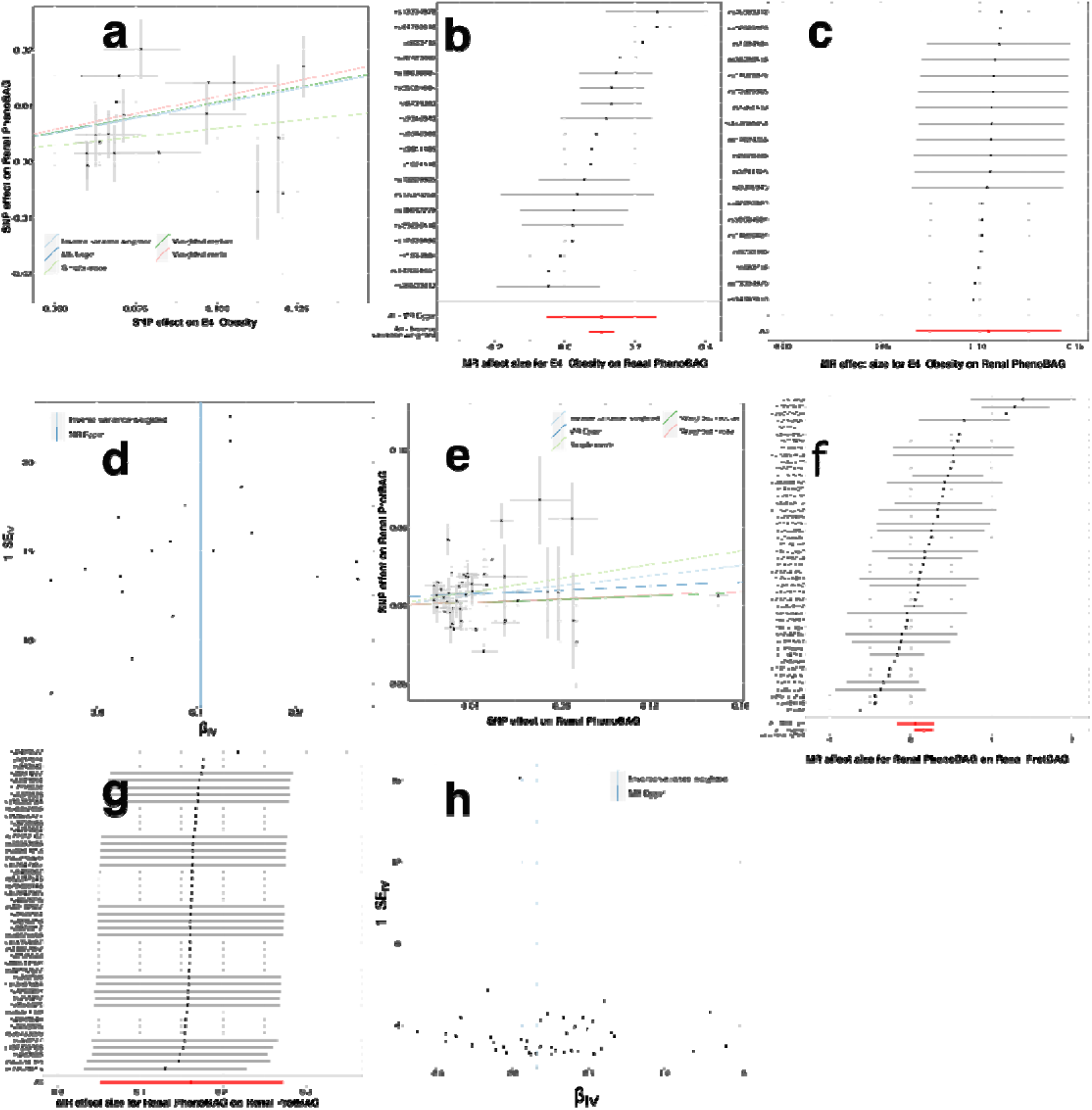
Sensitivity check analyses for the causal pathway of “obesity→renal PhenoBAG→renal ProtBAG”. **a)** Scatter plot for the MR effect sizes of the SNP-obesity association (*x*-axis, log OR) and the SNP-renal PhenoBAG associations (*y*-axis, log OR) with standard error bars. The slopes of the five lines correspond to the causal effect sizes estimated by the five MR estimators, respectively. **b)** Forest plot for the single-SNP MR results. Each dot represents the MR effect (log OR), and the error bar displays the 95% CI for Obesity on renal PhenoBAG using only one SNP; the red line shows the MR effect using all SNPs together for IVW and MR Egger estimators. **c**) Leave-one-SNP-out analysis of obesity on renal PhenoBAG. Each dot represents the MR effect (log OR), and the error bar displays the 95% CI by excluding that SNP from the analysis. The red line depicts the IVW estimator using all SNPs. **d**) Funnel plot for the relationship between the causal effect of obesity on renal PhenoBAG. Each dot represents MR effect sizes estimated using each SNP as a separate instrument against the inverse of the standard error of the causal estimate. **e**) Scatter plot for the MR effect sizes of the SNP-renal PhenoBAG association (*x*-axis, log OR) and the SNP-renal ProtBAG associations (*y*-axis, SD units) with standard error bars. The slopes of the five lines correspond to the causal effect sizes estimated by the five MR estimators, respectively. **f**) Forest plot for the single-SNP MR results. Each dot represents the MR effect (log OR), and the error bar displays the 95% CI for renal PhenoBAG on renal ProtBAG using only one SNP; the red line shows the MR effect using all SNPs together for IVW and MR Egger estimators. **g**) Leave-one-SNP-out analysis of renal PhenoBAG on renal ProtBAG. Each dot represents the MR effect (log OR), and the error bar displays the 95% CI by excluding that SNP from the analysis. The red line depicts the IVW estimator using all SNPs. **h**) Funnel plot for the relationship between the causal effect of renal PhenoBAG on renal ProtBAG. Each dot represents MR effect sizes estimated using each SNP as a separate instrument against the inverse of the standard error of the causal estimate.

