## Supplementary materials for "Refining the generation, interpretation, and application of multi-organ, multi-omics biological aging clocks"

### Online Supplementary Materials

**eNote 1: The tissue-enriched proteins to derive the 11 ProtBAGs**
**eNote 2: Sex-stratified brain ProtBAG**
**eNote 3: The definition of genomic loci, independent significant SNP, lead SNP, candidate SNP**
**eNote 4: The phenotypic and genetic correlation between the 11 ProtBAGs and 2448 proteins**
**eNote 5: Sensitivity check analyses for the MR results for the causal pathway of "obesity→renal PhenoBAG→renal ProtBAG"**
**eFigure 1: The LOWESS method for correcting the age bias, the comparison to the neuroimaging-based method, and the influence of sample size using the full set of 2448 proteins**
**eFigure 2: The U-shaped relationship between the beta coefficient and the number of epochs in the neural network for the association between the brain ProtBAG and age at death**
**eFigure 3: Organ specificity's influence on model overfitting using the 31,808 participants with mixed pathologies and protein collinearity among different categories using the down-sampled 53 proteins**
**eFigure 4: Organ specificity proteins vs. non-enriched proteins for predicting the brain ProtBAG**
**eFigure 5: Sex-stratified ML models for deriving the brain ProtBAG**
**eFigure 6: Feature importance of the organ-enriched proteins for deriving the 11 ProtBAGs**
**eFigure 7: Phenome-wide associations via the GWAS Atlas platform**
**eFigure 8: Multi-omic evidence for the *SERPINF2* gene in co-localization analysis**
**eFigure 9: The systemic disease category classification performance with age and sex as additional features**
**eFigure 10: Compare the brain ProtBAG derived from 4589 CN participants and 31,808 mixed-pathology participants**
**eFigure 11: Protein missing NPX value imputation quality**
**eFigure 12: UK biobank population selections and the nested cross-validation procedure to derive the 11 ProtBAGs**
**eTable 1: The characteristics of the MULTI consortium**
**eTable 2: Benchmarking the ProtBAG prediction performance for the CN training/validation/test (Training) and CN independent test (Ind. test) datasets**
**eTable 3: The relationship between the beta coefficient and the number of epochs by associating the brain ProtBAG with 8 cognitive scores and age at death**
**eTable 4: Genomic loci of the 11 ProtBAGs and 9 PhenoBAGs defined by FUMA**
**eTable 5a: Phenotypic and genetic correlations between the 11 ProtBAGs and 9 PhenoBAGs**
**eTable 5b: Phenotypic and genetic correlations between the 11 ProtBAGs and 2448 proteins**
**eTable 6: The incremental $R^2$ of the 20 ProtBAG-PRS and PhenoBAG-PRS**
**eTable 7: The Mendelian randomization results**
**eTable 8: The classification results to predict the 14 systemic disease categories**
**eTable 9: The survival analysis to predict mortality**

**eTable 10: Incremental R2 contributed by the 11 ProtBAGs on top of age, sex, and conventional non-organ specific ProtBAG in Argentieri**

**eTable 11: The statistics of the linear regression between the brain ProtBAG with/without age bias correction and age at death and DSST**

**eNote 1: The tissue-enriched proteins to derive the 11 ProtBAGs and the phenotypic features to derive the 9 PhenoBAGs from our previous study**

**Organ-specific proteins for the 11 ProtBAGs:**

I defined the organ-specific tissue-enriched proteins as detailed in **Method 3** using the HPA platform. Here is the detailed list of proteins for generating the 11 ProtBAGs.

**Brain ProtBAG**: 'PMCH', 'MOG', 'OXT', 'NCAN', 'MEPE', 'NRGN', 'MAG', 'MDGA1', 'CNTN2', 'CNP', 'VSNL1', 'GRIN2B', 'BCAN', 'CRTAM', 'GRIK2', 'CBLN1', 'CNDP1', 'NPTXR', 'LRTM2', 'SEPTIN8', 'C1QL2', 'CA11', 'SEPTIN3', 'ABCA2', 'APLP1', 'CRH', 'GFAP', 'IDS', 'IGSF21', 'RTN4R', 'FGFR2', 'IGLON5', 'LHPP', 'NPTX1', 'TUBB3', 'ADAM22', 'DNM1', 'ERC2', 'GP1BB', 'KLK6', 'PTPRZ1', 'SEZ6L', 'SLITRK1', 'SNAP25', 'CEND1', 'CLIP2', 'EDIL3', 'HPCAL1', 'ICAM5', 'OMG', 'PCDH9', 'SEZ6', 'TAFA5'.

**Eye ProtBAG**: 'CRX', 'IMPG1', 'PCARE', 'CLUL1', 'CABP2', 'RTBDN', 'STX3', 'WIF1'.

**Heart ProtBAG**: 'NPPB', 'BMP10', 'MYL4', 'PXDNL', 'CRIP2', 'FGF12'.

**Pulmonary ProtBAG**: 'SFTPA1', 'SFTPA2', 'SCGB3A2', 'SFTPD', 'AGER', 'SCGB1A1', 'LAMP3', 'CCL18', 'MSR1'.

**Renal ProtBAG**: 'UMOD', 'NPHS2', 'SOST', 'REN', 'PTH1R', 'SLC13A1', 'GGACT', 'PDZK1'.

**Hepatic ProtBAG**: 'AHSG', 'CFHR2', 'MBL2', 'F9', 'CFHR5', 'A1BG', 'SERPINC1', 'APOA2', 'F2', 'ITIH1', 'SERPINA7', 'HAO1', 'APOH', 'CFHR4', 'FGA', 'APOF', 'AGXT', 'F12', 'C9', 'F13B', 'ORM1', 'HRG', 'INHBC', 'GDF2', 'C8B', 'SAA4', 'FGF21', 'ITIH3', 'PON1', 'F7', 'SERPINA11', 'CPB2', 'APCS', 'AMBP', 'LPA', 'HGFAC', 'LECT2', 'ANG', 'ASGR2', 'CCL16', 'PGLYRP2', 'SERPINA1', 'CA5A', 'ASGR1', 'PROC', 'ITIH4', 'PZP', 'ADH4', 'C5', 'KLKB1', 'FETUB', 'LBP', 'LRG1', 'CFB', 'PON3', 'SERPINA6', 'AKR1C4', 'GC', 'SERPINF2', 'ANGPTL3', 'BCHE', 'C4BPB', 'HSD11B1', 'AFM', 'PLG', 'AFP', 'MST1', 'APOC1', 'C3', 'CES1', 'CLEC1B', 'UPB1', 'DCXR', 'AGT', 'C2', 'CFH', 'EPO', 'FGL1', 'FTCD', 'IGFBP1', 'IL1RAP', 'APOA1', 'C1S', 'CFI', 'GCHFR', 'PCSK9', 'SERPINA4', 'SERPIND1', 'SHBG', 'SULT2A1', 'THPO', 'ACADSB', 'APOM', 'ARG1', 'F11', 'FCN2', 'FUOM', 'LEPR', 'RNASE4'.

**Immune ProtBAG**: 'SH2D1A', 'GRAP2', 'SERPINA9', 'CD8A', 'TCL1A', 'CD1C', 'CR2', 'CTSV', 'CXCL9', 'CD3G', 'CXCL13', 'SIT1', 'SLAMF1', 'TIGIT', 'CD72', 'FCRL3', 'FCRL5', 'CCL17', 'CCL21', 'CD5L', 'LILRB1', 'STAB2', 'VCAM1', 'CCL19', 'CD27', 'CD79B', 'FCER2', 'HMOX1', 'LY75', 'NCR1', 'SLAMF6', 'MPO', 'RNASE3', 'RAB44', 'AHSP', 'PRTN3', 'AZU1', 'MMP8', 'CEACAM8', 'TARM1', 'SLC4A1', 'PGLYRP1', 'PRG3', 'CLC', 'EREG', 'CLEC4D', 'VSTM1', 'CCL7', 'ITGAM', 'OSM', 'CST7', 'CXCL8', 'RELT', 'S100A12', 'CLEC5A', 'HMBS', 'PLAUR', 'RETN', 'FOLR3'.

**Endocrine ProtBAG**: 'CCN3', 'AKR1B1', 'DBH', 'SCARB1', 'CELA2A', 'CPA1', 'CTRB1', 'CELA3A', 'PNLIP', 'AMY2A', 'CLPS', 'CTRC', 'PNLIPRP1', 'PLA2G1B', 'CPB1', 'CTRL', 'GP2', 'PRSS2', 'CPA2', 'SERPINI2', 'AMY2B', 'PNLIPRP2', 'SPINK1', 'PPY', 'REG3G', 'REG1A', 'REG1B', 'GCG', 'GPHA2', 'RNASE1', 'KIRREL2', 'SEL1L', 'COCH', 'IL22RA1', 'TG',

'IGFBPL1', 'PTH', 'CD109', 'ALCAM', 'CHGA', 'ATP6AP2', 'TSHB', 'PRL', 'GH1', 'POMC', 'GHRHR', 'FSHB', 'LHB', 'GAL', 'CGA', 'NPTX2'.

**Skin ProtBAG**: 'CDSN', 'FABP9', 'CST6', 'CCL27', 'SERPINA12', 'DSG4', 'CD207', 'EPPK1', 'S100A3', 'WFDC12'

**Reproductive female ProtBAG**: 'BTN1A1', 'STC2', 'MMP10', 'PSG1', 'SIGLEC6', 'PRG2', 'TFPI2', 'INSL4', 'ADAM12', 'HGF', 'FCGR2B', 'FLT1', 'PAPPA'.

**Reproductive male ProtBAG**: 'LYZL2', 'DNAJB8', 'ZNRF4', 'DKKL1', 'PDCL2', 'TMCO5A', 'GAGE2A', 'TEX101', 'BRDT', 'IL13', 'ACRV1', 'CRISP2', 'ACRBP', 'IZUMO1', 'LRRC37A2', 'IL5', 'SPESP1', 'PHOSPHO1', 'KLK3', 'KLK4', 'MSMB', 'EDDM3B', 'SPINK2', 'NPC2', 'ADGRG2', 'ENPP5'.

**Organ-specific phenotypes for the 9 PhenoBAGs:**
The phenotypic features used to derive the 9 PhenoBAGs are detailed in the Supplementary Materials from our previous studies[1.] A full list of the organ-specific feature is presented in **Supplementary eFile 21** in Wen et al.[2].

**Brain PhenoBAG**: Area of G+S-frontomargin (left hemisphere), Area of G+S-occipital-inf (left hemisphere), Area of G+S-paracentral (left hemisphere), Area of G+S-subcentral (left hemisphere), Area of G+S-transv-frontopol (left hemisphere), Area of G+S-cingul-Ant (left hemisphere), Area of G+S-cingul-Mid-Ant (left hemisphere), Area of G+S-cingul-Mid-Post (left hemisphere), Area of G-cingul-Post-dorsal (left hemisphere), Area of G-cingul-Post-ventral (left hemisphere), Area of G-cuneus (left hemisphere), Area of G-front-inf-Opercular (left hemisphere), Area of G-front-inf-Orbital (left hemisphere), Area of G-front-inf-Triangul (left hemisphere), Area of G-front-middle (left hemisphere), Area of G-front-sup (left hemisphere), Area of G-Ins-lg+S-cent-ins (left hemisphere), Area of G-insular-short (left hemisphere), Area of G-occipital-middle (left hemisphere), Area of G-occipital-sup (left hemisphere), Area of G-oc-temp-lat-fusifor (left hemisphere), Area of G-oc-temp-med-Lingual (left hemisphere), Area of G-oc-temp-med-Parahip (left hemisphere), Area of G-orbital (left hemisphere), Area of G-pariet-inf-Angular (left hemisphere), Area of G-pariet-inf-Supramar (left hemisphere), Area of G-parietal-sup (left hemisphere), Area of G-postcentral (left hemisphere), Area of G-precentral (left hemisphere), Area of G-precuneus (left hemisphere), Area of G-rectus (left hemisphere), Area of G-subcallosal (left hemisphere), Area of G-temp-sup-G-T-transv (left hemisphere), Area of G-temp-sup-Lateral (left hemisphere), Area of G-temp-sup-Plan-polar (left hemisphere), Area of G-temp-sup-Plan-tempo (left hemisphere), Area of G-temporal-inf (left hemisphere), Area of G-temporal-middle (left hemisphere), Area of Lat-Fis-ant-Horizont (left hemisphere), Area of Lat-Fis-ant-Vertical (left hemisphere), Area of Lat-Fis-post (left hemisphere), Area of Pole-occipital (left hemisphere), Area of Pole-temporal (left hemisphere), Area of S-calcarine (left hemisphere), Area of S-central (left hemisphere), Area of S-cingul-Marginalis (left hemisphere), Area of S-circular-insula-ant (left hemisphere), Area of S-circular-insula-inf (left hemisphere), Area of S-circular-insula-sup (left hemisphere), Area of S-collat-transv-ant (left hemisphere), Area of S-collat-transv-post (left hemisphere), Area of S-front-inf (left hemisphere), Area of S-front-middle (left hemisphere), Area of S-front-sup (left hemisphere), Area of S-interm-prim-Jensen (left hemisphere), Area of S-intrapariet+P-trans (left hemisphere), Area of S-oc-middle+Lunatus (left hemisphere), Area of S-oc-sup+transversal (left hemisphere), Area of S-occipital-ant (left hemisphere), Area of S-oc-temp-lat (left hemisphere), Area of S-oc-temp-med+Lingual (left hemisphere), Area of S-orbital-lateral (left hemisphere), Area of S-orbital-med-olfact (left hemisphere), Area of S-orbital-H-Shaped (left hemisphere), Area of S-parieto-occipital (left hemisphere), Area of S-pericallosal (left hemisphere), Area of S-postcentral (left hemisphere), Area of S-precentral-inf-part (left hemisphere), Area of S-precentral-sup-part (left hemisphere), Area of S-suborbital (left hemisphere), Area of S-subparietal (left hemisphere), Area of S-temporal-inf (left hemisphere), Area of S-temporal-sup (left hemisphere), Area of S-temporal-transverse (left hemisphere), Mean thickness of G+S-frontomargin (left hemisphere), Mean thickness of G+S-occipital-inf (left hemisphere), Mean thickness of G+S-paracentral (left hemisphere), Mean thickness of G+S-subcentral (left hemisphere), Mean thickness of G+S-transv-frontopol (left hemisphere), Mean thickness of G+S-cingul-Ant (left hemisphere), Mean thickness of G+S-cingul-Mid-Ant (left hemisphere), Mean thickness of G+S-cingul-Mid-Post (left hemisphere), Mean thickness of G-cingul-Post-dorsal (left hemisphere), Mean thickness of G-cingul-Post-ventral (left hemisphere), Mean thickness of G-cuneus (left hemisphere), Mean thickness of G-front-inf-Opercular (left hemisphere), Mean thickness of G-front-inf-Orbital (left hemisphere), Mean thickness of G-front-inf-Triangul (left hemisphere), Mean thickness of G-front-middle (left hemisphere), Mean thickness of G-front-sup (left hemisphere), Mean thickness of G-Ins-lg+S-cent-ins (left hemisphere), Mean thickness of G-insular-short (left hemisphere), Mean thickness of G-occipital-middle (left hemisphere), Mean thickness of G-occipital-sup (left hemisphere), Mean thickness of G-oc-temp-lat-fusifor (left hemisphere), Mean thickness of G-oc-temp-med-Lingual (left hemisphere), Mean thickness of G-oc-temp-med-Parahip (left hemisphere), Mean thickness of G-orbital (left hemisphere), Mean thickness of G-pariet-inf-Angular (left hemisphere), Mean thickness of G-pariet-inf-Supramar (left hemisphere), Mean thickness of G-parietal-sup (left hemisphere), Mean thickness of G-postcentral (left hemisphere), Mean thickness of G-precentral (left hemisphere), Mean thickness of G-precuneus (left hemisphere), Mean thickness of G-rectus (left hemisphere), Mean thickness of G-subcallosal (left hemisphere), Mean thickness of G-temp-sup-G-T-transv (left hemisphere), Mean thickness of G-temp-sup-Lateral (left hemisphere), Mean thickness of G-temp-sup-Plan-polar (left hemisphere), Mean thickness of G-temp-sup-Plan-tempo (left hemisphere), Mean thickness of G-temporal-inf (left hemisphere), Mean thickness of G-temporal-middle (left hemisphere), Mean thickness of Lat-Fis-ant-Horizont (left hemisphere), Mean thickness of Lat-Fis-ant-Vertical (left hemisphere), Mean thickness of Lat-Fis-post (left hemisphere), Mean thickness of Pole-occipital (left hemisphere), Mean thickness of Pole-temporal (left hemisphere), Mean thickness of S-calcarine (left hemisphere), Mean thickness of S-central (left hemisphere), Mean thickness of S-cingul-Marginalis (left hemisphere), Mean thickness of S-circular-insula-ant (left hemisphere), Mean thickness of S-circular-insula-inf (left hemisphere), Mean thickness of S-circular-insula-sup (left hemisphere), Mean thickness of S-collat-transv-ant (left hemisphere), Mean thickness of S-collat-transv-post (left hemisphere), Mean thickness of S-front-inf (left hemisphere), Mean thickness of S-front-middle (left hemisphere), Mean thickness of S-front-sup (left hemisphere), Mean thickness of S-interm-prim-Jensen (left hemisphere), Mean thickness of S-intrapariet+P-trans (left hemisphere), Mean thickness of S-oc-middle+Lunatus (left hemisphere), Mean thickness of S-oc-sup+transversal (left hemisphere), Mean thickness of S-occipital-ant (left hemisphere), Mean thickness of S-oc-temp-lat (left hemisphere), Mean thickness of S-oc-temp-med+Lingual (left hemisphere), Mean thickness of S-orbital-lateral (left hemisphere), Mean thickness of S-orbital-med-olfact (left hemisphere), Mean thickness of S-orbital-H-Shaped (left hemisphere), Mean thickness of S-parieto-occipital (left hemisphere), Mean thickness of S-pericallosal (left hemisphere), Mean thickness of S-postcentral (left hemisphere), Mean thickness of S-precentral-inf-part (left hemisphere), Mean thickness of S-precentral-sup-part (left hemisphere), Mean thickness of S-suborbital (left hemisphere), Mean thickness of S-subparietal (left hemisphere), Mean thickness of S-temporal-inf (left hemisphere), Mean thickness of S-temporal-sup (left hemisphere), Mean thickness of S-temporal-transverse (left hemisphere), Volume of G+S-frontomargin (left hemisphere), Volume of G+S-occipital-inf (left hemisphere), Volume of G+S-paracentral (left hemisphere), Volume of G+S-subcentral (left hemisphere), Volume of G+S-transv-frontopol (left hemisphere), Volume of G+S-cingul-Ant (left hemisphere), Volume of G+S-cingul-Mid-Ant (left hemisphere), Volume of G+S-cingul-Mid-Post (left hemisphere), Volume of G-cingul-Post-dorsal (left hemisphere), Volume of G-cingul-Post-ventral (left hemisphere), Volume of G-cuneus (left hemisphere), Volume of G-front-inf-Opercular (left hemisphere), Volume of G-front-inf-Orbital (left hemisphere), Volume of G-front-inf-Triangul (left hemisphere), Volume of G-front-middle (left hemisphere), Volume of G-front-sup (left hemisphere), Volume of G-Ins-lg+S-cent-ins (left hemisphere), Volume of G-insular-short (left hemisphere), Volume of G-occipital-middle (left hemisphere), Volume of G-occipital-sup (left hemisphere), Volume of G-oc-temp-lat-fusifor (left hemisphere), Volume of G-oc-temp-med-Lingual (left hemisphere), Volume of G-oc-temp-med-Parahip (left hemisphere), Volume of G-orbital (left hemisphere), Volume of G-pariet-inf-Angular (left hemisphere), Volume of G-pariet-inf-Supramar (left hemisphere), Volume of G-parietal-sup (left hemisphere), Volume of G-postcentral (left hemisphere), Volume of G-precentral (left hemisphere), Volume of G-precuneus (left hemisphere), Volume of G-rectus (left hemisphere), Volume of G-subcallosal (left hemisphere), Volume of G-temp-sup-G-T-transv (left hemisphere), Volume of G-temp-sup-Lateral (left hemisphere), Volume of G-temp-sup-Plan-polar (left hemisphere), Volume of G-temp-sup-Plan-tempo (left hemisphere), Volume of G-temporal-inf (left hemisphere), Volume of G-temporal-middle (left hemisphere), Volume of Lat-Fis-ant-Horizont (left hemisphere), Volume of Lat-Fis-ant-Vertical (left hemisphere), Volume of Lat-Fis-post (left hemisphere), Volume of Pole-occipital (left hemisphere), Volume of Pole-temporal (left hemisphere), Volume of S-calcarine (left hemisphere), Volume of S-central (left hemisphere), Volume of S-cingul-Marginalis (left hemisphere), Volume of S-circular-insula-ant (left hemisphere), Volume of S-circular-insula-inf (left hemisphere), Volume of S-circular-insula-sup (left hemisphere), Volume of S-collat-transv-ant (left hemisphere), Volume of S-collat-transv-post (left hemisphere), Volume of S-front-inf (left hemisphere), Volume of S-front-middle (left hemisphere), Volume of S-front-sup (left hemisphere), Volume of S-interm-prim-Jensen (left hemisphere), Volume of S-intrapariet+P-trans (left hemisphere), Volume of S-oc-middle+Lunatus (left hemisphere), Volume of S-oc-sup+transversal (left hemisphere), Volume of S-occipital-ant (left hemisphere), Volume of S-oc-temp-lat (left hemisphere), Volume of S-oc-temp-med+Lingual (left hemisphere), Volume of S-orbital-lateral (left hemisphere), Volume of S-orbital-med-olfact (left hemisphere), Volume of S-orbital-H-Shaped (left hemisphere), Volume of S-parieto-occipital (left hemisphere), Volume of S-pericallosal (left hemisphere), Volume of S-postcentral (left hemisphere), Volume of S-precentral-inf-part (left hemisphere), Volume of S-precentral-sup-part (left hemisphere), Volume of S-suborbital (left hemisphere), Volume of S-subparietal (left hemisphere), Volume of S-temporal-inf (left hemisphere), Volume of S-temporal-sup (left hemisphere), Volume of S-temporal-transverse (left hemisphere), Area of G+S-frontomargin (right hemisphere), Area of G+S-occipital-inf (right hemisphere), Area of G+S-paracentral (right hemisphere), Area of G+S-subcentral (right hemisphere), Area of G+S-transv-frontopol (right hemisphere), Area of G+S-cingul-Ant (right hemisphere), Area of G+S-cingul-Mid-Ant (right hemisphere), Area of G+S-cingul-Mid-Post (right hemisphere), Area of G-cingul-Post-dorsal (right hemisphere), Area of G-cingul-Post-ventral (right hemisphere), Area of G-cuneus (right hemisphere), Area of G-front-inf-Opercular (right hemisphere), Area of G-front-inf-Orbital (right hemisphere), Area of G-front-inf-Triangul (right hemisphere), Area of G-front-middle (right hemisphere), Area of G-front-sup (right hemisphere), Area of G-Ins-lg+S-cent-ins (right hemisphere), Area of G-insular-short (right hemisphere), Area of G-occipital-middle (right hemisphere), Area of G-occipital-sup (right hemisphere), Area of G-oc-temp-lat-fusifor (right hemisphere), Area of G-oc-temp-med-Lingual (right hemisphere), Area of G-oc-temp-med-Parahip (right hemisphere), Area of G-orbital (right hemisphere), Area of G-pariet-inf-Angular (right hemisphere), Area of G-pariet-inf-Supramar (right hemisphere), Area of G-parietal-sup (right hemisphere), Area of G-postcentral (right hemisphere), Area of G-precentral (right hemisphere), Area of G-precuneus (right hemisphere), Area of G-rectus (right hemisphere), Area of G-subcallosal (right hemisphere), Area of G-temp-sup-G-T-transv (right hemisphere), Area of G-temp-sup-Lateral (right hemisphere), Area of G-temp-sup-Plan-polar (right hemisphere), Area of G-temp-sup-Plan-tempo (right hemisphere), Area of G-temporal-inf (right hemisphere), Area of G-temporal-middle (right hemisphere), Area of Lat-Fis-ant-Horizont (right hemisphere), Area of Lat-Fis-ant-Vertical (right hemisphere), Area of Lat-Fis-post (right hemisphere), Area of Pole-occipital (right hemisphere), Area of Pole-temporal (right hemisphere), Area of S-calcarine (right hemisphere), Area of S-central (right hemisphere), Area of S-cingul-Marginalis (right hemisphere), Area of S-circular-insula-ant (right hemisphere), Area of S-circular-insula-inf (right hemisphere), Area of S-circular-insula-sup (right hemisphere), Area of S-collat-transv-ant (right hemisphere), Area of S-collat-transv-post (right hemisphere), Area of S-front-inf (right hemisphere), Area of S-front-middle (right hemisphere), Area of S-front-sup (right hemisphere), Area of S-interm-prim-Jensen (right hemisphere), Area of S-intrapariet+P-trans (right hemisphere), Area of S-oc-middle+Lunatus (right hemisphere), Area of S-oc-sup+transversal (right hemisphere), Area of S-occipital-ant (right hemisphere), Area of S-oc-temp-lat (right hemisphere), Area of S-oc-temp-med+Lingual (right hemisphere), Area of S-orbital-lateral (right hemisphere), Area of S-orbital-med-olfact (right hemisphere), Area of S-orbital-H-Shaped (right hemisphere), Area of S-parieto-occipital (right hemisphere), Area of S-pericallosal (right hemisphere), Area of S-postcentral (right hemisphere), Area of S-precentral-inf-part (right hemisphere), Area of S-precentral-sup-part (right hemisphere), Area of S-suborbital (right hemisphere), Area of S-subparietal (right hemisphere), Area of S-temporal-inf (right hemisphere), Area of S-temporal-sup (right hemisphere), Area of S-temporal-transverse (right hemisphere), Mean thickness of G+S-frontomargin (right hemisphere), Mean thickness of G+S-occipital-inf (right hemisphere), Mean thickness of G+S-paracentral (right hemisphere), Mean thickness of G+S-subcentral (right hemisphere), Mean thickness of G+S-transv-frontopol (right hemisphere), Mean thickness of G+S-cingul-Ant (right hemisphere), Mean thickness of G+S-cingul-Mid-Ant (right hemisphere), Mean thickness of G+S-cingul-Mid-Post (right hemisphere), Mean thickness of G-cingul-Post-dorsal (right hemisphere), Mean thickness of G-cingul-Post-ventral (right hemisphere), Mean thickness of G-cuneus (right hemisphere), Mean thickness of G-front-inf-Opercular (right hemisphere), Mean thickness of G-front-inf-Orbital (right hemisphere), Mean thickness of G-front-inf-Triangul (right hemisphere), Mean thickness of G-front-middle (right hemisphere), Mean thickness of G-front-sup (right hemisphere), Mean thickness of G-Ins-lg+S-cent-ins (right hemisphere), Mean thickness of G-insular-short (right hemisphere), Mean thickness of G-occipital-middle (right hemisphere), Mean thickness of G-occipital-sup (right hemisphere), Mean thickness of G-oc-temp-lat-fusifor (right hemisphere), Mean thickness of G-oc-temp-med-Lingual (right hemisphere), Mean thickness of G-oc-temp-med-Parahip (right hemisphere), Mean thickness of G-orbital (right hemisphere), Mean thickness of G-pariet-inf-Angular (right hemisphere), Mean thickness of G-pariet-inf-Supramar (right hemisphere), Mean thickness of G-parietal-sup (right hemisphere), Mean thickness of G-postcentral (right hemisphere), Mean thickness of G-precentral (right hemisphere), Mean thickness of G-precuneus (right hemisphere), Mean thickness of G-rectus (right hemisphere), Mean thickness of G-subcallosal (right hemisphere), Mean thickness of G-temp-sup-G-T-transv (right hemisphere), Mean thickness of G-temp-sup-Lateral (right hemisphere), Mean thickness of G-temp-sup-Plan-polar (right hemisphere), Mean thickness of G-temp-sup-Plan-tempo (right hemisphere), Mean thickness of G-temporal-inf (right hemisphere), Mean thickness of G-temporal-middle (right hemisphere), Mean thickness of Lat-Fis-ant-Horizont (right hemisphere), Mean thickness of Lat-Fis-ant-Vertical (right hemisphere), Mean thickness of Lat-Fis-post (right hemisphere), Mean thickness of Pole-occipital (right hemisphere), Mean thickness of Pole-temporal (right hemisphere), Mean thickness of S-calcarine (right hemisphere), Mean thickness of S-central (right hemisphere), Mean thickness of S-cingul-Marginalis (right hemisphere), Mean thickness of S-circular-insula-ant (right hemisphere), Mean thickness of S-circular-insula-inf (right hemisphere), Mean thickness of S-circular-insula-sup (right hemisphere), Mean thickness of S-collat-transv-ant (right hemisphere), Mean thickness of S-collat-transv-post (right hemisphere), Mean thickness of S-front-inf (right hemisphere), Mean thickness of S-front-middle (right hemisphere), Mean thickness of S-front-sup (right hemisphere), Mean thickness of S-interm-prim-Jensen (right hemisphere), Mean thickness of S-intrapariet+P-trans (right hemisphere), Mean thickness of S-oc-middle+Lunatus (right hemisphere), Mean thickness of S-oc-sup+transversal (right hemisphere), Mean thickness of S-occipital-ant (right hemisphere), Mean thickness of S-oc-temp-lat (right hemisphere), Mean thickness of S-oc-temp-med+Lingual (right hemisphere), Mean thickness of S-orbital-lateral (right hemisphere), Mean thickness of S-orbital-med-olfact (right hemisphere), Mean thickness of S-orbital-H-Shaped (right hemisphere), Mean thickness of S-parieto-occipital (right hemisphere), Mean thickness of S-pericallosal (right hemisphere), Mean thickness of S-postcentral (right hemisphere), Mean thickness of S-precentral-inf-part (right hemisphere), Mean thickness of S-precentral-sup-part (right hemisphere), Mean thickness of S-suborbital (right hemisphere), Mean thickness of S-subparietal (right hemisphere), Mean thickness of S-temporal-inf (right hemisphere), Mean thickness of S-temporal-sup (right hemisphere), Mean thickness of S-temporal-transverse (right hemisphere), Volume of G+S-frontomargin (right hemisphere), Volume of G+S-occipital-inf (right hemisphere), Volume of G+S-paracentral (right hemisphere), Volume of G+S-subcentral (right hemisphere), Volume of G+S-transv-frontopol (right hemisphere), Volume of G+S-cingul-Ant (right hemisphere), Volume of G+S-cingul-Mid-Ant (right hemisphere), Volume of G+S-cingul-Mid-Post (right hemisphere), Volume of G-cingul-Post-dorsal (right hemisphere), Volume of G-cingul-Post-ventral (right hemisphere), Volume of G-cuneus (right hemisphere), Volume of G-front-inf-Opercular (right hemisphere), Volume of G-front-inf-Orbital (right hemisphere), Volume of G-front-inf-Triangul (right hemisphere), Volume of G-front-middle (right hemisphere), Volume of G-front-sup (right hemisphere), Volume of G-Ins-lg+S-cent-ins (right hemisphere), Volume of G-insular-short (right hemisphere), Volume of G-occipital-middle (right hemisphere), Volume of G-occipital-sup (right hemisphere), Volume of G-oc-temp-lat-fusifor (right hemisphere), Volume of G-oc-temp-med-Lingual (right hemisphere), Volume of G-oc-temp-med-Parahip (right hemisphere), Volume of G-orbital (right hemisphere), Volume of G-pariet-inf-Angular (right hemisphere), Volume of G-pariet-inf-Supramar (right hemisphere), Volume of G-parietal-sup (right hemisphere), Volume of G-postcentral (right hemisphere), Volume of G-precentral (right hemisphere), Volume of G-precuneus (right hemisphere), Volume of G-rectus (right hemisphere), Volume of G-subcallosal (right hemisphere), Volume of G-temp-sup-G-T-transv (right hemisphere), Volume of G-temp-sup-Lateral (right hemisphere), Volume of G-temp-sup-Plan-polar (right hemisphere), Volume of G-temp-sup-Plan-tempo (right hemisphere), Volume of G-temporal-inf (right hemisphere), Volume of G-temporal-middle (right hemisphere), Volume of Lat-Fis-ant-Horizont (right hemisphere), Volume of Lat-Fis-ant-Vertical (right hemisphere), Volume of Lat-Fis-post (right hemisphere), Volume of Pole-occipital (right hemisphere), Volume of Pole-temporal (right hemisphere), Volume of S-calcarine (right hemisphere), Volume of S-central (right hemisphere), Volume of S-cingul-Marginalis (right hemisphere), Volume of S-circular-insula-ant (right hemisphere), Volume of S-circular-insula-inf (right hemisphere), Volume of S-circular-insula-sup (right hemisphere), Volume of S-collat-transv-ant (right hemisphere), Volume of S-collat-transv-post (right hemisphere), Volume of S-front-inf (right hemisphere), Volume of S-front-middle (right hemisphere), Volume of S-front-sup (right hemisphere), Volume of S-interm-prim-Jensen (right hemisphere), Volume of S-intrapariet+P-trans (right hemisphere), Volume of S-oc-middle+Lunatus (right hemisphere), Volume of S-oc-sup+transversal (right hemisphere), Volume of S-occipital-ant (right hemisphere), Volume of S-oc-temp-lat (right hemisphere), Volume of S-oc-temp-med+Lingual (right hemisphere), Volume of S-orbital-lateral (right hemisphere), Volume of S-orbital-med-olfact (right hemisphere), Volume of S-orbital-H-Shaped (right hemisphere), Volume of S-parieto-occipital (right hemisphere), Volume of S-pericallosal (right hemisphere), Volume of S-postcentral (right hemisphere), Volume of S-precentral-inf-part (right hemisphere), Volume of S-precentral-sup-part (right hemisphere), Volume of S-suborbital (right hemisphere), Volume of S-subparietal (right hemisphere), Volume of S-temporal-inf (right hemisphere), Volume of S-temporal-sup (right hemisphere), Volume of S-temporal-transverse (right hemisphere), Mean intensity of 3rd-Ventricle (whole brain), Mean intensity of 4th-Ventricle (whole brain), Mean intensity of 5th-Ventricle (whole brain), Mean intensity of Brain-Stem (whole brain), Mean intensity of CSF (whole brain), Mean intensity of WM-hypointensities (whole brain), Mean intensity of non-WM-hypointensities (whole brain), Mean intensity of Optic-Chiasm (whole brain), Mean intensity of CC-Posterior (whole brain), Mean intensity of CC-Mid-Posterior (whole brain), Mean intensity of CC-Central (whole brain), Mean intensity of CC-Mid-Anterior (whole brain), Mean intensity of CC-Anterior (whole brain), Volume of BrainSeg (whole brain), Volume of BrainSegNotVent (whole brain), Volume of BrainSegNotVentSurf (whole brain), Volume of SubCortGray (whole brain), Volume of TotalGray (whole brain), Volume of SupraTentorial (whole brain), Volume of SupraTentorialNotVent (whole brain), Volume of EstimatedTotalIntraCranial (whole brain), Volume of VentricleChoroid (whole brain), Volume of 3rd-Ventricle (whole brain), Volume of 4th-Ventricle (whole brain), Volume of 5th-Ventricle (whole brain), Volume of Brain-Stem (whole brain), Volume of CSF (whole brain), Volume of WM-hypointensities (whole brain), Volume of non-WM-hypointensities (whole brain), Volume of Optic-Chiasm (whole brain), Volume of CC-Posterior (whole brain), Volume of CC-Mid-Posterior (whole brain), Volume of CC-Central (whole brain), Volume of CC-Mid-Anterior (whole brain), Volume of CC-Anterior (whole brain), Volume-ratio of BrainSegVol-to-eTIV (whole brain), Volume-ratio of MaskVol-to-eTIV (whole brain), Mean intensity of Lateral-Ventricle (left hemisphere), Mean intensity of Inf-Lat-Vent (left hemisphere), Mean intensity of Cerebellum-White-Matter (left hemisphere),

Mean intensity of Cerebellum-Cortex (left hemisphere), Mean intensity of Thalamus-Proper (left hemisphere), Mean intensity of Caudate (left hemisphere), Mean intensity of Putamen (left hemisphere), Mean intensity of Pallidum (left hemisphere), Mean intensity of Hippocampus (left hemisphere), Mean intensity of Amygdala (left hemisphere), Mean intensity of Accumbens-area (left hemisphere), Mean intensity of VentralDC (left hemisphere), Mean intensity of vessel (left hemisphere), Mean intensity of choroid-plexus (left hemisphere), Volume of Cortex (left hemisphere), Volume of CerebralWhiteMatter (left hemisphere), Volume of Lateral-Ventricle (left hemisphere), Volume of Inf-Lat-Vent (left hemisphere), Volume of Cerebellum-White-Matter (left hemisphere), Volume of Cerebellum-Cortex (left hemisphere), Volume of Thalamus-Proper (left hemisphere), Volume of Caudate (left hemisphere), Volume of Putamen (left hemisphere), Volume of Pallidum (left hemisphere), Volume of Hippocampus (left hemisphere), Volume of Amygdala (left hemisphere), Volume of Accumbens-area (left hemisphere), Volume of VentralDC (left hemisphere), Volume of vessel (left hemisphere), Volume of choroid-plexus (left hemisphere), Mean intensity of Lateral-Ventricle (right hemisphere), Mean intensity of Inf-Lat-Vent (right hemisphere), Mean intensity of Cerebellum-White-Matter (right hemisphere), Mean intensity of Cerebellum-Cortex (right hemisphere), Mean intensity of Thalamus-Proper (right hemisphere), Mean intensity of Caudate (right hemisphere), Mean intensity of Putamen (right hemisphere), Mean intensity of Pallidum (right hemisphere), Mean intensity of Hippocampus (right hemisphere), Mean intensity of Amygdala (right hemisphere), Mean intensity of Accumbens-area (right hemisphere), Mean intensity of VentralDC (right hemisphere), Mean intensity of vessel (right hemisphere), Mean intensity of choroid-plexus (right hemisphere), Volume of Cortex (right hemisphere), Volume of CerebralWhiteMatter (right hemisphere), Volume of Lateral-Ventricle (right hemisphere), Volume of Inf-Lat-Vent (right hemisphere), Volume of Cerebellum-White-Matter (right hemisphere), Volume of Cerebellum-Cortex (right hemisphere), Volume of Thalamus-Proper (right hemisphere), Volume of Caudate (right hemisphere), Volume of Putamen (right hemisphere), Volume of Pallidum (right hemisphere), Volume of Hippocampus (right hemisphere), Volume of Amygdala (right hemisphere), Volume of Accumbens-area (right hemisphere), Volume of VentralDC (right hemisphere), Volume of vessel (right hemisphere), Volume of choroid-plexus (right hemisphere), Volume of Lateral-nucleus (left hemisphere), Volume of Basal-nucleus (left hemisphere), Volume of Accessory-Basal-nucleus (left hemisphere), Volume of Anterior-amygdaloid-area-AAA (left hemisphere), Volume of Central-nucleus (left hemisphere), Volume of Medial-nucleus (left hemisphere), Volume of Cortical-nucleus (left hemisphere), Volume of Corticoamygdaloid-transitio (left hemisphere), Volume of Paralaminar-nucleus (left hemisphere), Volume of Whole-amygdala (left hemisphere), Volume of Lateral-nucleus (right hemisphere), Volume of Basal-nucleus (right hemisphere), Volume of Accessory-Basal-nucleus (right hemisphere), Volume of Anterior-amygdaloid-area-AAA (right hemisphere), Volume of Central-nucleus (right hemisphere), Volume of Medial-nucleus (right hemisphere), Volume of Cortical-nucleus (right hemisphere), Volume of Corticoamygdaloid-transitio (right hemisphere), Volume of Paralaminar-nucleus (right hemisphere), Volume of Whole-amygdala (right hemisphere), Volume of Hippocampal-tail (left hemisphere), Volume of subiculum-body (left hemisphere), Volume of CA1-body (left hemisphere), Volume of subiculum-head (left hemisphere), Volume of hippocampal-fissure (left hemisphere), Volume of presubiculum-head (left hemisphere), Volume of CA1-head (left hemisphere), Volume of presubiculum-body (left hemisphere), Volume of parasubiculum (left hemisphere), Volume of molecular-layer-HP-head (left hemisphere), Volume of molecular-layer-HP-body (left hemisphere), Volume of GC-ML-

DG-head (left hemisphere), Volume of CA3-body (left hemisphere), Volume of GC-ML-DG-body (left hemisphere), Volume of CA4-head (left hemisphere), Volume of CA4-body (left hemisphere), Volume of fimbria (left hemisphere), Volume of CA3-head (left hemisphere), Volume of HATA (left hemisphere), Volume of Whole-hippocampal-body (left hemisphere), Volume of Whole-hippocampal-head (left hemisphere), Volume of Whole-hippocampus (left hemisphere), Volume of Hippocampal-tail (right hemisphere), Volume of subiculum-body (right hemisphere), Volume of CA1-body (right hemisphere), Volume of subiculum-head (right hemisphere), Volume of hippocampal-fissure (right hemisphere), Volume of presubiculum-head (right hemisphere), Volume of CA1-head (right hemisphere), Volume of presubiculum-body (right hemisphere), Volume of parasubiculum (right hemisphere), Volume of molecular-layer-HP-head (right hemisphere), Volume of molecular-layer-HP-body (right hemisphere), Volume of GC-ML-DG-head (right hemisphere), Volume of CA3-body (right hemisphere), Volume of GC-ML-DG-body (right hemisphere), Volume of CA4-head (right hemisphere), Volume of CA4-body (right hemisphere), Volume of fimbria (right hemisphere), Volume of CA3-head (right hemisphere), Volume of HATA (right hemisphere), Volume of Whole-hippocampal-body (right hemisphere), Volume of Whole-hippocampal-head (right hemisphere), Volume of Whole-hippocampus (right hemisphere), Volume of MGN (left hemisphere), Volume of LGN (left hemisphere), Volume of PuI (left hemisphere), Volume of PuM (left hemisphere), Volume of L-Sg (left hemisphere), Volume of VPL (left hemisphere), Volume of CM (left hemisphere), Volume of VLa (left hemisphere), Volume of PuA (left hemisphere), Volume of MDm (left hemisphere), Volume of Pf (left hemisphere), Volume of VAmc (left hemisphere), Volume of MDl (left hemisphere), Volume of CeM (left hemisphere), Volume of VA (left hemisphere), Volume of MV(Re) (left hemisphere), Volume of VM (left hemisphere), Volume of CL (left hemisphere), Volume of PuL (left hemisphere), Volume of Pt (left hemisphere), Volume of AV (left hemisphere), Volume of Pc (left hemisphere), Volume of VLp (left hemisphere), Volume of LP (left hemisphere), Volume of LGN (right hemisphere), Volume of MGN (right hemisphere), Volume of PuI (right hemisphere), Volume of PuM (right hemisphere), Volume of L-Sg (right hemisphere), Volume of VPL (right hemisphere), Volume of CM (right hemisphere), Volume of VLa (right hemisphere), Volume of PuA (right hemisphere), Volume of MDm (right hemisphere), Volume of Pf (right hemisphere), Volume of VAmc (right hemisphere), Volume of MDl (right hemisphere), Volume of VA (right hemisphere), Volume of MV(Re) (right hemisphere), Volume of CeM (right hemisphere), Volume of VM (right hemisphere), Volume of PuL (right hemisphere), Volume of CL (right hemisphere), Volume of VLp (right hemisphere), Volume of Pc (right hemisphere), Volume of Pt (right hemisphere), Volume of AV (right hemisphere), Volume of LP (right hemisphere), Volume of LD (left hemisphere), Volume of LD (right hemisphere), Volume of Whole-thalamus (left hemisphere), Volume of Whole-thalamus (right hemisphere), Volume of Medulla (whole brain), Volume of Pons (whole brain), Volume of SCP (whole brain), Volume of Midbrain (whole brain), Volume of Whole-brainstem (whole brain), Mean FA in middle cerebellar peduncle on FA skeleton, Mean FA in pontine crossing tract on FA skeleton, Mean FA in genu of corpus callosum on FA skeleton, Mean FA in body of corpus callosum on FA skeleton, Mean FA in splenium of corpus callosum on FA skeleton, Mean FA in fornix on FA skeleton, Mean FA in corticospinal tract on FA skeleton (right), Mean FA in corticospinal tract on FA skeleton (left), Mean FA in medial lemniscus on FA skeleton (right), Mean FA in medial lemniscus on FA skeleton (left), Mean FA in inferior cerebellar peduncle on FA skeleton (right), Mean FA in inferior cerebellar peduncle on FA skeleton (left), Mean FA in superior cerebellar peduncle on FA skeleton (right), Mean FA in superior cerebellar peduncle on FA skeleton (left), Mean FA in cerebral peduncle on FA skeleton (right), Mean FA in cerebral peduncle on FA skeleton (left), Mean FA in anterior limb of internal capsule on FA skeleton (right), Mean FA in anterior limb of internal capsule on FA skeleton (left), Mean FA in posterior limb of internal capsule on FA skeleton (right), Mean FA in posterior limb of internal capsule on FA skeleton (left), Mean FA in retrolenticular part of internal capsule on FA skeleton (right), Mean FA in retrolenticular part of internal capsule on FA skeleton (left), Mean FA in anterior corona radiata on FA skeleton (right), Mean FA in anterior corona radiata on FA skeleton (left), Mean FA in superior corona radiata on FA skeleton (right), Mean FA in superior corona radiata on FA skeleton (left), Mean FA in posterior corona radiata on FA skeleton (right), Mean FA in posterior corona radiata on FA skeleton (left), Mean FA in posterior thalamic radiation on FA skeleton (right), Mean FA in posterior thalamic radiation on FA skeleton (left), Mean FA in sagittal stratum on FA skeleton (right), Mean FA in sagittal stratum on FA skeleton (left), Mean FA in external capsule on FA skeleton (right), Mean FA in external capsule on FA skeleton (left), Mean FA in cingulum cingulate gyrus on FA skeleton (right), Mean FA in cingulum cingulate gyrus on FA skeleton (left), Mean FA in cingulum hippocampus on FA skeleton (right), Mean FA in cingulum hippocampus on FA skeleton (left), Mean FA in fornix cres+stria terminalis on FA skeleton (right), Mean FA in fornix cres+stria terminalis on FA skeleton (left), Mean FA in superior longitudinal fasciculus on FA skeleton (right), Mean FA in superior longitudinal fasciculus on FA skeleton (left), Mean FA in superior fronto-occipital fasciculus on FA skeleton (right), Mean FA in superior fronto-occipital fasciculus on FA skeleton (left), Mean MD in middle cerebellar peduncle on FA skeleton, Mean MD in pontine crossing tract on FA skeleton, Mean MD in genu of corpus callosum on FA skeleton, Mean MD in body of corpus callosum on FA skeleton, Mean MD in splenium of corpus callosum on FA skeleton, Mean MD in fornix on FA skeleton, Mean MD in corticospinal tract on FA skeleton (right), Mean MD in corticospinal tract on FA skeleton (left), Mean MD in medial lemniscus on FA skeleton (right), Mean MD in medial lemniscus on FA skeleton (left), Mean MD in inferior cerebellar peduncle on FA skeleton (right), Mean MD in inferior cerebellar peduncle on FA skeleton (left), Mean MD in superior cerebellar peduncle on FA skeleton (right), Mean MD in superior cerebellar peduncle on FA skeleton (left), Mean MD in cerebral peduncle on FA skeleton (right), Mean MD in cerebral peduncle on FA skeleton (left), Mean MD in anterior limb of internal capsule on FA skeleton (right), Mean MD in anterior limb of internal capsule on FA skeleton (left), Mean MD in posterior limb of internal capsule on FA skeleton (right), Mean MD in posterior limb of internal capsule on FA skeleton (left), Mean MD in retrolenticular part of internal capsule on FA skeleton (right), Mean MD in retrolenticular part of internal capsule on FA skeleton (left), Mean MD in anterior corona radiata on FA skeleton (right), Mean MD in anterior corona radiata on FA skeleton (left), Mean MD in superior corona radiata on FA skeleton (right), Mean MD in superior corona radiata on FA skeleton (left), Mean MD in posterior corona radiata on FA skeleton (right), Mean MD in posterior corona radiata on FA skeleton (left), Mean MD in posterior thalamic radiation on FA skeleton (right), Mean MD in posterior thalamic radiation on FA skeleton (left), Mean MD in sagittal stratum on FA skeleton (right), Mean MD in sagittal stratum on FA skeleton (left), Mean MD in external capsule on FA skeleton (right), Mean MD in external capsule on FA skeleton (left), Mean MD in cingulum cingulate gyrus on FA skeleton (right), Mean MD in cingulum cingulate gyrus on FA skeleton (left), Mean MD in cingulum hippocampus on FA skeleton (right), Mean MD in cingulum hippocampus on FA skeleton (left), Mean MD in fornix cres+stria terminalis on FA skeleton (right), Mean MD in fornix cres+stria terminalis on FA skeleton (left), Mean MD in superior longitudinal fasciculus on FA skeleton (right), Mean MD in superior longitudinal fasciculus on FA skeleton (left), Mean MD in superior fronto-occipital fasciculus on FA skeleton (right), Mean MD in superior fronto-occipital fasciculus on FA skeleton (left), Mean MD in uncinate fasciculus on FA skeleton (right), Mean MD in uncinate fasciculus on FA skeleton (left), Mean MD in tapetum on FA skeleton (right), Mean MD in tapetum on FA skeleton (left), Resting-state functional connectivity 1, Resting-state functional connectivity 2, Resting-state functional connectivity 3, Resting-state functional connectivity 4, Resting-state functional connectivity 5, Resting-state functional connectivity 6, Resting-state functional connectivity 7, Resting-state functional connectivity 8, Resting-state functional connectivity 9, Resting-state functional connectivity 10, Resting-state functional connectivity 11, Resting-state functional connectivity 12, Resting-state functional connectivity 13, Resting-state functional connectivity 14, Resting-state functional connectivity 15, Resting-state functional connectivity 16, Resting-state functional connectivity 17, Resting-state functional connectivity 18, Resting-state functional connectivity 19, Resting-state functional connectivity 20, Resting-state functional connectivity 21, Resting-state functional connectivity 22, Resting-state functional connectivity 23, Resting-state functional connectivity 24, Resting-state functional connectivity 25, Resting-state functional connectivity 26, Resting-state functional connectivity 27, Resting-state functional connectivity 28, Resting-state functional connectivity 29, Resting-state functional connectivity 30, Resting-state functional connectivity 31, Resting-state functional connectivity 32, Resting-state functional connectivity 33, Resting-state functional connectivity 34, Resting-state functional connectivity 35, Resting-state functional connectivity 36, Resting-state functional connectivity 37, Resting-state functional connectivity 38, Resting-state functional connectivity 39, Resting-state functional connectivity 40, Resting-state functional connectivity 41, Resting-state functional connectivity 42, Resting-state functional connectivity 43, Resting-state functional connectivity 44, Resting-state functional connectivity 45, Resting-state functional connectivity 46, Resting-state functional connectivity 47, Resting-state functional connectivity 48, Resting-state functional connectivity 49, Resting-state functional connectivity 50, Resting-state functional connectivity 51, Resting-state functional connectivity 52, Resting-state functional connectivity 53, Resting-state functional connectivity 54, Resting-state functional connectivity 55, Resting-state functional connectivity 56, Resting-state functional connectivity 57, Resting-state functional connectivity 58, Resting-state functional connectivity 59, Resting-state functional connectivity 60, Resting-state functional connectivity 61, Resting-state functional connectivity 62, Resting-state functional connectivity 63, Resting-state functional connectivity 64, Resting-state functional connectivity 65, Resting-state functional connectivity 66, Resting-state functional connectivity 67, Resting-state functional connectivity 68, Resting-state functional connectivity 69, Resting-state functional connectivity 70, Resting-state functional connectivity 71, Resting-state functional connectivity 72, Resting-state functional connectivity 73, Resting-state functional connectivity 74, Resting-state functional connectivity 75, Resting-state functional connectivity 76, Resting-state functional connectivity 77, Resting-state functional connectivity 78, Resting-state functional connectivity 79, Resting-state functional connectivity 80, Resting-state functional connectivity 81, Resting-state functional connectivity 82, Resting-state functional connectivity 83, Resting-state functional connectivity 84, Resting-state functional connectivity 85, Resting-state functional connectivity 86, Resting-state functional connectivity 87, Resting-state functional connectivity 88, Resting-state functional connectivity 89, Resting-state functional connectivity 90, Resting-state functional connectivity 91, Resting-state functional connectivity 92, Resting-state functional connectivity

93, Resting-state functional connectivity 94, Resting-state functional connectivity 95, Resting-state functional connectivity 96, Resting-state functional connectivity 97, Resting-state functional connectivity 98, Resting-state functional connectivity 99, Resting-state functional connectivity 100, Resting-state functional connectivity 101, Resting-state functional connectivity 102, Resting-state functional connectivity 103, Resting-state functional connectivity 104, Resting-state functional connectivity 105, Resting-state functional connectivity 106, Resting-state functional connectivity 107, Resting-state functional connectivity 108, Resting-state functional connectivity 109, Resting-state functional connectivity 110, Resting-state functional connectivity 111, Resting-state functional connectivity 112, Resting-state functional connectivity 113, Resting-state functional connectivity 114, Resting-state functional connectivity 115, Resting-state functional connectivity 116, Resting-state functional connectivity 117, Resting-state functional connectivity 118, Resting-state functional connectivity 119, Resting-state functional connectivity 120, Resting-state functional connectivity 121, Resting-state functional connectivity 122, Resting-state functional connectivity 123, Resting-state functional connectivity 124, Resting-state functional connectivity 125, Resting-state functional connectivity 126, Resting-state functional connectivity 127, Resting-state functional connectivity 128, Resting-state functional connectivity 129, Resting-state functional connectivity 130, Resting-state functional connectivity 131, Resting-state functional connectivity 132, Resting-state functional connectivity 133, Resting-state functional connectivity 134, Resting-state functional connectivity 135, Resting-state functional connectivity 136, Resting-state functional connectivity 137, Resting-state functional connectivity 138, Resting-state functional connectivity 139, Resting-state functional connectivity 140, Resting-state functional connectivity 141, Resting-state functional connectivity 142, Resting-state functional connectivity 143, Resting-state functional connectivity 144, Resting-state functional connectivity 145, Resting-state functional connectivity 146, Resting-state functional connectivity 147, Resting-state functional connectivity 148, Resting-state functional connectivity 149, Resting-state functional connectivity 150, Resting-state functional connectivity 151, Resting-state functional connectivity 152, Resting-state functional connectivity 153, Resting-state functional connectivity 154, Resting-state functional connectivity 155, Resting-state functional connectivity 156, Resting-state functional connectivity 157, Resting-state functional connectivity 158, Resting-state functional connectivity 159, Resting-state functional connectivity 160, Resting-state functional connectivity 161, Resting-state functional connectivity 162, Resting-state functional connectivity 163, Resting-state functional connectivity 164, Resting-state functional connectivity 165, Resting-state functional connectivity 166, Resting-state functional connectivity 167, Resting-state functional connectivity 168, Resting-state functional connectivity 169, Resting-state functional connectivity 170, Resting-state functional connectivity 171, Resting-state functional connectivity 172, Resting-state functional connectivity 173, Resting-state functional connectivity 174, Resting-state functional connectivity 175, Resting-state functional connectivity 176, Resting-state functional connectivity 177, Resting-state functional connectivity 178, Resting-state functional connectivity 179, Resting-state functional connectivity 180, Resting-state functional connectivity 181, Resting-state functional connectivity 182, Resting-state functional connectivity 183, Resting-state functional connectivity 184, Resting-state functional connectivity 185, Resting-state functional connectivity 186, Resting-state functional connectivity 187, Resting-state functional connectivity 188, Resting-state functional connectivity 189, Resting-state functional connectivity 190, Resting-state functional connectivity 191, Resting-state functional connectivity 192, Resting-state functional connectivity 193, Resting-state functional connectivity 194, Resting-state functional connectivity 195, Resting-state functional connectivity 196, Resting-state functional connectivity 197, Resting-state functional connectivity 198, Resting-state functional connectivity 199, Resting-state functional connectivity 200, Resting-state functional connectivity 201, Resting-state functional connectivity 202, Resting-state functional connectivity 203, Resting-state functional connectivity 204, Resting-state functional connectivity 205, Resting-state functional connectivity 206, Resting-state functional connectivity 207, Resting-state functional connectivity 208, Resting-state functional connectivity 209, Resting-state functional connectivity 210, Resting-state functional connectivity 211, Resting-state functional connectivity 212, Resting-state functional connectivity 213, Resting-state functional connectivity 214, Resting-state functional connectivity 215, Resting-state functional connectivity 216, Resting-state functional connectivity 217, Resting-state functional connectivity 218, Resting-state functional connectivity 219, Resting-state functional connectivity 220, Resting-state functional connectivity 221, Resting-state functional connectivity 222, Resting-state functional connectivity 223, Resting-state functional connectivity 224, Resting-state functional connectivity 225, Resting-state functional connectivity 226, Resting-state functional connectivity 227, Resting-state functional connectivity 228, Resting-state functional connectivity 229, Resting-state functional connectivity 230, Resting-state functional connectivity 231, Resting-state functional connectivity 232, Resting-state functional connectivity 233, Resting-state functional connectivity 234, Resting-state functional connectivity 235, Resting-state functional connectivity 236, Resting-state functional connectivity 237, Resting-state functional connectivity 238, Resting-state functional connectivity 239, Resting-state functional connectivity 240, Resting-state functional connectivity 241, Resting-state functional connectivity 242, Resting-state functional connectivity 243, Resting-state functional connectivity 244, Resting-state functional connectivity 245, Resting-state functional connectivity 246, Resting-state functional connectivity 247, Resting-state functional connectivity 248, Resting-state functional connectivity 249, Resting-state functional connectivity 250, Resting-state functional connectivity 251, Resting-state functional connectivity 252, Resting-state functional connectivity 253, Resting-state functional connectivity 254, Resting-state functional connectivity 255, Resting-state functional connectivity 256, Resting-state functional connectivity 257, Resting-state functional connectivity 258, Resting-state functional connectivity 259, Resting-state functional connectivity 260, Resting-state functional connectivity 261, Resting-state functional connectivity 262, Resting-state functional connectivity 263, Resting-state functional connectivity 264, Resting-state functional connectivity 265, Resting-state functional connectivity 266, Resting-state functional connectivity 267, Resting-state functional connectivity 268, Resting-state functional connectivity 269, Resting-state functional connectivity 270, Resting-state functional connectivity 271, Resting-state functional connectivity 272, Resting-state functional connectivity 273, Resting-state functional connectivity 274, Resting-state functional connectivity 275, Resting-state functional connectivity 276, Resting-state functional connectivity 277, Resting-state functional connectivity 278, Resting-state functional connectivity 279, Resting-state functional connectivity 280, Resting-state functional connectivity 281, Resting-state functional connectivity 282, Resting-state functional connectivity 283, Resting-state functional connectivity 284, Resting-state functional connectivity 285, Resting-state functional connectivity 286, Resting-state functional connectivity 287, Resting-state functional connectivity 288, Resting-state functional connectivity 289, Resting-state functional connectivity 290, Resting-state functional connectivity 291, Resting-state functional connectivity 292, Resting-state functional connectivity 293, Resting-state functional connectivity 294, Resting-state functional connectivity 295, Resting-state functional connectivity 296, Resting-state functional connectivity 297, Resting-state functional connectivity 298, Resting-state functional connectivity 299, Resting-state functional connectivity 300, Resting-state functional connectivity 301, Resting-state functional connectivity 302, Resting-state functional connectivity 303, Resting-state functional connectivity 304, Resting-state functional connectivity 305, Resting-state functional connectivity 306, Resting-state functional connectivity 307, Resting-state functional connectivity 308, Resting-state functional connectivity 309, Resting-state functional connectivity 310, Resting-state functional connectivity 311, Resting-state functional connectivity 312, Resting-state functional connectivity 313, Resting-state functional connectivity 314, Resting-state functional connectivity 315, Resting-state functional connectivity 316, Resting-state functional connectivity 317, Resting-state functional connectivity 318, Resting-state functional connectivity 319, Resting-state functional connectivity 320, Resting-state functional connectivity 321, Resting-state functional connectivity 322, Resting-state functional connectivity 323, Resting-state functional connectivity 324, Resting-state functional connectivity 325, Resting-state functional connectivity 326, Resting-state functional connectivity 327, Resting-state functional connectivity 328, Resting-state functional connectivity 329, Resting-state functional connectivity 330, Resting-state functional connectivity 331, Resting-state functional connectivity 332, Resting-state functional connectivity 333, Resting-state functional connectivity 334, Resting-state functional connectivity 335, Resting-state functional connectivity 336, Resting-state functional connectivity 337, Resting-state functional connectivity 338, Resting-state functional connectivity 339, Resting-state functional connectivity 340, Resting-state functional connectivity 341, Resting-state functional connectivity 342, Resting-state functional connectivity 343, Resting-state functional connectivity 344, Resting-state functional connectivity 345, Resting-state functional connectivity 346, Resting-state functional connectivity 347, Resting-state functional connectivity 348, Resting-state functional connectivity 349, Resting-state functional connectivity 350, Resting-state functional connectivity 351, Resting-state functional connectivity 352, Resting-state functional connectivity 353, Resting-state functional connectivity 354, Resting-state functional connectivity 355, Resting-state functional connectivity 356, Resting-state functional connectivity 357, Resting-state functional connectivity 358, Resting-state functional connectivity 359, Resting-state functional connectivity 360, Resting-state functional connectivity 361, Resting-state functional connectivity 362, Resting-state functional connectivity 363, Resting-state functional connectivity 364, Resting-state functional connectivity 365, Resting-state functional connectivity 366, Resting-state functional connectivity 367, Resting-state functional connectivity 368, Resting-state functional connectivity 369, Resting-state functional connectivity 370, Resting-state functional connectivity 371, Resting-state functional connectivity 372, Resting-state functional connectivity 373, Resting-state functional connectivity 374, Resting-state functional connectivity 375, Resting-state functional connectivity 376, Resting-state functional connectivity 377, Resting-state functional connectivity 378, Resting-state functional connectivity 379, Resting-state functional connectivity 380, Resting-state functional connectivity 381, Resting-state functional connectivity 382, Resting-state functional connectivity 383, Resting-state functional connectivity 384, Resting-state functional connectivity 385, Resting-state functional connectivity 386, Resting-state functional connectivity 387, Resting-state functional connectivity 388, Resting-state functional connectivity 389, Resting-state functional connectivity 390, Resting-state functional connectivity 391, Resting-state functional connectivity 392, Resting-state functional connectivity 393, Resting-state functional connectivity 394, Resting-state functional connectivity 395, Resting-state functional connectivity 396, Resting-state functional connectivity 397, Resting-state functional connectivity 398, Resting-state functional connectivity 399, Resting-state functional connectivity 400, Resting-state functional connectivity 401, Resting-state functional connectivity 402, Resting-state functional connectivity 403, Resting-state functional connectivity 404, Resting-state functional connectivity 405, Resting-state functional connectivity 406, Resting-state functional connectivity 407, Resting-state functional connectivity 408, Resting-state functional connectivity 409, Resting-state functional connectivity 410, Resting-state functional connectivity 411, Resting-state functional connectivity 412, Resting-state functional connectivity 413, Resting-state functional connectivity 414, Resting-state functional connectivity 415, Resting-state functional connectivity 416, Resting-state functional connectivity 417, Resting-state functional connectivity 418, Resting-state functional connectivity 419, Resting-state functional connectivity 420, Resting-state functional connectivity 421, Resting-state functional connectivity 422, Resting-state functional connectivity 423, Resting-state functional connectivity 424, Resting-state functional connectivity 425, Resting-state functional connectivity 426, Resting-state functional connectivity 427, Resting-state functional connectivity 428, Resting-state functional connectivity 429, Resting-state functional connectivity 430, Resting-state functional connectivity 431, Resting-state functional connectivity 432, Resting-state functional connectivity 433, Resting-state functional connectivity 434, Resting-state functional connectivity 435, Resting-state functional connectivity 436, Resting-state functional connectivity 437, Resting-state functional connectivity 438, Resting-state functional connectivity 439, Resting-state functional connectivity 440, Resting-state functional connectivity 441, Resting-state functional connectivity 442, Resting-state functional connectivity 443, Resting-state functional connectivity 444, Resting-state functional connectivity 445, Resting-state functional connectivity 446, Resting-state functional connectivity 447, Resting-state functional connectivity 448, Resting-state functional connectivity 449, Resting-state functional connectivity 450, Resting-state functional connectivity 451, Resting-state functional connectivity 452, Resting-state functional connectivity 453, Resting-state functional connectivity 454, Resting-state functional connectivity 455, Resting-state functional connectivity 456, Resting-state functional connectivity 457, Resting-state functional connectivity 458, Resting-state functional connectivity 459, Resting-state functional connectivity 460, Resting-state functional connectivity 461, Resting-state functional connectivity 462, Resting-state functional connectivity 463, Resting-state functional connectivity 464, Resting-state functional connectivity 465, Resting-state functional connectivity 466, Resting-state functional connectivity 467, Resting-state functional connectivity 468, Resting-state functional connectivity 469, Resting-state functional connectivity 470, Resting-state functional connectivity 471, Resting-state functional connectivity 472, Resting-state functional connectivity 473, Resting-state functional connectivity 474, Resting-state functional connectivity 475, Resting-state functional connectivity 476, Resting-state functional connectivity 477, Resting-state functional connectivity 478, Resting-state functional connectivity 479, Resting-state functional connectivity 480, Resting-state functional connectivity 481, Resting-state functional connectivity 482, Resting-state functional connectivity 483, Resting-state functional connectivity 484, Resting-state functional connectivity 485, Resting-state functional connectivity 486, Resting-state functional connectivity 487, Resting-state functional connectivity 488, Resting-state functional connectivity 489, Resting-state functional connectivity 490, Resting-state functional connectivity 491, Resting-state functional connectivity 492, Resting-state functional connectivity 493, Resting-state functional connectivity 494, Resting-state functional connectivity 495, Resting-state functional connectivity 496, Resting-state functional connectivity 497, Resting-state functional connectivity

498, Resting-state functional connectivity 499, Resting-state functional connectivity 500, Resting-state functional connectivity 501, Resting-state functional connectivity 502, Resting-state functional connectivity 503, Resting-state functional connectivity 504, Resting-state functional connectivity 505, Resting-state functional connectivity 506, Resting-state functional connectivity 507, Resting-state functional connectivity 508, Resting-state functional connectivity 509, Resting-state functional connectivity 510, Resting-state functional connectivity 511, Resting-state functional connectivity 512, Resting-state functional connectivity 513, Resting-state functional connectivity 514, Resting-state functional connectivity 515, Resting-state functional connectivity 516, Resting-state functional connectivity 517, Resting-state functional connectivity 518, Resting-state functional connectivity 519, Resting-state functional connectivity 520, Resting-state functional connectivity 521, Resting-state functional connectivity 522, Resting-state functional connectivity 523, Resting-state functional connectivity 524, Resting-state functional connectivity 525, Resting-state functional connectivity 526, Resting-state functional connectivity 527, Resting-state functional connectivity 528, Resting-state functional connectivity 529, Resting-state functional connectivity 530, Resting-state functional connectivity 531, Resting-state functional connectivity 532, Resting-state functional connectivity 533, Resting-state functional connectivity 534, Resting-state functional connectivity 535, Resting-state functional connectivity 536, Resting-state functional connectivity 537, Resting-state functional connectivity 538, Resting-state functional connectivity 539, Resting-state functional connectivity 540, Resting-state functional connectivity 541, Resting-state functional connectivity 542, Resting-state functional connectivity 543, Resting-state functional connectivity 544, Resting-state functional connectivity 545, Resting-state functional connectivity 546, Resting-state functional connectivity 547, Resting-state functional connectivity 548, Resting-state functional connectivity 549, Resting-state functional connectivity 550, Resting-state functional connectivity 551, Resting-state functional connectivity 552, Resting-state functional connectivity 553, Resting-state functional connectivity 554, Resting-state functional connectivity 555, Resting-state functional connectivity 556, Resting-state functional connectivity 557, Resting-state functional connectivity 558, Resting-state functional connectivity 559, Resting-state functional connectivity 560, Resting-state functional connectivity 561, Resting-state functional connectivity 562, Resting-state functional connectivity 563, Resting-state functional connectivity 564, Resting-state functional connectivity 565, Resting-state functional connectivity 566, Resting-state functional connectivity 567, Resting-state functional connectivity 568, Resting-state functional connectivity 569, Resting-state functional connectivity 570, Resting-state functional connectivity 571, Resting-state functional connectivity 572, Resting-state functional connectivity 573, Resting-state functional connectivity 574, Resting-state functional connectivity 575, Resting-state functional connectivity 576, Resting-state functional connectivity 577, Resting-state functional connectivity 578, Resting-state functional connectivity 579, Resting-state functional connectivity 580, Resting-state functional connectivity 581, Resting-state functional connectivity 582, Resting-state functional connectivity 583, Resting-state functional connectivity 584, Resting-state functional connectivity 585, Resting-state functional connectivity 586, Resting-state functional connectivity 587, Resting-state functional connectivity 588, Resting-state functional connectivity 589, Resting-state functional connectivity 590, Resting-state functional connectivity 591, Resting-state functional connectivity 592, Resting-state functional connectivity 593, Resting-state functional connectivity 594, Resting-state functional connectivity 595, Resting-state functional connectivity 596, Resting-state functional connectivity 597, Resting-state functional connectivity 598, Resting-state functional connectivity 599,

Resting-state functional connectivity 600, Resting-state functional connectivity 601, Resting-state functional connectivity 602, Resting-state functional connectivity 603, Resting-state functional connectivity 604, Resting-state functional connectivity 605, Resting-state functional connectivity 606, Resting-state functional connectivity 607, Resting-state functional connectivity 608, Resting-state functional connectivity 609, Resting-state functional connectivity 610, Resting-state functional connectivity 611, Resting-state functional connectivity 612, Resting-state functional connectivity 613, Resting-state functional connectivity 614, Resting-state functional connectivity 615, Resting-state functional connectivity 616, Resting-state functional connectivity 617, Resting-state functional connectivity 618, Resting-state functional connectivity 619, Resting-state functional connectivity 620, Resting-state functional connectivity 621, Resting-state functional connectivity 622, Resting-state functional connectivity 623, Resting-state functional connectivity 624, Resting-state functional connectivity 625, Resting-state functional connectivity 626, Resting-state functional connectivity 627, Resting-state functional connectivity 628, Resting-state functional connectivity 629, Resting-state functional connectivity 630, Resting-state functional connectivity 631, Resting-state functional connectivity 632, Resting-state functional connectivity 633, Resting-state functional connectivity 634, Resting-state functional connectivity 635, Resting-state functional connectivity 636, Resting-state functional connectivity 637, Resting-state functional connectivity 638, Resting-state functional connectivity 639, Resting-state functional connectivity 640, Resting-state functional connectivity 641, Resting-state functional connectivity 642, Resting-state functional connectivity 643, Resting-state functional connectivity 644, Resting-state functional connectivity 645, Resting-state functional connectivity 646, Resting-state functional connectivity 647, Resting-state functional connectivity 648, Resting-state functional connectivity 649, Resting-state functional connectivity 650, Resting-state functional connectivity 651, Resting-state functional connectivity 652, Resting-state functional connectivity 653, Resting-state functional connectivity 654, Resting-state functional connectivity 655, Resting-state functional connectivity 656, Resting-state functional connectivity 657, Resting-state functional connectivity 658, Resting-state functional connectivity 659, Resting-state functional connectivity 660, Resting-state functional connectivity 661, Resting-state functional connectivity 662, Resting-state functional connectivity 663, Resting-state functional connectivity 664, Resting-state functional connectivity 665, Resting-state functional connectivity 666, Resting-state functional connectivity 667, Resting-state functional connectivity 668, Resting-state functional connectivity 669, Resting-state functional connectivity 670, Resting-state functional connectivity 671, Resting-state functional connectivity 672, Resting-state functional connectivity 673, Resting-state functional connectivity 674, Resting-state functional connectivity 675, Resting-state functional connectivity 676, Resting-state functional connectivity 677, Resting-state functional connectivity 678, Resting-state functional connectivity 679, Resting-state functional connectivity 680, Resting-state functional connectivity 681, Resting-state functional connectivity 682, Resting-state functional connectivity 683, Resting-state functional connectivity 684, Resting-state functional connectivity 685, Resting-state functional connectivity 686, Resting-state functional connectivity 687, Resting-state functional connectivity 688, Resting-state functional connectivity 689, Resting-state functional connectivity 690, Resting-state functional connectivity 691, Resting-state functional connectivity 692, Resting-state functional connectivity 693, Resting-state functional connectivity 694, Resting-state functional connectivity 695, Resting-state functional connectivity 696, Resting-state functional connectivity 697, Resting-state functional connectivity 698, Resting-state functional connectivity 699, Resting-state functional connectivity 700, Resting-state functional connectivity 701, Resting-state functional connectivity 702, Resting-state functional connectivity 703, Resting-state functional connectivity 704, Resting-state functional connectivity 705, Resting-state functional connectivity 706, Resting-state functional connectivity 707, Resting-state functional connectivity 708, Resting-state functional connectivity 709, Resting-state functional connectivity 710, Resting-state functional connectivity 711, Resting-state functional connectivity 712, Resting-state functional connectivity 713, Resting-state functional connectivity 714, Resting-state functional connectivity 715, Resting-state functional connectivity 716, Resting-state functional connectivity 717, Resting-state functional connectivity 718, Resting-state functional connectivity 719, Resting-state functional connectivity 720, Resting-state functional connectivity 721, Resting-state functional connectivity 722, Resting-state functional connectivity 723, Resting-state functional connectivity 724, Resting-state functional connectivity 725, Resting-state functional connectivity 726, Resting-state functional connectivity 727, Resting-state functional connectivity 728, Resting-state functional connectivity 729, Resting-state functional connectivity 730, Resting-state functional connectivity 731, Resting-state functional connectivity 732, Resting-state functional connectivity 733, Resting-state functional connectivity 734, Resting-state functional connectivity 735, Resting-state functional connectivity 736, Resting-state functional connectivity 737, Resting-state functional connectivity 738, Resting-state functional connectivity 739, Resting-state functional connectivity 740, Resting-state functional connectivity 741, Resting-state functional connectivity 742, Resting-state functional connectivity 743, Resting-state functional connectivity 744, Resting-state functional connectivity 745, Resting-state functional connectivity 746, Resting-state functional connectivity 747, Resting-state functional connectivity 748, Resting-state functional connectivity 749, Resting-state functional connectivity 750, Resting-state functional connectivity 751, Resting-state functional connectivity 752, Resting-state functional connectivity 753, Resting-state functional connectivity 754, Resting-state functional connectivity 755, Resting-state functional connectivity 756, Resting-state functional connectivity 757, Resting-state functional connectivity 758, Resting-state functional connectivity 759, Resting-state functional connectivity 760, Resting-state functional connectivity 761, Resting-state functional connectivity 762, Resting-state functional connectivity 763, Resting-state functional connectivity 764, Resting-state functional connectivity 765, Resting-state functional connectivity 766, Resting-state functional connectivity 767, Resting-state functional connectivity 768, Resting-state functional connectivity 769, Resting-state functional connectivity 770, Resting-state functional connectivity 771, Resting-state functional connectivity 772, Resting-state functional connectivity 773, Resting-state functional connectivity 774, Resting-state functional connectivity 775, Resting-state functional connectivity 776, Resting-state functional connectivity 777, Resting-state functional connectivity 778, Resting-state functional connectivity 779, Resting-state functional connectivity 780, Resting-state functional connectivity 781, Resting-state functional connectivity 782, Resting-state functional connectivity 783, Resting-state functional connectivity 784, Resting-state functional connectivity 785, Resting-state functional connectivity 786, Resting-state functional connectivity 787, Resting-state functional connectivity 788, Resting-state functional connectivity 789, Resting-state functional connectivity 790, Resting-state functional connectivity 791, Resting-state functional connectivity 792, Resting-state functional connectivity 793, Resting-state functional connectivity 794, Resting-state functional connectivity 795, Resting-state functional connectivity 796, Resting-state functional connectivity 797, Resting-state functional connectivity 798, Resting-state functional connectivity 799, Resting-state functional connectivity 800, Resting-state functional connectivity 801, Resting-state functional connectivity 802, Resting-state functional connectivity 803, Resting-state functional connectivity 804, Resting-state functional connectivity 805, Resting-state functional connectivity 806, Resting-state functional connectivity 807, Resting-state functional connectivity 808, Resting-state functional connectivity 809, Resting-state functional connectivity 810, Resting-state functional connectivity 811, Resting-state functional connectivity 812, Resting-state functional connectivity 813, Resting-state functional connectivity 814, Resting-state functional connectivity 815, Resting-state functional connectivity 816, Resting-state functional connectivity 817, Resting-state functional connectivity 818, Resting-state functional connectivity 819, Resting-state functional connectivity 820, Resting-state functional connectivity 821, Resting-state functional connectivity 822, Resting-state functional connectivity 823, Resting-state functional connectivity 824, Resting-state functional connectivity 825, Resting-state functional connectivity 826, Resting-state functional connectivity 827, Resting-state functional connectivity 828, Resting-state functional connectivity 829, Resting-state functional connectivity 830, Resting-state functional connectivity 831, Resting-state functional connectivity 832, Resting-state functional connectivity 833, Resting-state functional connectivity 834, Resting-state functional connectivity 835, Resting-state functional connectivity 836, Resting-state functional connectivity 837, Resting-state functional connectivity 838, Resting-state functional connectivity 839, Resting-state functional connectivity 840, Resting-state functional connectivity 841, Resting-state functional connectivity 842, Resting-state functional connectivity 843, Resting-state functional connectivity 844, Resting-state functional connectivity 845, Resting-state functional connectivity 846, Resting-state functional connectivity 847, Resting-state functional connectivity 848, Resting-state functional connectivity 849, Resting-state functional connectivity 850, Resting-state functional connectivity 851, Resting-state functional connectivity 852, Resting-state functional connectivity 853, Resting-state functional connectivity 854, Resting-state functional connectivity 855, Resting-state functional connectivity 856, Resting-state functional connectivity 857, Resting-state functional connectivity 858, Resting-state functional connectivity 859, Resting-state functional connectivity 860, Resting-state functional connectivity 861, Resting-state functional connectivity 862, Resting-state functional connectivity 863, Resting-state functional connectivity 864, Resting-state functional connectivity 865, Resting-state functional connectivity 866, Resting-state functional connectivity 867, Resting-state functional connectivity 868, Resting-state functional connectivity 869, Resting-state functional connectivity 870, Resting-state functional connectivity 871, Resting-state functional connectivity 872, Resting-state functional connectivity 873, Resting-state functional connectivity 874, Resting-state functional connectivity 875, Resting-state functional connectivity 876, Resting-state functional connectivity 877, Resting-state functional connectivity 878, Resting-state functional connectivity 879, Resting-state functional connectivity 880, Resting-state functional connectivity 881, Resting-state functional connectivity 882, Resting-state functional connectivity 883, Resting-state functional connectivity 884, Resting-state functional connectivity 885, Resting-state functional connectivity 886, Resting-state functional connectivity 887, Resting-state functional connectivity 888, Resting-state functional connectivity 889, Resting-state functional connectivity 890, Resting-state functional connectivity 891, Resting-state functional connectivity 892, Resting-state functional connectivity 893, Resting-state functional connectivity 894, Resting-state functional connectivity 895, Resting-state functional connectivity 896, Resting-state functional connectivity 897, Resting-state functional connectivity 898, Resting-state functional connectivity 899, Resting-state functional connectivity 900, Resting-state functional connectivity 901, Resting-state functional connectivity 902, Resting-state functional connectivity 903, Resting-state functional connectivity 904, Resting-state functional connectivity 905, Resting-state functional connectivity 906, Resting-state functional connectivity 907, Resting-state functional connectivity 908, Resting-state functional connectivity 909, Resting-state functional connectivity 910, Resting-state functional connectivity 911, Resting-state functional connectivity 912, Resting-state functional connectivity 913, Resting-state functional connectivity 914, Resting-state functional connectivity 915, Resting-state functional connectivity 916, Resting-state functional connectivity 917, Resting-state functional connectivity 918, Resting-state functional connectivity 919, Resting-state functional connectivity 920, Resting-state functional connectivity 921, Resting-state functional connectivity 922, Resting-state functional connectivity 923, Resting-state functional connectivity 924, Resting-state functional connectivity 925, Resting-state functional connectivity 926, Resting-state functional connectivity 927, Resting-state functional connectivity 928, Resting-state functional connectivity 929, Resting-state functional connectivity 930, Resting-state functional connectivity 931, Resting-state functional connectivity 932, Resting-state functional connectivity 933, Resting-state functional connectivity 934, Resting-state functional connectivity 935, Resting-state functional connectivity 936, Resting-state functional connectivity 937, Resting-state functional connectivity 938, Resting-state functional connectivity 939, Resting-state functional connectivity 940, Resting-state functional connectivity 941, Resting-state functional connectivity 942, Resting-state functional connectivity 943, Resting-state functional connectivity 944, Resting-state functional connectivity 945, Resting-state functional connectivity 946, Resting-state functional connectivity 947, Resting-state functional connectivity 948, Resting-state functional connectivity 949, Resting-state functional connectivity 950, Resting-state functional connectivity 951, Resting-state functional connectivity 952, Resting-state functional connectivity 953, Resting-state functional connectivity 954, Resting-state functional connectivity 955, Resting-state functional connectivity 956, Resting-state functional connectivity 957, Resting-state functional connectivity 958, Resting-state functional connectivity 959, Resting-state functional connectivity 960, Resting-state functional connectivity 961, Resting-state functional connectivity 962, Resting-state functional connectivity 963, Resting-state functional connectivity 964, Resting-state functional connectivity 965, Resting-state functional connectivity 966, Resting-state functional connectivity 967, Resting-state functional connectivity 968, Resting-state functional connectivity 969, Resting-state functional connectivity 970, Resting-state functional connectivity 971, Resting-state functional connectivity 972, Resting-state functional connectivity 973, Resting-state functional connectivity 974, Resting-state functional connectivity 975, Resting-state functional connectivity 976, Resting-state functional connectivity 977, Resting-state functional connectivity 978, Resting-state functional connectivity 979, Resting-state functional connectivity 980, Resting-state functional connectivity 981, Resting-state functional connectivity 982, Resting-state functional connectivity 983, Resting-state functional connectivity 984, Resting-state functional connectivity 985, Resting-state functional connectivity 986, Resting-state functional connectivity 987, Resting-state functional connectivity 988, Resting-state functional connectivity 989, Resting-state functional connectivity 990, Resting-state functional connectivity 991, Resting-state functional connectivity 992, Resting-state functional connectivity 993, Resting-state functional connectivity 994, Resting-state functional connectivity 995, Resting-state functional connectivity 996, Resting-state functional connectivity 997, Resting-state functional connectivity 998, Resting-state functional connectivity 999, Resting-state functional connectivity 1000, Resting-state functional connectivity 1001, Resting-state functional connectivity 1002, Resting-state functional connectivity 1003, Resting-state functional

connectivity 1004, Resting-state functional connectivity 1005, Resting-state functional connectivity 1006, Resting-state functional connectivity 1007, Resting-state functional connectivity 1008, Resting-state functional connectivity 1009, Resting-state functional connectivity 1010, Resting-state functional connectivity 1011, Resting-state functional connectivity 1012, Resting-state functional connectivity 1013, Resting-state functional connectivity 1014, Resting-state functional connectivity 1015, Resting-state functional connectivity 1016, Resting-state functional connectivity 1017, Resting-state functional connectivity 1018, Resting-state functional connectivity 1019, Resting-state functional connectivity 1020, Resting-state functional connectivity 1021, Resting-state functional connectivity 1022, Resting-state functional connectivity 1023, Resting-state functional connectivity 1024, Resting-state functional connectivity 1025, Resting-state functional connectivity 1026, Resting-state functional connectivity 1027, Resting-state functional connectivity 1028, Resting-state functional connectivity 1029, Resting-state functional connectivity 1030, Resting-state functional connectivity 1031, Resting-state functional connectivity 1032, Resting-state functional connectivity 1033, Resting-state functional connectivity 1034, Resting-state functional connectivity 1035, Resting-state functional connectivity 1036, Resting-state functional connectivity 1037, Resting-state functional connectivity 1038, Resting-state functional connectivity 1039, Resting-state functional connectivity 1040, Resting-state functional connectivity 1041, Resting-state functional connectivity 1042, Resting-state functional connectivity 1043, Resting-state functional connectivity 1044, Resting-state functional connectivity 1045, Resting-state functional connectivity 1046, Resting-state functional connectivity 1047, Resting-state functional connectivity 1048, Resting-state functional connectivity 1049, Resting-state functional connectivity 1050, Resting-state functional connectivity 1051, Resting-state functional connectivity 1052, Resting-state functional connectivity 1053, Resting-state functional connectivity 1054, Resting-state functional connectivity 1055, Resting-state functional connectivity 1056, Resting-state functional connectivity 1057, Resting-state functional connectivity 1058, Resting-state functional connectivity 1059, Resting-state functional connectivity 1060, Resting-state functional connectivity 1061, Resting-state functional connectivity 1062, Resting-state functional connectivity 1063, Resting-state functional connectivity 1064, Resting-state functional connectivity 1065, Resting-state functional connectivity 1066, Resting-state functional connectivity 1067, Resting-state functional connectivity 1068, Resting-state functional connectivity 1069, Resting-state functional connectivity 1070, Resting-state functional connectivity 1071, Resting-state functional connectivity 1072, Resting-state functional connectivity 1073, Resting-state functional connectivity 1074, Resting-state functional connectivity 1075, Resting-state functional connectivity 1076, Resting-state functional connectivity 1077, Resting-state functional connectivity 1078, Resting-state functional connectivity 1079, Resting-state functional connectivity 1080, Resting-state functional connectivity 1081, Resting-state functional connectivity 1082, Resting-state functional connectivity 1083, Resting-state functional connectivity 1084, Resting-state functional connectivity 1085, Resting-state functional connectivity 1086, Resting-state functional connectivity 1087, Resting-state functional connectivity 1088, Resting-state functional connectivity 1089, Resting-state functional connectivity 1090, Resting-state functional connectivity 1091, Resting-state functional connectivity 1092, Resting-state functional connectivity 1093, Resting-state functional connectivity 1094, Resting-state functional connectivity 1095, Resting-state functional

connectivity 1096, Resting-state functional connectivity 1097, Resting-state functional connectivity 1098, Resting-state functional connectivity 1099, Resting-state functional connectivity 1100, Resting-state functional connectivity 1101, Resting-state functional connectivity 1102, Resting-state functional connectivity 1103, Resting-state functional connectivity 1104, Resting-state functional connectivity 1105, Resting-state functional connectivity 1106, Resting-state functional connectivity 1107, Resting-state functional connectivity 1108, Resting-state functional connectivity 1109, Resting-state functional connectivity 1110, Resting-state functional connectivity 1111, Resting-state functional connectivity 1112, Resting-state functional connectivity 1113, Resting-state functional connectivity 1114, Resting-state functional connectivity 1115, Resting-state functional connectivity 1116, Resting-state functional connectivity 1117, Resting-state functional connectivity 1118, Resting-state functional connectivity 1119, Resting-state functional connectivity 1120, Resting-state functional connectivity 1121, Resting-state functional connectivity 1122, Resting-state functional connectivity 1123, Resting-state functional connectivity 1124, Resting-state functional connectivity 1125, Resting-state functional connectivity 1126, Resting-state functional connectivity 1127, Resting-state functional connectivity 1128, Resting-state functional connectivity 1129, Resting-state functional connectivity 1130, Resting-state functional connectivity 1131, Resting-state functional connectivity 1132, Resting-state functional connectivity 1133, Resting-state functional connectivity 1134, Resting-state functional connectivity 1135, Resting-state functional connectivity 1136, Resting-state functional connectivity 1137, Resting-state functional connectivity 1138, Resting-state functional connectivity 1139, Resting-state functional connectivity 1140, Resting-state functional connectivity 1141, Resting-state functional connectivity 1142, Resting-state functional connectivity 1143, Resting-state functional connectivity 1144, Resting-state functional connectivity 1145, Resting-state functional connectivity 1146, Resting-state functional connectivity 1147, Resting-state functional connectivity 1148, Resting-state functional connectivity 1149, Resting-state functional connectivity 1150, Resting-state functional connectivity 1151, Resting-state functional connectivity 1152, Resting-state functional connectivity 1153, Resting-state functional connectivity 1154, Resting-state functional connectivity 1155, Resting-state functional connectivity 1156, Resting-state functional connectivity 1157, Resting-state functional connectivity 1158, Resting-state functional connectivity 1159, Resting-state functional connectivity 1160, Resting-state functional connectivity 1161, Resting-state functional connectivity 1162, Resting-state functional connectivity 1163, Resting-state functional connectivity 1164, Resting-state functional connectivity 1165, Resting-state functional connectivity 1166, Resting-state functional connectivity 1167, Resting-state functional connectivity 1168, Resting-state functional connectivity 1169, Resting-state functional connectivity 1170, Resting-state functional connectivity 1171, Resting-state functional connectivity 1172, Resting-state functional connectivity 1173, Resting-state functional connectivity 1174, Resting-state functional connectivity 1175, Resting-state functional connectivity 1176, Resting-state functional connectivity 1177, Resting-state functional connectivity 1178, Resting-state functional connectivity 1179, Resting-state functional connectivity 1180, Resting-state functional connectivity 1181, Resting-state functional connectivity 1182, Resting-state functional connectivity 1183, Resting-state functional connectivity 1184, Resting-state functional connectivity 1185, Resting-state functional connectivity 1186, Resting-state functional connectivity 1187, Resting-state functional

connectivity 1188, Resting-state functional connectivity 1189, Resting-state functional connectivity 1190, Resting-state functional connectivity 1191, Resting-state functional connectivity 1192, Resting-state functional connectivity 1193, Resting-state functional connectivity 1194, Resting-state functional connectivity 1195, Resting-state functional connectivity 1196, Resting-state functional connectivity 1197, Resting-state functional connectivity 1198, Resting-state functional connectivity 1199, Resting-state functional connectivity 1200, Resting-state functional connectivity 1201, Resting-state functional connectivity 1202, Resting-state functional connectivity 1203, Resting-state functional connectivity 1204, Resting-state functional connectivity 1205, Resting-state functional connectivity 1206, Resting-state functional connectivity 1207, Resting-state functional connectivity 1208, Resting-state functional connectivity 1209, Resting-state functional connectivity 1210, Resting-state functional connectivity 1211, Resting-state functional connectivity 1212, Resting-state functional connectivity 1213, Resting-state functional connectivity 1214, Resting-state functional connectivity 1215, Resting-state functional connectivity 1216, Resting-state functional connectivity 1217, Resting-state functional connectivity 1218, Resting-state functional connectivity 1219, Resting-state functional connectivity 1220, Resting-state functional connectivity 1221, Resting-state functional connectivity 1222, Resting-state functional connectivity 1223, Resting-state functional connectivity 1224, Resting-state functional connectivity 1225, Resting-state functional connectivity 1226, Resting-state functional connectivity 1227, Resting-state functional connectivity 1228, Resting-state functional connectivity 1229, Resting-state functional connectivity 1230, Resting-state functional connectivity 1231, Resting-state functional connectivity 1232, Resting-state functional connectivity 1233, Resting-state functional connectivity 1234, Resting-state functional connectivity 1235, Resting-state functional connectivity 1236, Resting-state functional connectivity 1237, Resting-state functional connectivity 1238, Resting-state functional connectivity 1239, Resting-state functional connectivity 1240, Resting-state functional connectivity 1241, Resting-state functional connectivity 1242, Resting-state functional connectivity 1243, Resting-state functional connectivity 1244, Resting-state functional connectivity 1245, Resting-state functional connectivity 1246, Resting-state functional connectivity 1247, Resting-state functional connectivity 1248, Resting-state functional connectivity 1249, Resting-state functional connectivity 1250, Resting-state functional connectivity 1251, Resting-state functional connectivity 1252, Resting-state functional connectivity 1253, Resting-state functional connectivity 1254, Resting-state functional connectivity 1255, Resting-state functional connectivity 1256, Resting-state functional connectivity 1257, Resting-state functional connectivity 1258, Resting-state functional connectivity 1259, Resting-state functional connectivity 1260, Resting-state functional connectivity 1261, Resting-state functional connectivity 1262, Resting-state functional connectivity 1263, Resting-state functional connectivity 1264, Resting-state functional connectivity 1265, Resting-state functional connectivity 1266, Resting-state functional connectivity 1267, Resting-state functional connectivity 1268, Resting-state functional connectivity 1269, Resting-state functional connectivity 1270, Resting-state functional connectivity 1271, Resting-state functional connectivity 1272, Resting-state functional connectivity 1273, Resting-state functional connectivity 1274, Resting-state functional connectivity 1275, Resting-state functional connectivity 1276, Resting-state functional connectivity 1277, Resting-state functional connectivity 1278, Resting-state functional connectivity 1279, Resting-state functional

connectivity 1280, Resting-state functional connectivity 1281, Resting-state functional connectivity 1282, Resting-state functional connectivity 1283, Resting-state functional connectivity 1284, Resting-state functional connectivity 1285, Resting-state functional connectivity 1286, Resting-state functional connectivity 1287, Resting-state functional connectivity 1288, Resting-state functional connectivity 1289, Resting-state functional connectivity 1290, Resting-state functional connectivity 1291, Resting-state functional connectivity 1292, Resting-state functional connectivity 1293, Resting-state functional connectivity 1294, Resting-state functional connectivity 1295, Resting-state functional connectivity 1296, Resting-state functional connectivity 1297, Resting-state functional connectivity 1298, Resting-state functional connectivity 1299, Resting-state functional connectivity 1300, Resting-state functional connectivity 1301, Resting-state functional connectivity 1302, Resting-state functional connectivity 1303, Resting-state functional connectivity 1304, Resting-state functional connectivity 1305, Resting-state functional connectivity 1306, Resting-state functional connectivity 1307, Resting-state functional connectivity 1308, Resting-state functional connectivity 1309, Resting-state functional connectivity 1310, Resting-state functional connectivity 1311, Resting-state functional connectivity 1312, Resting-state functional connectivity 1313, Resting-state functional connectivity 1314, Resting-state functional connectivity 1315, Resting-state functional connectivity 1316, Resting-state functional connectivity 1317, Resting-state functional connectivity 1318, Resting-state functional connectivity 1319, Resting-state functional connectivity 1320, Resting-state functional connectivity 1321, Resting-state functional connectivity 1322, Resting-state functional connectivity 1323, Resting-state functional connectivity 1324, Resting-state functional connectivity 1325, Resting-state functional connectivity 1326, Resting-state functional connectivity 1327, Resting-state functional connectivity 1328, Resting-state functional connectivity 1329, Resting-state functional connectivity 1330, Resting-state functional connectivity 1331, Resting-state functional connectivity 1332, Resting-state functional connectivity 1333, Resting-state functional connectivity 1334, Resting-state functional connectivity 1335, Resting-state functional connectivity 1336, Resting-state functional connectivity 1337, Resting-state functional connectivity 1338, Resting-state functional connectivity 1339, Resting-state functional connectivity 1340, Resting-state functional connectivity 1341, Resting-state functional connectivity 1342, Resting-state functional connectivity 1343, Resting-state functional connectivity 1344, Resting-state functional connectivity 1345, Resting-state functional connectivity 1346, Resting-state functional connectivity 1347, Resting-state functional connectivity 1348, Resting-state functional connectivity 1349, Resting-state functional connectivity 1350, Resting-state functional connectivity 1351, Resting-state functional connectivity 1352, Resting-state functional connectivity 1353, Resting-state functional connectivity 1354, Resting-state functional connectivity 1355, Resting-state functional connectivity 1356, Resting-state functional connectivity 1357, Resting-state functional connectivity 1358, Resting-state functional connectivity 1359, Resting-state functional connectivity 1360, Resting-state functional connectivity 1361, Resting-state functional connectivity 1362, Resting-state functional connectivity 1363, Resting-state functional connectivity 1364, Resting-state functional connectivity 1365, Resting-state functional connectivity 1366, Resting-state functional connectivity 1367, Resting-state functional connectivity 1368, Resting-state functional connectivity 1369, Resting-state functional connectivity 1370, Resting-state functional connectivity 1371, Resting-state functional

connectivity 1372, Resting-state functional connectivity 1373, Resting-state functional connectivity 1374, Resting-state functional connectivity 1375, Resting-state functional connectivity 1376, Resting-state functional connectivity 1377, Resting-state functional connectivity 1378, Resting-state functional connectivity 1379, Resting-state functional connectivity 1380, Resting-state functional connectivity 1381, Resting-state functional connectivity 1382, Resting-state functional connectivity 1383, Resting-state functional connectivity 1384, Resting-state functional connectivity 1385, Resting-state functional connectivity 1386, Resting-state functional connectivity 1387, Resting-state functional connectivity 1388, Resting-state functional connectivity 1389, Resting-state functional connectivity 1390, Resting-state functional connectivity 1391, Resting-state functional connectivity 1392, Resting-state functional connectivity 1393, Resting-state functional connectivity 1394, Resting-state functional connectivity 1395, Resting-state functional connectivity 1396, Resting-state functional connectivity 1397, Resting-state functional connectivity 1398, Resting-state functional connectivity 1399, Resting-state functional connectivity 1400, Resting-state functional connectivity 1401, Resting-state functional connectivity 1402, Resting-state functional connectivity 1403, Resting-state functional connectivity 1404, Resting-state functional connectivity 1405, Resting-state functional connectivity 1406, Resting-state functional connectivity 1407, Resting-state functional connectivity 1408, Resting-state functional connectivity 1409, Resting-state functional connectivity 1410, Resting-state functional connectivity 1411, Resting-state functional connectivity 1412, Resting-state functional connectivity 1413, Resting-state functional connectivity 1414, Resting-state functional connectivity 1415, Resting-state functional connectivity 1416, Resting-state functional connectivity 1417, Resting-state functional connectivity 1418, Resting-state functional connectivity 1419, Resting-state functional connectivity 1420, Resting-state functional connectivity 1421, Resting-state functional connectivity 1422, Resting-state functional connectivity 1423, Resting-state functional connectivity 1424, Resting-state functional connectivity 1425, Resting-state functional connectivity 1426, Resting-state functional connectivity 1427, Resting-state functional connectivity 1428, Resting-state functional connectivity 1429, Resting-state functional connectivity 1430, Resting-state functional connectivity 1431, Resting-state functional connectivity 1432, Resting-state functional connectivity 1433, Resting-state functional connectivity 1434, Resting-state functional connectivity 1435, Resting-state functional connectivity 1436, Resting-state functional connectivity 1437, Resting-state functional connectivity 1438, Resting-state functional connectivity 1439, Resting-state functional connectivity 1440, Resting-state functional connectivity 1441, Resting-state functional connectivity 1442, Resting-state functional connectivity 1443, Resting-state functional connectivity 1444, Resting-state functional connectivity 1445, Resting-state functional connectivity 1446, Resting-state functional connectivity 1447, Resting-state functional connectivity 1448, Resting-state functional connectivity 1449, Resting-state functional connectivity 1450, Resting-state functional connectivity 1451, Resting-state functional connectivity 1452, Resting-state functional connectivity 1453, Resting-state functional connectivity 1454, Resting-state functional connectivity 1455, Resting-state functional connectivity 1456, Resting-state functional connectivity 1457, Resting-state functional connectivity 1458, Resting-state functional connectivity 1459, Resting-state functional connectivity 1460, Resting-state functional connectivity 1461, Resting-state functional connectivity 1462, Resting-state functional connectivity 1463, Resting-state functional

connectivity 1464, Resting-state functional connectivity 1465, Resting-state functional connectivity 1466, Resting-state functional connectivity 1467, Resting-state functional connectivity 1468, Resting-state functional connectivity 1469, Resting-state functional connectivity 1470, Resting-state functional connectivity 1471, Resting-state functional connectivity 1472, Resting-state functional connectivity 1473, Resting-state functional connectivity 1474, Resting-state functional connectivity 1475, Resting-state functional connectivity 1476, Resting-state functional connectivity 1477, Resting-state functional connectivity 1478, Resting-state functional connectivity 1479, Resting-state functional connectivity 1480, Resting-state functional connectivity 1481, Resting-state functional connectivity 1482, Resting-state functional connectivity 1483, Resting-state functional connectivity 1484, Resting-state functional connectivity 1485, Grey-white contrast in unknown (left hemisphere), Grey-white contrast in bankssts (left hemisphere), Grey-white contrast in caudalanteriorcingulate (left hemisphere), Grey-white contrast in caudalmiddlefrontal (left hemisphere), Grey-white contrast in cuneus (left hemisphere), Grey-white contrast in entorhinal (left hemisphere), Grey-white contrast in fusiform (left hemisphere), Grey-white contrast in inferiorparietal (left hemisphere), Grey-white contrast in inferiortemporal (left hemisphere), Grey-white contrast in isthmuscingulate (left hemisphere), Grey-white contrast in lateraloccipital (left hemisphere), Grey-white contrast in lateralorbitofrontal (left hemisphere), Grey-white contrast in lingual (left hemisphere), Grey-white contrast in medialorbitofrontal (left hemisphere), Grey-white contrast in middletemporal (left hemisphere), Grey-white contrast in parahippocampal (left hemisphere), Grey-white contrast in paracentral (left hemisphere), Grey-white contrast in parsopercularis (left hemisphere), Grey-white contrast in parsorbitalis (left hemisphere), Grey-white contrast in parstriangularis (left hemisphere), Grey-white contrast in pericalcarine (left hemisphere), Grey-white contrast in postcentral (left hemisphere), Grey-white contrast in posteriorcingulate (left hemisphere), Grey-white contrast in precentral (left hemisphere), Grey-white contrast in precuneus (left hemisphere), Grey-white contrast in rostralanteriorcingulate (left hemisphere), Grey-white contrast in rostralmiddlefrontal (left hemisphere), Grey-white contrast in superiorfrontal (left hemisphere), Grey-white contrast in superiorparietal (left hemisphere), Grey-white contrast in superiortemporal (left hemisphere), Grey-white contrast in supramarginal (left hemisphere), Grey-white contrast in frontalpole (left hemisphere), Grey-white contrast in temporalpole (left hemisphere), Grey-white contrast in transversetemporal (left hemisphere), Grey-white contrast in insula (left hemisphere), Grey-white contrast in unknown (right hemisphere), Grey-white contrast in bankssts (right hemisphere), Grey-white contrast in caudalanteriorcingulate (right hemisphere), Grey-white contrast in caudalmiddlefrontal (right hemisphere), Grey-white contrast in cuneus (right hemisphere), Grey-white contrast in entorhinal (right hemisphere), Grey-white contrast in fusiform (right hemisphere), Grey-white contrast in inferiorparietal (right hemisphere), Grey-white contrast in inferiortemporal (right hemisphere), Grey-white contrast in isthmuscingulate (right hemisphere), Grey-white contrast in lateraloccipital (right hemisphere), Grey-white contrast in lateralorbitofrontal (right hemisphere), Grey-white contrast in lingual (right hemisphere), Grey-white contrast in medialorbitofrontal (right hemisphere), Grey-white contrast in middletemporal (right hemisphere), Grey-white contrast in parahippocampal (right hemisphere), Grey-white contrast in paracentral (right hemisphere), Grey-white contrast in parsopercularis (right hemisphere), Grey-white contrast in parsorbitalis (right hemisphere), Grey-white contrast in parstriangularis (right hemisphere), Grey-white contrast in pericalcarine (right hemisphere), Grey-white contrast in postcentral (right hemisphere), Grey-white contrast in posteriorcingulate (right hemisphere), Grey-white contrast in precentral (right hemisphere), Grey-white contrast in precuneus (right hemisphere), Grey-white contrast in rostralanteriorcingulate (right hemisphere), Grey-white contrast in rostralmiddlefrontal (right hemisphere), Grey-white contrast in superiorfrontal (right hemisphere), Grey-white contrast in superiorparietal (right hemisphere), Grey-white contrast in superiortemporal (right hemisphere), Grey-white contrast in supramarginal (right hemisphere), Grey-white contrast in frontalpole (right hemisphere), Grey-white contrast in temporalpole (right hemisphere), Grey-white contrast in transversetemporal (right hemisphere), Grey-white contrast in insula (right hemisphere)

**Eye PhenoBAG**: Overall macular thickness (left), Overall macular thickness (right), Macular thickness at the central subfield (left), Macular thickness at the central subfield (right), Macular thickness at the inner inferior subfield (left), Macular thickness at the inner inferior subfield (right), Macular thickness at the inner nasal subfield (left), Macular thickness at the inner nasal subfield (right), Macular thickness at the inner superior subfield (left), Macular thickness at the inner superior subfield (right), Macular thickness at the inner temporal subfield (left), Macular thickness at the inner temporal subfield (right), Macular thickness at the outer inferior subfield (left), Macular thickness at the outer inferior subfield (right), Macular thickness at the outer nasal subfield (left), Macular thickness at the outer nasal subfield (right), Macular thickness at the outer superior subfield (left), Macular thickness at the outer superior subfield (right), Macular thickness at the outer temporal subfield (left), Macular thickness at the outer temporal subfield (right), Overall average retinal pigment epithelium thickness (left), Overall average retinal pigment epithelium thickness (right), Average retinal nerve fibre layer thickness (left), Average retinal nerve fibre layer thickness (right), Average inner nuclear layer thickness (left), Average inner nuclear layer thickness (right), Average ganglion cell-inner plexiform layer thickness (left), Average ganglion cell-inner plexiform layer thickness (right), INL-ELM thickness of the central subfield (left), INL-ELM thickness of the central subfield (right), INL-ELM thickness of the inner subfield (left), INL-ELM thickness of the inner subfield (right), INL-ELM thickness of the outer subfield (left), INL-ELM thickness of the outer subfield (right), Average INL-ELM thickness (left), Average INL-ELM thickness (right), ELM-ISOS thickness of central subfield (left), ELM-ISOS thickness of central subfield (right), ELM-ISOS thickness of inner subfield (left), ELM-ISOS thickness of inner subfield (right), ELM-ISOS thickness of outer subfield (left), ELM-ISOS thickness of outer subfield (right), Average ELM-ISOS thickness (left), Average ELM-ISOS thickness (right), ISOS-RPE thickness of central subfield (left), ISOS-RPE thickness of central subfield (right), ISOS-RPE thickness of inner subfield (left), ISOS-RPE thickness of inner subfield (right), ISOS-RPE thickness of outer subfield (left), ISOS-RPE thickness of outer subfield (right), Average ISOS-RPE thickness (left), Average ISOS-RPE thickness (right), INL-RPE thickness of central subfield (left), INL-RPE thickness of central subfield (right), INL-RPE thickness of inner subfield (left), INL-RPE thickness of inner subfield (right), INL-RPE thickness of outer subfield (left), INL-RPE thickness of outer subfield (right), Average INL-RPE thickness (left), Average INL-RPE thickness (right)

**Cardiovascular PhenoBAG**: "Pulse rate, automated reading", "Diastolic blood pressure, automated reading", "Systolic blood pressure, automated reading"

**Hepatic PhenoBAG**: Alkaline phosphatase (**Musculoskeletal**), Alanine aminotransferase (ALT), Aspartate aminotransferase (AST), Direct bilirubin, Gamma glutamyltransferase, Total bilirubin, Albumin (**Renal**), Total protein (**Renal**)

**Immune PhenoBAG**: White blood cell (leukocyte) count, Red blood cell (erythrocyte) count, Haematocrit percentage, Mean corpuscular volume, Mean corpuscular haemoglobin, Mean corpuscular haemoglobin concentration, Red blood cell (erythrocyte) distribution width, Platelet count, Platelet crit, Mean platelet (thrombocyte) volume, Platelet distribution width, Lymphocyte count, Monocyte count, Neutrophill count, Eosinophill count, Basophill count, Nucleated red blood cell count, Lymphocyte percentage, Monocyte percentage, Neutrophill percentage, Eosinophill percentage, Basophill percentage, Nucleated red blood cell percentage, Reticulocyte percentage, Reticulocyte count, Mean reticulocyte volume, Mean sphered cell volume, Immature reticulocyte fraction, High light scatter reticulocyte percentage, High light scatter reticulocyte count, C-reactive protein, Haemoglobin concentration

**Metabolic PhenoBAG**: Glycated haemoglobin (HbA1c), Apolipoprotein A, Apolipoprotein B, Cholesterol, Glucose, HDL cholesterol, LDL direct, Lipoprotein A, Triglycerides

**Musculoskeletal PhenoBAG**: Handgrip strength (average), Waist circumference, Hip circumference, Standing height, Body mass index (BMI), Weight, Calcium (**Renal**), Vitamin D, Waist-hip circumference ratio, Heel bone mineral density (average), Ankle spacing width (average), Phosphate (**Renal**)

**Pulmonary PhenoBAG**: FEV1-FVC ratio, Forced vital capacity (FVC), Forced expiratory volume in 1-second (FEV1), Peak expiratory flow (PEF)

**Renal PhenoBAG**: Creatinine (enzymatic) in urine, Potassium in urine, Sodium in urine, Urea, Creatinine, Cystatin C, Urate

**eNote 2: Sex-stratified brain ProtBAG**

Sex differences in aging clocks have been identified in recent studies[3].

For example, Moguilner et al.[4] studied brain aging clocks derived from EEG data in aging and dementia across geographically diverse populations. They analyzed data from 5,306 participants across 15 countries, including both Latin American and Caribbean (LAC) and non-LAC regions. Participants comprised healthy controls and individuals diagnosed with mild cognitive impairment (MCI), Alzheimer's disease (AD), and behavioral variant frontotemporal dementia (bvFTD). Specifically for sex differences, they found that, in LAC regions, the study reported larger brain-age gaps in females compared to males within both the control and AD groups, in which the model was trained on combined data from both sexes (Fig. 4 in their paper).

In a recent study, Argentieri et al[3] explored the effect of sex on proteome-based aging clocks using UK Biobank data. They found similar age prediction performance between sex-specific models and a combined-sex model. Predicted ages from both approaches were nearly perfectly correlated. Notably, the female-only model showed a slightly better fit (MAE = 2.25) compared to the male-only model (MAE = 2.45). Both models used over 2,000 proteins and were not organ specific. Consistency was observed in many of the most important proteins identified through feature selection between sexes.

The two studies above-mentioned addressed different questions: one focused on sex differences in brain aging clocks, while the other examined whether aging clocks trained on combined-sex data are generalizable to sex-specific models. They also employed distinct modeling techniques (trained on both sexes' data or sex-specific data) and data types (EEG vs. proteomics).

Here, I build upon these two studies to examine sex differences in greater detail, focusing on organ-specific analyses and providing additional interpretation of ProtBAG, using the brain as a case study (**eFigure 5**). First, using linear (Lasso) and nonlinear (NN) models, I showed that the models trained on male-only data could not be generalize well from the training/validation/test data to the independent test data using both 4589 CN participants and 31,808 mixed-pathology participants, albeit the latter alleviated such overfitting with the increased sample size (**eFigure 5a-d**). The poor generalization of the male-only model could be due to inherent biological differences between sexes that influence the aging process, leading to distinct patterns in brain aging or pathology across males and females. This could cause the model to learn sex-specific features that are not representative of the broader population, in the context of the brain ProtBAG. More interestingly, a nonlinear model like NN seemed to mitigate the overfitting phenomenon to a certain extent compared to the situation in the linear Lasso model (i.e., Cohen's D between the training and ind. test data). The potential variability in male's and female's proteomics data may arise from differences in hormonal regulation and how male biology responds to aging. Males might experience greater fluctuations in biological aging markers, creating a more nonlinear relationship between biological and chronological age. These fluctuations can introduce more noise, leading to overfitting in linear models like Lasso. Nonlinear models, such as NN, are better equipped to capture this complexity, allowing them to handle the larger variability and reduce overfitting by modeling more intricate, non-linear patterns in the data.

Secondly, I found that the predicted ages from models using combined data from both sexes and single-sex data were highly correlated in both cases: *i*) when the model generalized well to independent test data (Pearson's $r > 0.94$, as shown in Argentieri et al.) (**eFig. 5e-f**), and *ii*) when it failed to generalize (Pearson's $r = 0.94$) (**eFig. 5g**). Further investigation revealed that the predicted age from the male-only Lasso model was consistently higher (potential domain shift due to underlying biology in male proteomics data) in mean compared to the model using both sexes (**eFig. 5h-i**).

Finally, the predicted ages from male- and female-only models cannot be directly compared. Following the method of Moguilner et al. using the model trained on both sexes, I found sex differences in the brain ProtBAG, with females showing a higher mean BAG (**eFig. J**; Cohen's D=0.36; P-value=$7.08 \times 10^{-5}$), supporting the findings in Moguilner et al. demonstrated using EEG data.

Relatedly, understanding the genetic underpinnings contributing to these sex differences is crucial. In our previous studies, we conducted sex-stratified GWAS on 12 multi-organ aging clocks derived from 9 organs using neuroimaging[5] and clinical data[2]. We observed that, in certain organs such as the cardiovascular system, the genetic architecture differed between male- and female-specific GWAS signals. Similarly, future studies should explore this in the context of ProtBAG.

In summary, sex differences are evident in the brain ProtBAG, with male-specific models being more susceptible to overfitting from female-specific models. While the predicted ages from sex-specific models and models trained on combined data are highly consistent (i.e., correlated), it is crucial to address issues such as model overfitting and domain shift, rather than relying solely on the Pearson's *r* coefficient for evaluation.

**eNote 3: The definition of genomic loci, independent significant SNP, lead SNP, candidate SNP**

FUMA defined the significant independent SNPs, lead SNPs, candidate SNPs, and genomic risk loci as follows (https://fuma.ctglab.nl/tutorial#snp2gene):

*Independent significant SNPs*

They are defined as SNPs with $P \leq 5 \times 10^{-8}$ that are independent of each other at the user-defined $r^2$ (set to 0.6 in the current study). I further describe *candidate SNPs* as those in linkage disequilibrium (LD) with independent significant SNPs. FUMA then queries each candidate SNP in the GWAS Catalog to check whether any clinical traits have been reported to be associated with previous GWAS studies.

*Lead SNPs*

Lead SNPs are defined as independent significant SNPs that are also independent of each other at $r^2 < 0.1$. If multiple independent significant SNPs are correlated at $r^2 \geq 0.1$, then the one with the lowest individual *P*-value becomes the lead SNP. If $r^2$ threshold is set to 0.1 for the independent significant SNPs, then they would constitute the identical set as the lead SNPs. FUMA thus advises setting $r^2$ to be 0.6 or higher.

*Genomic risk loci*

FUMA defines genomic risk loci to include all independent signals physically close or overlapping in a single locus. First, independent significant SNPs dependent on each other at $r^2 \geq 0.1$ are assigned to the same genomic risk locus. Then, independent significant SNPs with less than the user-defined distance (250 kilobases by default) away from one another are merged into the same genomic risk locus - the distance between two LD blocks of two independent significant SNPs is the distance between the closest points from each LD block. Each locus is represented by the SNP within the locus with the lowest *P*-value.

**eNote 4: The phenotypic and genetic correlation between the 11 ProtBAGs and 2448 proteins**

A total of 132 significant ProtBAG-protein genetic correlations were identified after Bonferroni correction, encompassing the Pulmonary, Heart, Brain, Hepatic, Renal, Endocrine, Immune, and Skin ProtBAGs, involving 119 unique plasma proteins (**eTable 5b**).These ProtBAG-protein pairs showed significant results for both phenotypic correlation (P-value<0.05/2923/11) and genetic correlation (P-value<0.05/2448/11) after Bonferroni correction.

The phenotypic correlation analysis identified 19,503 significant associations, which is expected due to circular bias[6], as ProtBAGs were derived from a subset of these proteins. However, this post-hoc analysis was conducted to validate organ-specific patterns – demonstrating, for instance, that brain ProtBAGs are most strongly linked to brain-enriched proteins – rather than to claim predictive power. Given this context, the approach remains justifiable.

I also conducted large-scale genetic correlation analyses using GWAS summary statistics from the 11 ProtBAGs and the GWAS data for 2,448 plasma proteins (processed using the same pipeline). The goal was *i*) to validate the organ-specificity at an additional genetic level and *ii*) to provide supporting evidence for Cheverud's Conjecture, which posits that the genetic correlation between two traits (in this case, ProtBAGs and plasma proteins) largely reflects their phenotypic correlation.

An interactive webpage is available online for users to browse these significant signals (https://labs-laboratory.com/medicine/protbag_protein_interaction), and detailed statistics are present in **eTable 5b**.

**Genetic correlation and phenotypic correlation support this organ specificity pattern**: As expected, the most significant proteins within each organ-specific ProtBAG were those classified as organ-specific in **eNote 1**, based on the HPA platform. For instance, the pulmonary ProtBAG showed the highest genetic correlation with the LAMP3 protein ($g_c = 0.60 \pm 0.05$). Analyzing its expression profile in the HPA platform confirms that LAMP3 is indeed highly expressed in lung tissue (https://www.proteinatlas.org/ENSG00000078081-LAMP3/tissue). Another example is the NPPB protein, which exhibited a genetic correlation of 0.88 with the heart ProtBAG and demonstrated an nTPM value in heart tissue that was more than four times higher than in any other tissue or organ (https://www.proteinatlas.org/ENSG00000120937-NPPB/tissue).

**Cheverud's Conjecture: the genetic correlation mirrors their phenotypic correlation**

Taking the LAMP3-Pulmonary ProtBAG pair as an example, their phenotypic correlation ($p_c$ = 0.60) largely mirrors the genetic correlation ($g_c$ = 0.60). I calculated the Pearson's correlation coefficient between the 132 genetic correlations and 132 phenotypic correlations and found them to be strongly correlated ($r$ = 0.86).

**eNote 5: Sensitivity check analyses for the MR results for the causal pathway of "obesity→renal PhenoBAG→renal ProtBAG"**

As Mendelian randomization is sensitive to underlying IV assumptions, I performed sensitivity analyses to investigate the potential violation, exemplified by the three-layer causal pathway of obesity→renal PhenoBAG→renal ProtBAG.

For the causal relationship from obesity to the renal PhenoBAG, I observed two potential outlier instrumental variables (IVs, i.e., independent SNPs: rs76503812 and rs147001051) for the effect sizes on the exposure (i.e., E4_Obesity) and outcome variables (i.e., the renal PhenoBAG) (**Extended Fig. 4a**). I excluded those two outliers to rerun the analysis; the causal effect persisted, and the *beta* value slightly increased. I then showed the forest plot for the individual-SNP level of the causal effect sizes, indicating that most IVs exert positive effects (**Extended Fig. 4b**). I then performed a leave-one-IV-out analysis and found that no single SNPs largely dominated the causal effect (**Extended Fig. 4c**). Finally, I showed a symmetry funnel plot that indicates no potential "directional pleiotropy" (**Extended Fig. 4d**). To further scrutinize this bias, I applied MR-Egger regression with MAF-corrected weights to the summarized data, yielding an intercept estimate of -0.00629 with an associated P-value of 0.99. This further supports the absence of directional pleiotropy. There was also no apparent heterogeneity in the IV estimates from each genetic variant individually, as evidenced by Cochran's Q test (P = 0.06). In summary, there is no evidence that directional pleiotropy biases the conclusion.

Similarly, for the causal effect from the renal PhenoBAG to renal ProtBAG, I showed the scatter plot between the IV effect sizes from the exposure and outcome variables with one IV as a potential outlier (rs2424577; **Extended Fig. 4e**). Similarly, I redid the analysis by excluding the outlier, and the relationship persisted. Most SNPs exerted a positive causal effect in our individual-level IV analysis, as shown in the forest plot (**Extended Fig. 4f**), but some SNPs showed heterogeneity, as evidenced by Cochran's Q test ($P = 4.2 \times 10^{-8}$). The outlier (rs2424577) also dominated the effect in the leave-one-IV-out analysis (**Extended Fig. 4g**). Finally, I observed a slight asymmetry in the funnel plot (**Extended Fig. 4h**). MR-Egger regression showed an intercept estimate of 0.0053 with a P-value of 0.34. This indicated that there was no strong evidence for potential horizontal pleiotropy. In summary, I did not observe clear violations that would bias the causal relationship factor for these data.

**eFigure 1: The LOWESS method for correcting the age bias, the comparison to the neuroimaging-based method, and the influence of sample size using the full set of 2448 proteins**

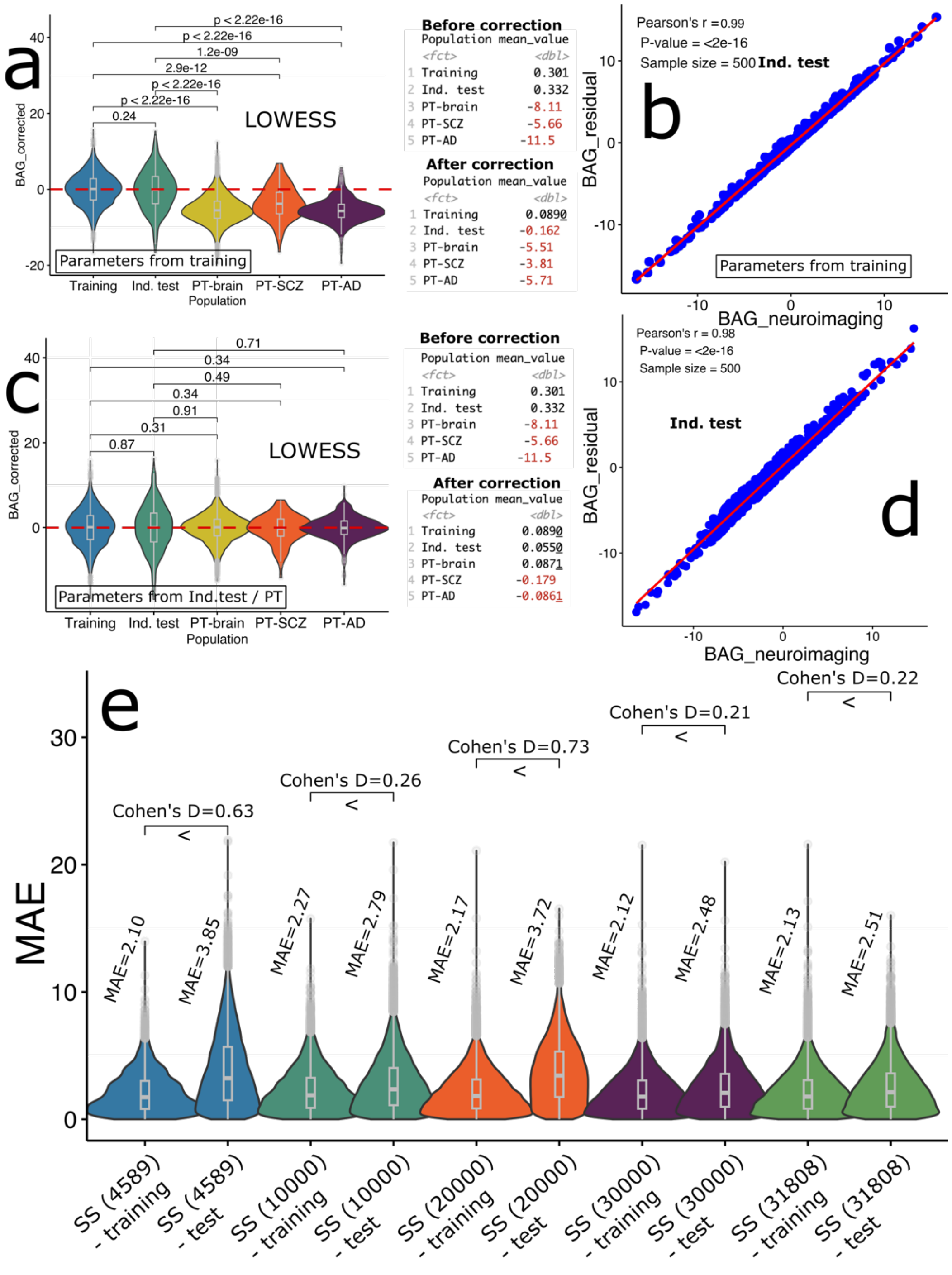


**a**) LOWESS: A nonlinear regression technique that models the relationship between Chronological Age and Predicted Age using local smoothing (weighted least squares). Plots show the BAG after the respective correction methods using parameters from the training dataset (i.e., **Parameters from training**); the table on the right shows the mean BAG in each population before and after the correction. I summarize the main difference between the two correction methods: *i*) the neuroimaging-based method implemented in Beheshti et al.[7] fits a linear regression between BAG and chronological age to remove systematic bias, deriving the $\alpha$ and $\beta$ from the training data; *ii*) the residual-based method introduced in Oh et al.[8] and Teschendorff and Horvath[9] first fits a LOWESS regression model between the predicted age and chronological age, assuming local nonlinearity of the aging trajectory. **b**) I also calculated the Pearson's $r$ for the corrected BAG between the two methods (Beheshti et al.[7] for neuroimaging-based and Oh et al.[8] for residual-based approach) using the **independent test data** ($N$=500). **c**) These plots are similar to those in figure a), but here, to correct the age bias in the independent test and PT populations (as observed in the potential domain shift in **Fig. 1b**), we used the parameters derived specifically from the independent test and PT populations. **d**) The same plot as in figure **b**, but we used the age-bias corrected BAG from figure c). **e**) Model's poor generalizability was mitigated by increasing the sample size using the full set of 2448 plasma proteins, guided by the approach in Argentieri et al.

**eFigure 2: The U-shaped relationship between the beta coefficient and the number of epochs in the neural network for the association between the brain ProtBAG and age at death**

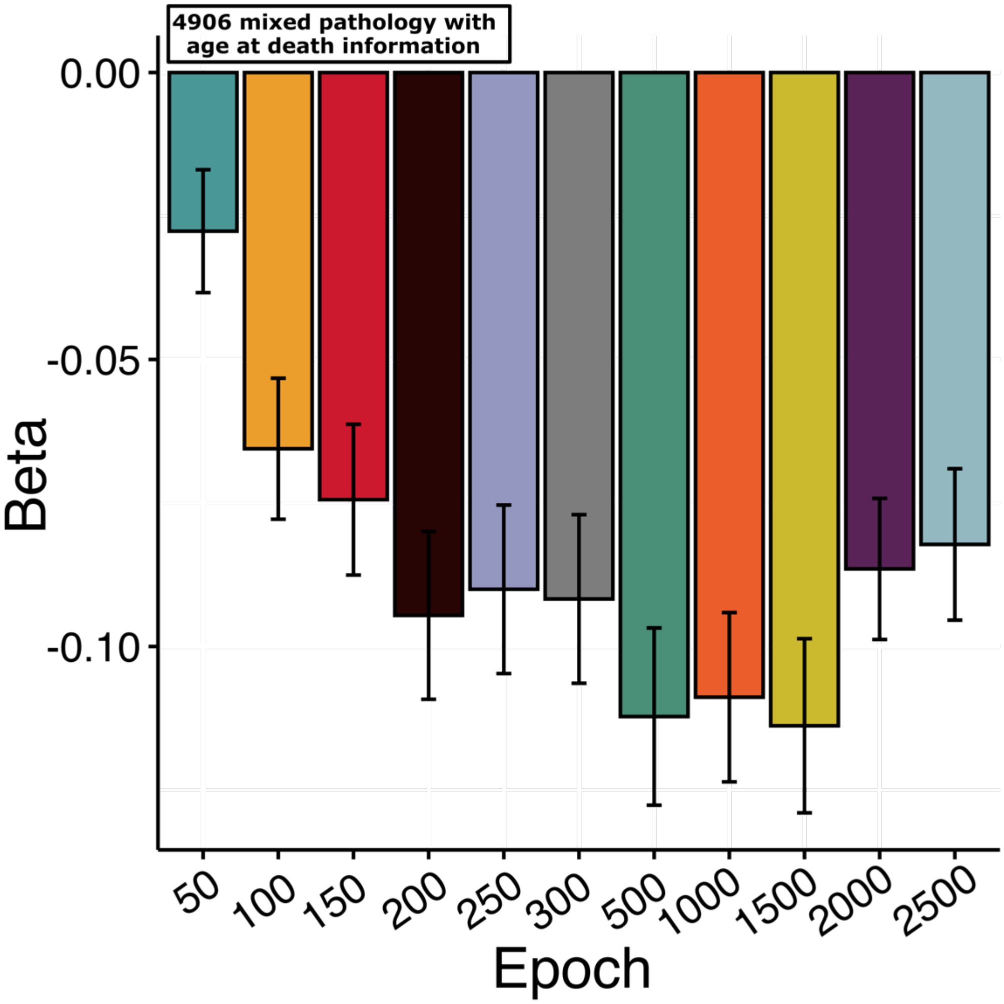


A more "tightly-fitted" model (Epoch = 2500) did not yield greater statistical power in predicting cognition (i.e., digital symbol substitution test) compared to a "moderately-fitted" model (Epoch = 500 and 1500), as indicated by the U-shaped dotted line. The $\beta$ coefficient from the linear regression model associating the brain ProtBAG with age at death was evaluated at different epochs.

**eFigure 3: Organ specificity's influence on model overfitting using the 31,808 participants with mixed pathologies and protein collinearity among different categories using the down-sampled 53 proteins**

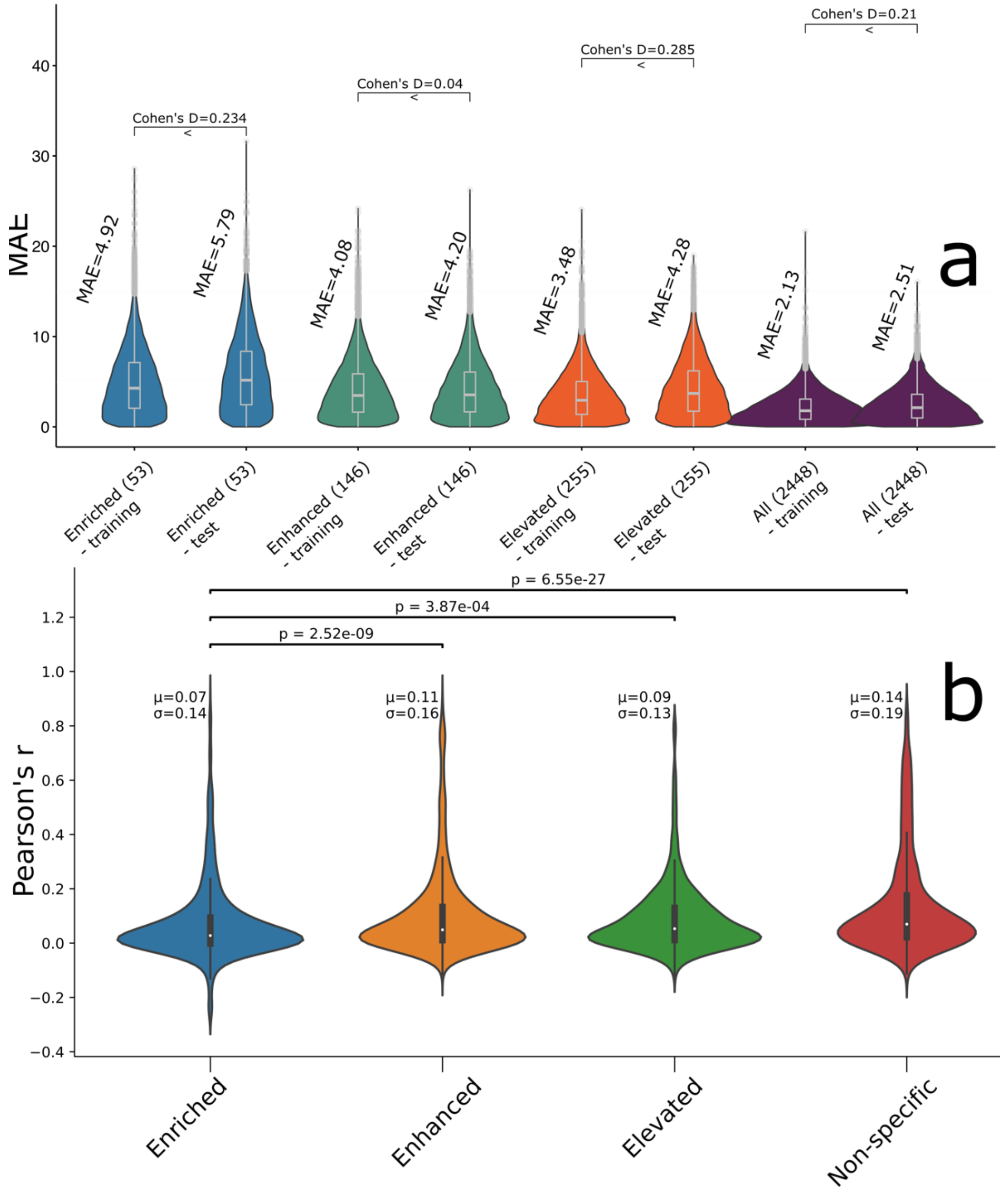


**a**) I repeated the organ specificity analyses using tissue-enriched, tissue-enhanced, tissue-elevated, and all 2,448 proteins as features. Our results showed that the overfitting issue was mitigated with the larger sample size, suggesting that sample size plays a more critical role than organ specificity or underlying pathologies in addressing overfitting. **b**) The distribution of pairwise Pearson's *r* correlation coefficients were calculated for the 53 brain-enriched proteins and, for comparison, for 53 randomly selected non-brain-enriched proteins from other categories (tissue-enhanced, tissue-elevated, and non-specific proteins) in the training dataset.

**eFigure 4: Organ specificity proteins vs. non-enriched proteins for predicting the brain ProtBAG**

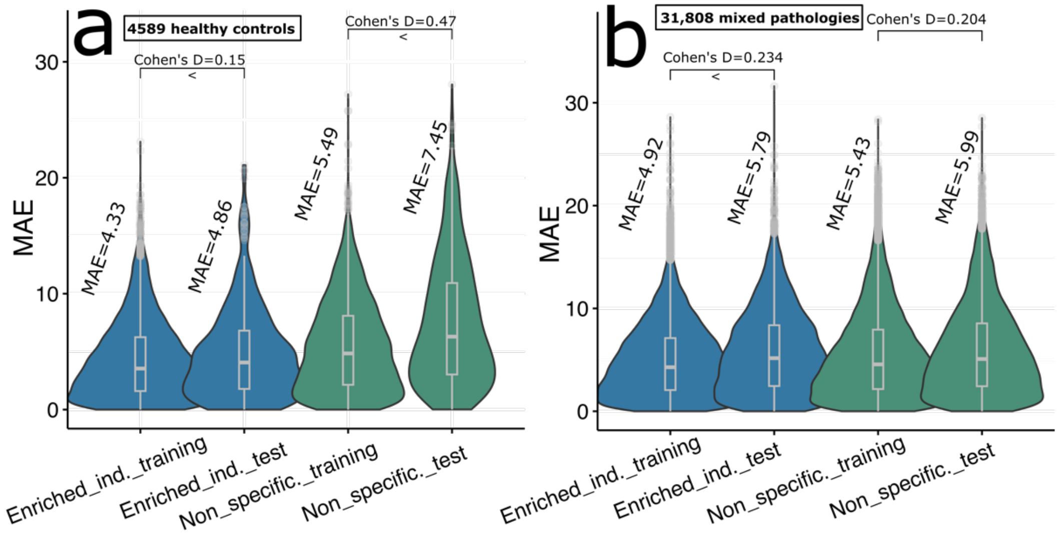


**a**) When training the model on 4589 CN participants, organ specificity (53 brain-enriched proteins) was the primary factor contributing to potential overfitting, rather than the number of features (53 randomly selected non-enriched proteins). **b**) When the sample size was increased to 31,808 participants with mixed pathologies, organ specificity was no longer a key driver of overfitting.

**eFigure 5: Sex-stratified ML models for deriving the brain ProtBAG**

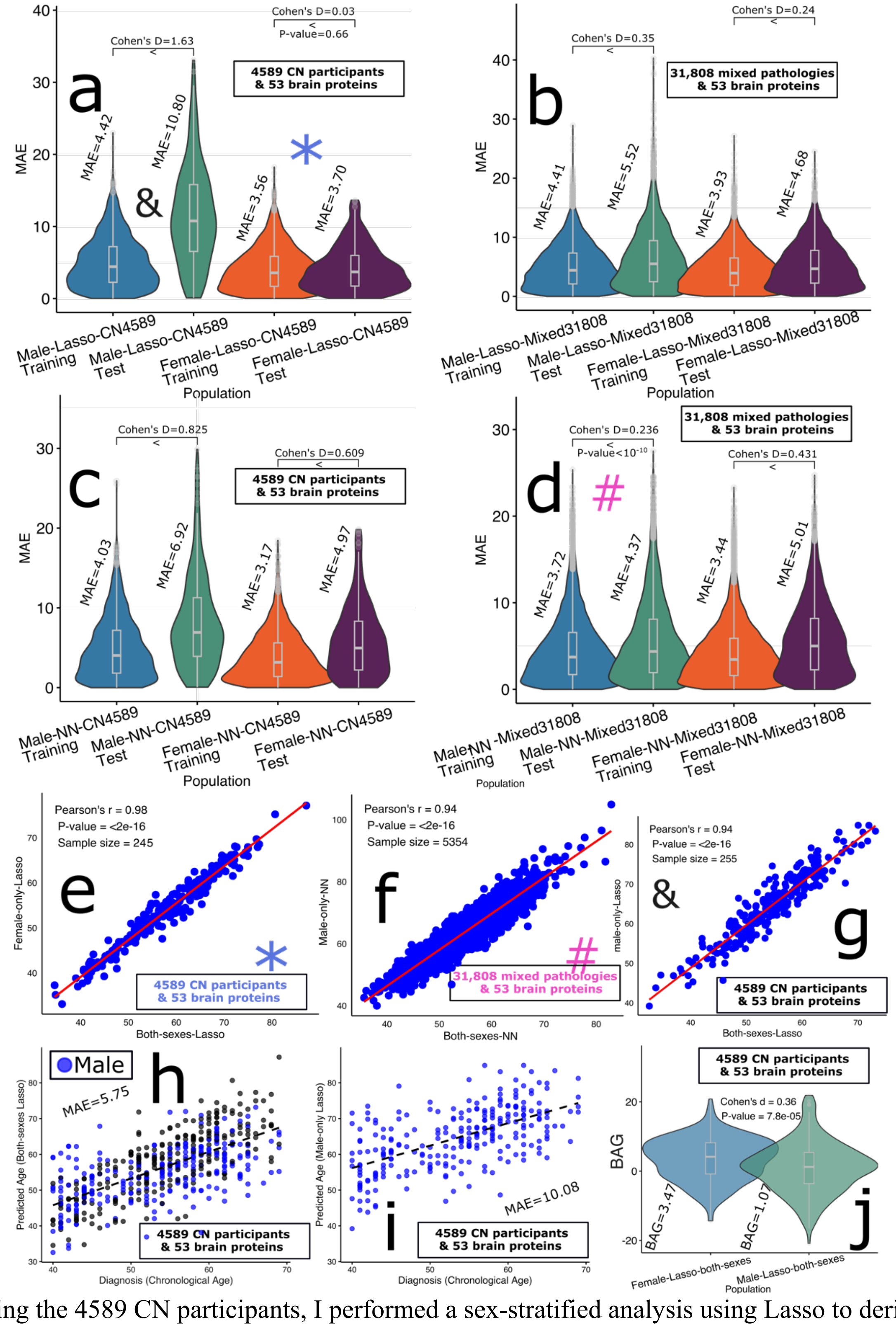


**a**) Using the 4589 CN participants, I performed a sex-stratified analysis using Lasso to derive the male- and female-specific brain ProtBAG. MAE and Cohen's D between the training/validation/test data and the ind. test data are presented. * denotes the best model for female-specific analyses; # denotes the best model for male-specific models (i.e., the most generalizable model from the training to ind. test datasets); & denotes a scenario where the model trained on male-specific data could not generalize well to independent test data. **b**) Using the 31,808 mixed-pathology participants, I performed a sex-stratified analysis using Lasso to derive the male- and female-specific brain ProtBAG. **c**) Using the 4589 CN participants, I performed a sex-stratified analysis using NN to derive the male- and female-specific brain ProtBAG. **d**) Using the 31,808 mixed-pathology participants, I performed a sex-stratified analysis using NN to derive the male- and female-specific brain ProtBAG. Following the approach of Argentieri et al.[3], I generated a scatter plot comparing the two best models: the female-specific Lasso model applied to 4,589 cognitively normal (CN) **e**) participants and the male-specific neural network (NN) model applied to 31,808 mixed-pathology participants **f**). I then compared these models to a model trained on both sexes. To ensure a fair comparison between the single-sex and mixed-sex models, all models were evaluated using the same cross-validation split. **g**) As an example, I demonstrated that the Lasso model failed to generalize to the independent test data in male-specific analyses, potentially due to domain shifts or the inherent biological complexity of male proteomics data. Despite this, I observed a very high correlation of predicted ages between models trained on combined data from both sexes and those trained specifically on male data. **h**) Scatter plot comparing predicted age and chronological age for the model trained on combined data from both sexes, with males represented by blue data points. **i**) Scatter plot comparing predicted age and chronological age for the Lasso model trained exclusively on male data. **j**) Since I observed that models trained on sex-stratified data and those trained on combined data from both sexes were highly correlated, I compared the mean BAG between females and males derived from the model trained on both sexes using the 4,589 CN participants. This approach was necessary because sex-stratified models are not ideal for directly comparing the mean BAG between males and females, as the normative reference – defined by the health status of the training data – varies between the two sexes.

**eFigure 6: Feature importance of the organ-enriched proteins for deriving the 11 ProtBAGs**

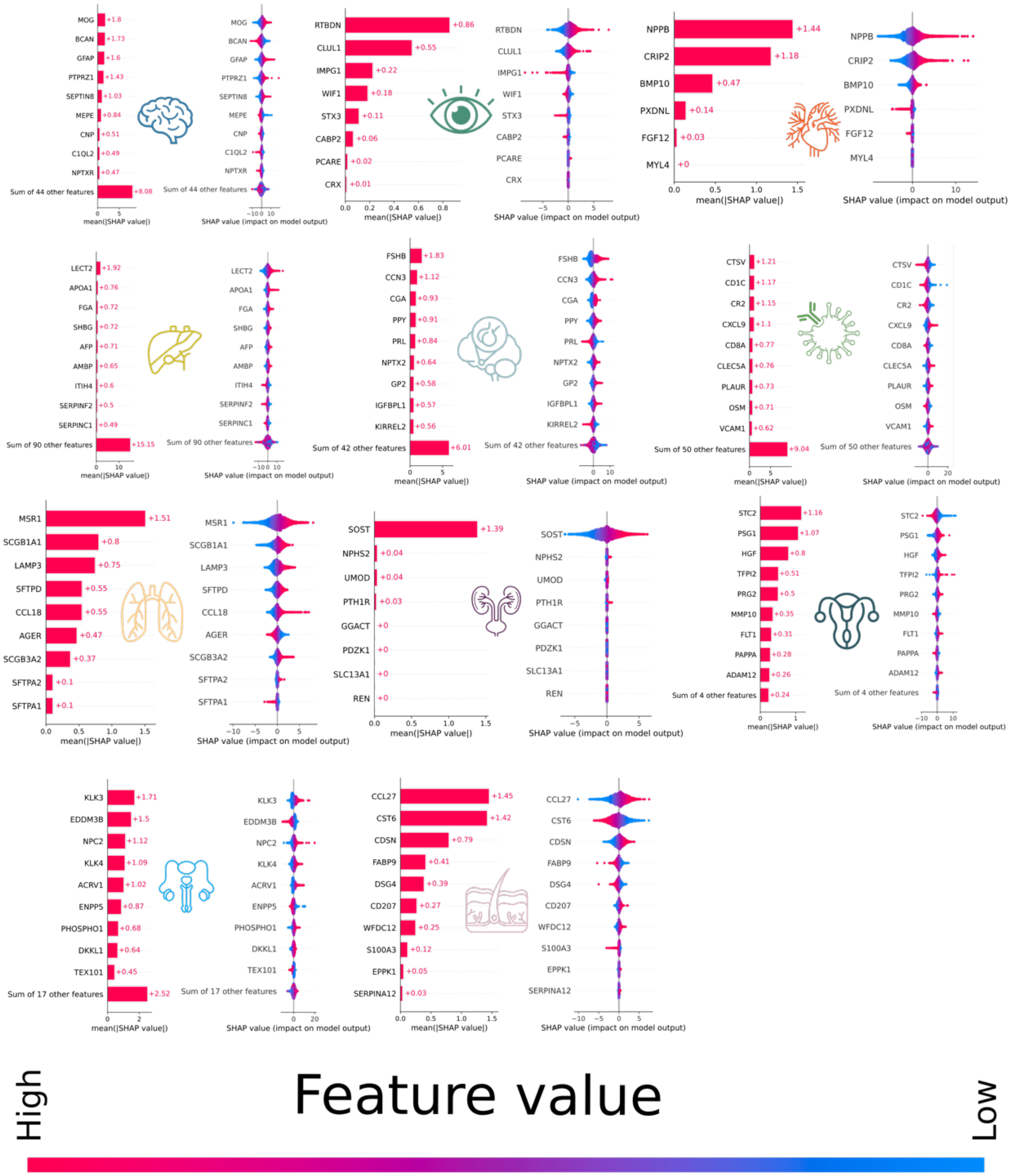


SHAP analysis of the most important organ-enriched proteins for the 11 ProtBAG. For each ProtBAG, the left panel shows the bar plot representing the mean absolute SHAP values for each protein, highlighting their overall contribution to the model's predictions. The right panel displays the bee swarm plot, visualizing the distribution of SHAP values for individual samples in the training data. A positive SHAP value means that the feature in question is increasing the predicted age. In other words, the presence or higher value of this feature makes the model predict an older biological age. A negative SHAP value means that the feature is decreasing the predicted age. A lower value or the absence of this feature contributes to the model predicting a younger biological age. Red color indicates high feature values, blue indicates low values. For organs with more than 10 organ-enriched proteins, I display the top 9 protein.

**eFigure 7: Phenome-wide associations via the GWAS Atlas platform**

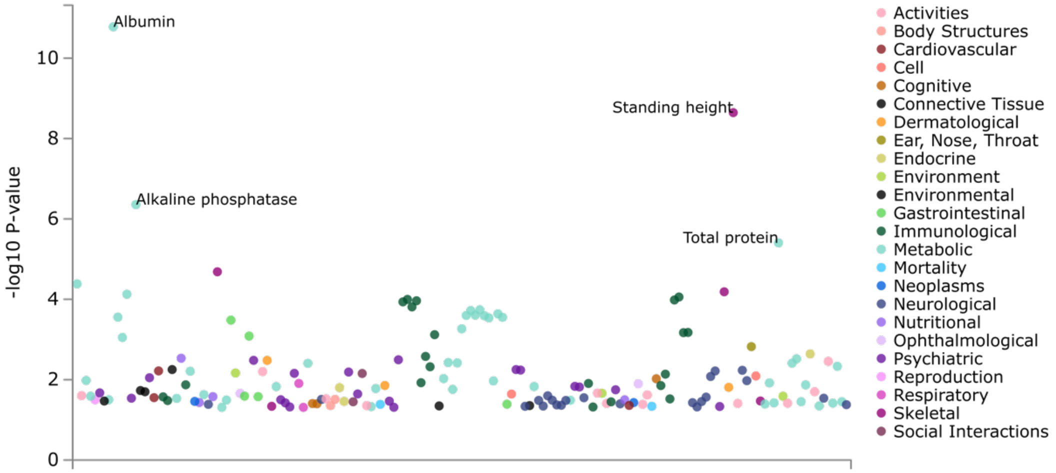


I used the PheWAS functionality on the potential causal SNP (rs7212936) in the GWAS Atlas platform to validate and complement the traits identified in the GWAS Catalog platform. I then annotated the four most significant traits based on existing literature.

**eFigure 8: Multi-omic evidence for the *SERPINF2* gene in co-localization analysis**

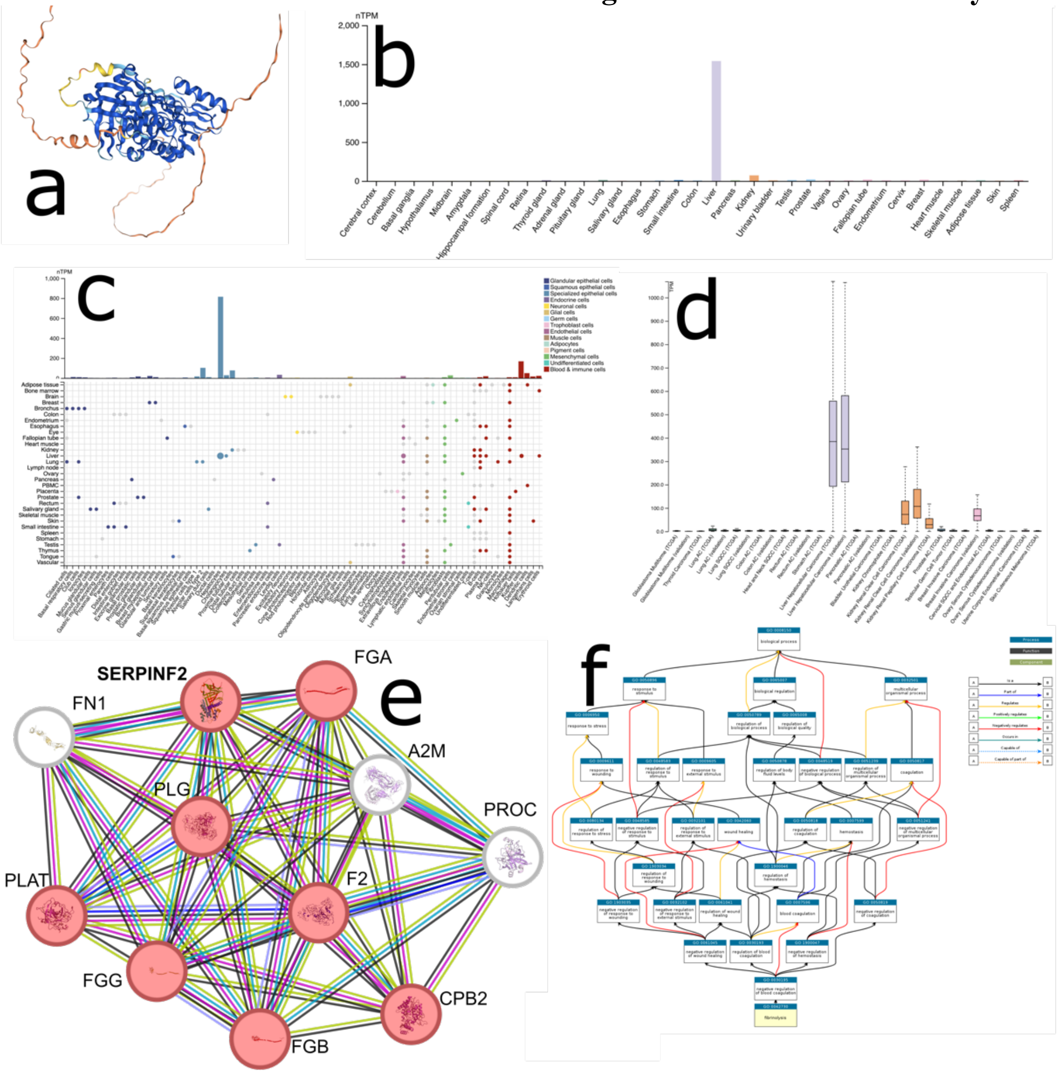


**a**) Structure prediction of *SERPINF2* from Alphafold v2.3.2. **b**) The expression profile of the SERPINF2 protein was analyzed using GTEx RNA-seq data across 50 different tissues. As anticipated, this protein exhibited liver-enriched expression, with mRNA levels more than four times higher in the liver compared to other tissues. Notably, SERPINF2 is one of the features used to derive the hepatic ProtBAG, which incorporates a broader range of data beyond GTEx (**Method 3c**). **c**) Using the HPA-curated data, a summary of normalized single cell RNA (nTPM) from all single cell types is presented. Color-coding is based on cell type groups, each consisting of cell types with functional features in common. As expected, hepatocytes are cell-type enriched (814.9 nTPM). **d**) RNA expression overview shows RNA-seq data from The Cancer Genome Atlas, and the most enriched cancer types are related to the liver and kidney. **e**) The protein-protein interaction network of the SERPINF2 protein was constructed using the STRING database, which integrates multiple sources of interaction evidence, including curated databases and experimentally validated data. Node colors indicate different interaction shells, with red nodes representing the first shell and white nodes representing the second shell. Edge colors correspond to various interaction types, such as gene neighborhood, gene fusions, gene co-occurrence, as well as co-expression and protein homology. **f**) An example of the biological pathway analysis highlights the fibrinolysis pathway (GO:0042730), which has been associated with both liver and kidney diseases.

**eFigure 9: The systemic disease category classification performance with age and sex as additional features**

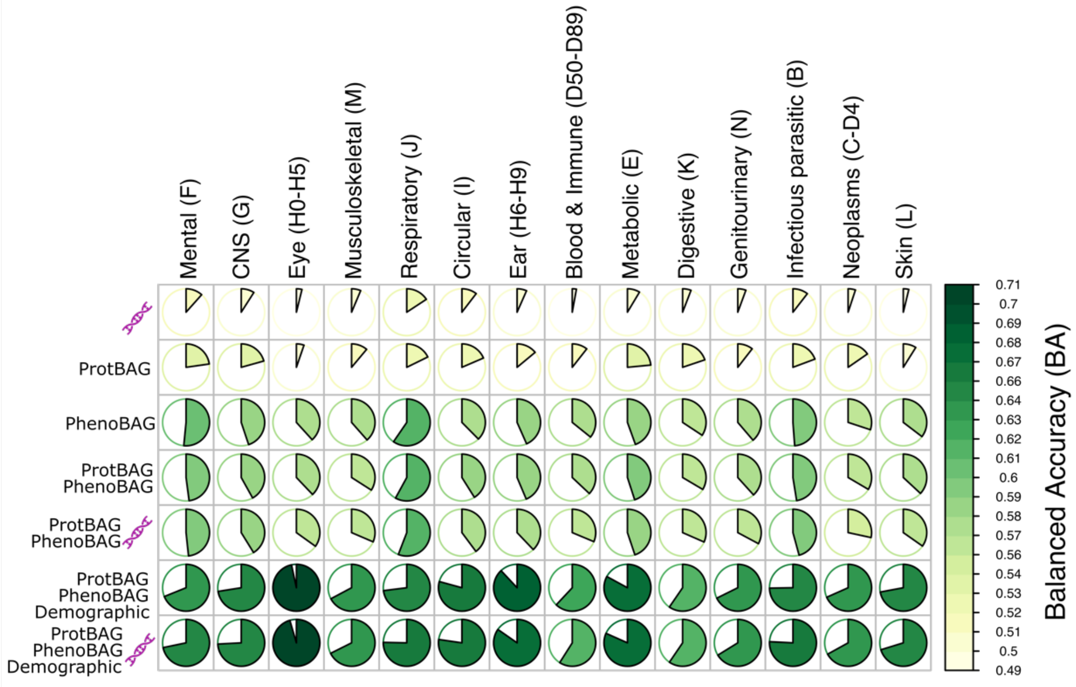


Adding age and sex as additional features improves classification accuracy.

**eFigure 10: Compare the brain ProtBAG derived from 4589 CN participants and 31,808 mixed-pathology participants, and also the two different age correction approaches**

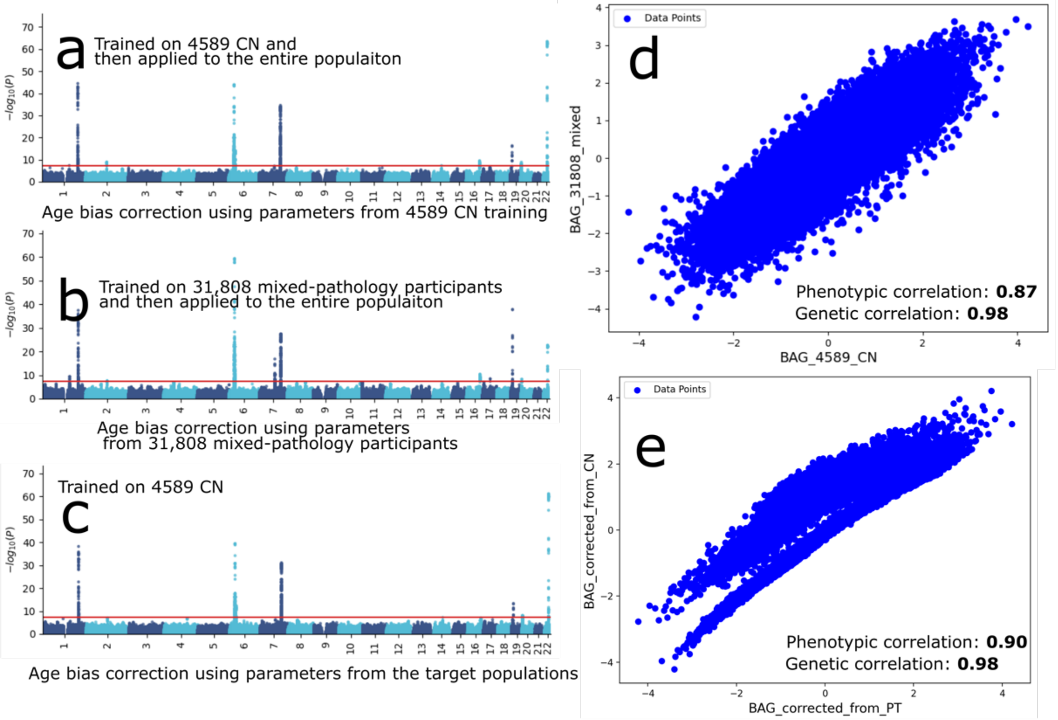


**a**) GWAS signals identified by the brain ProtBAG in the European ancestry population using the model trained on 4,589 cognitively normal (CN) participants and the age bias correction using parameters from the CN population, and **b**) the model trained on 31,808 participants with mixed pathologies. **c**) GWAS signals associated with the brain ProtBAG were identified in the European ancestry population using a model trained on 4,589 cognitively normal (CN) individuals. Age bias correction was performed using parameters (alpha and beta) derived specifically from each target population, meaning the PT population's own parameters were used to adjust the ProtBAG values within that group. **d**) Phenotypic and genetic correlations between the two approaches in a and b. **e**) Phenotypic and genetic correlations between the two approaches in a and c. The high correlations, especially the genetic correlations, suggest that the underlying genetic variants associated with both methods are highly similar. Figure e clearly illustrates evidence of domain shift, as the alpha and beta parameters derived from the CN population did not generalize well to the PT population.

**eFigure 11: Protein missing NPX value imputation quality**

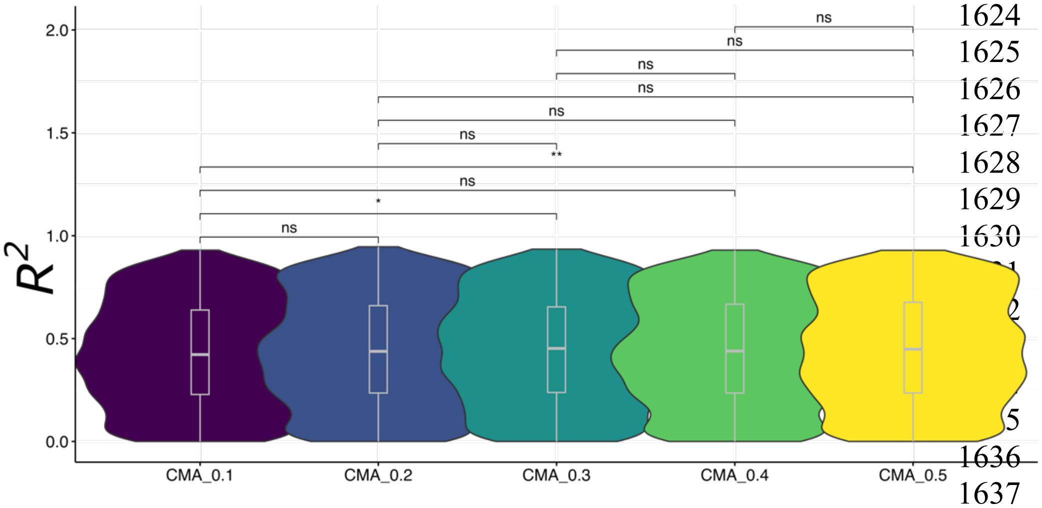


Imputation performance is unrelated to copy mask amount (CMA) using AutoComplete. The original study has assessed the impact of missing value rate on model performance.

**eFigure 12: UK biobank population selections and the nested cross-validation procedure to derive the 11 ProtBAGs.**

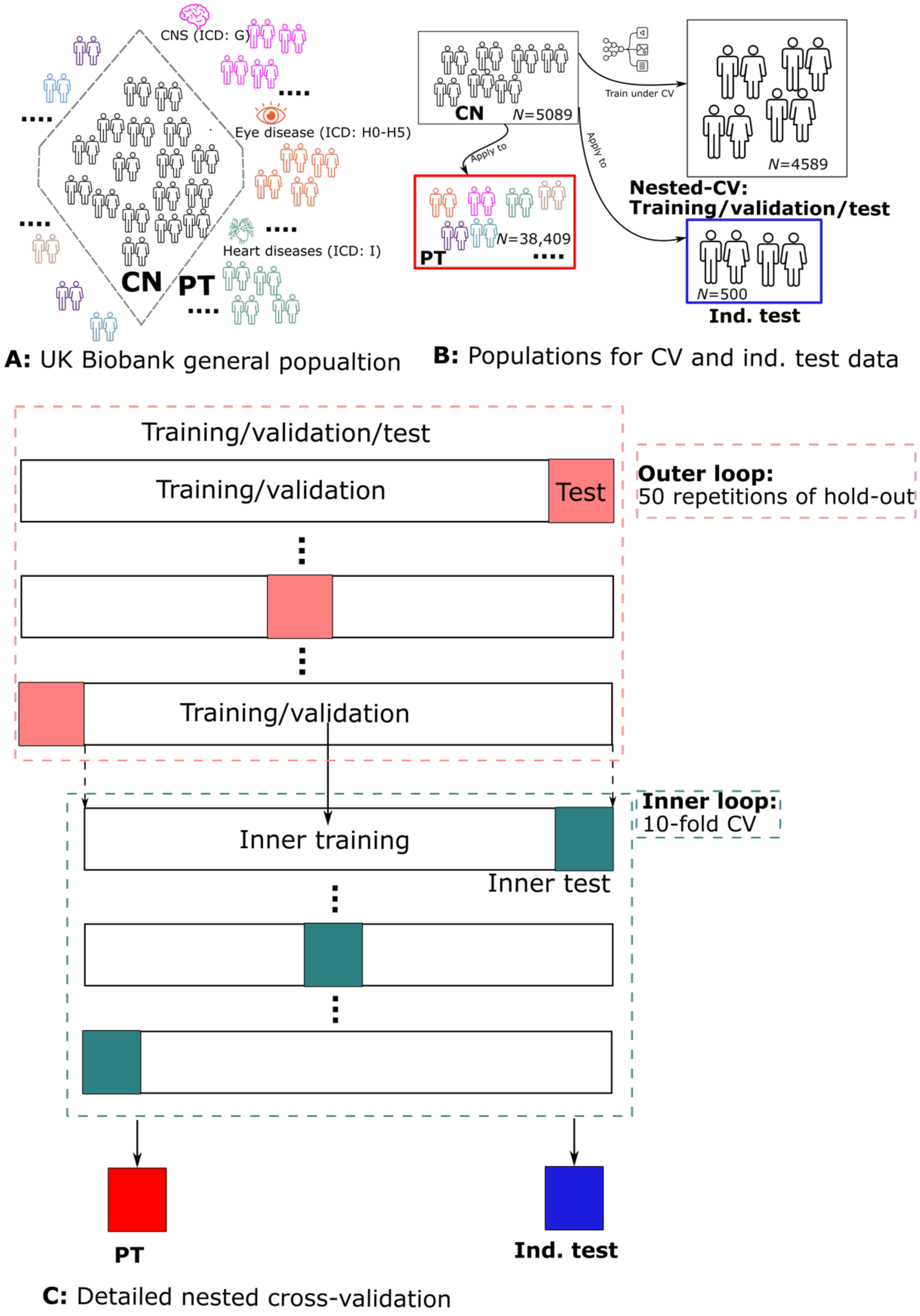


**a**): The UK Biobank is a UK-based general population. I define a healthy control population (CN) based on the ICD code and the in-patient medical history (i.e., participants that did not have any disease diagnosis records in the system), and a patient population (PT) (i.e., participants that had any single or multiple disease diagnosis records in the system). The motivation is both for methodological modeling (e.g., protein levels in CN are more stable compared to PT, where various diseases, due to pathological perturbations, may lead to both up- and down-regulation of plasma protein levels) and clinical interpretation (e.g., modeling the effects of "pure" biological aging and then applying the model to diseased populations to identify disease-specific deviations, such as how ProtBAG shifts in Alzheimer's disease patients compared to CN). **b**): The CN population was further split into the CN independent test (ind. test; $N$=500) dataset and the CN nested CV dataset ($N$=4589 for training/validation/test datasets). The ML models were trained on the CN nested CV dataset and then applied to the CN ind. test dataset and the PT dataset ($N$=38,409). **c**): Details on the nested CV procedure include a repeated hold-out cross-validation with 50 repetitions (80% training/validation and 20% testing) for the outer loop and 10-fold cross-validation for the inner loop for hyperparameter selection (e.g., $C$ for SVR).

**eTable 1: The characteristics of the MULTI consortium**

| Data type | Data type | Study | *N* | Age [year (mean/std)] | | | Sex (female) | | |
|---|---|---|---|---|---|---|---|---|---|
| | | | | CN training[d] | CN ind. test | PT | CN training | CN ind. test | PT |
| Individual-level | Plasma proteomics (ProtBAG) | UKBB | 43,498[a] | 54.11±7.69 | 54.00±7.84 | 57.68±8.05 | 2382/52% | 255/51% | 20,746/54% |
| Individual-level | Phenotypic and clinical variables (PhenoBAG) | UKBB | 193,895[b] | | 56.09±8.06[e] | | | 92,685/48% | |
| Summary-level | GWAS summary statistics | FinnGen | 521[c] | NA | NA | NA | NA | NA | NA |
| Summary-level | GWAS summary statistics | PGC | 4[c] | NA | NA | NA | NA | NA | NA |

[a]This is the proteomics population after quality check and imputation for missing NPX value.
[b]This is the entire population that merged the 9 PhenoBAGs from our two previous studies[2,1].
[c]I only used the publicly available GWAS summary statistics from FinnGen and PGC for our Mendelian randomization analyses.
[d]The training data here was used to cross-validate the AI/ML models that comprised training/validation/test splits.
[e]The exact training/validation/test splits for the 9 PhenoBAGs are detailed in our previous studies following the same procedure as shown here for the 11 ProtBAGs[2,1]. Here, I showed the age and sex information for the entire population.

**eTable 2: Benchmarking the ProtBAG prediction performance for the CN training/validation/test (Training) and CN independent test (Ind. test) datasets**

a) **Before age bias correction**

| Organ | Dataset | Model | MAE | r |
|---|---|---|---|---|
| Reproductive_female | Training | svr | 5.99006817 | 0.34587722 |
| Reproductive_female | Ind. test | svr | 6.54194349 | 0.41402729 |
| Pulmonary | Training | svr | 5.79416383 | 0.41905129 |
| Pulmonary | Ind. test | svr | 5.72462223 | 0.45383097 |
| Heart | Training | svr | 5.88557966 | 0.38723536 |
| Heart | Ind. test | svr | 7.7771664 | 0.45078717 |
| Brain | Training | svr | 4.63735083 | 0.6590637 |
| Brain | Ind. test | svr | 5.65644408 | 0.66636339 |
| Eye | Training | svr | 6.4121788 | 0.16993856 |
| Eye | Ind. test | svr | 6.52493531 | 0.13236604 |
| Hepatic | Training | svr | 4.66034005 | 0.65023348 |
| Hepatic | Ind. test | svr | 8.49060491 | 0.61888877 |
| Renal | Training | svr | 6.2531785 | 0.24087996 |
| Renal | Ind. test | svr | 6.36008307 | 0.24339464 |
| Reproductive_male | Training | svr | 5.53587713 | 0.48564069 |
| Reproductive_male | Ind. test | svr | 7.06727651 | 0.4653191 |
| Endocrine | Training | svr | 4.86848409 | 0.6193155 |
| Endocrine | Ind. test | svr | 5.2220332 | 0.62839946 |
| Immune | Training | svr | 4.94458621 | 0.60410782 |
| Immune | Ind. test | svr | 6.56786232 | 0.556987 |
| Skin | Training | svr | 5.8491002 | 0.38758097 |
| Skin | Ind. test | svr | 6.14092739 | 0.39687219 |
| Reproductive_female | Training | lasso | 6.01990917 | 0.34773675 |
| Reproductive_female | Ind. test | lasso | 6.30463135 | 0.41722829 |
| Pulmonary | Training | lasso | 5.76098417 | 0.42359797 |
| Pulmonary | Ind. test | lasso | 5.77063271 | 0.44909376 |
| Heart | Training | lasso | 5.93206564 | 0.38571369 |
| Heart | Ind. test | lasso | 7.00113562 | 0.44689485 |
| Brain | Training | lasso | 4.65728178 | 0.66333101 |
| Brain | Ind. test | lasso | 5.7564748 | 0.66637912 |
| Eye | Training | lasso | 6.39042697 | 0.17813699 |
| Eye | Ind. test | lasso | 6.58410894 | 0.09628249 |
| Hepatic | Training | lasso | 4.73716085 | 0.65173784 |
| Hepatic | Ind. test | lasso | 5.56640792 | 0.63062858 |
| Renal | Training | lasso | 6.28482581 | 0.23930918 |
| Renal | Ind. test | lasso | 6.27430335 | 0.26557103 |
| Reproductive_male | Training | lasso | 5.57365101 | 0.4875842 |
| Reproductive_male | Ind. test | lasso | 6.32156164 | 0.46372775 |
| Endocrine | Training | lasso | 4.90511008 | 0.62089172 |
| Endocrine | Ind. test | lasso | 5.17779923 | 0.62814763 |
| Immune | Training | lasso | 4.96017428 | 0.60819404 |
| Immune | Ind. test | lasso | 6.59580643 | 0.5569291 |
| Skin | Training | lasso | 5.87835848 | 0.38813625 |
| Skin | Ind. test | lasso | 6.03622279 | 0.39676416 |
| Reproductive_female | Training | nn | 5.83139788 | 0.39359695 |
| Reproductive_female | Ind. test | nn | 6.69302985 | 0.44250494 |
| Pulmonary | Training | nn | 5.64647227 | 0.44430181 |
| Pulmonary | Ind. test | nn | 5.76512078 | 0.45341088 |
| Heart | Training | nn | 5.71610384 | 0.43175327 |
| Heart | Ind. test | nn | 7.03096169 | 0.42391373 |
| Brain | Training | nn | 4.33149819 | 0.69451192 |

| | | | | |
|---|---|---|---|---|
| Brain | Ind. test | nn | 4.86072404 | 0.64649974 |
| Eye | Training | nn | 6.39909597 | 0.16990408 |
| Eye | Ind. test | nn | 6.77741838 | 0.12981237 |
| Hepatic | Training | nn | 4.51712292 | 0.67045638 |
| Hepatic | Ind. test | nn | 10.1979413 | 0.6057521 |
| Renal | Training | nn | 6.16728536 | 0.27803434 |
| Renal | Ind. test | nn | 6.39473452 | 0.2639637 |
| Reproductive_male | Training | nn | 5.25469234 | 0.540281 |
| Reproductive_male | Ind. test | nn | 6.60320145 | 0.49581386 |
| Endocrine | Training | nn | 4.6419709 | 0.64662078 |
| Endocrine | Ind. test | nn | 5.00420552 | 0.6239082 |
| Immune | Training | nn | 4.65947126 | 0.64352651 |
| Immune | Ind. test | nn | 7.30030081 | 0.53638486 |
| Skin | Training | nn | 5.76946166 | 0.40245339 |
| Skin | Ind. test | nn | 6.15225224 | 0.39743693 |

b) **After age bias correction**

| **Organ** | **Dataset** | **Model** | **MAE** | **r** |
|---|---|---|---|---|
| Reproductive_female | Training | svr | 2.39776922 | 0.92664071 |
| Reproductive_female | Ind. test | svr | 4.0817444 | 0.83709446 |
| Pulmonary | Training | svr | 2.09760081 | 0.94496487 |
| Pulmonary | Ind. test | svr | 2.65679853 | 0.91618328 |
| Heart | Training | svr | 2.57506373 | 0.91414 |
| Heart | Ind. test | svr | 3.63590713 | 0.85988929 |
| Brain | Training | svr | 3.06460847 | 0.89148935 |
| Brain | Ind. test | svr | 5.32951935 | 0.7637381 |
| Eye | Training | svr | 0.73470349 | 0.99278978 |
| Eye | Ind. test | svr | 0.91450498 | 0.98949015 |
| Hepatic | Training | svr | 3.21801797 | 0.88292658 |
| Hepatic | Ind. test | svr | 4.60023987 | 0.79862744 |
| Renal | Training | svr | 1.86940311 | 0.95579281 |
| Renal | Ind. test | svr | 2.08551474 | 0.94484391 |
| Reproductive_male | Training | svr | 2.91132954 | 0.89817926 |
| Reproductive_male | Ind. test | svr | 4.83627378 | 0.78107469 |
| Endocrine | Training | svr | 3.15443808 | 0.88898069 |
| Endocrine | Ind. test | svr | 4.19694787 | 0.82789062 |
| Immune | Training | svr | 3.0189385 | 0.89457886 |
| Immune | Ind. test | svr | 5.10266644 | 0.76410503 |
| Skin | Training | svr | 2.66989033 | 0.91526295 |
| Skin | Ind. test | svr | 3.98320289 | 0.83998484 |
| Reproductive_female | Training | lasso | 1.92738181 | 0.95074547 |
| Reproductive_female | Ind. test | lasso | 3.28836875 | 0.88529969 |
| Pulmonary | Training | lasso | 2.31849818 | 0.93362726 |
| Pulmonary | Ind. test | lasso | 3.02624626 | 0.89326741 |
| Heart | Training | lasso | 1.93286486 | 0.94877539 |
| Heart | Ind. test | lasso | 2.75995775 | 0.91161738 |
| Brain | Training | lasso | 2.95385292 | 0.89762687 |
| Brain | Ind. test | lasso | 5.211114 | 0.76738641 |
| Eye | Training | lasso | 1.05842823 | 0.98498285 |
| Eye | Ind. test | lasso | 1.39207557 | 0.97573181 |
| Hepatic | Training | lasso | 2.86013931 | 0.90431087 |
| Hepatic | Ind. test | lasso | 3.84637339 | 0.84428636 |
| Renal | Training | lasso | 1.34497825 | 0.97631656 |
| Renal | Ind. test | lasso | 1.39304529 | 0.97453238 |
| Reproductive_male | Training | lasso | 2.53569743 | 0.9201013 |

| | | | | |
|---|---|---|---|---|
| Reproductive_male | Ind. test | lasso | 4.20151639 | 0.82056009 |
| Endocrine | Training | lasso | 2.91085803 | 0.90326211 |
| Endocrine | Ind. test | lasso | 3.86339811 | 0.84850843 |
| Immune | Training | lasso | 2.87970169 | 0.90254394 |
| Immune | Ind. test | lasso | 5.05701879 | 0.76936607 |
| Skin | Training | lasso | 2.17036054 | 0.94157427 |
| Skin | Ind. test | lasso | 3.25414357 | 0.88445897 |
| Reproductive_female | Training | nn | 2.69137381 | 0.91530922 |
| Reproductive_female | Ind. test | nn | 4.3465821 | 0.82358999 |
| Pulmonary | Training | nn | 2.82917129 | 0.90736528 |
| Pulmonary | Ind. test | nn | 3.40468825 | 0.87569297 |
| Heart | Training | nn | 2.81560211 | 0.90826774 |
| Heart | Ind. test | nn | 3.38123682 | 0.874591 |
| Brain | Training | nn | 3.35620877 | 0.87863142 |
| Brain | Ind. test | nn | 4.26791781 | 0.82469056 |
| Eye | Training | nn | 1.45168463 | 0.97249359 |
| Eye | Ind. test | nn | 1.90108955 | 0.95687703 |
| Hepatic | Training | nn | 3.29312929 | 0.87964641 |
| Hepatic | Ind. test | nn | 3.88452377 | 0.84810163 |
| Renal | Training | nn | 2.08223895 | 0.9434552 |
| Renal | Ind. test | nn | 2.44876428 | 0.92490279 |
| Reproductive_male | Training | nn | 3.0972128 | 0.88907816 |
| Reproductive_male | Ind. test | nn | 4.96961795 | 0.77707802 |
| Endocrine | Training | nn | 3.35018979 | 0.87886601 |
| Endocrine | Ind. test | nn | 4.24517205 | 0.82375595 |
| Immune | Training | nn | 3.27047553 | 0.88424686 |
| Immune | Ind. test | nn | 5.46340475 | 0.75038402 |
| Skin | Training | nn | 2.72348142 | 0.91413666 |
| Skin | Ind. test | nn | 3.87705129 | 0.85361898 |

**eTable 3: The relationship between the beta coefficient and the number of epochs by associating the brain ProtBAG with 8 cognitive scores and age at death**

A linear regression model was applied to examine the association between the brain ProtBAG and cognition/age at death after combining the respective populations. Covariates such as age, sex, and dataset status (e.g., Training, Independent Test, and PT) were included in the model to account for potential confounding factors.

**a) 8 cognitive scores**

| Cognition | Beta | SE | Epoch | -log$_{10}$(P) |
|---|---|---|---|---|
| duration_to_complete_alphanumeric_path_trail_2_f6350_2_0 | 1.00E-05 | 0.00015992 | 50 | 0.02226832 |
| duration_to_complete_alphanumeric_path_trail_2_f6350_2_0 | 4.55E-05 | 0.00017223 | 100 | 0.10149717 |
| duration_to_complete_alphanumeric_path_trail_2_f6350_2_0 | 4.46E-05 | 0.00018274 | 150 | 0.0929076 |
| duration_to_complete_alphanumeric_path_trail_2_f6350_2_0 | 3.70E-05 | 0.00019664 | 200 | 0.07023304 |
| duration_to_complete_alphanumeric_path_trail_2_f6350_2_0 | 7.48E-05 | 0.0001985 | 250 | 0.15092401 |
| duration_to_complete_alphanumeric_path_trail_2_f6350_2_0 | 1.91E-05 | 0.00019512 | 300 | 0.03517732 |
| duration_to_complete_alphanumeric_path_trail_2_f6350_2_0 | 7.38E-05 | 0.00019798 | 500 | 0.14919847 |
| duration_to_complete_alphanumeric_path_trail_2_f6350_2_0 | 8.71E-05 | 0.00020167 | 1000 | 0.17657828 |
| duration_to_complete_alphanumeric_path_trail_2_f6350_2_0 | 1.05E-05 | 0.00020636 | 1500 | 0.01802069 |
| duration_to_complete_alphanumeric_path_trail_2_f6350_2_0 | 9.55E-05 | 0.00019159 | 2000 | 0.20892074 |
| duration_to_complete_alphanumeric_path_trail_2_f6350_2_0 | 3.47E-05 | 0.0002065 | 2500 | 0.06227215 |
| duration_to_complete_numeric_path_trail_1_f6348_2_0 | 0.00082878 | 0.00051325 | 50 | 0.97284453 |
| duration_to_complete_numeric_path_trail_1_f6348_2_0 | 0.00140784 | 0.00055245 | 100 | 1.96389572 |
| duration_to_complete_numeric_path_trail_1_f6348_2_0 | 0.00148365 | 0.0005862 | 150 | 1.94236942 |
| duration_to_complete_numeric_path_trail_1_f6348_2_0 | 0.00129214 | 0.00063097 | 200 | 1.39095638 |
| duration_to_complete_numeric_path_trail_1_f6348_2_0 | 0.00125931 | 0.00063699 | 250 | 1.31765203 |
| duration_to_complete_numeric_path_trail_1_f6348_2_0 | 0.00122733 | 0.00062611 | 300 | 1.30062352 |
| duration_to_complete_numeric_path_trail_1_f6348_2_0 | 0.00110244 | 0.00063538 | 500 | 1.08189631 |
| duration_to_complete_numeric_path_trail_1_f6348_2_0 | 0.00099881 | 0.0006473 | 1000 | 0.91039414 |
| duration_to_complete_numeric_path_trail_1_f6348_2_0 | 0.00100996 | 0.00066232 | 1500 | 0.8948885 |
| duration_to_complete_numeric_path_trail_1_f6348_2_0 | 0.00076694 | 0.00061502 | 2000 | 0.67269606 |
| duration_to_complete_numeric_path_trail_1_f6348_2_0 | 0.00088465 | 0.00066283 | 2500 | 0.73975357 |
| fluid_intelligence_score_f20016_2_0 | -0.0054258 | 0.01590833 | 50 | 0.13485787 |

| | | | | |
|---|---|---|---|---|
| fluid_intelligence_score_f20016_2_0 | -0.005068 | 0.01729724 | 100 | 0.11377216 |
| fluid_intelligence_score_f20016_2_0 | -0.0045783 | 0.01834211 | 150 | 0.09533764 |
| fluid_intelligence_score_f20016_2_0 | -0.0077254 | 0.01975233 | 200 | 0.15756023 |
| fluid_intelligence_score_f20016_2_0 | -0.0037381 | 0.01996795 | 250 | 0.06981313 |
| fluid_intelligence_score_f20016_2_0 | -0.0079033 | 0.01961542 | 300 | 0.16302725 |
| fluid_intelligence_score_f20016_2_0 | -0.0026135 | 0.01984606 | 500 | 0.04806274 |
| fluid_intelligence_score_f20016_2_0 | -0.0068265 | 0.02011973 | 1000 | 0.13406707 |
| fluid_intelligence_score_f20016_2_0 | -0.0036582 | 0.02065367 | 1500 | 0.06579409 |
| fluid_intelligence_score_f20016_2_0 | 0.00501929 | 0.01913969 | 2000 | 0.10064944 |
| fluid_intelligence_score_f20016_2_0 | 0.00393785 | 0.02049776 | 2500 | 0.07177766 |
| maximum_digits_remembered_correctly_f4282_2_0 | -0.0257551 | 0.03034651 | 50 | 0.40218834 |
| maximum_digits_remembered_correctly_f4282_2_0 | -0.0240616 | 0.03266354 | 100 | 0.3359375 |
| maximum_digits_remembered_correctly_f4282_2_0 | -0.0193249 | 0.03462912 | 150 | 0.23894166 |
| maximum_digits_remembered_correctly_f4282_2_0 | -0.0189212 | 0.03725902 | 200 | 0.21352878 |
| maximum_digits_remembered_correctly_f4282_2_0 | -0.0132085 | 0.03763685 | 250 | 0.1392731 |
| maximum_digits_remembered_correctly_f4282_2_0 | -0.0053619 | 0.03698108 | 300 | 0.05319073 |
| maximum_digits_remembered_correctly_f4282_2_0 | -0.0248063 | 0.03751309 | 500 | 0.29372486 |
| maximum_digits_remembered_correctly_f4282_2_0 | -0.0228933 | 0.0381457 | 1000 | 0.26086943 |
| maximum_digits_remembered_correctly_f4282_2_0 | -0.0162946 | 0.03904571 | 1500 | 0.16975166 |
| maximum_digits_remembered_correctly_f4282_2_0 | -0.0091626 | 0.03625493 | 2000 | 0.0966418 |
| maximum_digits_remembered_correctly_f4282_2_0 | -0.0128981 | 0.03903029 | 2500 | 0.13014123 |
| mean_time_to_correctly_identify_matches_f20023_2_0 | -7.53E-05 | 0.00031279 | 50 | 0.09168723 |
| mean_time_to_correctly_identify_matches_f20023_2_0 | 2.29E-05 | 0.00033873 | 100 | 0.0240858 |
| mean_time_to_correctly_identify_matches_f20023_2_0 | 0.00010095 | 0.00035911 | 150 | 0.10867059 |
| mean_time_to_correctly_identify_matches_f20023_2_0 | 0.00022347 | 0.00038711 | 200 | 0.24889027 |
| mean_time_to_correctly_identify_matches_f20023_2_0 | 0.00028366 | 0.0003912 | 250 | 0.32936279 |
| mean_time_to_correctly_identify_matches_f20023_2_0 | 0.00039962 | 0.00038476 | 300 | 0.52429046 |
| mean_time_to_correctly_identify_matches_f20023_2_0 | 0.00054511 | 0.0003897 | 500 | 0.79066401 |

| | | | | |
|---|---|---|---|---|
| mean_time_to_correctly_identify_matches_f20023_2_0 | 0.00044397 | 0.00039539 | 1000 | 0.58246468 |
| mean_time_to_correctly_identify_matches_f20023_2_0 | 0.00058972 | 0.00040567 | 1500 | 0.83538499 |
| mean_time_to_correctly_identify_matches_f20023_2_0 | 0.00040068 | 0.00037433 | 2000 | 0.5459442 |
| mean_time_to_correctly_identify_matches_f20023_2_0 | 0.00044658 | 0.00040129 | 2500 | 0.57542711 |
| number_of_puzzles_correct_f21004_2_0 | 0.00386429 | 0.01402095 | 50 | 0.10631272 |
| number_of_puzzles_correct_f21004_2_0 | 0.00140162 | 0.01509141 | 100 | 0.0333853 |
| number_of_puzzles_correct_f21004_2_0 | 0.00070989 | 0.01600485 | 150 | 0.01564186 |
| number_of_puzzles_correct_f21004_2_0 | 0.0011568 | 0.01722286 | 200 | 0.02390089 |
| number_of_puzzles_correct_f21004_2_0 | -0.0018796 | 0.01738643 | 250 | 0.03909281 |
| number_of_puzzles_correct_f21004_2_0 | -0.003882 | 0.01708731 | 300 | 0.08603063 |
| number_of_puzzles_correct_f21004_2_0 | -0.006902 | 0.01734845 | 500 | 0.16066815 |
| number_of_puzzles_correct_f21004_2_0 | -0.0061779 | 0.01766193 | 1000 | 0.13875244 |
| number_of_puzzles_correct_f21004_2_0 | -0.0074063 | 0.01806325 | 1500 | 0.16633151 |
| number_of_puzzles_correct_f21004_2_0 | -0.0076978 | 0.0167748 | 2000 | 0.18953881 |
| number_of_puzzles_correct_f21004_2_0 | -0.0045277 | 0.01808417 | 2500 | 0.09565363 |
| number_of_puzzles_correctly_solved_f6373_2_0 | 0.01203234 | 0.02128748 | 50 | 0.24263921 |
| number_of_puzzles_correctly_solved_f6373_2_0 | -0.0071752 | 0.02292411 | 100 | 0.12245568 |
| number_of_puzzles_correctly_solved_f6373_2_0 | -0.0036319 | 0.02431707 | 150 | 0.05488534 |
| number_of_puzzles_correctly_solved_f6373_2_0 | -0.0058741 | 0.02617045 | 200 | 0.08490824 |
| number_of_puzzles_correctly_solved_f6373_2_0 | -0.0087589 | 0.02642269 | 250 | 0.1305966 |
| number_of_puzzles_correctly_solved_f6373_2_0 | -0.0105321 | 0.02596574 | 300 | 0.16427654 |
| number_of_puzzles_correctly_solved_f6373_2_0 | -0.0076389 | 0.02635667 | 500 | 0.11240231 |
| number_of_puzzles_correctly_solved_f6373_2_0 | -0.0119399 | 0.02684266 | 1000 | 0.18277557 |
| number_of_puzzles_correctly_solved_f6373_2_0 | -0.0028653 | 0.02746697 | 1500 | 0.03766743 |
| number_of_puzzles_correctly_solved_f6373_2_0 | -0.0038259 | 0.02550319 | 2000 | 0.05514245 |
| number_of_puzzles_correctly_solved_f6373_2_0 | -0.0024999 | 0.02748848 | 2500 | 0.03266597 |
| number_of_symbol_digit_matches_made_correctly_f23324_2_0 | -0.0032996 | 0.00896907 | 50 | 0.14692212 |
| number_of_symbol_digit_matches_made_correctly_f23324_2_0 | -0.0220887 | 0.00965332 | 100 | 1.65390226 |

| | | | | |
|---|---|---|---|---|
| number_of_symbol_digit_matches_made_correctly_f23324_2_0 | -0.0251473 | 0.01023961 | 150 | 1.85068076 |
| number_of_symbol_digit_matches_made_correctly_f23324_2_0 | -0.0297129 | 0.01101675 | 200 | 2.15306925 |
| number_of_symbol_digit_matches_made_correctly_f23324_2_0 | -0.029885 | 0.01112256 | 250 | 2.13983331 |
| number_of_symbol_digit_matches_made_correctly_f23324_2_0 | -0.0312221 | 0.01093147 | 300 | 2.36514757 |
| number_of_symbol_digit_matches_made_correctly_f23324_2_0 | -0.0307388 | 0.01109437 | 500 | 2.24993364 |
| number_of_symbol_digit_matches_made_correctly_f23324_2_0 | -0.0333073 | 0.0112985 | 1000 | 2.49206258 |
| number_of_symbol_digit_matches_made_correctly_f23324_2_0 | -0.0306259 | 0.01156833 | 1500 | 2.08890672 |
| number_of_symbol_digit_matches_made_correctly_f23324_2_0 | -0.0260268 | 0.01073521 | 2000 | 1.81293756 |
| number_of_symbol_digit_matches_made_correctly_f23324_2_0 | -0.0245552 | 0.01158159 | 2500 | 1.46773243 |

#### b) Age at death

| Variable | Beta | SE | Epoch | $-\log_{10}(P)$ |
|---|---|---|---|---|
| age_at_death_f40007_0_0 | -0.0276447 | 0.01070479 | 50 | 2.00706993 |
| age_at_death_f40007_0_0 | -0.0655473 | 0.01228292 | 100 | 7.00415659 |
| age_at_death_f40007_0_0 | -0.0744235 | 0.01313374 | 150 | 7.81248708 |
| age_at_death_f40007_0_0 | -0.0945895 | 0.01461604 | 200 | 9.97295271 |
| age_at_death_f40007_0_0 | -0.090036 | 0.01467968 | 250 | 9.03250229 |
| age_at_death_f40007_0_0 | -0.0917235 | 0.01468873 | 300 | 9.33617649 |
| age_at_death_f40007_0_0 | -0.1122137 | 0.01544037 | 500 | 12.3727765 |
| age_at_death_f40007_0_0 | -0.1088401 | 0.0147478 | 1000 | 12.7332291 |
| age_at_death_f40007_0_0 | -0.1138174 | 0.01517105 | 1500 | 13.1304976 |
| age_at_death_f40007_0_0 | -0.0864915 | 0.01229123 | 2000 | 11.6502271 |
| age_at_death_f40007_0_0 | -0.08222 | 0.01319252 | 2500 | 11.4523289 |

### eTable 4: Genomic loci of the 11 ProtBAGs and 9 PhenoBAGs defined by FUMA

| TopLeadSNP | Chromosome | Position | P-value | MappedGene | BAG | BAG_type | Cytogenetic_region |
|---|---|---|---|---|---|---|---|
| rs76560665 | 1 | 180956936 | 1.57E-15 | ['KIAA1614', 'AL162431.1', 'STX6', 'MR1'] | Brain_age_gap_brain | PhenoBAG | 1q25.3 |
| rs6442411 | 3 | 13836296 | 1.85E-10 | ['WNT7A'] | Brain_age_gap_brain | PhenoBAG | 3p25.1 |
| rs11708828 | 3 | 39499649 | 2.71E-10 | ['SLC25A38', 'RPSA', 'MOBP'] | Brain_age_gap_brain | PhenoBAG | 3p22.1 |
| rs12263364 | 10 | 134555548 | 4.44E-09 | ['INPP5A', 'NKX6-2'] | Brain_age_gap_brain | PhenoBAG | 10q26.3 |
| rs1470 | 16 | 70127276 | 2.43E-11 | ['WWP2', 'CLEC18A', 'PDPR', 'CLEC18C', 'AC009060.1', 'FKSG63', 'EXOSC6', 'AARS', 'DDX19B', 'RP11-529K1.3', 'DDX19A', 'ST3GAL2', 'FUK', 'COG4', 'SF3B3', 'CMTR2'] | Brain_age_gap_brain | PhenoBAG | 16q22.1 |
| rs371185851 | 17 | 43568278 | 8.76E-17 | ['C17orf104', 'NMT1', 'FMNL1', 'ARHGAP27', 'PLEKHM1', 'CRHR1', 'SPPL2C', 'MAPT', 'STH', 'KANSL1', 'ARL17B', 'LRRC37A', 'LRRC37A2', 'ARL17A', 'NSF', 'WNT3', 'EFCAB13'] | Brain_age_gap_brain | PhenoBAG | 17q21.31 |
| rs769449 | 19 | 45410002 | 1.29E-10 | ['PVRL2', 'TOMM40', 'APOE', 'APOC1'] | Brain_age_gap_brain | PhenoBAG | 19q13.32 |
| rs142005893 | 1 | 11873512 | 6.62E-10 | ['C1orf167', 'MTHFR', 'CLCN6', 'NPPA'] | Cardiovascular_age_gap_brain | PhenoBAG | 1p36.22 |
| rs28611172 | 1 | 38410321 | 1.78E-09 | ['MTF1', 'INPP5B', 'SF3A3', 'FHL3'] | Cardiovascular_age_gap_brain | PhenoBAG | 1p34.3 |
| rs12733512 | 1 | 59646978 | 3.20E-17 | | Cardiovascular_age_gap_brain | PhenoBAG | 1p32.1 |
| rs55779904 | 2 | 19723238 | 2.68E-17 | | Cardiovascular_age_gap_brain | PhenoBAG | 2p24.1 |
| rs67343025 | 2 | 164934626 | 4.50E-12 | | Cardiovascular_age_gap_brain | PhenoBAG | 2q24.3 |
| rs1250259 | 2 | 216300482 | 5.19E-10 | ['FN1'] | Cardiovascular_age_gap_brain | PhenoBAG | 2q35 |
| rs9848992 | 3 | 41875297 | 8.24E-16 | ['ULK4', 'TRAK1'] | Cardiovascular_age_gap_brain | PhenoBAG | 3p22.1 |
| rs12509595 | 4 | 81182554 | 8.44E-12 | ['PAQR3', 'FGF5', 'BMP3'] | Cardiovascular_age_gap_brain | PhenoBAG | 4q21.21 |
| rs5863154 | 4 | 156410257 | 3.54E-09 | | Cardiovascular_age_gap_brain | PhenoBAG | 4q32.1 |
| rs1173727 | 5 | 32830521 | 6.99E-12 | ['PDZD2', 'NPR3', 'AC026703.1'] | Cardiovascular_age_gap_brain | PhenoBAG | 5p13.3 |
| rs2964325 | 5 | 157726670 | 6.99E-10 | | Cardiovascular_age_gap_brain | PhenoBAG | 5q33.3 |
| rs4704968 | 5 | 158264020 | 8.42E-11 | ['EBF1'] | Cardiovascular_age_gap_brain | PhenoBAG | 5q33.3 |
| rs546130170 | 6 | 56010655 | 2.35E-11 | ['COL21A1'] | Cardiovascular_age_gap_brain | PhenoBAG | 6p12.1 |
| rs5880952 | 6 | 152319503 | 1.48E-13 | ['ESR1'] | Cardiovascular_age_gap_brain | PhenoBAG | 6q25.1 |
| rs419009 | 6 | 159692596 | 1.11E-13 | ['FNDC1'] | Cardiovascular_age_gap_brain | PhenoBAG | 6q25.3 |
| rs11759438 | 6 | 169633335 | 8.65E-11 | ['THBS2'] | Cardiovascular_age_gap_brain | PhenoBAG | 6q27 |
| rs2107595 | 7 | 19049388 | 6.50E-10 | ['HDAC9', 'TWIST1'] | Cardiovascular_age_gap_brain | PhenoBAG | 7p21.1 |
| rs75487554 | 7 | 28618547 | 3.82E-09 | ['CREB5'] | Cardiovascular_age_gap_brain | PhenoBAG | 7p15.1 |
| rs10260816 | 7 | 46010100 | 2.85E-20 | ['IGFBP3'] | Cardiovascular_age_gap_brain | PhenoBAG | 7p12.3 |
| rs10269774 | 7 | 92253972 | 9.55E-16 | ['RBM48', 'FAM133B', 'CDK6'] | Cardiovascular_age_gap_brain | PhenoBAG | 7q21.2 |
| rs17477177 | 7 | 106411858 | 1.37E-38 | | Cardiovascular_age_gap_brain | PhenoBAG | 7q22.3 |
| rs2071526 | 8 | 120426597 | 3.26E-17 | ['MAL2', 'NOV', 'ENPP2'] | Cardiovascular_age_gap_brain | PhenoBAG | 8q24.12 |
| rs7107356 | 11 | 47676170 | 1.86E-10 | ['NR1H3', 'SPI1', 'SLC39A13', 'CELF1', 'NDUFS3', 'PTPMT1', 'KBTBD4', 'KBTBD4', 'FAM180B', 'C1QTNF4', 'MTCH2', 'AGBL2', 'FNBP4', 'NUP160', 'PTPRJ'] | Cardiovascular_age_gap_brain | PhenoBAG | 11p11.2 |
| rs7936928 | 11 | 130279168 | 1.83E-26 | ['ADAMTS8', 'ADAMTS15'] | Cardiovascular_age_gap_brain | PhenoBAG | 11q24.3 |
| rs10841385 | 12 | 20007757 | 5.50E-10 | | Cardiovascular_age_gap_brain | PhenoBAG | 12p12.2 |
| rs1401981 | 12 | 90091446 | 6.85E-11 | ['ATP2B1'] | Cardiovascular_age_gap_brain | PhenoBAG | 12q21.33 |
| rs452036 | 14 | 23865885 | 7.78E-12 | ['MYH6', 'MYH7'] | Cardiovascular_age_gap_brain | PhenoBAG | 14q11.2 |
| rs4775769 | 15 | 48939888 | 5.08E-13 | ['FBN1', 'CEP152', 'SHC4'] | Cardiovascular_age_gap_brain | PhenoBAG | 15q21.1 |
| rs77870048 | 16 | 69965021 | 6.72E-24 | ['NFAT5', 'NOB1', 'WWP2', 'CLEC18A', 'EXOSC6', 'AARS', 'DDX19B', 'RP11-529K1.3', 'DDX19A', 'ST3GAL2', 'FUK', 'COG4', 'IL34', 'MTSS1L', 'FLJ00418', 'VAC14', 'HYDIN', 'CALB2', 'MARVELD3', 'PHLPP2', 'AP1G1', 'ATXN1L', 'IST1', 'ZNF821', 'TXNL4B', 'HP', 'HPR', 'PMFBP1', 'ZFHX3'] | Cardiovascular_age_gap_brain | PhenoBAG | 16q22.1 |
| rs7500448 | 16 | 83045790 | 5.49E-10 | ['CDH13'] | Cardiovascular_age_gap_brain | PhenoBAG | 16q23.3 |
| rs12603813 | 17 | 43196584 | 2.68E-09 | ['C1QL1', 'DCAKD', 'NMT1', 'PLCD3', 'ACBD4', | Cardiovascular_age_gap_brain | PhenoBAG | 17q21.31 |

| | | | | | | | |
|---|---|---|---|---|---|---|---|
| | | | | 'HEXIM1', 'HEXIM2', 'SPATA32'] | | | |
| rs80335285 | 17 | 45055107 | 4.27E-09 | ['KANSL1', 'LRRC37A2', 'WNT3', 'WNT9B', 'GOSR2', 'RP11-156P1.2', 'RPRML', 'CDC27'] | Cardiovascular_age_gap_brain | PhenoBAG | 17q21.32 |
| rs7236548 | 18 | 43097750 | 4.35E-12 | ['SLC14A2'] | Cardiovascular_age_gap_brain | PhenoBAG | 18q12.3 |
| rs2206815 | 20 | 10669188 | 4.19E-13 | ['JAG1'] | Cardiovascular_age_gap_brain | PhenoBAG | 20p12.2 |
| rs10801558 | 1 | 196699044 | 1.18E-13 | ['KCNT2', 'CFH', 'CFHR1', 'CFHR2', 'CFHR4'] | Eye_age_gap_brain | PhenoBAG | 1q31.3 |
| rs17279437 | 3 | 45814094 | 1.16E-24 | ['LARS2', 'LIMD1', 'SACM1L', 'SLC6A20', 'LZTFL1', 'CCR9', 'FYCO1', 'ALS2CL'] | Eye_age_gap_brain | PhenoBAG | 3p21.31 |
| rs1970896 | 4 | 184943118 | 1.78E-09 | ['STOX2'] | Eye_age_gap_brain | PhenoBAG | 4q35.1 |
| rs2004187 | 5 | 2612747 | 1.10E-14 | | Eye_age_gap_brain | PhenoBAG | 5p15.33 |
| rs35259275 | 6 | 70414557 | 1.27E-09 | ['LMBRD1'] | Eye_age_gap_brain | PhenoBAG | 6q13 |
| rs1536057 | 6 | 108885623 | 3.92E-15 | ['FOXO3', 'ARMC2', 'SESN1', 'CEP57L1'] | Eye_age_gap_brain | PhenoBAG | 6q21 |
| rs2345701 | 7 | 46634598 | 8.74E-16 | | Eye_age_gap_brain | PhenoBAG | 7p12.3 |
| rs13309489 | 7 | 51011324 | 1.09E-13 | ['GRB10'] | Eye_age_gap_brain | PhenoBAG | 7p12.1 |
| rs393155 | 8 | 9033093 | 1.81E-10 | ['MFHAS1', 'ERI1', 'RP11-10A14.4', 'TNKS', 'MSRA', 'PRSS55', 'RP1L1', 'SOX7', 'XKR6', 'AF131215.5', 'MTMR9', 'SLC35G5', 'C8orf12', 'FAM167A', 'BLK', 'GATA4', 'C8orf49', 'NEIL2', 'CTSB', 'DEFB136', 'DEFB135', 'DEFB134', 'RP11-481A20.11', 'USP17L2', 'FAM86B1', 'DEFB130', 'FAM86B2'] | Eye_age_gap_brain | PhenoBAG | 8p23.1 |
| rs4871827 | 8 | 121061879 | 2.57E-15 | ['TAF2', 'DSCC1', 'DEPTOR', 'COL14A1'] | Eye_age_gap_brain | PhenoBAG | 8q24.12 |
| rs10830240 | 11 | 88938356 | 2.41E-20 | ['GRM5', 'TYR', 'NOX4'] | Eye_age_gap_brain | PhenoBAG | 11q14.3 |
| rs1010053 | 14 | 61005625 | 4.49E-10 | ['C14orf39', 'SIX6', 'SIX1'] | Eye_age_gap_brain | PhenoBAG | 14q23.1 |
| rs1800407 | 15 | 28230318 | 2.25E-37 | ['OCA2', 'HERC2'] | Eye_age_gap_brain | PhenoBAG | 15q13.1 |
| rs2010428 | 16 | 71772115 | 2.86E-09 | ['ZNF23', 'AC010547.9', 'ZNF19', 'MARVELD3', 'PHLPP2', 'AP1G1', 'ATXN1L', 'IST1', 'ZNF821'] | Eye_age_gap_brain | PhenoBAG | 16q22.2 |
| rs9747093 | 17 | 79584280 | 5.73E-32 | ['RP11-1055B8.7', 'ACTG1', 'FSCN2', 'C17orf70', 'NPLOC4', 'PDE6G', 'OXLD1', 'CCDC137', 'ARL16', 'HGS', 'MRPL12', 'SLC25A10', 'SLC25A10'] | Eye_age_gap_brain | PhenoBAG | 17q25.3 |
| rs149344982 | 1 | 21889760 | 2.50E-85 | ['EIF4G3', 'ECE1', 'NBPF3', 'ALPL', 'RAP1GAP', 'USP48'] | Hepatic_age_gap_brain | PhenoBAG | 1p36.12 |
| rs114384494 | 1 | 161653554 | 1.17E-11 | ['FCGR2A', 'HSPA6', 'FCGR3A', 'FCGR2B', 'FCRLA'] | Hepatic_age_gap_brain | PhenoBAG | 1q23.3 |
| rs2093770 | 1 | 178604633 | 4.27E-09 | ['TEX35', 'C1orf220', 'C1ORF220'] | Hepatic_age_gap_brain | PhenoBAG | 1q25.2 |
| rs28899170 | 2 | 234604230 | 6.28E-22 | ['USP40', 'UGT1A8', 'UGT1A10', 'UGT1A9', 'UGT1A7', 'UGT1A6', 'UGT1A5', 'UGT1A4', 'UGT1A3', 'UGT1A1', 'MROH2A'] | Hepatic_age_gap_brain | PhenoBAG | 2q37.1 |
| rs146948889 | 3 | 48477252 | 4.61E-14 | ['SCAP', 'SMARCC1', 'DHX30', 'MAP4', 'CDC25A', 'ZNF589', 'NME6', 'SPINK8', 'FBXW12', 'PLXNB1', 'CCDC51', 'TMA7', 'ATRIP', 'TREX1', 'PFKFB4', 'UCN2', 'COL7A1', 'UQCRC1', 'TMEM89', 'SLC26A6', 'IP6K2', 'PRKAR2A', 'SLC25A20', 'ARIH2OS', 'ARIH2', 'QRICH1', | Hepatic_age_gap_brain | PhenoBAG | 3p21.31 |

| | | | | | | | |
|---|---|---|---|---|---|---|---|
| | | | | 'QARS', 'CCDC36', 'GPX1', 'RHOA', 'DAG1', 'BSN', 'RNF123', 'IP6K1', 'CDHR4', 'MST1R', 'CTD-2330K9.3', 'MON1A', 'RBM6', 'RBM5', 'SEMA3F', 'GNAI2', 'HYAL1', 'HYAL2', 'TUSC2', 'ZMYND10', 'NPRL2', 'CYB561D2', 'XXcos-LUCA11.5', 'TMEM115', 'CACNA2D2', 'C3orf18', 'HEMK1', 'MANF', 'RBM15B'] | | | |
| rs4228 | 3 | 58413669 | 2.48E-11 | ['ABHD6', 'RPP14', 'RPP14', 'PXK', 'PDHB', 'KCTD6', 'ACOX2'] | Hepatic_age_gap_brain | PhenoBAG | 3p14.3 |
| rs80120242 | 3 | 132235344 | 2.17E-11 | ['DNAJC13', 'ACAD11', 'NPHP3'] | Hepatic_age_gap_brain | PhenoBAG | 3q22.1 |
| rs166304 | 5 | 72351708 | 5.25E-17 | ['MAP1B', 'TNPO1', 'FCHO2', 'TMEM171'] | Hepatic_age_gap_brain | PhenoBAG | 5q13.2 |
| rs62401887 | 6 | 24416482 | 8.63E-25 | ['NRSN1', 'DCDC2', 'MRS2', 'GPLD1', 'ALDH5A1', 'KIAA0319', 'C6orf62'] | Hepatic_age_gap_brain | PhenoBAG | 6p22.3 |
| rs13234131 | 7 | 73025975 | 3.83E-09 | ['POM121', 'FZD9', 'BAZ1B', 'BCL7B', 'TBL2', 'MLXIPL', 'VPS37D'] | Hepatic_age_gap_brain | PhenoBAG | 7q11.23 |
| rs115946508 | 7 | 150497496 | 5.37E-10 | ['TMEM176B', 'TMEM176A'] | Hepatic_age_gap_brain | PhenoBAG | 7q36.1 |
| rs4841133 | 8 | 9183664 | 3.47E-26 | | Hepatic_age_gap_brain | PhenoBAG | 8p23.1 |
| rs2062377 | 8 | 120007420 | 1.49E-09 | ['TNFRSF11B', 'COLEC10'] | Hepatic_age_gap_brain | PhenoBAG | 8q24.12 |
| rs149092047 | 9 | 136139907 | 6.24E-165 | ['RALGDS', 'GBGT1', 'OBP2B', 'SURF6', 'MED22', 'RPL7A', 'SURF1', 'SURF2', 'SURF4', 'C9orf96', 'REXO4', 'ADAMTS13', 'CACFD1', 'SLC2A6', 'TMEM8C', 'ADAMTSL2'] | Hepatic_age_gap_brain | PhenoBAG | 9q34.2 |
| rs4307445 | 9 | 139279776 | 3.87E-09 | ['GPSM1', 'DNLZ', 'CARD9', 'SNAPC4', 'SDCCAG3', 'PMPCA', 'INPP5E', 'SEC16A', 'C9orf163', 'NOTCH1'] | Hepatic_age_gap_brain | PhenoBAG | 9q34.3 |
| rs553304 | 10 | 18266410 | 3.25E-11 | ['TMEM236', 'MRC1L1', 'TMEM236', 'MRC1', 'SLC39A12'] | Hepatic_age_gap_brain | PhenoBAG | 10p12.33 |
| rs10509186 | 10 | 65207018 | 1.10E-42 | ['NRBF2', 'JMJD1C', 'REEP3'] | Hepatic_age_gap_brain | PhenoBAG | 10q21.3 |
| rs76998820 | 10 | 103877902 | 1.05E-11 | ['NPM3', 'MGEA5', 'KCNIP2', 'C10orf76', 'HPS6', 'LDB1', 'PPRC1', 'NOLC1', 'ELOVL3', 'PITX3', 'GBF1', 'NFKB2', 'PSD', 'FBXL15', 'CUEDC2', 'ACTR1A', 'SUFU'] | Hepatic_age_gap_brain | PhenoBAG | 10q24.32 |
| rs7949566 | 11 | 126285301 | 1.23E-10 | ['DCPS', 'ST3GAL4', 'KIRREL3'] | Hepatic_age_gap_brain | PhenoBAG | 11q24.2 |
| rs7955258 | 12 | 570947 | 2.27E-12 | ['CCDC77', 'B4GALNT3'] | Hepatic_age_gap_brain | PhenoBAG | 12p13.33 |
| rs10498635 | 14 | 93103309 | 1.50E-09 | ['RIN3'] | Hepatic_age_gap_brain | PhenoBAG | 14q32.12 |
| rs28929474 | 14 | 94844947 | 1.98E-16 | ['PPP4R4', 'SERPINA10', 'SERPINA6', 'SERPINA1'] | Hepatic_age_gap_brain | PhenoBAG | 14q32.13 |
| rs139097404 | 15 | 43933941 | 3.19E-22 | ['STARD9', 'TTBK2', 'UBR1', 'EPB42', 'CCNDBP1', 'TGM7', 'LCMT2', 'ADAL', 'ZSCAN29', 'TUBGCP4', 'TP53BP1', 'MAP1A', 'PPIP5K1', 'CKMT1B', 'STRC', 'CATSPER2', 'CKMT1A', 'PDIA3', 'ELL3', 'RP11-296A16.1', 'SERF2', 'AC018512.1', 'SERINC4', 'HYPK', 'MFAP1', 'WDR76', 'FRMD5', 'CASC4', 'CTDSPL2', 'EIF3J', 'SPG11'] | Hepatic_age_gap_brain | PhenoBAG | 15q15.3 |

| | | | | | | | |
|---|---|---|---|---|---|---|---|
| rs11078597 | 17 | 1618363 | 2.82E-22 | ['TLCD2', 'WDR81', 'SERPINF2', 'SERPINF1'] | Hepatic_age_gap_brain | PhenoBAG | 17p13.3 |
| rs55714927 | 17 | 7080316 | 2.75E-22 | ['CLEC10A', 'ASGR2', 'ASGR1', 'DLG4', 'ACADVL', 'DVL2', 'PHF23', 'GABARAP', 'CTD-2545G14.7', 'CTDNEP1', 'RP1-4G17.5', 'ELP5', 'CLDN7', 'SLC2A4'] | Hepatic_age_gap_brain | PhenoBAG | 17p13.1 |
| rs77542162 | 17 | 67081278 | 2.15E-31 | ['ABCA8', 'ABCA9', 'ABCA6', 'ABCA10', 'ABCA5', 'MAP2K6'] | Hepatic_age_gap_brain | PhenoBAG | 17q24.2 |
| rs883921 | 18 | 60068549 | 3.50E-10 | ['PIGN', 'TNFRSF11A'] | Hepatic_age_gap_brain | PhenoBAG | 18q21.33 |
| rs273488 | 19 | 18236654 | 4.71E-09 | ['IL12RB1', 'MAST3', 'PIK3R2', 'PIK3R2', 'IFI30', 'MPV17L2', 'RAB3A', 'PDE4C', 'KIAA1683'] | Hepatic_age_gap_brain | PhenoBAG | 19p13.11 |
| rs58542926 | 19 | 19379549 | 2.36E-45 | ['UBA52', 'SUGP2', 'ARMC6', 'SLC25A42', 'MEF2BNB-MEF2B', 'MEF2B', 'MEF2BNB', 'RFXANK', 'NR2C2AP', 'NCAN', 'HAPLN4', 'TM6SF2', 'SUGP1', 'MAU2', 'GATAD2A', 'TSSK6', 'NDUFA13', 'YJEFN3', 'CTC-260F20.3', 'CILP2', 'PBX4', 'LPAR2', 'GMIP', 'ATP13A1', 'ZNF101', 'ZNF14', 'ZNF90', 'ZNF486'] | Hepatic_age_gap_brain | PhenoBAG | 19p13.11 |
| rs45512696 | 19 | 35550878 | 3.41E-27 | ['HPN', 'TMEM147'] | Hepatic_age_gap_brain | PhenoBAG | 19q13.12 |
| rs429358 | 19 | 45411941 | 2.29E-18 | ['PVRL2', 'TOMM40', 'APOE', 'APOC1'] | Hepatic_age_gap_brain | PhenoBAG | 19q13.32 |
| rs35728097 | 19 | 49213853 | 1.94E-14 | ['FAM83E', 'NTN5', 'FUT2', 'MAMSTR', 'RASIP1', 'IZUMO1', 'FUT1', 'FGF21'] | Hepatic_age_gap_brain | PhenoBAG | 19q13.33 |
| rs113886122 | 19 | 50044741 | 1.29E-21 | ['ALDH16A1', 'CTD-3148I10.9', 'FLT3LG', 'RPL13A', 'RPS11', 'hsa-mir-150', 'FCGRT', 'RCN3', 'NOSIP', 'PRRG2', 'PRR12', 'RRAS', 'SCAF1', 'IRF3', 'CPT1C', 'TSKS', 'AP2A1'] | Hepatic_age_gap_brain | PhenoBAG | 19q13.33 |
| rs3806401 | 1 | 43426212 | 1.03E-12 | ['SLC2A1'] | Immune_age_gap_brain | PhenoBAG | 1p34.2 |
| rs139795227 | 1 | 92842367 | 2.39E-10 | ['TGFBR3', 'GLMN', 'RPAP2'] | Immune_age_gap_brain | PhenoBAG | 1p22.1 |
| rs11102809 | 1 | 115064222 | 1.10E-09 | ['TRIM33'] | Immune_age_gap_brain | PhenoBAG | 1p13.2 |
| rs857693 | 1 | 158556301 | 9.28E-25 | ['OR10K2', 'OR10K1', 'OR10R2', 'OR6Y1', 'OR6P1', 'OR10X1', 'OR10Z1', 'SPTA1', 'OR6K6'] | Immune_age_gap_brain | PhenoBAG | 1q23.1 |
| rs57346671 | 1 | 205260165 | 2.56E-15 | ['NFASC', 'CNTN2', 'TMEM81', 'RBBP5', 'DSTYK', 'TMCC2', 'NUAK2'] | Immune_age_gap_brain | PhenoBAG | 1q32.1 |
| rs3811444 | 1 | 248039451 | 3.62E-59 | ['OR14A16', 'OR11L1', 'TRIM58', 'OR2W3', 'OR2T8', 'OR2T33'] | Immune_age_gap_brain | PhenoBAG | 1q44 |
| rs6730558 | 2 | 8756183 | 1.36E-18 | | Immune_age_gap_brain | PhenoBAG | 2p25.1 |
| rs11925835 | 3 | 56865445 | 4.75E-26 | ['ARHGEF3'] | Immune_age_gap_brain | PhenoBAG | 3p14.3 |
| rs9828178 | 3 | 142171303 | 1.42E-14 | ['XRN1', 'ATR', 'PLS1', 'TRPC1', 'PCOLCE2', 'PAQR9', 'U2SURP'] | Immune_age_gap_brain | PhenoBAG | 3q23 |
| rs4243419 | 3 | 171507381 | 1.47E-09 | ['TNIK', 'PLD1', 'PP13439'] | Immune_age_gap_brain | PhenoBAG | 3q26.31 |
| rs2736100 | 5 | 1286516 | 1.28E-10 | ['TERT'] | Immune_age_gap_brain | PhenoBAG | 5p15.33 |
| rs144215134 | 5 | 76069008 | 3.93E-11 | ['IQGAP2', 'F2R'] | Immune_age_gap_brain | PhenoBAG | 5q13.3 |
| rs2617617 | 5 | 127455008 | 1.41E-19 | ['SLC12A2'] | Immune_age_gap_brain | PhenoBAG | 5q23.3 |
| rs80215559 | 6 | 25918225 | 6.20E-26 | ['LRRC16A', 'SCGN', 'HIST1H2AA', 'HIST1H2BA', 'SLC17A4', 'SLC17A1', 'SLC17A3', 'SLC17A2', 'TRIM38', 'HIST1H1A', 'HIST1H3A', 'HIST1H4A', 'HIST1H4B', 'HIST1H3B', 'HIST1H2AB', 'HIST1H2BB', 'HIST1H3C', | Immune_age_gap_brain | PhenoBAG | 6p22.2 |

| | | | | | | | |
|---|---|---|---|---|---|---|---|
| | | | | 'HIST1H1C', 'HFE', 'HIST1H4C', 'HIST1H1T', 'HIST1H2BC', 'HIST1H2AC'] | | | |
| rs80215559 | 6 | 25918225 | 6.20E-26 | ['BTN2A1'] | Immune_age_gap_brain | PhenoBAG | 6p22.2 |
| rs13194491 | 6 | 27037080 | 2.16E-09 | ['ZNF322', 'HIST1H2BK', 'HIST1H2AH', 'PRSS16', 'HIST1H3H', 'HIST1H2AJ', 'HIST1H2BM', 'HIST1H4J', 'HIST1H4K', 'HIST1H2AK', 'HIST1H3J', 'HIST1H2AM', 'HIST1H2BO'] | Immune_age_gap_brain | PhenoBAG | 6p22.1 |
| rs9471708 | 6 | 41956353 | 1.14E-21 | ['OARD1', 'NFYA', 'FOXP4', 'PGC', 'FRS3', 'PRICKLE4', 'TOMM6', 'USP49', 'MED20', 'BYSL', 'CCND3', 'GUCA1A', 'RP1-139D8.6', 'GUCA1B', 'MRPS10'] | Immune_age_gap_brain | PhenoBAG | 6p21.1 |
| rs11755527 | 6 | 90958231 | 6.04E-11 | ['BACH2'] | Immune_age_gap_brain | PhenoBAG | 6q15 |
| rs592423 | 6 | 139840693 | 1.04E-24 | | Immune_age_gap_brain | PhenoBAG | 6q24.1 |
| rs4316067 | 7 | 33081514 | 4.40E-09 | ['AVL9', 'FKBP9', 'NT5C3A', 'RP9'] | Immune_age_gap_brain | PhenoBAG | 7p14.3 |
| rs7840344 | 8 | 10726464 | 2.52E-10 | ['MSRA', 'PRSS55', 'RP1L1', 'C8orf74', 'SOX7', 'SOX7', 'PINX1', 'XKR6', 'AF131215.5', 'MTMR9', 'SLC35G5', 'C8orf12', 'FAM167A', 'BLK', 'GATA4', 'C8orf49', 'NEIL2', 'FDFT1', 'RP11-297N6.4', 'CTSB', 'DEFB136', 'DEFB135', 'DEFB134', 'FAM86B1'] | Immune_age_gap_brain | PhenoBAG | 8p23.1 |
| rs35091702 | 8 | 30279470 | 6.48E-11 | ['RBPMS', 'GTF2E2'] | Immune_age_gap_brain | PhenoBAG | 8p12 |
| rs7461534 | 8 | 41628650 | 2.34E-18 | ['GINS4', 'ANK1'] | Immune_age_gap_brain | PhenoBAG | 8p11.21 |
| rs10974815 | 9 | 4854253 | 3.68E-20 | ['AK3', 'RCL1', 'AL158147.2'] | Immune_age_gap_brain | PhenoBAG | 9p24.1 |
| rs61469632 | 9 | 135861990 | 7.71E-10 | ['GFI1B', 'SURF4', 'C9orf96', 'REXO4', 'ADAMTS13', 'CACFD1', 'SLC2A6'] | Immune_age_gap_brain | PhenoBAG | 9q34.13 |
| rs10793620 | 10 | 46087283 | 6.17E-11 | ['OR13A1', 'ALOX5', 'MARCH8', 'ZFAND4', 'FAM21C'] | Immune_age_gap_brain | PhenoBAG | 10q11.21 |
| rs17476364 | 10 | 71094504 | 1.63E-27 | ['HK1'] | Immune_age_gap_brain | PhenoBAG | 10q22.1 |
| rs2294081 | 11 | 193863 | 5.59E-10 | ['BET1L', 'SCGB1C1', 'ODF3', 'RIC8A', 'SIRT3', 'PSMD13', 'NLRP6'] | Immune_age_gap_brain | PhenoBAG | 11p15.5 |
| rs12419046 | 11 | 9822520 | 3.00E-09 | ['SWAP70', 'SBF2', 'ADM', 'AMPD3'] | Immune_age_gap_brain | PhenoBAG | 11p15.4 |
| rs201519335 | 11 | 16239763 | 7.16E-16 | ['SOX6', 'C11orf58', 'PLEKHA7'] | Immune_age_gap_brain | PhenoBAG | 11p15.1 |
| rs3842642 | 11 | 119145705 | 2.88E-09 | ['NLRX1', 'PDZD3', 'CCDC153', 'CBL', 'MCAM'] | Immune_age_gap_brain | PhenoBAG | 11q23.3 |
| rs35039023 | 12 | 6291665 | 2.19E-11 | ['CD9'] | Immune_age_gap_brain | PhenoBAG | 12p13.31 |
| rs7957424 | 12 | 120894318 | 4.16E-12 | ['COX6A1', 'AL021546.6', 'TRIAP1', 'GATC', 'SRSF9', 'DYNLL1', 'COQ5', 'RNF10', 'POP5', 'P2RX4'] | Immune_age_gap_brain | PhenoBAG | 12q24.31 |
| rs11615667 | 12 | 122399173 | 6.09E-36 | ['KDM2B', 'ORAI1', 'MORN3', 'TMEM120B', 'RHOF', 'HPD', 'PSMD9', 'RP11-87C12.2', 'WDR66'] | Immune_age_gap_brain | PhenoBAG | 12q24.31 |
| rs2713931 | 15 | 56550975 | 1.44E-09 | ['PIGB', 'RFX7', 'TEX9', 'MNS1'] | Immune_age_gap_brain | PhenoBAG | 15q21.3 |
| rs1550026 | 15 | 65705116 | 8.54E-11 | ['IGDCC4', 'DPP8', 'PTPLAD1', 'VWA9', 'SLC24A1', 'DENND4A', 'RAB11A', 'DIS3L'] | Immune_age_gap_brain | PhenoBAG | 15q22.31 |
| rs12594925 | 15 | 91520287 | 1.36E-11 | ['MAN2A2', 'UNC45A', 'RCCD1', 'PRC1', 'VPS33B'] | Immune_age_gap_brain | PhenoBAG | 15q26.1 |
| rs12931752 | 16 | 159710 | 2.19E-17 | ['SNRNP25', 'RHBDF1', 'MPG', 'NPRL3', 'HBZ', 'HBM', 'HBA2', 'HBA1', 'HBQ1', 'LUC7L', 'ITFG3', 'ITFG3', 'RGS11', 'ARHGDIG', 'PDIA2', 'AXIN1', 'TMEM8A'] | Immune_age_gap_brain | PhenoBAG | 16p13.3 |

| | | | | | | | |
|---|---|---|---|---|---|---|---|
| rs112180638 | 17 | 19923166 | 6.04E-10 | ['AKAP10', 'SPECC1', 'LGALS9B'] | Immune_age_gap_brain | PhenoBAG | 17p11.2 |
| rs536327 | 17 | 27857698 | 8.12E-11 | ['TAOK1', 'ABHD15', 'TP53I13'] | Immune_age_gap_brain | PhenoBAG | 17q11.2 |
| rs7503168 | 17 | 33885904 | 4.21E-15 | ['SLFN12L', 'SLFN14', 'PEX12', 'AP2B1', 'RASL10B', 'GAS2L2'] | Immune_age_gap_brain | PhenoBAG | 17q12 |
| rs78310935 | 17 | 55449072 | 1.94E-09 | ['MSI2'] | Immune_age_gap_brain | PhenoBAG | 17q22 |
| rs34381780 | 17 | 58005749 | 1.69E-09 | ['VMP1', 'TUBD1', 'RPS6KB1', 'RP11-178C3.1', 'RNFT1'] | Immune_age_gap_brain | PhenoBAG | 17q23.1 |
| rs11085076 | 19 | 4476270 | 5.33E-19 | ['HDGFRP2', 'PLIN4'] | Immune_age_gap_brain | PhenoBAG | 19p13.3 |
| rs17678527 | 19 | 11294120 | 1.59E-09 | ['KANK2', 'DOCK6'] | Immune_age_gap_brain | PhenoBAG | 19p13.2 |
| rs7412 | 19 | 45412079 | 3.85E-10 | ['PVRL2', 'TOMM40', 'APOE', 'APOC1'] | Immune_age_gap_brain | PhenoBAG | 19q13.32 |
| rs6014993 | 20 | 55991637 | 1.33E-13 | ['RBM38'] | Immune_age_gap_brain | PhenoBAG | 20q13.31 |
| rs415064 | 20 | 57597971 | 2.68E-84 | ['GNAS', 'NELFCD', 'CTSZ', 'TUBB1', 'ATP5E', 'SLMO2', 'ZNF831', 'EDN3'] | Immune_age_gap_brain | PhenoBAG | 20q13.32 |
| rs2072860 | 22 | 37470604 | 1.86E-11 | ['KCTD17', 'TMPRSS6'] | Immune_age_gap_brain | PhenoBAG | 22q12.3 |
| rs140522 | 22 | 50971266 | 3.12E-21 | ['MIOX', 'LMF2', 'NCAPH2', 'SCO2', 'TYMP', 'ODF3B', 'KLHDC7B', 'SYCE3', 'CPT1B'] | Immune_age_gap_brain | PhenoBAG | 22q13.33 |
| rs12045096 | 1 | 46114428 | 1.94E-10 | ['TESK2', 'AKR1A1', 'NASP', 'CCDC17', 'GPBP1L1', 'TMEM69', 'IPP', 'MAST2', 'PIK3R3'] | Metabolic_age_gap_brain | PhenoBAG | 1p34.1 |
| rs11591147 | 1 | 55505647 | 4.12E-17 | ['PCSK9', 'USP24'] | Metabolic_age_gap_brain | PhenoBAG | 1p32.3 |
| rs10789112 | 1 | 62957758 | 1.20E-26 | ['USP1', 'DOCK7', 'ANGPTL3', 'AL138847.1', 'ATG4C'] | Metabolic_age_gap_brain | PhenoBAG | 1p31.3 |
| rs12740374 | 1 | 109817590 | 1.67E-18 | ['SARS', 'CELSR2', 'PSRC1', 'AMIGO1'] | Metabolic_age_gap_brain | PhenoBAG | 1p13.3 |
| rs857721 | 1 | 158612548 | 3.91E-27 | ['OR10K2', 'OR10K1', 'OR10R2', 'OR6Y1', 'OR6P1', 'OR10X1', 'OR10Z1', 'SPTA1'] | Metabolic_age_gap_brain | PhenoBAG | 1q23.1 |
| rs79687284 | 1 | 214150821 | 1.07E-11 | ['PROX1'] | Metabolic_age_gap_brain | PhenoBAG | 1q32.3 |
| rs556107 | 1 | 234853059 | 4.49E-10 | ['IRF2BP2'] | Metabolic_age_gap_brain | PhenoBAG | 1q42.3 |
| rs907867 | 2 | 20371301 | 3.53E-10 | ['SDC1'] | Metabolic_age_gap_brain | PhenoBAG | 2p24.1 |
| rs34722314 | 2 | 21271707 | 3.54E-26 | ['APOB', 'TDRD15'] | Metabolic_age_gap_brain | PhenoBAG | 2p24.1 |
| rs1260326 | 2 | 27730940 | 4.30E-10 | ['TRIM54', 'EIF2B4', 'SNX17', 'ZNF513', 'PPM1G', 'KRTCAP3', 'GCKR', 'AC109829.1'] | Metabolic_age_gap_brain | PhenoBAG | 2p23.3 |
| rs112259853 | 2 | 169790258 | 7.03E-46 | ['NOSTRIN', 'SPC25', 'G6PC2', 'ABCB11'] | Metabolic_age_gap_brain | PhenoBAG | 2q31.1 |
| rs2885296 | 2 | 234659061 | 3.18E-16 | ['USP40', 'UGT1A8', 'UGT1A10', 'UGT1A9', 'UGT1A7', 'UGT1A6', 'UGT1A5', 'UGT1A4', 'UGT1A3', 'UGT1A1', 'MROH2A'] | Metabolic_age_gap_brain | PhenoBAG | 2q37.1 |
| rs35939242 | 3 | 12301893 | 1.41E-27 | ['TIMP4', 'PPARG', 'TSEN2', 'C3orf83', 'MKRN2', 'RAF1', 'TMEM40', 'CAND2'] | Metabolic_age_gap_brain | PhenoBAG | 3p25.2 |
| rs11708067 | 3 | 123065778 | 2.27E-18 | ['PDIA5', 'SEC22A', 'ADCY5'] | Metabolic_age_gap_brain | PhenoBAG | 3q21.1 |
| rs139750425 | 4 | 3470174 | 1.28E-09 | ['DOK7', 'LRPAP1'] | Metabolic_age_gap_brain | PhenoBAG | 4p16.3 |
| rs55935372 | 4 | 144621779 | 1.25E-09 | ['FREM3', 'GYPE', 'GYPB', 'GYPA'] | Metabolic_age_gap_brain | PhenoBAG | 4q31.21 |
| rs4704727 | 5 | 156380067 | 3.97E-13 | ['TIMD4', 'HAVCR1'] | Metabolic_age_gap_brain | PhenoBAG | 5q33.3 |
| rs7766070 | 6 | 20686573 | 4.11E-15 | ['CDKAL1'] | Metabolic_age_gap_brain | PhenoBAG | 6p22.3 |
| rs79220007 | 6 | 26098474 | 9.46E-22 | ['LRRC16A', 'SCGN', 'SLC17A1', 'SLC17A3', 'SLC17A2', 'TRIM38', 'HIST1H1A', 'HIST1H3A', 'HIST1H4A', 'HIST1H4B', 'HIST1H3B', 'HIST1H2AB', 'HIST1H2BB', 'HIST1H3C', 'HIST1H1C', 'HFE', 'HIST1H4C', 'HIST1H1T', 'HIST1H2BC', 'HIST1H2AC', 'BTN3A2', 'BTN2A2', 'BTN2A1', 'BTN1A1', 'ABT1', 'ZNF322', 'HIST1H2BK', | Metabolic_age_gap_brain | PhenoBAG | 6p22.2 |

| | | | | | | | |
|---|---|---|---|---|---|---|---|
| | | | | 'HIST1H2AH', 'PRSS16', 'HIST1H3H', 'HIST1H2AJ', 'HIST1H2BM', 'HIST1H4J', 'HIST1H4K', 'HIST1H2AK', 'HIST1H3J', 'HIST1H2AM', 'HIST1H2BO', 'ZNF165', 'ZSCAN16', 'ZKSCAN8', 'PGBD1', 'ZSCAN31', 'ZKSCAN3', 'ZSCAN12', 'ZSCAN23', 'GPX6', 'GPX5', 'SCAND3'] | | | |
| rs76967117 | 6 | 34603691 | 3.91E-09 | ['SPDEF', 'C6orf106', 'SNRPC', 'UHRF1BP1', 'ANKS1A', 'SCUBE3', 'ZNF76', 'DEF6', 'PPARD', 'FANCE', 'MAPK13'] | Metabolic_age_gap_brain | PhenoBAG | 6p21.31 |
| rs56293029 | 6 | 135419039 | 3.86E-13 | ['ALDH8A1', 'HBS1L'] | Metabolic_age_gap_brain | PhenoBAG | 6q23.3 |
| rs74617384 | 6 | 160997118 | 3.86E-26 | ['SLC22A2', 'SLC22A3', 'LPA', 'PLG'] | Metabolic_age_gap_brain | PhenoBAG | 6q25.3 |
| rs2971667 | 7 | 44245060 | 1.80E-43 | ['URGCP-MRPS24', 'MRPS24', 'URGCP', 'UBE2D4', 'DBNL', 'AEBP1', 'POLD2', 'GCK', 'YKT6', 'CAMK2B'] | Metabolic_age_gap_brain | PhenoBAG | 7p13 |
| rs4841132 | 8 | 9183596 | 6.23E-12 | | Metabolic_age_gap_brain | PhenoBAG | 8p23.1 |
| rs115849089 | 8 | 19912370 | 2.86E-12 | ['LPL'] | Metabolic_age_gap_brain | PhenoBAG | 8p21.3 |
| rs72638977 | 8 | 41540622 | 5.22E-18 | ['GINS4', 'ANK1'] | Metabolic_age_gap_brain | PhenoBAG | 8p11.21 |
| rs3802177 | 8 | 118185025 | 1.30E-20 | ['SLC30A8'] | Metabolic_age_gap_brain | PhenoBAG | 8q24.11 |
| rs1215112 | 9 | 15303583 | 4.56E-09 | ['TTC39B'] | Metabolic_age_gap_brain | PhenoBAG | 9p22.3 |
| rs10811660 | 9 | 22134068 | 6.17E-11 | | Metabolic_age_gap_brain | PhenoBAG | 9p21.3 |
| rs146783254 | 9 | 79946049 | 2.92E-09 | ['VPS13A', 'GNA14'] | Metabolic_age_gap_brain | PhenoBAG | 9q21.2 |
| rs142550358 | 9 | 91392686 | 7.77E-10 | | Metabolic_age_gap_brain | PhenoBAG | 9q22.1 |
| rs2740488 | 9 | 107661742 | 2.93E-31 | ['NIPSNAP3A', 'NIPSNAP3B', 'ABCA1'] | Metabolic_age_gap_brain | PhenoBAG | 9q31.1 |
| rs635634 | 9 | 136155000 | 1.33E-27 | ['SURF6', 'MED22', 'RPL7A', 'SURF1', 'SURF2', 'SURF4', 'C9orf96', 'REXO4', 'ADAMTS13', 'CACFD1', 'SLC2A6'] | Metabolic_age_gap_brain | PhenoBAG | 9q34.2 |
| rs17476364 | 10 | 71094504 | 2.68E-175 | ['SRGN', 'VPS26A', 'SUPV3L1', 'HKDC1', 'HK1', 'TACR2', 'TSPAN15'] | Metabolic_age_gap_brain | PhenoBAG | 10q22.1 |
| rs34872471 | 10 | 114754071 | 6.71E-28 | ['TCF7L2'] | Metabolic_age_gap_brain | PhenoBAG | 10q25.2 |
| rs415895 | 11 | 9769562 | 1.47E-10 | ['SWAP70', 'SBF2', 'ADM', 'AMPD3'] | Metabolic_age_gap_brain | PhenoBAG | 11p15.4 |
| rs138135047 | 11 | 47337383 | 8.33E-15 | ['PACSIN3', 'DDB2', 'ACP2', 'NR1H3', 'MADD', 'MYBPC3', 'SPI1', 'SLC39A13', 'PSMC3', 'RAPSN', 'CELF1', 'C1QTNF4', 'MTCH2', 'FNBP4', 'NUP160', 'PTPRJ'] | Metabolic_age_gap_brain | PhenoBAG | 11p11.2 |
| rs174567 | 11 | 61593005 | 4.16E-42 | ['MYRF', 'TMEM258', 'FEN1', 'FADS2', 'FADS1', 'FADS3', 'RAB3IL1'] | Metabolic_age_gap_brain | PhenoBAG | 11q12.2 |
| rs12721030 | 11 | 116705278 | 2.05E-30 | ['BUD13', 'ZNF259', 'APOA5', 'APOA4', 'APOC3', 'APOA1', 'SIK3', 'PAFAH1B2', 'SIDT2', 'TAGLN', 'PCSK7', 'RNF214', 'BACE1'] | Metabolic_age_gap_brain | PhenoBAG | 11q23.3 |
| rs7930518 | 11 | 122523596 | 4.85E-11 | ['UBASH3B'] | Metabolic_age_gap_brain | PhenoBAG | 11q24.1 |
| rs117233107 | 12 | 4328521 | 3.21E-14 | | Metabolic_age_gap_brain | PhenoBAG | 12p13.32 |
| ss1388090533 | 12 | 9104207 | 1.91E-09 | ['PHC1', 'M6PR', 'KLRG1'] | Metabolic_age_gap_brain | PhenoBAG | 12p13.31 |
| rs4760682 | 12 | 48512285 | 4.08E-20 | ['TMEM106C', 'COL2A1', 'RP1-228P16.5', 'SENP1', 'PFKM', 'ASB8', 'C12orf68', 'DKFZP779L1853', 'OR10AD1', 'H1FNT', 'ZNF641', 'AC024257.1'] | Metabolic_age_gap_brain | PhenoBAG | 12q13.11 |
| rs2701180 | 12 | 121451866 | 1.13E-10 | ['HNF1A', 'C12orf43', 'OASL'] | Metabolic_age_gap_brain | PhenoBAG | 12q24.31 |
| rs10773112 | 12 | 125338529 | 2.47E-09 | ['SCARB1'] | Metabolic_age_gap_brain | PhenoBAG | 12q24.31 |
| rs60353775 | 13 | 28498265 | 1.24E-10 | ['PDX1'] | Metabolic_age_gap_brain | PhenoBAG | 13q12.2 |
| rs7140110 | 13 | 114544024 | 8.17E-13 | ['GAS6'] | Metabolic_age_gap_brain | PhenoBAG | 13q34 |
| rs71884092 | 15 | 58679807 | 8.60E-96 | ['ALDH1A2', 'LIPC', 'ADAM10', | Metabolic_age_gap_brain | PhenoBAG | 15q21.3 |

| | | | | | | | |
|---|---|---|---|---|---|---|---|
| | | | | 'FAM63B', 'MYO1E'] | | | |
| rs17231506 | 16 | 56994528 | 5.64E-86 | ['NUP93', 'SLC12A3', 'HERPUD1', 'CETP', 'NLRC5', 'FAM192A', 'RSPRY1'] | Metabolic_age_gap_brain | PhenoBAG | 16q13 |
| rs837763 | 16 | 88853729 | 6.92E-16 | ['CTU2', 'PIEZO1', 'CDT1'] | Metabolic_age_gap_brain | PhenoBAG | 16q24.3 |
| rs112885647 | 17 | 7532813 | 9.08E-12 | ['BCL6B', 'CHRNB1', 'TNFSF12', 'TNFSF12-TNFSF13', 'TNFSF13', 'SENP3', 'CD68', 'MPDU1', 'SOX15', 'FXR2', 'AC007421.1', 'SHBG', 'SAT2', 'ATP1B2', 'TP53', 'WRAP53'] | Metabolic_age_gap_brain | PhenoBAG | 17p13.1 |
| rs74311766 | 17 | 45457136 | 4.36E-13 | ['ITGB3', 'ITGB3', 'EFCAB13', 'NPEPPS', 'KPNB1', 'TBKBP1'] | Metabolic_age_gap_brain | PhenoBAG | 17q21.32 |
| rs2748424 | 17 | 76124865 | 1.41E-16 | ['TNRC6C', 'TMC6', 'TMC8', 'SYNGR2', 'PGS1'] | Metabolic_age_gap_brain | PhenoBAG | 17q25.3 |
| rs3848403 | 17 | 80693899 | 8.26E-18 | ['FASN', 'FN3KRP', 'FN3K', 'TBCD', 'ZNF750', 'B3GNTL1'] | Metabolic_age_gap_brain | PhenoBAG | 17q25.3 |
| rs6507938 | 18 | 47168648 | 1.96E-26 | ['DYM', 'LIPG', 'ACAA2', 'RP11-886H22.1', 'MYO5B'] | Metabolic_age_gap_brain | PhenoBAG | 18q21.1 |
| rs6511720 | 19 | 11202306 | 6.77E-27 | ['SMARCA4', 'LDLR', 'SPC24', 'DOCK6', 'C19orf80'] | Metabolic_age_gap_brain | PhenoBAG | 19p13.2 |
| rs1065853 | 19 | 45413233 | 4.87E-162 | ['ZNF221', 'ZNF155', 'ZNF224', 'ZNF225', 'ZNF112', 'CTC-512J12.6', 'ZNF285', 'IGSF23', 'PVR', 'CEACAM19', 'CEACAM16', 'BCL3', 'CBLC', 'BCAM', 'PVRL2', 'TOMM40', 'APOE', 'APOC1', 'APOC4-APOC2', 'APOC4', 'APOC2', 'CTB-129P6.11', 'CLPTM1', 'RELB', 'CLASRP', 'MARK4', 'PPP1R37', 'NKPD1', 'TRAPPC6A', 'BLOC1S3', 'AC005779.2', 'AC006126.3', 'DMPK'] | Metabolic_age_gap_brain | PhenoBAG | 19q13.32 |
| rs67546213 | 19 | 50082587 | 3.89E-09 | ['FCGRT', 'NOSIP', 'PRRG2', 'PRR12', 'RRAS', 'SCAF1', 'IRF3', 'CPT1C'] | Metabolic_age_gap_brain | PhenoBAG | 19q13.33 |
| rs367070 | 19 | 54800500 | 7.84E-12 | ['LILRB2', 'LILRA3', 'LILRA5'] | Metabolic_age_gap_brain | PhenoBAG | 19q13.42 |
| rs1800961 | 20 | 43042364 | 1.46E-09 | ['HNF4A'] | Metabolic_age_gap_brain | PhenoBAG | 20q13.12 |
| rs2076085 | 22 | 37470041 | 1.98E-11 | ['KCTD17', 'TMPRSS6'] | Metabolic_age_gap_brain | PhenoBAG | 22q12.3 |
| rs149344982 | 1 | 21889760 | 2.71E-42 | ['EIF4G3', 'NBPF3', 'ALPL', 'RAP1GAP', 'USP48'] | Musculoskeletal_age_gap_brain | PhenoBAG | 1p36.12 |
| rs1991083 | 2 | 23887437 | 3.96E-10 | ['KLHL29', 'ATAD2B', 'MFSD2B', 'C2orf44', 'FAM228B'] | Musculoskeletal_age_gap_brain | PhenoBAG | 2p24.1 |
| rs1344840 | 2 | 25070645 | 2.25E-09 | ['NCOA1', 'PTRHD1', 'CENPO', 'ADCY3'] | Musculoskeletal_age_gap_brain | PhenoBAG | 2p23.3 |
| rs13426845 | 2 | 42222529 | 2.62E-13 | ['PKDCC'] | Musculoskeletal_age_gap_brain | PhenoBAG | 2p21 |
| rs17275273 | 3 | 64944311 | 8.65E-11 | | Musculoskeletal_age_gap_brain | PhenoBAG | 3p14.1 |
| rs4588 | 4 | 72618323 | 4.22E-21 | ['RUFY3', 'GRSF1', 'SLC4A4', 'GC', 'ADAMTS3'] | Musculoskeletal_age_gap_brain | PhenoBAG | 4q13.3 |
| rs2744575 | 6 | 24494975 | 8.18E-17 | ['MRS2', 'GPLD1', 'ALDH5A1'] | Musculoskeletal_age_gap_brain | PhenoBAG | 6p22.3 |
| rs36018387 | 6 | 35386872 | 2.98E-09 | ['SPDEF', 'C6orf106', 'SNRPC', 'UHRF1BP1', 'SCUBE3', 'PPARD', 'FANCE', 'MAPK13'] | Musculoskeletal_age_gap_brain | PhenoBAG | 6p21.31 |
| rs10628041 | 7 | 27613821 | 3.18E-19 | ['HIBADH', 'TAX1BP1', 'JAZF1'] | Musculoskeletal_age_gap_brain | PhenoBAG | 7p15.2 |
| rs34060476 | 7 | 73037956 | 3.02E-10 | ['POM121', 'FZD9', 'BAZ1B', 'BCL7B', 'TBL2', 'MLXIPL'] | Musculoskeletal_age_gap_brain | PhenoBAG | 7q11.23 |
| rs142005327 | 7 | 120969969 | 4.42E-11 | ['WNT16', 'FAM3C'] | Musculoskeletal_age_gap_brain | PhenoBAG | 7q31.31 |
| rs550057 | 9 | 136146597 | 2.80E-63 | ['RALGDS', 'GBGT1', 'OBP2B', 'SURF6', 'MED22', | Musculoskeletal_age_gap_brain | PhenoBAG | 9q34.2 |

| | | | | | | | |
|---|---|---|---|---|---|---|---|
| | | | | 'RPL7A', 'SURF1', 'SURF2', 'SURF4', 'C9orf96', 'REXO4', 'ADAMTS13', 'CACFD1', 'SLC2A6'] | | | |
| rs2393969 | 10 | 65140440 | 5.64E-17 | ['NRBF2', 'JMJD1C', 'REEP3'] | Musculoskeletal_age_gap_brain | PhenoBAG | 10q21.3 |
| rs116970203 | 11 | 14876718 | 1.77E-15 | ['COPB1', 'PSMA1', 'PDE3B', 'CYP2R1', 'CALCB', 'INSC'] | Musculoskeletal_age_gap_brain | PhenoBAG | 11p15.2 |
| rs61734601 | 11 | 67184725 | 2.16E-10 | ['RNASEH2C', 'SF3B2', 'MRPL11', 'PELI3', 'CTD-3074O7.11', 'BBS1', 'ZDHHC24', 'CCS', 'RBM14', 'RBM4', 'RBM14-RBM4', 'RBM4B', 'SPTBN2', 'C11orf80', 'PC', 'C11orf86', 'KDM2A', 'ADRBK1', 'ANKRD13D', 'SSH3', 'RAD9A', 'PPP1CA', 'TBC1D10C', 'CARNS1', 'NDUFV1'] | Musculoskeletal_age_gap_brain | PhenoBAG | 11q13.2 |
| rs2264782 | 12 | 121432603 | 1.62E-11 | ['POP5', 'MLEC', 'UNC119B', 'SPPL3', 'AC079602.1', 'HNF1A', 'C12orf43', 'OASL'] | Musculoskeletal_age_gap_brain | PhenoBAG | 12q24.31 |
| rs9533090 | 13 | 42951449 | 1.35E-09 | ['AKAP11'] | Musculoskeletal_age_gap_brain | PhenoBAG | 13q14.11 |
| rs12449427 | 17 | 7173279 | 1.32E-18 | ['ASGR1', 'DLG4', 'ACADVL', 'DVL2', 'PHF23', 'GABARAP', 'CTD-2545G14.7', 'CTDNEP1', 'RP1-4G17.5', 'ELP5', 'CLDN7', 'SLC2A4', 'YBX2'] | Musculoskeletal_age_gap_brain | PhenoBAG | 17p13.1 |
| rs35728097 | 19 | 49213853 | 7.20E-12 | ['CYTH2', 'FAM83E', 'NTN5', 'FUT2', 'MAMSTR', 'RASIP1', 'IZUMO1', 'FUT1', 'FGF21'] | Musculoskeletal_age_gap_brain | PhenoBAG | 19q13.33 |
| rs2425052 | 20 | 33880729 | 3.27E-19 | ['AHCY', 'ITCH', 'DYNLRB1', 'MAP1LC3A', 'PIGU', 'NCOA6', 'TP53INP2', 'GGT7', 'ACSS2', 'GSS', 'MYH7B', 'TRPC4AP', 'EDEM2', 'PROCR', 'MMP24', 'EIF6', 'FAM83C', 'UQCC1', 'GDF5OS', 'GDF5', 'ERGIC3', 'SPAG4', 'CPNE1'] | Musculoskeletal_age_gap_brain | PhenoBAG | 20q11.22 |
| rs6683792 | 1 | 26743903 | 9.92E-10 | ['EXTL1', 'CATSPER4', 'CEP85', 'SH3BGRL3', 'UBXN11', 'CD52', 'AIM1L', 'LIN28A', 'DHDDS', 'HMGN2', 'RPS6KA1'] | Pulmonary_age_gap_brain | PhenoBAG | 1p36.11 |
| rs28736888 | 1 | 218857609 | 5.09E-10 | | Pulmonary_age_gap_brain | PhenoBAG | 1q41 |
| rs62126405 | 2 | 18303119 | 5.59E-10 | ['KCNS3'] | Pulmonary_age_gap_brain | PhenoBAG | 2p24.2 |
| rs6734395 | 2 | 24054632 | 3.99E-09 | ['KLHL29', 'ATAD2B', 'UBXN2A', 'MFSD2B', 'C2orf44', 'FAM228B'] | Pulmonary_age_gap_brain | PhenoBAG | 2p23.3 |
| rs10084382 | 2 | 36768447 | 2.13E-10 | ['CRIM1', 'AC007401.2', 'FEZ2'] | Pulmonary_age_gap_brain | PhenoBAG | 2p22.2 |
| rs1465685 | 2 | 56186471 | 1.67E-11 | ['PNPT1', 'EFEMP1'] | Pulmonary_age_gap_brain | PhenoBAG | 2p16.1 |
| rs55681051 | 2 | 100620655 | 1.47E-09 | ['AFF3'] | Pulmonary_age_gap_brain | PhenoBAG | 2q11.2 |
| rs7424080 | 2 | 156997602 | 3.57E-11 | ['NR4A2'] | Pulmonary_age_gap_brain | PhenoBAG | 2q24.1 |
| rs2571445 | 2 | 218683154 | 2.41E-11 | ['DIRC3', 'TNS1'] | Pulmonary_age_gap_brain | PhenoBAG | 2q35 |
| rs11124193 | 2 | 239875322 | 6.89E-14 | | Pulmonary_age_gap_brain | PhenoBAG | 2q37.3 |
| rs784286 | 3 | 168970043 | 7.04E-11 | ['MECOM'] | Pulmonary_age_gap_brain | PhenoBAG | 3q26.2 |
| rs78101726 | 3 | 169295436 | 5.43E-11 | | Pulmonary_age_gap_brain | PhenoBAG | 3q26.2 |
| rs11938781 | 4 | 17924734 | 1.85E-12 | ['FAM184B', 'DCAF16', 'NCAPG', 'LCORL'] | Pulmonary_age_gap_brain | PhenoBAG | 4p15.31 |
| rs6830501 | 4 | 57808917 | 4.44E-09 | ['REST', 'NOA1', 'POLR2B', 'IGFBP7'] | Pulmonary_age_gap_brain | PhenoBAG | 4q12 |
| rs7664805 | 4 | 106843958 | 2.85E-21 | ['TET2', 'PPA2', 'ARHGEF38', 'INTS12', 'GSTCD', 'NPNT', 'TBCK', 'AIMP1'] | Pulmonary_age_gap_brain | PhenoBAG | 4q24 |
| rs62321845 | 4 | 124819607 | 9.18E-11 | | Pulmonary_age_gap_brain | PhenoBAG | 4q28.1 |
| rs12504628 | 4 | 145436324 | 4.16E-43 | ['HHIP', 'ANAPC10', 'ABCE1', 'OTUD4'] | Pulmonary_age_gap_brain | PhenoBAG | 4q31.21 |
| rs2407778 | 5 | 122700656 | 7.02E-10 | ['CEP120'] | Pulmonary_age_gap_brain | PhenoBAG | 5q23.2 |

| | | | | | | | |
|---|---|---|---|---|---|---|---|
| rs7733410 | 5 | 147856522 | 3.29E-21 | ['SPINK7', 'AC091948.1', 'SPINK9', 'FBXO38', 'HTR4'] | Pulmonary_age_gap_brain | PhenoBAG | 5q32 |
| rs13355228 | 5 | 156969683 | 2.82E-12 | ['ITK', 'CYFIP2', 'FNDC9', 'ADAM19', 'NIPAL4', 'SOX30', 'C5orf52'] | Pulmonary_age_gap_brain | PhenoBAG | 5q33.3 |
| rs411535 | 6 | 22061040 | 3.55E-09 | | Pulmonary_age_gap_brain | PhenoBAG | 6p22.3 |
| rs34613987 | 6 | 27433029 | 2.57E-11 | ['SCGN', 'SLC17A4', 'SLC17A1', 'SLC17A3', 'SLC17A2', 'TRIM38', 'HIST1H1A', 'HIST1H3A', 'HIST1H4A', 'HIST1H4B', 'HIST1H3B', 'HIST1H2AB', 'HIST1H2BB', 'HIST1H3C', 'HIST1H1C', 'HFE', 'HIST1H4C', 'HIST1H1T', 'HIST1H2BC', 'HIST1H2AC', 'HIST1H2BF', 'HIST1H4E', 'HIST1H2BG', 'HIST1H2AE', 'HIST1H3E', 'HIST1H1D', 'HIST1H2BH', 'HIST1H3G', 'HIST1H2BI', 'HIST1H4H', 'BTN3A2', 'BTN3A1', 'BTN3A3', 'BTN2A1', 'BTN1A1', 'HMGN4', 'ABT1', 'ZNF322', 'HIST1H2BJ', 'HIST1H2AG', 'HIST1H2BK', 'HIST1H4I', 'HIST1H2AH', 'PRSS16', 'POM121L2', 'ZNF391', 'ZNF184', 'HIST1H2BL', 'HIST1H2AI', 'HIST1H3H', 'HIST1H2AJ', 'HIST1H2BM', 'HIST1H4J', 'HIST1H4K', 'HIST1H2AK', 'HIST1H2BN', 'HIST1H2AL', 'HIST1H1B', 'HIST1H3I', 'HIST1H4L', 'HIST1H3J', 'HIST1H2AM', 'HIST1H2BO', 'OR2B2', 'OR2B6', 'ZNF165', 'ZSCAN16', 'ZKSCAN8', 'ZSCAN9', 'ZKSCAN4', 'NKAPL', 'PGBD1', 'ZSCAN31', 'ZKSCAN3', 'ZSCAN12', 'ZSCAN23', 'GPX6', 'GPX5', 'SCAND3', 'TRIM27', 'C6orf100', 'ZNF311', 'OR2W1', 'OR2B3', 'OR2J1', 'OR2J3', 'OR2J2', 'OR14J1'] | Pulmonary_age_gap_brain | PhenoBAG | 6p22.1 |
| rs75104038 | 6 | 34190104 | 1.79E-14 | ['GRM4', 'HMGA1', 'C6orf1', 'NUDT3', 'RPS10-NUDT3', 'RPS10', 'C6orf106', 'SNRPC', 'UHRF1BP1', 'TAF11', 'ANKS1A', 'TCP11', 'SCUBE3', 'ZNF76', 'DEF6', 'PPARD', 'FANCE', 'RPL10A', 'TEAD3', 'CLPS'] | Pulmonary_age_gap_brain | PhenoBAG | 6p21.31 |
| rs2798641 | 6 | 109268050 | 7.11E-12 | ['FOXO3', 'ARMC2'] | Pulmonary_age_gap_brain | PhenoBAG | 6q21 |
| rs12524502 | 6 | 109654221 | 2.17E-09 | ['CD164', 'PPIL6', 'SMPD2', 'MICAL1', 'ZBTB24', 'AK9', 'FIG4'] | Pulmonary_age_gap_brain | PhenoBAG | 6q21 |
| rs113898003 | 6 | 130341235 | 2.36E-11 | ['L3MBTL3'] | Pulmonary_age_gap_brain | PhenoBAG | 6q23.1 |
| rs56364507 | 6 | 140246095 | 2.09E-10 | | Pulmonary_age_gap_brain | PhenoBAG | 6q24.1 |
| rs4594972 | 6 | 142561244 | 4.55E-13 | ['NMBR', 'GJE1', 'VTA1', 'GPR126'] | Pulmonary_age_gap_brain | PhenoBAG | 6q24.1 |

| | | | | | | | |
|---|---|---|---|---|---|---|---|
| rs13271434 | 8 | 57158821 | 2.92E-09 | ['RPS20', 'PLAG1', 'CHCHD7', 'SDR16C5'] | Pulmonary_age_gap_brain | PhenoBAG | 8q12.1 |
| rs10116426 | 9 | 4145648 | 1.55E-10 | ['GLIS3'] | Pulmonary_age_gap_brain | PhenoBAG | 9p24.2 |
| rs10810579 | 9 | 16625061 | 4.69E-10 | ['BNC2'] | Pulmonary_age_gap_brain | PhenoBAG | 9p22.2 |
| rs2271804 | 10 | 12252217 | 2.87E-23 | ['SEC61A2', 'NUDT5', 'CDC123'] | Pulmonary_age_gap_brain | PhenoBAG | 10p13 |
| rs2509967 | 11 | 62312786 | 2.27E-09 | ['AHNAK', 'EEF1G', 'MIR3654', 'TUT1', 'MTA2', 'EML3', 'ROM1', 'B3GAT3', 'GANAB', 'INTS5', 'RP11-831H9.11', 'C11orf48', 'METTL12', 'C11orf83', 'TTC9C'] | Pulmonary_age_gap_brain | PhenoBAG | 11q12.3 |
| rs35099456 | 11 | 66649527 | 1.51E-09 | ['RNASEH2C', 'SF3B2', 'MRPL11', 'PELI3', 'CTD-3074O7.11', 'BBS1', 'ZDHHC24', 'CCS', 'RBM14', 'RBM4', 'RBM14-RBM4', 'RBM4B', 'SPTBN2', 'C11orf80', 'PC', 'C11orf86', 'KDM2A', 'ANKRD13D', 'SSH3', 'RAD9A', 'NDUFV1'] | Pulmonary_age_gap_brain | PhenoBAG | 11q13.2 |
| rs7968682 | 12 | 66371880 | 1.70E-17 | ['HMGA2', 'AC090673.2'] | Pulmonary_age_gap_brain | PhenoBAG | 12q14.3 |
| rs6538668 | 12 | 96128771 | 1.71E-09 | ['NTN4'] | Pulmonary_age_gap_brain | PhenoBAG | 12q22 |
| rs2812208 | 13 | 50707087 | 2.09E-12 | ['EBPL', 'KPNA3', 'TRIM13', 'DLEU1'] | Pulmonary_age_gap_brain | PhenoBAG | 13q14.2 |
| rs2588808 | 14 | 68660181 | 4.52E-09 | ['RAD51B', 'ZFP36L1'] | Pulmonary_age_gap_brain | PhenoBAG | 14q24.1 |
| rs72697299 | 14 | 93070367 | 1.72E-09 | ['RIN3'] | Pulmonary_age_gap_brain | PhenoBAG | 14q32.12 |
| rs2415116 | 15 | 71673185 | 2.91E-21 | ['THSD4'] | Pulmonary_age_gap_brain | PhenoBAG | 15q23 |
| rs7162542 | 15 | 84514290 | 2.05E-18 | ['HOMER2', 'C15orf40', 'SH3GL3', 'ADAMTSL3', 'GOLGA6L4'] | Pulmonary_age_gap_brain | PhenoBAG | 15q25.2 |
| rs72755233 | 15 | 100692953 | 4.34E-11 | ['ADAMTS17'] | Pulmonary_age_gap_brain | PhenoBAG | 15q26.3 |
| rs11645303 | 16 | 88814504 | 5.33E-10 | ['RNF166', 'CTU2', 'PIEZO1'] | Pulmonary_age_gap_brain | PhenoBAG | 16q24.3 |
| rs151088031 | 17 | 30405857 | 1.91E-09 | ['COPRS', 'UTP6', 'SUZ12', 'LRRC37B', 'AC090616.2', 'RHOT1', 'AC116407.2'] | Pulmonary_age_gap_brain | PhenoBAG | 17q11.2 |
| rs77804065 | 17 | 43810896 | 4.73E-16 | ['NMT1', 'FMNL1', 'ARHGAP27', 'PLEKHM1', 'CRHR1', 'SPPL2C', 'MAPT', 'STH', 'KANSL1', 'ARL17B', 'LRRC37A', 'LRRC37A2', 'ARL17A', 'NSF', 'WNT3', 'EFCAB13'] | Pulmonary_age_gap_brain | PhenoBAG | 17q21.31 |
| rs34500422 | 17 | 79906216 | 8.94E-11 | ['ARHGDIA', 'PYCR1', 'MYADML2', 'NOTUM', 'ASPSCR1', 'STRA13', 'LRRC45', 'RAC3', 'RFNG'] | Pulmonary_age_gap_brain | PhenoBAG | 17q25.3 |
| rs513953 | 18 | 8801351 | 7.59E-12 | ['SOGA2'] | Pulmonary_age_gap_brain | PhenoBAG | 18p11.22 |
| rs62621197 | 19 | 8670147 | 3.16E-09 | ['MYO1F', 'ADAMTS10'] | Pulmonary_age_gap_brain | PhenoBAG | 19p13.2 |
| rs7285566 | 22 | 18445288 | 1.86E-09 | ['MICAL3'] | Pulmonary_age_gap_brain | PhenoBAG | 22q11.21 |
| rs333947 | 1 | 110470764 | 6.46E-15 | ['CSF1'] | Renal_age_gap_brain | PhenoBAG | 1p13.3 |
| rs760077 | 1 | 155178782 | 1.38E-19 | ['PMVK', 'PYGO2', 'ZBTB7B', 'DCST2', 'DCST1', 'ADAM15', 'EFNA1', 'KRTCAP2', 'RP11-201K10.3', 'TRIM46', 'MUC1', 'THBS3', 'MTX1', 'GBA', 'SCAMP3', 'CLK2', 'HCN3', 'PKLR', 'FDPS', 'RUSC1', 'ASH1L', 'MSTO1', 'YY1AP1', 'DAP3', 'GON4L', 'SYT11', 'RIT1', 'KIAA0907', 'RXFP4', 'ARHGEF2', 'SSR2', 'UBQLN4', 'SEMA4A', 'SLC25A44', 'PMF1-BGLAP', 'PMF1', 'BGLAP', 'PAQR6', 'C1orf85', 'TSACC'] | Renal_age_gap_brain | PhenoBAG | 1q22 |
| rs66822347 | 2 | 15733023 | 3.19E-12 | ['DDX1'] | Renal_age_gap_brain | PhenoBAG | 2p24.3 |
| rs1260326 | 2 | 27730940 | 2.72E-13 | ['TRIM54', 'EIF2B4', 'SNX17', 'ZNF513', 'PPM1G', 'KRTCAP3', | Renal_age_gap_brain | PhenoBAG | 2p23.3 |

| | | | | | | | |
|---|---|---|---|---|---|---|---|
| | | | | 'IFT172', 'GCKR', 'AC109829.1'] | | | |
| rs190303734 | 2 | 203491226 | 1.33E-11 | ['FAM117B'] | Renal_age_gap_brain | PhenoBAG | 2q33.2 |
| rs1047891 | 2 | 211540507 | 1.14E-34 | ['CPS1'] | Renal_age_gap_brain | PhenoBAG | 2q34 |
| rs12694435 | 2 | 219279673 | 1.35E-10 | ['SLC11A1', 'CTDSP1', 'VIL1', 'USP37', 'RQCD1', 'PLCD4', 'ZNF142', 'BCS1L', 'RNF25', 'STK36', 'TTLL4', 'CYP27A1'] | Renal_age_gap_brain | PhenoBAG | 2q35 |
| rs62189028 | 2 | 226924209 | 3.48E-09 | | Renal_age_gap_brain | PhenoBAG | 2q36.3 |
| rs140414074 | 3 | 121486554 | 6.54E-10 | ['GOLGB1', 'IQCB1', 'EAF2', 'SLC15A2', 'ILDR1'] | Renal_age_gap_brain | PhenoBAG | 3q13.33 |
| rs10512987 | 3 | 135823077 | 3.20E-10 | ['PPP2R3A', 'MSL2', 'PCCB', 'STAG1', 'SLC35G2', 'NCK1', 'IL20RB'] | Renal_age_gap_brain | PhenoBAG | 3q22.3 |
| rs9820812 | 3 | 187736055 | 4.34E-10 | | Renal_age_gap_brain | PhenoBAG | 3q27.3 |
| rs1986734 | 4 | 77420784 | 1.07E-19 | ['FAM47E', 'FAM47E-STBD1', 'FAM47E-STBD1', 'CCDC158', 'SHROOM3'] | Renal_age_gap_brain | PhenoBAG | 4q21.1 |
| rs112647987 | 5 | 40635920 | 1.89E-12 | ['PTGER4', 'TTC33', 'PRKAA1', 'RPL37', 'CARD6', 'C7'] | Renal_age_gap_brain | PhenoBAG | 5p13.1 |
| rs10051765 | 5 | 176799992 | 9.37E-29 | ['FGFR4', 'NSD1', 'RAB24', 'MXD3', 'PRELID1', 'LMAN2', 'RGS14', 'SLC34A1', 'PFN3', 'F12', 'GRK6'] | Renal_age_gap_brain | PhenoBAG | 5q35.3 |
| rs12192672 | 6 | 7229619 | 2.70E-14 | ['RREB1', 'SSR1', 'CAGE1'] | Renal_age_gap_brain | PhenoBAG | 6p24.3 |
| rs55925606 | 6 | 25878848 | 3.26E-09 | ['LRRC16A', 'SCGN', 'SLC17A3', 'SLC17A2', 'HIST1H1A', 'HIST1H3A', 'HIST1H4A', 'HIST1H4B', 'HIST1H3B', 'HIST1H2AB', 'HFE', 'HIST1H4C', 'HIST1H1T', 'HIST1H2BC', 'HIST1H2AC'] | Renal_age_gap_brain | PhenoBAG | 6p22.2 |
| rs4714684 | 6 | 43396856 | 1.16E-11 | ['TTBK1', 'SLC22A7', 'CRIP3', 'ZNF318', 'ABCC10'] | Renal_age_gap_brain | PhenoBAG | 6p21.1 |
| rs9496567 | 6 | 100602753 | 3.70E-12 | | Renal_age_gap_brain | PhenoBAG | 6q16.3 |
| rs2004640 | 7 | 128578301 | 1.38E-20 | ['IRF5', 'TNPO3'] | Renal_age_gap_brain | PhenoBAG | 7q32.1 |
| rs55779150 | 7 | 151406449 | 7.56E-16 | ['PRKAG2'] | Renal_age_gap_brain | PhenoBAG | 7q36.1 |
| rs3758086 | 8 | 23714992 | 4.02E-13 | ['STC1'] | Renal_age_gap_brain | PhenoBAG | 8p21.2 |
| rs71461799 | 10 | 64914518 | 1.28E-11 | ['NRBF2', 'JMJD1C', 'REEP3'] | Renal_age_gap_brain | PhenoBAG | 10q21.3 |
| rs12247543 | 10 | 126419743 | 4.52E-10 | ['LHPP', 'RP11-12J10.3', 'FAM53B', 'METTL10', 'FAM175B'] | Renal_age_gap_brain | PhenoBAG | 10q26.13 |
| rs55733296 | 11 | 30754837 | 5.30E-15 | | Renal_age_gap_brain | PhenoBAG | 11p14.1 |
| rs7123220 | 11 | 122508163 | 2.40E-13 | ['UBASH3B'] | Renal_age_gap_brain | PhenoBAG | 11q24.1 |
| rs3809272 | 12 | 111800258 | 1.39E-20 | ['MYL2', 'CUX2', 'FAM109A', 'SH2B3', 'ATXN2', 'BRAP', 'ACAD10', 'RP11-162P23.2', 'ALDH2', 'MAPKAPK5', 'TMEM116', 'ERP29', 'NAA25', 'HECTD4', 'PTPN11', 'RPH3A'] | Renal_age_gap_brain | PhenoBAG | 12q24.12 |
| rs7310409 | 12 | 121424861 | 9.35E-21 | ['POP5', 'MLEC', 'UNC119B', 'ACADS', 'SPPL3', 'AC079602.1', 'HNF1A', 'C12orf43', 'OASL'] | Renal_age_gap_brain | PhenoBAG | 12q24.31 |
| rs60569686 | 13 | 49744296 | 5.34E-10 | ['FNDC3A', 'MLNR', 'CDADC1'] | Renal_age_gap_brain | PhenoBAG | 13q14.2 |
| rs141866277 | 15 | 43950699 | 9.63E-10 | ['LCMT2', 'ADAL', 'TUBGCP4', 'TP53BP1', 'MAP1A', 'PPIP5K1', 'STRC', 'CATSPER2', 'CKMT1A', 'FRMD5', 'CASC4', 'EIF3J', 'SPG11'] | Renal_age_gap_brain | PhenoBAG | 15q15.3 |
| rs11072567 | 15 | 76298744 | 3.69E-22 | ['MAN2C1', 'SNUPN', 'CSPG4', 'AC105020.1', 'ODF3L1', 'AC019294.1', 'UBE2Q2', 'FBXO22', 'NRG4', 'C15orf27'] | Renal_age_gap_brain | PhenoBAG | 15q24.2 |
| rs4966021 | 15 | 99285056 | 7.50E-17 | ['IGF1R'] | Renal_age_gap_brain | PhenoBAG | 15q26.3 |
| rs77924615 | 16 | 20392332 | 3.98E-23 | ['UMOD', 'PDILT'] | Renal_age_gap_brain | PhenoBAG | 16p12.3 |
| rs140851213 | 16 | 79754433 | 1.53E-11 | | Renal_age_gap_brain | PhenoBAG | 16q23.2 |
| rs754492 | 17 | 1639985 | 1.09E-09 | ['TLCD2', 'WDR81', 'SERPINF2'] | Renal_age_gap_brain | PhenoBAG | 17p13.3 |
| rs77542162 | 17 | 67081278 | 1.26E-09 | ['ABCA9', 'ABCA6', 'ABCA10', 'ABCA5'] | Renal_age_gap_brain | PhenoBAG | 17q24.2 |
| rs45512696 | 19 | 35550878 | 1.13E-10 | ['HPN', 'TMEM147'] | Renal_age_gap_brain | PhenoBAG | 19q13.12 |

| | | | | | | | |
|---|---|---|---|---|---|---|---|
| rs76782803 | 20 | 23665192 | 2.9e-322 | ['FOXA2', 'SSTR4', 'THBD', 'CD93', 'AL096677.1', 'NXT1', 'GZF1', 'NAPB', 'CSTL1', 'CST11', 'CST8', 'CST9L', 'CST9', 'CST3', 'CST4', 'CST1', 'CST2', 'CST5', 'GGTLC1', 'SYNDIG1', 'ENTPD6'] | Renal_age_gap_brain | PhenoBAG | 20p11.21 |
| rs209958 | 20 | 52717172 | 2.46E-10 | | Renal_age_gap_brain | PhenoBAG | 20q13.2 |
| rs8135578 | 22 | 38182546 | 3.99E-09 | ['NOL12', 'TRIOBP', 'H1F0', 'GCAT', 'GALR3', 'ANKRD54', 'EIF3L', 'MICALL1', 'PICK1'] | Renal_age_gap_brain | PhenoBAG | 22q13.1 |
| rs61804208 | 1 | 161661411 | 2.20E-09 | ['FCGR2B', 'FCRLA'] | Reproductive_female | ProtBAG | 1q23.3 |
| rs6796854 | 3 | 126244591 | 6.89E-10 | ['UROC1', 'CHST13', 'C3orf22'] | Reproductive_female | ProtBAG | 3q21.3 |
| rs4055121 | 11 | 126232337 | 1.33E-11 | ['DCPS', 'ST3GAL4'] | Reproductive_female | ProtBAG | 11q24.2 |
| rs8003309 | 14 | 103050143 | 1.19E-10 | ['ANKRD9', 'RCOR1'] | Reproductive_female | ProtBAG | 14q32.31 |
| rs4030933 | 19 | 43237764 | 1.15E-281 | ['ADCK4', 'CYP2A13', 'CYP2F1', 'CYP2S1', 'AXL', 'HNRNPUL1', 'TGFB1', 'CTC-435M10.3', 'TMEM91', 'B9D2', 'BCKDHA', 'EXOSC5', 'B3GNT8', 'ATP5SL', 'C19orf69', 'CEACAM21', 'CEACAM4', 'CEACAM7', 'CEACAM5', 'CEA', 'CEACAM6', 'CEACAM3', 'LYPD4', 'DMRTC2', 'RPS19', 'CD79A', 'ARHGEF1', 'RABAC1', 'ATP1A3', 'GRIK5', 'ZNF574', 'POU2F2', 'DEDD2', 'ZNF526', 'GSK3A', 'AC006486.9', 'AC006486.1', 'ERF', 'CIC', 'PAFAH1B3', 'PRR19', 'TMEM145', 'MEGF8', 'CNFN', 'LIPE', 'CXCL17', 'CEACAM1', 'CEACAM8', 'PSG3', 'PSG8', 'PSG1', 'PSG6', 'PSG11', 'PSG2', 'PSG5', 'PSG4', 'PSG9', 'TEX101', 'LYPD3', 'PHLDB3', 'ETHE1', 'ZNF575', 'XRCC1', 'L34079.2', 'PINLYP', 'IRGQ', 'ZNF576', 'SRRM5', 'ZNF428', 'CADM4', 'PLAUR', 'ZNF155'] | Reproductive_female | ProtBAG | 19q13.2 |
| rs61740142 | 20 | 2464112 | 4.76E-17 | ['TGM6', 'RP4-734P14.4', 'ZNF343'] | Reproductive_female | ProtBAG | 20p13 |
| rs831256 | 3 | 182858461 | 1.25E-17 | ['MCCC1', 'LAMP3'] | Pulmonary | ProtBAG | 3q27.1 |
| rs1515496 | 3 | 189508566 | 1.77E-20 | ['TP63'] | Pulmonary | ProtBAG | 3q28 |
| rs41341748 | 8 | 16012594 | 5.57E-122 | ['TUSC3', 'MSR1', 'FGF20', 'ZDHHC2', 'CNOT7', 'VPS37A'] | Pulmonary | ProtBAG | 8p22 |
| rs7869487 | 9 | 117580914 | 3.62E-09 | ['TNFSF15', 'TNFSF8'] | Pulmonary | ProtBAG | 9q32 |
| rs721917 | 10 | 81706324 | 1.82E-123 | ['AL133481.1', 'EIF5AL1', 'SFTPA2', 'SFTPA1', 'NUTM2B', 'NUTM2E', 'SFTPD', 'TMEM254', 'PLAC9', 'ANXA11', 'AL359195.1', 'MAT1A', 'DYDC1', 'DYDC2', 'FAM213A', 'TSPAN14'] | Pulmonary | ProtBAG | 10q22.3 |
| rs3741240 | 11 | 62186542 | 2.52E-44 | ['SCGB2A1', 'SCGB1D2', 'SCGB2A2', 'SCGB1D4', 'ASRGL1', 'SCGB1A1', 'AHNAK'] | Pulmonary | ProtBAG | 11q12.3 |

| | | | | | | | |
|---|---|---|---|---|---|---|---|
| rs4601794 | 11 | 126245145 | 9.51E-22 | ['PUS3', 'DDX25', 'RPUSD4', 'FAM118B', 'SRPR', 'FOXRED1', 'TIRAP', 'RP11-712L6.5', 'DCPS', 'ST3GAL4', 'KIRREL3'] | Pulmonary | ProtBAG | 11q24.2 |
| rs1755775 | 14 | 36691187 | 3.86E-10 | | Pulmonary | ProtBAG | 14q13.3 |
| rs55714927 | 17 | 7080316 | 3.07E-17 | ['ASGR1', 'DLG4', 'DVL2', 'PHF23', 'GABARAP', 'CTD-2545G14.7', 'CTDNEP1', 'RP1-4G17.5', 'ELP5', 'CLDN7'] | Pulmonary | ProtBAG | 17p13.1 |
| rs58818008 | 17 | 34377725 | 1.15E-55 | ['NLE1', 'CCL16', 'CCL14', 'CCL15-CCL14', 'CTB-186H2.3', 'CCL15', 'CCL23', 'CCL18', 'CCL3'] | Pulmonary | ProtBAG | 17q12 |
| rs77804065 | 17 | 43810896 | 1.22E-15 | ['NMT1', 'FMNL1', 'ARHGAP27', 'PLEKHM1', 'CRHR1', 'SPPL2C', 'MAPT', 'STH', 'KANSL1', 'ARL17B', 'LRRC37A', 'LRRC37A2', 'ARL17A', 'NSF', 'WNT3', 'EFCAB13'] | Pulmonary | ProtBAG | 17q21.31 |
| rs198379 | 1 | 11915467 | 2.52E-28 | ['AGTRAP', 'C1orf167', 'MTHFR', 'CLCN6', 'NPPA', 'NPPB', 'KIAA2013', 'PLOD1', 'MFN2', 'MIIP'] | Heart | ProtBAG | 1p36.22 |
| rs34008398 | 2 | 69093413 | 9.60E-25 | ['BMP10'] | Heart | ProtBAG | 2p13.3 |
| 9:136149709_AC_A | 9 | 136149709 | 1.47E-09 | ['SURF1'] | Heart | ProtBAG | 9q34.2 |
| rs1572995 | 1 | 205024012 | 3.63E-45 | ['ETNK2', 'PLEKHA6', 'NFASC', 'CNTN2', 'TMEM81', 'RBBP5', 'DSTYK', 'TMCC2', 'NUAK2', 'KLHDC8A'] | Brain | ProtBAG | 1q32.1 |
| rs2491393 | 1 | 207300259 | 3.85E-11 | ['C4BPA'] | Brain | ProtBAG | 1q32.2 |
| rs7597287 | 2 | 119903969 | 1.13E-09 | ['C1QL2', 'STEAP3'] | Brain | ProtBAG | 2q14.2 |
| rs146950657 | 6 | 29614405 | 4.16E-44 | ['ZNF165', 'ZSCAN16', 'ZSCAN31', 'ZKSCAN3', 'ZNF311', 'OR2J1', 'OR2J3', 'OR5V1', 'OR12D3', 'OR12D2', 'OR11A1', 'OR10C1', 'MAS1L', 'UBD', 'GABBR1'] | Brain | ProtBAG | 6p22.1 |
| rs4711510 | 6 | 37669641 | 9.39E-15 | ['CPNE5', 'RNF8', 'CMTR1', 'MDGA1', 'ZFAND3', 'BTBD9', 'GLO1'] | Brain | ProtBAG | 6p21.2 |
| rs772220206 | 7 | 121502883 | 2.73E-35 | ['PTPRZ1', 'AASS', 'FEZF1'] | Brain | ProtBAG | 7q31.32 |
| rs12924867 | 16 | 75207097 | 3.06E-10 | ['LDHD', 'ZFP1'] | Brain | ProtBAG | 16q23.1 |
| rs281440 | 19 | 10400304 | 5.14E-17 | ['ICAM1', 'ICAM4', 'ICAM5', 'ZGLP1', 'CTD-2369P2.10', 'FDX1L', 'TYK2'] | Brain | ProtBAG | 19p13.2 |
| rs150622725 | 20 | 3692345 | 1.35E-09 | ['SIGLEC1', 'HSPA12B'] | Brain | ProtBAG | 20p13 |
| rs117773903 | 22 | 39226589 | 4.14E-64 | ['SUN2', 'DNAL4', 'NPTXR', 'CBX6'] | Brain | ProtBAG | 22q13.1 |
| rs9939224 | 16 | 57002732 | 4.77E-10 | ['HERPUD1', 'CETP', 'FAM192A', 'RSPRY1'] | Eye | ProtBAG | 16q13 |
| rs72865429 | 18 | 599618 | 9.36E-302 | ['RP11-683L23.1', 'USP14', 'THOC1', 'COLEC12', 'CETN1', 'CLUL1', 'C18orf56', 'TYMS'] | Eye | ProtBAG | 18p11.32 |
| rs62108432 | 19 | 12964033 | 3.75E-27 | ['ZNF763', 'ZNF490', 'ZNF791', 'WDR83', 'WDR83OS', 'DHPS', 'FBXW9', 'ASNA1', 'BEST2', 'HOOK2', 'JUNB', 'PRDX2', 'RNASEH2A', 'RTBDN', 'MAST1', 'DNASE2', 'KLF1', 'GCDH', 'SYCE2', 'FARSA', 'CALR', 'RAD23A', 'GADD45GIP1', 'DAND5'] | Eye | ProtBAG | 19p13.2 |
| rs6065904 | 20 | 44534651 | 3.68E-12 | ['SNX21', 'CTSA', 'PLTP', 'PCIF1', 'ZNF335'] | Eye | ProtBAG | 20q13.12 |
| rs3832016 | 1 | 109818158 | 3.28E-13 | ['CELSR2', 'PSRC1', 'AMIGO1'] | Hepatic | ProtBAG | 1p13.3 |
| 2:17959473_GTA_G | 2 | 17959473 | 2.57E-22 | ['VSNL1', 'SMC6', 'GEN1', 'MSGN1'] | Hepatic | ProtBAG | 2p24.2 |
| rs1260326 | 2 | 27730940 | 8.78E-13 | ['TRIM54', 'EIF2B4', 'SNX17', | Hepatic | ProtBAG | 2p23.3 |

| | | | | | | | |
|---|---|---|---|---|---|---|---|
| | | | | 'ZNF513', 'PPM1G', 'KRTCAP3', 'GCKR', 'AC109829.1'] | | | |
| rs62280667 | 3 | 101084604 | 1.33E-12 | ['IMPG2', 'SENP7', 'PCNP'] | Hepatic | ProtBAG | 3q12.3 |
| rs5030103 | 3 | 186448928 | 2.68E-40 | ['TBCCD1', 'DNAJB11', 'AHSG', 'FETUB', 'HRG', 'KNG1'] | Hepatic | ProtBAG | 3q27.3 |
| rs2498323 | 4 | 3451109 | 6.03E-13 | ['RGS12', 'HGFAC', 'DOK7'] | Hepatic | ProtBAG | 4p16.3 |
| rs1352846 | 4 | 72617775 | 2.10E-26 | ['GC'] | Hepatic | ProtBAG | 4q13.3 |
| rs556235831 | 4 | 73991462 | 1.16E-10 | ['COX18', 'ANKRD17', 'ALB', 'AFP', 'AFM', 'RASSF6'] | Hepatic | ProtBAG | 4q13.3 |
| rs62384157 | 5 | 131707537 | 5.75E-13 | ['RAPGEF6', 'ACSL6', 'P4HA2', 'PDLIM4', 'SLC22A4', 'SLC22A5', 'C5orf56', 'IRF1', 'IL13', 'GDF9', 'UQCRQ', 'LEAP2'] | Hepatic | ProtBAG | 5q31.1 |
| rs2023696 | 5 | 135269687 | 4.23946e-311 | ['CAMLG', 'CXCL14', 'SLC25A48', 'IL9', 'LECT2', 'TGFBI', 'SMAD5', 'TRPC7', 'SPOCK1'] | Hepatic | ProtBAG | 5q31.1 |
| rs4252165 | 6 | 161161559 | 3.76E-18 | ['SLC22A3', 'LPA', 'PLG'] | Hepatic | ProtBAG | 6q26 |
| rs854562 | 7 | 94947969 | 1.47E-09 | ['PON1'] | Hepatic | ProtBAG | 7q21.3 |
| rs2148412 | 13 | 46595849 | 2.99E-12 | ['ZC3H13', 'CPB2'] | Hepatic | ProtBAG | 13q14.13 |
| rs8004382 | 14 | 21155393 | 3.34E-17 | ['TTC5', 'RNASE4', 'ANG', 'AL163636.6'] | Hepatic | ProtBAG | 14q11.2 |
| rs10147337 | 14 | 95025560 | 1.30E-19 | ['SERPINA1', 'SERPINA4', 'SERPINA5', 'RP11-986E7.7', 'SERPINA3'] | Hepatic | ProtBAG | 14q32.13 |
| rs147233090 | 15 | 44028047 | 4.11E-15 | ['STARD9', 'TTBK2', 'UBR1', 'TMEM62', 'TGM7', 'LCMT2', 'ADAL', 'ZSCAN29', 'TUBGCP4', 'TP53BP1', 'MAP1A', 'PPIP5K1', 'CKMT1B', 'STRC', 'CATSPER2', 'CKMT1A', 'PDIA3', 'ELL3', 'RP11-296A16.1', 'SERF2', 'AC018512.1', 'SERINC4', 'HYPK', 'MFAP1', 'WDR76', 'FRMD5', 'CASC4', 'EIF3J', 'SPG11'] | Hepatic | ProtBAG | 15q15.3 |
| rs7212936 | 17 | 1646651 | 1.14E-09 | ['TLCD2', 'WDR81', 'SERPINF2', 'SERPINF1'] | Hepatic | ProtBAG | 17p13.3 |
| rs10445391 | 17 | 34306106 | 2.50E-45 | ['LYZL6', 'CCL16', 'CCL14', 'CCL15-CCL14', 'CTB-186H2.3', 'CCL15'] | Hepatic | ProtBAG | 17q12 |
| rs58088385 | 20 | 33748229 | 4.77E-18 | ['E2F1', 'CHMP4B', 'RALY', 'EIF2S2', 'ASIP', 'AHCY', 'ITCH', 'PIGU', 'NCOA6', 'TP53INP2', 'GGT7', 'ACSS2', 'GSS', 'MYH7B', 'TRPC4AP', 'EDEM2', 'PROCR', 'MMP24', 'EIF6', 'FAM83C', 'UQCC1'] | Hepatic | ProtBAG | 20q11.22 |
| rs77623237 | 2 | 230114499 | 7.75E-17 | ['PID1'] | Renal | ProtBAG | 2q36.3 |
| rs34498262 | 5 | 115330494 | 1.34E-20 | ['ATG12', 'AQPEP', 'CTD-2287O16.3', 'COMMD10'] | Renal | ProtBAG | 5q23.1 |
| rs2038760 | 6 | 2680732 | 7.32E-10 | ['MYLK4'] | Renal | ProtBAG | 6p25.2 |
| rs34817919 | 6 | 44766127 | 6.99E-15 | ['SUPT3H', 'RUNX2'] | Renal | ProtBAG | 6p21.1 |
| rs3012471 | 6 | 133356235 | 4.56E-27 | ['EYA4'] | Renal | ProtBAG | 6q23.2 |
| rs78740585 | 7 | 150944302 | 1.05E-12 | ['ABCF2', 'CHPF2', 'SMARCD3'] | Renal | ProtBAG | 7q36.1 |
| rs1485307 | 8 | 120007395 | 1.65E-45 | ['TNFRSF11B', 'COLEC10', 'MAL2', 'NOV'] | Renal | ProtBAG | 8q24.12 |
| rs2241003 | 9 | 123666777 | 6.77E-10 | ['PSMD5', 'PHF19', 'TRAF1', 'C5'] | Renal | ProtBAG | 9q33.2 |
| rs6489548 | 12 | 588604 | 9.80E-152 | ['SLC6A13', 'KDM5A', 'CCDC77', 'B4GALNT3', 'NINJ2', 'RAD52'] | Renal | ProtBAG | 12p13.33 |
| rs61928096 | 12 | 53780633 | 2.45E-10 | ['SP7', 'SP1', 'AMHR2', 'PRR13', 'PCBP2', 'MAP3K12', 'TARBP2', 'RP11-793H13.10', 'NPFF', 'ATF7', 'ATP5G2'] | Renal | ProtBAG | 12q13.13 |
| rs9533090 | 13 | 42951449 | 2.73E-18 | ['AKAP11', 'TNFSF11'] | Renal | ProtBAG | 13q14.11 |
| rs9521792 | 13 | 111120089 | 3.98E-10 | ['COL4A2'] | Renal | ProtBAG | 13q34 |

| | | | | | | | |
|---|---|---|---|---|---|---|---|
| rs28929474 | 14 | 94844947 | 6.86E-14 | ['SERPINA1'] | Renal | ProtBAG | 14q32.13 |
| 15:51203676_ATT_A | 15 | 51203676 | 1.19E-14 | ['HDC', 'GABPB1', 'TRPM7', 'SPPL2A', 'AP4E1', 'TNFAIP8L3'] | Renal | ProtBAG | 15q21.2 |
| rs77924615 | 16 | 20392332 | 2.93E-13 | ['UMOD', 'PDILT'] | Renal | ProtBAG | 16p12.3 |
| rs6416905 | 17 | 41804464 | 4.20E-15 | ['SOST', 'DUSP3', 'C17orf105', 'MPP3', 'CD300LG'] | Renal | ProtBAG | 17q21.31 |
| rs2532269 | 17 | 44250108 | 5.41E-11 | ['NMT1', 'FMNL1', 'ARHGAP27', 'PLEKHM1', 'CRHR1', 'SPPL2C', 'MAPT', 'STH', 'KANSL1', 'ARL17B', 'LRRC37A', 'LRRC37A2', 'ARL17A', 'NSF', 'WNT3', 'EFCAB13'] | Renal | ProtBAG | 17q21.31 |
| rs884205 | 18 | 60054857 | 1.53E-13 | ['PIGN', 'TNFRSF11A'] | Renal | ProtBAG | 18q21.33 |
| rs17087194 | 4 | 57690811 | 6.81E-15 | ['SPINK2'] | Reproductive_male | ProtBAG | 4q12 |
| rs9472712 | 6 | 46134213 | 1e-323 | ['CLIC5', 'ENPP4', 'ENPP5', 'RCAN2', 'CYP39A1', 'SLC25A27', 'TDRD6', 'PLA2G7', 'ANKRD66'] | Reproductive_male | ProtBAG | 6p21.1 |
| rs61847060 | 10 | 51510203 | 9.92E-18 | ['AGAP7', 'MSMB'] | Reproductive_male | ProtBAG | 10q11.23 |
| rs7152772 | 14 | 21244502 | 4.28E-17 | ['EDDM3A', 'EDDM3B', 'RNASE6'] | Reproductive_male | ProtBAG | 14q11.2 |
| rs28929474 | 14 | 94844947 | 1.22E-10 | ['SERPINA1'] | Reproductive_male | ProtBAG | 14q32.13 |
| 17:44161091_GCTCCCCTGGTAAGTCCTAAA_G | 17 | 44161091 | 1.29E-36 | ['NMT1', 'FMNL1', 'ARHGAP27', 'PLEKHM1', 'CRHR1', 'SPPL2C', 'MAPT', 'STH', 'KANSL1', 'ARL17B', 'LRRC37A', 'LRRC37A2', 'ARL17A', 'NSF', 'WNT3', 'EFCAB13'] | Reproductive_male | ProtBAG | 17q21.31 |
| rs2288481 | 19 | 49878196 | 1.19826e-315 | ['BCAT2', 'GYS1', 'RUVBL2', 'CGB1', 'CGB5', 'CGB8', 'CGB7', 'NTF4', 'KCNA7', 'SNRNP70', 'LIN7B', 'C19orf73', 'PPFIA3', 'HRC', 'TRPM4', 'SLC6A16', 'CD37', 'TEAD2', 'DKKL1', 'CCDC155', 'PTH2', 'GFY', 'SLC17A7', 'PIH1D1', 'ALDH16A1', 'CTD-3148I10.9', 'FLT3LG', 'RPL13A', 'FCGRT', 'RCN3', 'NOSIP', 'PRRG2', 'PRR12', 'RRAS', 'SCAF1', 'IRF3', 'BCL2L12', 'PRMT1', 'ADM5', 'CPT1C', 'TSKS', 'NUP62', 'VRK3'] | Reproductive_male | ProtBAG | 19q13.33 |
| rs2979451 | 19 | 51411388 | 2.73E-26 | ['KLK15', 'KLK3', 'KLK2', 'AC037199.1', 'KLK4', 'KLK5'] | Reproductive_male | ProtBAG | 19q13.41 |
| rs951807 | 2 | 60513389 | 2.42E-13 | | Endocrine | ProtBAG | 2p16.1 |
| rs310752 | 3 | 12272683 | 3.24E-09 | | Endocrine | ProtBAG | 3p25.2 |
| rs73174306 | 3 | 169194244 | 8.54E-16 | ['MECOM'] | Endocrine | ProtBAG | 3q26.2 |
| rs2351528 | 6 | 74497009 | 2.05E-47 | ['CD109'] | Endocrine | ProtBAG | 6q13 |
| rs62472465 | 7 | 98281512 | 4.40E-14 | ['NPTX2', 'TMEM130'] | Endocrine | ProtBAG | 7q22.1 |
| rs13226219 | 7 | 130019491 | 1.18E-10 | ['CPA5', 'CPA1', 'CEP41'] | Endocrine | ProtBAG | 7q32.2 |
| rs11538929 | 8 | 120431506 | 5.32E-21 | ['MAL2', 'NOV', 'ENPP2', 'TAF2', 'DEPTOR'] | Endocrine | ProtBAG | 8q24.12 |
| rs16402 | 9 | 38406788 | 4.58E-10 | ['ALDH1B1', 'IGFBPL1'] | Endocrine | ProtBAG | 9p13.1 |
| rs2519093 | 9 | 136141870 | 9.63E-37 | ['OBP2B', 'MED22', 'RPL7A', 'SURF1', 'SURF2', 'SURF4', 'C9orf96', 'REXO4', 'ADAMTS13', 'CACFD1', 'SLC2A6', 'FAM163B', 'DBH'] | Endocrine | ProtBAG | 9q34.2 |
| rs7910135 | 10 | 118398046 | 1.21E-41 | ['PNLIPRP1', 'C10orf82'] | Endocrine | ProtBAG | 10q25.3 |
| rs10489671 | 1 | 157538725 | 1.61E-42 | ['FCRL5', 'FCRL4', 'FCRL3', 'FCRL2', 'FCRL1', 'CD5L'] | Immune | ProtBAG | 1q23.1 |
| rs12059417 | 1 | 207654301 | 3.14E-10 | ['C1orf116', 'YOD1', 'PFKFB2', 'C4BPA', 'CD55', 'CR2', 'CR1'] | Immune | ProtBAG | 1q32.2 |
| rs60264981 | 2 | 160677998 | 1.34E-11 | ['LY75', 'LY75-CD302', 'PLA2R1'] | Immune | ProtBAG | 2q24.2 |
| rs72928971 | 2 | 203591967 | 1.15E-11 | ['FAM117B'] | Immune | ProtBAG | 2q33.2 |
| rs2228467 | 3 | 42906116 | 9.29E-10 | ['HIGD1A', 'ACKR2', 'KRBOX1', | Immune | ProtBAG | 3p22.1 |

| | | | | | | | |
|---|---|---|---|---|---|---|---|
| | | | | 'KRBOX1', 'CYP8B1', 'POMGNT2'] | | | |
| rs71657399 | 4 | 76986705 | 1.07E-12 | ['PPEF2', 'NAAA', 'SDAD1', 'AC110615.1', 'CXCL9', 'ART3', 'CXCL10', 'CXCL11', 'NUP54'] | Immune | ProtBAG | 4q21.1 |
| rs2893910 | 6 | 27283254 | 1.35E-09 | ['BTN3A2', 'PRSS16', 'POM121L2'] | Immune | ProtBAG | 6p22.1 |
| rs2893910 | 6 | 27283254 | 1.35E-09 | ['ZNF391', 'ZKSCAN8'] | Immune | ProtBAG | 6p22.1 |
| rs1535039 | 6 | 29411432 | 6.33E-10 | ['ZSCAN16', 'ZSCAN9', 'PGBD1', 'ZSCAN31', 'ZKSCAN3', 'ZSCAN12', 'ZSCAN23', 'GPX6', 'SCAND3', 'TRIM27', 'C6orf100', 'ZNF311', 'OR2W1', 'OR2B3', 'OR2J1', 'OR2J3', 'OR2J2', 'OR14J1', 'OR5V1', 'OR12D3', 'OR12D2', 'OR11A1', 'OR10C1', 'OR2H1', 'MAS1L', 'UBD', 'GABBR1', 'OR2H2'] | Immune | ProtBAG | 6p22.1 |
| rs2608639 | 6 | 52632213 | 8.32E-17 | ['GSTA2', 'GSTA1', 'GSTA5'] | Immune | ProtBAG | 6p12.2 |
| rs1285968 | 7 | 141647022 | 3.79E-19 | ['AGK', 'KIAA1147', 'WEE2', 'SSBP1', 'TAS2R3', 'TAS2R4', 'TAS2R5', 'PRSS37', 'MGAM', 'OR9A4', 'CLEC5A', 'TAS2R38'] | Immune | ProtBAG | 7q34 |
| rs4743059 | 9 | 99899313 | 1.90E-31 | ['ZNF782', 'CTSV', 'CCDC180'] | Immune | ProtBAG | 9q22.33 |
| rs25680 | 12 | 6554628 | 4.38E-17 | ['CD27', 'TAPBPL', 'VAMP1'] | Immune | ProtBAG | 12p13.31 |
| rs1182933 | 12 | 121454622 | 2.50E-19 | ['GATC', 'POP5', 'MLEC', 'UNC119B', 'SPPL3', 'AC079602.1', 'HNF1A', 'C12orf43', 'OASL'] | Immune | ProtBAG | 12q24.31 |
| rs2038499 | 14 | 94949291 | 1.08E-19 | ['SERPINA11', 'SERPINA9', 'SERPINA12'] | Immune | ProtBAG | 14q32.13 |
| rs12599288 | 16 | 11822066 | 1.39E-13 | ['SNN', 'TXNDC11', 'ZC3H7A'] | Immune | ProtBAG | 16p13.13 |
| rs77924615 | 16 | 20392332 | 2.99E-09 | ['PDILT'] | Immune | ProtBAG | 16p12.3 |
| rs74562379 | 1 | 8269962 | 1.28E-26 | | Skin | ProtBAG | 1p36.23 |
| rs552693039 | 1 | 109816863 | 8.01E-10 | ['SARS', 'CELSR2', 'PSRC1'] | Skin | ProtBAG | 1p13.3 |
| rs11803731 | 1 | 152083325 | 6.48E-32 | ['SELENBP1', 'RIIAD1', 'TDRKH', 'LINGO4', 'RORC', 'THEM4', 'S100A10', 'S100A11', 'TCHHL1', 'TCHH', 'RPTN', 'HRNR', 'FLG', 'FLG2', 'LCE5A', 'CRCT1', 'LCE3E', 'LCE3A', 'KPRP', 'LCE1E', 'LCE1D', 'LCE1C', 'LCE1B', 'LCE1A', 'LCE6A', 'SMCP', 'IVL', 'SPRR2B', 'SPRR2F'] | Skin | ProtBAG | 1q21.3 |
| rs6674686 | 1 | 218859416 | 1.25E-09 | | Skin | ProtBAG | 1q41 |
| rs1260326 | 2 | 27730940 | 9.72E-17 | ['TRIM54', 'EIF2B4', 'SNX17', 'ZNF513', 'PPM1G', 'KRTCAP3', 'GCKR', 'AC109829.1', 'C2orf16', 'ZNF512', 'CCDC121', 'GPN1', 'SUPT7L', 'SLC4A1AP', 'AC074091.13'] | Skin | ProtBAG | 2p23.3 |
| rs17006436 | 2 | 71060111 | 1.02E-12 | ['CLEC4F', 'CD207'] | Skin | ProtBAG | 2p13.3 |
| rs833270 | 3 | 181520166 | 2.38E-12 | | Skin | ProtBAG | 3q26.33 |
| rs155942 | 5 | 139532538 | 4.29E-14 | ['CYSTM1', 'PFDN1', 'TMCO6', 'PCDHA1', 'PCDHA3', 'PCDHA9', 'PCDHA13'] | Skin | ProtBAG | 5q31.3 |
| rs3095268 | 6 | 29606761 | 7.94E-18 | ['SCGN', 'SLC17A4', 'SLC17A1', 'SLC17A3', 'SLC17A2', 'TRIM38', 'HIST1H1A', 'HIST1H3A', 'HIST1H4A', 'HIST1H4B', | Skin | ProtBAG | 6p22.1 |

| | | | | | | | |
|---|---|---|---|---|---|---|---|
| | | | | 'HIST1H3B', 'HIST1H2AB', 'HIST1H2BB', 'HIST1H3C', 'HIST1H1C', 'HFE', 'HIST1H4C', 'HIST1H1T', 'HIST1H2BC', 'HIST1H2AC', 'HIST1H2BD', 'HIST1H2BE', 'HIST1H4D', 'HIST1H3D', 'HIST1H2AD', 'HIST1H2BF', 'HIST1H4E', 'HIST1H2BG', 'HIST1H2AE', 'HIST1H3E', 'HIST1H1D', 'HIST1H3G', 'HIST1H2BI', 'HIST1H4H', 'BTN3A2', 'BTN2A2', 'BTN3A1', 'BTN3A3', 'BTN2A1', 'BTN1A1', 'HMGN4', 'ABT1', 'ZNF322', 'HIST1H2BJ', 'HIST1H2AG', 'HIST1H2BK', 'HIST1H4I', 'HIST1H2AH', 'PRSS16', 'POM121L2', 'ZNF391', 'ZNF184', 'HIST1H2BL', 'HIST1H2AI', 'HIST1H3H', 'HIST1H2AJ', 'HIST1H2BM', 'HIST1H4J', 'HIST1H4K', 'HIST1H2AK', 'HIST1H2BN', 'HIST1H2AL', 'HIST1H1B', 'HIST1H3I', 'HIST1H4L', 'HIST1H3J', 'HIST1H2AM', 'HIST1H2BO', 'OR2B2', 'OR2B6', 'ZNF165', 'ZSCAN16', 'ZKSCAN8', 'ZSCAN9', 'ZKSCAN4', 'NKAPL', 'PGBD1', 'ZSCAN31', 'ZKSCAN3', 'ZSCAN12', 'ZSCAN23', 'GPX6', 'SCAND3', 'TRIM27', 'C6orf100', 'ZNF311', 'OR2W1', 'OR2B3', 'OR2J1', 'OR2J3', 'OR2J2', 'OR14J1', 'OR5V1', 'OR12D3', 'OR12D2', 'OR11A1', 'OR10C1', 'OR2H1', 'MAS1L', 'UBD', 'GABBR1', 'OR2H2'] | | | |
| rs17145750 | 7 | 73026378 | 4.05E-11 | ['POM121', 'FZD9', 'BAZ1B', 'BCL7B', 'TBL2', 'MLXIPL'] | Skin | ProtBAG | 7q11.23 |
| rs2954024 | 8 | 126482930 | 5.25E-13 | | Skin | ProtBAG | 8q24.13 |
| rs2070074 | 9 | 34649442 | 2.23E-76 | ['PRSS3', 'UBE2R2', 'UBAP2', 'DCAF12', 'UBAP1', 'KIF24', 'NUDT2', 'C9orf24', 'FAM219A', 'DNAI1', 'ENHO', 'CNTFR', 'RPP25L', 'DCTN3', 'ARID3C', 'SIGMAR1', 'GALT', 'GALT', 'IL11RA', 'CCL27', 'RP11-195F19.5', 'CCL19', 'CCL21', 'FAM205A'] | Skin | ProtBAG | 9p13.3 |
| rs11002517 | 10 | 54315112 | 7.54E-12 | | Skin | ProtBAG | 10q21.1 |
| rs12577165 | 11 | 65776429 | 1.77E-52 | ['PPP2R5B', 'ZFPL1', 'TM7SF2', 'ZNHIT2', 'FAU', 'SYVN1', 'MRPL49', 'SLC25A45', 'FRMD8', 'MAP3K11', 'RELA', 'KAT5', 'AP5B1', 'OVOL1', 'CFL1', 'SNX32', 'MUS81', 'EFEMP2', 'CTSW', 'FIBP', 'CCDC85B', | Skin | ProtBAG | 11q13.1 |

| | | | | | | | |
|---|---|---|---|---|---|---|---|
| | | | | 'FOSL1', 'C11orf68', 'DRAP1', 'SART1', 'EIF1AD', 'BANF1', 'CST6', 'CATSPER1', 'GAL3ST3', 'SF3B2', 'RP11-1167A19.2', 'PACS1', 'KLC2', 'RAB1B', 'CNIH2', 'YIF1A', 'TMEM151A', 'RIN1', 'NPAS4', 'CCDC87', 'CCS', 'RBM14', 'RBM4', 'RBM14-RBM4', 'SPTBN2', 'C11orf80', 'PC'] | | | |
| rs964184 | 11 | 116648917 | 5.64E-14 | ['BUD13', 'ZNF259', 'APOA5', 'APOA4', 'APOC3', 'APOA1', 'SIK3', 'PAFAH1B2', 'SIDT2', 'TAGLN', 'PCSK7'] | Skin | ProtBAG | 11q23.3 |
| rs3859191 | 17 | 38128714 | 1.21E-23 | ['ERBB2', 'MIEN1', 'GRB7', 'IKZF3', 'ZPBP2', 'GSDMB', 'ORMDL3', 'LRRC3C', 'GSDMA', 'PSMD3', 'CSF3', 'MED24', 'THRA', 'NR1D1', 'MSL1', 'RAPGEFL1', 'CDC6', 'RARA'] | Skin | ProtBAG | 17q21.1 |
| rs415430 | 17 | 44859144 | 2.52E-13 | ['NMT1', 'FMNL1', 'ARHGAP27', 'PLEKHM1', 'CRHR1', 'SPPL2C', 'MAPT', 'STH', 'KANSL1', 'ARL17B', 'LRRC37A', 'LRRC37A2', 'ARL17A', 'NSF', 'WNT3', 'EFCAB13'] | Skin | ProtBAG | 17q21.31 |
| rs77542162 | 17 | 67081278 | 1.24E-20 | ['ABCA9', 'ABCA6', 'ABCA10', 'ABCA5', 'MAP2K6'] | Skin | ProtBAG | 17q24.2 |
| rs71354994 | 19 | 38789249 | 1.64E-14 | ['SIPA1L3', 'SPINT2', 'PPP1R14A', 'CTB-102L5.4', 'C19orf33', 'YIF1B', 'KCNK6', 'CATSPERG', 'PSMD8', 'FAM98C', 'RINL'] | Skin | ProtBAG | 19q13.2 |
| rs190712692 | 19 | 45425178 | 2.52E-17 | ['PVRL2', 'TOMM40', 'APOE', 'APOC1', 'APOC4-APOC2', 'APOC4'] | Skin | ProtBAG | 19q13.32 |
| rs9304708 | 19 | 51583936 | 9.00E-10 | ['KLK14'] | Skin | ProtBAG | 19q13.41 |
| rs6104027 | 20 | 43760461 | 7.93E-19 | ['KCNS1', 'WFDC5', 'WFDC12', 'PI3', 'SEMG1', 'SEMG2', 'SLPI', 'MATN4', 'RBPJL', 'SDC4', 'WFDC3'] | Skin | ProtBAG | 20q13.12 |
| rs135542 | 22 | 46556037 | 4.34E-25 | ['FLJ27365', 'PPARA'] | Skin | ProtBAG | 22q13.31 |

**eTable 5a: Phenotypic and genetic correlations between the 11 ProtBAGs and 9 PhenoBAGs**

a) **Phenotypic correlation**

| PhenoBAG | ProtBAG | Pearson_r | P-value | N |
|---|---|---|---|---|
| Brain_age_gap | Brain | 0.08061404 | 1.76E-07 | 4187 |
| Cardiovascular_age_gap | Brain | 0.0027792 | 0.75413314 | 12702 |
| Eye_age_gap | Brain | 0.00916593 | 0.57222512 | 3799 |
| Hepatic_age_gap | Brain | 0.12863793 | 5.27E-48 | 12702 |
| Immune_age_gap | Brain | 0.06888649 | 7.71E-15 | 12702 |
| Metabolic_age_gap | Brain | 0.08538404 | 5.43E-22 | 12702 |
| Musculoskeletal_age_gap | Brain | 0.09556607 | 3.66E-27 | 12702 |
| Pulmonary_age_gap | Brain | 0.07397483 | 6.95E-17 | 12702 |
| Renal_age_gap | Brain | 0.22901374 | 8.34E-151 | 12702 |
| Brain_age_gap | Endocrine | 0.01366985 | 0.37652745 | 4187 |
| Cardiovascular_age_gap | Endocrine | 0.00286918 | 0.74644168 | 12702 |
| Eye_age_gap | Endocrine | 0.02053382 | 0.20574954 | 3799 |
| Hepatic_age_gap | Endocrine | 0.12844301 | 7.30E-48 | 12702 |
| Immune_age_gap | Endocrine | 0.04157226 | 2.77E-06 | 12702 |
| Metabolic_age_gap | Endocrine | 0.10542588 | 1.00E-32 | 12702 |
| Musculoskeletal_age_gap | Endocrine | 0.1242874 | 6.62E-45 | 12702 |
| Pulmonary_age_gap | Endocrine | 0.05760039 | 8.22E-11 | 12702 |
| Renal_age_gap | Endocrine | 0.3051886 | 5.56E-272 | 12702 |
| Brain_age_gap | Eye | -0.0467586 | 0.00247518 | 4187 |
| Cardiovascular_age_gap | Eye | -0.0288994 | 0.00112433 | 12702 |
| Eye_age_gap | Eye | -0.0168589 | 0.29887484 | 3799 |
| Hepatic_age_gap | Eye | 0.03535288 | 6.74E-05 | 12702 |
| Immune_age_gap | Eye | -0.0017433 | 0.84425638 | 12702 |
| Metabolic_age_gap | Eye | 0.02613133 | 0.00322645 | 12702 |
| Musculoskeletal_age_gap | Eye | 0.04183178 | 2.40E-06 | 12702 |
| Pulmonary_age_gap | Eye | -0.0040265 | 0.65000541 | 12702 |
| Renal_age_gap | Eye | 0.11409485 | 4.52E-38 | 12702 |
| Brain_age_gap | Heart | 0.05878505 | 0.00014125 | 4187 |
| Cardiovascular_age_gap | Heart | 0.03541162 | 6.56E-05 | 12702 |
| Eye_age_gap | Heart | 0.03906549 | 0.01604157 | 3799 |
| Hepatic_age_gap | Heart | 0.08614672 | 2.33E-22 | 12702 |
| Immune_age_gap | Heart | 0.04685075 | 1.27E-07 | 12702 |
| Metabolic_age_gap | Heart | 0.00431327 | 0.62691525 | 12702 |
| Musculoskeletal_age_gap | Heart | 0.02644898 | 0.00287204 | 12702 |
| Pulmonary_age_gap | Heart | 0.08217612 | 1.75E-20 | 12702 |
| Renal_age_gap | Heart | 0.23427793 | 6.37E-158 | 12702 |
| Brain_age_gap | Hepatic | 0.01584437 | 0.30536301 | 4187 |
| Cardiovascular_age_gap | Hepatic | -0.0105758 | 0.23332539 | 12702 |
| Eye_age_gap | Hepatic | -0.0079598 | 0.62380986 | 3799 |
| Hepatic_age_gap | Hepatic | 0.09171062 | 3.89E-25 | 12702 |
| Immune_age_gap | Hepatic | 0.03140034 | 0.00040097 | 12702 |
| Metabolic_age_gap | Hepatic | 0.11905653 | 2.53E-41 | 12702 |
| Musculoskeletal_age_gap | Hepatic | 0.08282023 | 8.83E-21 | 12702 |
| Pulmonary_age_gap | Hepatic | 0.05088464 | 9.58E-09 | 12702 |
| Renal_age_gap | Hepatic | 0.25900302 | 8.72E-194 | 12702 |
| Brain_age_gap | Immune | 0.03818201 | 0.01348083 | 4187 |
| Cardiovascular_age_gap | Immune | -0.0141768 | 0.11011186 | 12702 |
| Eye_age_gap | Immune | 0.03291056 | 0.04252372 | 3799 |
| Hepatic_age_gap | Immune | 0.14191881 | 3.85E-58 | 12702 |
| Immune_age_gap | Immune | 0.11326109 | 1.54E-37 | 12702 |
| Metabolic_age_gap | Immune | 0.0039542 | 0.65588032 | 12702 |

| | | | | |
|---|---|---|---|---|
| Musculoskeletal_age_gap | Immune | 0.12447114 | 4.92E-45 | 12702 |
| Pulmonary_age_gap | Immune | 0.07715884 | 3.09E-18 | 12702 |
| Renal_age_gap | Immune | 0.33391125 | 0 | 12702 |
| Brain_age_gap | Pulmonary | 0.02796114 | 0.07043612 | 4187 |
| Cardiovascular_age_gap | Pulmonary | 0.0157238 | 0.07638501 | 12702 |
| Eye_age_gap | Pulmonary | 0.03752031 | 0.02074156 | 3799 |
| Hepatic_age_gap | Pulmonary | 0.13616874 | 1.26E-53 | 12702 |
| Immune_age_gap | Pulmonary | 0.08062508 | 8.98E-20 | 12702 |
| Metabolic_age_gap | Pulmonary | 0.07204835 | 4.30E-16 | 12702 |
| Musculoskeletal_age_gap | Pulmonary | 0.11851316 | 5.83E-41 | 12702 |
| Pulmonary_age_gap | Pulmonary | 0.06397525 | 5.32E-13 | 12702 |
| Renal_age_gap | Pulmonary | 0.27503738 | 3.05E-219 | 12702 |
| Brain_age_gap | Renal | -0.001254 | 0.93534661 | 4187 |
| Cardiovascular_age_gap | Renal | -0.0152671 | 0.08532662 | 12702 |
| Eye_age_gap | Renal | 0.00472951 | 0.77073457 | 3799 |
| Hepatic_age_gap | Renal | 0.0299143 | 0.00074663 | 12702 |
| Immune_age_gap | Renal | 0.01856608 | 0.03640002 | 12702 |
| Metabolic_age_gap | Renal | 0.11344822 | 1.17E-37 | 12702 |
| Musculoskeletal_age_gap | Renal | 0.0105282 | 0.23543385 | 12702 |
| Pulmonary_age_gap | Renal | 0.01570837 | 0.07667429 | 12702 |
| Renal_age_gap | Renal | 0.13234242 | 9.94E-51 | 12702 |
| Brain_age_gap | Reproductive_female | 0.0433506 | 0.00502277 | 4187 |
| Cardiovascular_age_gap | Reproductive_female | -0.0058129 | 0.51242068 | 12702 |
| Eye_age_gap | Reproductive_female | 0.0022315 | 0.89063906 | 3799 |
| Hepatic_age_gap | Reproductive_female | 0.12115393 | 9.66E-43 | 12702 |
| Immune_age_gap | Reproductive_female | 0.01310079 | 0.13983159 | 12702 |
| Metabolic_age_gap | Reproductive_female | 0.07970028 | 2.34E-19 | 12702 |
| Musculoskeletal_age_gap | Reproductive_female | 0.08078878 | 7.57E-20 | 12702 |
| Pulmonary_age_gap | Reproductive_female | 0.0452416 | 3.38E-07 | 12702 |
| Renal_age_gap | Reproductive_female | 0.15638753 | 2.34E-70 | 12702 |
| Brain_age_gap | Reproductive_male | 0.01891859 | 0.22098634 | 4187 |
| Cardiovascular_age_gap | Reproductive_male | -0.0016441 | 0.85300898 | 12702 |
| Eye_age_gap | Reproductive_male | 0.01391099 | 0.39134681 | 3799 |
| Hepatic_age_gap | Reproductive_male | 0.08981435 | 3.60E-24 | 12702 |
| Immune_age_gap | Reproductive_male | 0.08373203 | 3.31E-21 | 12702 |
| Metabolic_age_gap | Reproductive_male | 0.01981359 | 0.02554543 | 12702 |
| Musculoskeletal_age_gap | Reproductive_male | 0.05378584 | 1.31E-09 | 12702 |
| Pulmonary_age_gap | Reproductive_male | 0.07160602 | 6.49E-16 | 12702 |
| Renal_age_gap | Reproductive_male | 0.28433707 | 8.07E-235 | 12702 |
| Brain_age_gap | Skin | 0.05357694 | 0.00052384 | 4187 |
| Cardiovascular_age_gap | Skin | 0.00086984 | 0.92191339 | 12702 |
| Eye_age_gap | Skin | 0.01951363 | 0.22918492 | 3799 |
| Hepatic_age_gap | Skin | 0.05816893 | 5.35E-11 | 12702 |
| Immune_age_gap | Skin | 0.02066317 | 0.01986841 | 12702 |
| Metabolic_age_gap | Skin | 0.12109873 | 1.05E-42 | 12702 |
| Musculoskeletal_age_gap | Skin | 0.03582295 | 5.38E-05 | 12702 |
| Pulmonary_age_gap | Skin | 0.05243049 | 3.37E-09 | 12702 |
| Renal_age_gap | Skin | 0.04911023 | 3.07E-08 | 12702 |

b) **Genetic correlation**

[a]For genetic correlations with the eye ProtBAG, LDSC analysis failed to converge, prompting us to use an alternative tool, GNOVA, to estimate genetic correlations. In our previous research[10], I found that while LDSC and GNOVA produced genetic correlation estimates of differing magnitudes, the results were highly correlated.

| **PhenoBAG** | **ProtBAG** | **gc_mean** | **gc_std** | **Z** | **P** |
|---|---|---|---|---|---|
| Brain_age_gap | Eye[a] | | | | |

| | | | | | |
|---|---|---|---|---|---|
| Brain_age_gap | Reproductive_female | 0.3106 | 0.1899 | 1.6358 | 0.1019 |
| Brain_age_gap | Pulmonary | -0.0101 | 0.0644 | -0.1575 | 0.8748 |
| Brain_age_gap | Heart | 0.1161 | 0.0824 | 1.4076 | 0.1593 |
| Brain_age_gap | Brain | -0.0118 | 0.0812 | -0.145 | 0.8847 |
| Brain_age_gap | Hepatic | 0.0255 | 0.076 | 0.336 | 0.7369 |
| Brain_age_gap | Renal | -0.0487 | 0.0617 | -0.7892 | 0.43 |
| Brain_age_gap | Reproductive_male | 0.0547 | 0.0657 | 0.8335 | 0.4045 |
| Brain_age_gap | Endocrine | 0.1183 | 0.0843 | 1.4032 | 0.1606 |
| Brain_age_gap | Immune | 0.0689 | 0.0579 | 1.1901 | 0.234 |
| Brain_age_gap | Skin | -0.0202 | 0.087 | -0.2324 | 0.8162 |
| Cardiovascular_age_gap | Eye | | | | |
| Cardiovascular_age_gap | Reproductive_female | -0.0826 | 0.1133 | -0.7292 | 0.4659 |
| Cardiovascular_age_gap | Pulmonary | 0.0275 | 0.0513 | 0.537 | 0.5913 |
| Cardiovascular_age_gap | Heart | 0.0807 | 0.0753 | 1.0715 | 0.2839 |
| Cardiovascular_age_gap | Brain | 0.0074 | 0.058 | 0.1275 | 0.8985 |
| Cardiovascular_age_gap | Hepatic | 0.1199 | 0.0588 | 2.0394 | 0.0414 |
| Cardiovascular_age_gap | Renal | 0.0037 | 0.0488 | 0.0751 | 0.9402 |
| Cardiovascular_age_gap | Reproductive_male | -0.0036 | 0.0465 | -0.0772 | 0.9384 |
| Cardiovascular_age_gap | Endocrine | -0.0279 | 0.0692 | -0.4034 | 0.6866 |
| Cardiovascular_age_gap | Immune | 0.0129 | 0.0455 | 0.2832 | 0.777 |
| Cardiovascular_age_gap | Skin | 0.0187 | 0.0622 | 0.3001 | 0.7641 |
| Eye_age_gap | Eye | | | | |
| Eye_age_gap | Reproductive_female | -0.0833 | 0.131 | -0.6358 | 0.5249 |
| Eye_age_gap | Pulmonary | 0.0742 | 0.0618 | 1.2004 | 0.23 |
| Eye_age_gap | Heart | -0.0509 | 0.0855 | -0.5957 | 0.5513 |
| Eye_age_gap | Brain | 0.1952 | 0.0862 | 2.2649 | 0.0235 |
| Eye_age_gap | Hepatic | 0.0677 | 0.0774 | 0.8749 | 0.3816 |
| Eye_age_gap | Renal | 0.0065 | 0.0599 | 0.1088 | 0.9133 |
| Eye_age_gap | Reproductive_male | 0.0306 | 0.0659 | 0.464 | 0.6427 |
| Eye_age_gap | Endocrine | 0.0092 | 0.079 | 0.1168 | 0.9071 |
| Eye_age_gap | Immune | 0.0151 | 0.0568 | 0.2662 | 0.7901 |
| Eye_age_gap | Skin | 0.0941 | 0.0755 | 1.2456 | 0.2129 |
| Hepatic_age_gap | Eye | | | | |
| Hepatic_age_gap | Reproductive_female | 0.3782 | 0.1741 | 2.1724 | 0.0298 |
| Hepatic_age_gap | Pulmonary | 0.1587 | 0.0659 | 2.409 | 0.016 |
| Hepatic_age_gap | Heart | 0.2536 | 0.0868 | 2.9203 | 0.0035 |
| Hepatic_age_gap | Brain | 0.0451 | 0.0689 | 0.6543 | 0.5129 |
| Hepatic_age_gap | Hepatic | 0.3226 | 0.0787 | 4.0965 | 4.19E-05 |
| Hepatic_age_gap | Renal | -0.0438 | 0.0654 | -0.6693 | 0.5033 |
| Hepatic_age_gap | Reproductive_male | 0.0904 | 0.0681 | 1.3272 | 0.1844 |
| Hepatic_age_gap | Endocrine | 0.0987 | 0.0669 | 1.477 | 0.1397 |
| Hepatic_age_gap | Immune | 0.0946 | 0.057 | 1.661 | 0.0967 |
| Hepatic_age_gap | Skin | 0.0803 | 0.0796 | 1.0083 | 0.3133 |
| Immune_age_gap | Eye | | | | |
| Immune_age_gap | Reproductive_female | -0.002 | 0.1178 | -0.0169 | 0.9865 |
| Immune_age_gap | Pulmonary | 0.0654 | 0.0636 | 1.0277 | 0.3041 |
| Immune_age_gap | Heart | -0.0322 | 0.0833 | -0.3867 | 0.699 |
| Immune_age_gap | Brain | 0.1232 | 0.0868 | 1.4188 | 0.156 |
| Immune_age_gap | Hepatic | -0.0022 | 0.0698 | -0.0322 | 0.9743 |
| Immune_age_gap | Renal | -0.0544 | 0.0548 | -0.9943 | 0.3201 |
| Immune_age_gap | Reproductive_male | -0.0544 | 0.0574 | -0.9484 | 0.3429 |
| Immune_age_gap | Endocrine | 0.0063 | 0.0674 | 0.0933 | 0.9257 |
| Immune_age_gap | Immune | 0.0584 | 0.0521 | 1.1197 | 0.2629 |
| Immune_age_gap | Skin | -0.1082 | 0.0726 | -1.4899 | 0.1363 |
| Metabolic_age_gap | Eye | | | | |
| Metabolic_age_gap | Reproductive_female | 0.4393 | 0.2071 | 2.1213 | 0.0339 |

| | | | | | |
|---|---|---|---|---|---|
| Metabolic_age_gap | Pulmonary | 0.0627 | 0.0697 | 0.8996 | 0.3683 |
| Metabolic_age_gap | Heart | 0.0902 | 0.0862 | 1.0466 | 0.2953 |
| Metabolic_age_gap | Brain | 0.0074 | 0.0709 | 0.1039 | 0.9173 |
| Metabolic_age_gap | Hepatic | 0.3052 | 0.0845 | 3.6109 | 0.0003 |
| Metabolic_age_gap | Renal | 0.1868 | 0.0584 | 3.1979 | 0.0014 |
| Metabolic_age_gap | Reproductive_male | -0.0024 | 0.0571 | -0.0416 | 0.9668 |
| Metabolic_age_gap | Endocrine | 0.023 | 0.0675 | 0.3409 | 0.7332 |
| Metabolic_age_gap | Immune | -0.0286 | 0.0569 | -0.5031 | 0.6149 |
| Metabolic_age_gap | Skin | 0.1423 | 0.0734 | 1.9391 | 0.0525 |
| Musculoskeletal_age_gap | Eye | | | | |
| Musculoskeletal_age_gap | Reproductive_female | 0.2179 | 0.136 | 1.6026 | 0.109 |
| Musculoskeletal_age_gap | Pulmonary | -0.0014 | 0.0584 | -0.0248 | 0.9802 |
| Musculoskeletal_age_gap | Heart | -0.0471 | 0.0645 | -0.7314 | 0.4646 |
| Musculoskeletal_age_gap | Brain | 0.0437 | 0.0641 | 0.6819 | 0.4953 |
| Musculoskeletal_age_gap | Hepatic | 0.1454 | 0.0708 | 2.0542 | 0.04 |
| Musculoskeletal_age_gap | Renal | -0.0696 | 0.0568 | -1.2255 | 0.2204 |
| Musculoskeletal_age_gap | Reproductive_male | -0.0718 | 0.0602 | -1.1916 | 0.2334 |
| Musculoskeletal_age_gap | Endocrine | -0.0879 | 0.0665 | -1.3214 | 0.1864 |
| Musculoskeletal_age_gap | Immune | -0.0245 | 0.0506 | -0.4833 | 0.6289 |
| Musculoskeletal_age_gap | Skin | 0.0158 | 0.0482 | 0.3284 | 0.7426 |
| Pulmonary_age_gap | Eye | | | | |
| Pulmonary_age_gap | Reproductive_female | 0.1563 | 0.1128 | 1.385 | 0.1661 |
| Pulmonary_age_gap | Pulmonary | -0.0968 | 0.0533 | -1.8154 | 0.0695 |
| Pulmonary_age_gap | Heart | 0.0815 | 0.0698 | 1.1689 | 0.2424 |
| Pulmonary_age_gap | Brain | 0.0239 | 0.0669 | 0.3572 | 0.7209 |
| Pulmonary_age_gap | Hepatic | 0.0348 | 0.0553 | 0.6297 | 0.5289 |
| Pulmonary_age_gap | Renal | 0.0592 | 0.0448 | 1.3212 | 0.1864 |
| Pulmonary_age_gap | Reproductive_male | 0.036 | 0.0597 | 0.6035 | 0.5462 |
| Pulmonary_age_gap | Endocrine | 0.056 | 0.0593 | 0.9448 | 0.3448 |
| Pulmonary_age_gap | Immune | -0.0286 | 0.0518 | -0.5523 | 0.5807 |
| Pulmonary_age_gap | Skin | 0.1282 | 0.0616 | 2.0829 | 0.0373 |
| Renal_age_gap | Eye | | | | |
| Renal_age_gap | Reproductive_female | 0.1898 | 0.1127 | 1.6839 | 0.0922 |
| Renal_age_gap | Pulmonary | 0.3017 | 0.0546 | 5.5255 | 3.29E-08 |
| Renal_age_gap | Heart | 0.2595 | 0.0723 | 3.591 | 0.0003 |
| Renal_age_gap | Brain | 0.1772 | 0.0591 | 2.9995 | 0.0027 |
| Renal_age_gap | Hepatic | 0.2948 | 0.0612 | 4.8161 | 1.46E-06 |
| Renal_age_gap | Renal | 0.1762 | 0.0507 | 3.4728 | 0.0005 |
| Renal_age_gap | Reproductive_male | 0.3412 | 0.1138 | 2.9986 | 0.0027 |
| Renal_age_gap | Endocrine | 0.2263 | 0.0704 | 3.2141 | 0.0013 |
| Renal_age_gap | Immune | 0.205 | 0.0516 | 3.9691 | 7.21E-05 |
| Renal_age_gap | Skin | 0.0362 | 0.0484 | 0.7475 | 0.4548 |

**eTable 5b: Phenotypic and genetic correlations between the 11 ProtBAGs and 2448 proteins**

- **Phenotypic correlation**

I show significant results after Bonferroni correction based on the unique number of ProtBAGs and proteins (P-value < 0.05/2923/11). Here, I show only the 132 ProtBAG-protein pairs that are both genetically and phenotypically significant. Inf indicates the P-value's precision exceeds the number of decimals.

| Protein | ProtBAG | N | Beta | SE | -log$_{10}$(P) | r_coef |
|---|---|---|---|---|---|---|
| CST3 | Hepatic | 42658 | 0.98844335 | 0.01357811 | inf | 0.27147985 |
| AFP | Hepatic | 42083 | 0.37141558 | 0.00613606 | inf | 0.28468758 |
| AMBP | Hepatic | 42830 | 1.54202523 | 0.01924937 | inf | 0.32169633 |
| ELN | Hepatic | 36597 | 0.85052586 | 0.0135495 | inf | 0.27816653 |
| HSPG2 | Heart | 42238 | 0.99179547 | 0.01524247 | inf | 0.28720775 |
| NTproBNP | Heart | 40341 | 0.50843467 | 0.00364804 | inf | 0.53553548 |
| NPPB | Heart | 18389 | 0.84126532 | 0.00258803 | inf | 0.90497439 |
| LTBP2 | Heart | 43063 | 0.91354155 | 0.01279535 | inf | 0.27908302 |
| SMOC2 | Heart | 42729 | 0.49676541 | 0.01082249 | inf | 0.19254988 |
| COL6A3 | Heart | 43063 | 0.72543766 | 0.01096384 | inf | 0.29157605 |
| COLEC12 | Heart | 42519 | 0.88921295 | 0.01506547 | inf | 0.25764191 |
| CX3CL1 | Heart | 42210 | 0.42911547 | 0.0129497 | 237.003649 | 0.15975097 |
| CRIM1 | Heart | 42729 | 1.26008584 | 0.02040869 | inf | 0.27748098 |
| CD93 | Heart | 40947 | 0.72265164 | 0.01403127 | inf | 0.24327323 |
| EDA2R | Heart | 42890 | 0.69440456 | 0.0109029 | inf | 0.23640379 |
| EFEMP1 | Heart | 42660 | 0.79566092 | 0.01307874 | inf | 0.26108375 |
| AGER | Heart | 42444 | 0.40477127 | 0.01041444 | inf | 0.18689696 |
| BMP10 | Heart | 35769 | 0.9073669 | 0.01704242 | inf | 0.26545437 |
| PDCD1 | Pulmonary | 42820 | 0.7231143 | 0.00947118 | inf | 0.33104124 |
| PHOSPHO1 | Pulmonary | 42207 | 0.66139599 | 0.01406 | inf | 0.23033648 |
| OGFR | Pulmonary | 42327 | 0.60849781 | 0.01261556 | inf | 0.2375024 |
| POLR2F | Pulmonary | 42327 | 0.63377553 | 0.01171713 | inf | 0.25537588 |
| PLAU | Pulmonary | 42545 | 1.09777528 | 0.01391087 | inf | 0.33242727 |
| PLAUR | Pulmonary | 42234 | 1.57062388 | 0.01260368 | inf | 0.48827787 |
| IL2RA | Pulmonary | 42240 | 0.8678876 | 0.00966931 | inf | 0.40138246 |
| IL18BP | Pulmonary | 42730 | 1.49200026 | 0.01192256 | inf | 0.50513967 |
| ITGB2 | Pulmonary | 42301 | 0.91972755 | 0.01444129 | inf | 0.26030783 |
| HAVCR2 | Pulmonary | 42545 | 1.21736183 | 0.01039029 | inf | 0.47254407 |
| IFNGR1 | Pulmonary | 42729 | 1.58931079 | 0.01930492 | inf | 0.36377952 |
| IL10RB | Pulmonary | 42729 | 1.23430979 | 0.01328997 | inf | 0.40927713 |
| MSR1 | Pulmonary | 42901 | 1.20974292 | 0.00710549 | inf | 0.59655315 |
| NBL1 | Pulmonary | 42671 | 1.22672752 | 0.01485994 | inf | 0.36584818 |
| MMP12 | Pulmonary | 42553 | 0.52005904 | 0.0074174 | inf | 0.30048899 |
| NPDC1 | Pulmonary | 42569 | 1.02680015 | 0.01258957 | inf | 0.35386 |
| LGALS9 | Pulmonary | 42523 | 1.29050771 | 0.01097825 | inf | 0.46437125 |
| LRRC25 | Pulmonary | 42327 | 0.89249859 | 0.01001125 | inf | 0.37146601 |
| LAMP3 | Pulmonary | 42924 | 1.02583705 | 0.00579108 | inf | 0.63206389 |
| MANSC1 | Pulmonary | 42671 | 0.978635 | 0.01498128 | inf | 0.29770092 |
| SIGLEC10 | Pulmonary | 42454 | 0.98945777 | 0.0116411 | inf | 0.37127555 |
| SIGLEC1 | Pulmonary | 42728 | 0.93032483 | 0.00884754 | inf | 0.4429158 |
| SFTPD | Pulmonary | 42569 | 0.55399398 | 0.00584317 | inf | 0.41311384 |
| RELT | Pulmonary | 42545 | 1.25893011 | 0.01356599 | inf | 0.40842292 |
| PTGDS | Pulmonary | 42239 | 1.08436943 | 0.01342768 | inf | 0.37031078 |
| SCGB1A1 | Pulmonary | 42583 | 0.68610763 | 0.00691593 | inf | 0.4231482 |
| XCL1 | Pulmonary | 42892 | 0.38341511 | 0.00747562 | inf | 0.23400756 |
| ULBP2 | Pulmonary | 42210 | 0.78754517 | 0.01093866 | inf | 0.31584725 |

| | | | | | | |
|---|---|---|---|---|---|---|
| VCAM1 | Pulmonary | 42656 | 1.18285829 | 0.01481869 | inf | 0.35335327 |
| TGFBR2 | Pulmonary | 42671 | 1.03742717 | 0.01173861 | inf | 0.37545185 |
| THBD | Pulmonary | 42569 | 1.169621 | 0.01410143 | inf | 0.37684393 |
| TREML2 | Pulmonary | 42545 | 0.73144353 | 0.01220358 | inf | 0.26456505 |
| TNFRSF9 | Pulmonary | 42210 | 0.8974331 | 0.00951755 | inf | 0.40184474 |
| TNF | Pulmonary | 41693 | 0.77430067 | 0.01080668 | inf | 0.32280286 |
| TNFRSF8 | Pulmonary | 42909 | 0.76487033 | 0.0087645 | inf | 0.3751608 |
| TNFRSF12A | Pulmonary | 42670 | 0.83501668 | 0.01122574 | inf | 0.33132691 |
| TNFRSF14 | Pulmonary | 42590 | 1.10456695 | 0.01293862 | inf | 0.38091154 |
| TNFRSF19 | Pulmonary | 42892 | 0.89054832 | 0.0128367 | inf | 0.31627383 |
| TNFRSF1A | Pulmonary | 42545 | 1.30320856 | 0.01233315 | inf | 0.444282 |
| TNFRSF1B | Pulmonary | 42545 | 1.10777947 | 0.01016995 | inf | 0.45687788 |
| TNFRSF21 | Pulmonary | 42909 | 1.33237535 | 0.01419113 | inf | 0.3932833 |
| TNFRSF4 | Pulmonary | 42924 | 1.05517314 | 0.00996258 | inf | 0.44071236 |
| COLEC12 | Pulmonary | 42519 | 1.33692855 | 0.01414905 | inf | 0.37542366 |
| CLEC7A | Pulmonary | 42684 | 0.655648 | 0.00867429 | inf | 0.33399569 |
| CX3CL1 | Pulmonary | 42210 | 0.61473396 | 0.01267622 | inf | 0.21596788 |
| CXCL10 | Pulmonary | 42729 | 0.4694427 | 0.00594459 | inf | 0.33872995 |
| CSF1 | Pulmonary | 42729 | 1.3244525 | 0.01396452 | inf | 0.40649131 |
| CD74 | Pulmonary | 42909 | 1.18155017 | 0.01068471 | inf | 0.45484252 |
| CD79B | Pulmonary | 42995 | 0.74048981 | 0.0087848 | inf | 0.37394416 |
| CD83 | Pulmonary | 42454 | 1.33721622 | 0.01039571 | inf | 0.50520099 |
| CD93 | Pulmonary | 40947 | 0.91949543 | 0.01364052 | inf | 0.31579315 |
| CD38 | Pulmonary | 42327 | 0.87895776 | 0.01285702 | inf | 0.31616657 |
| CD163 | Pulmonary | 42730 | 0.8834456 | 0.00866385 | inf | 0.4303866 |
| CD27 | Pulmonary | 42671 | 0.80419659 | 0.00966505 | inf | 0.36800999 |
| CD274 | Pulmonary | 42210 | 0.88585421 | 0.01201976 | inf | 0.34626867 |
| CDCP1 | Pulmonary | 42210 | 0.85799501 | 0.00716934 | inf | 0.45271041 |
| CHIT1 | Pulmonary | 42684 | 0.07617076 | 0.00284515 | 155.869235 | 0.12880035 |
| CKAP4 | Pulmonary | 42789 | 1.17798337 | 0.01451625 | inf | 0.35588154 |
| CDH3 | Pulmonary | 42909 | 0.77262231 | 0.01140864 | inf | 0.31617315 |
| CXCL9 | Pulmonary | 42729 | 0.41920543 | 0.00614789 | inf | 0.28683001 |
| FOLR1 | Pulmonary | 42471 | 1.1786812 | 0.01382789 | inf | 0.33757125 |
| FOLR2 | Pulmonary | 42909 | 1.03308699 | 0.01344212 | inf | 0.30931874 |
| DLL1 | Pulmonary | 42671 | 1.19492325 | 0.01301549 | inf | 0.40371443 |
| EPHA2 | Pulmonary | 40983 | 1.06806255 | 0.01328986 | inf | 0.35148515 |
| EDA2R | Pulmonary | 42890 | 0.75738155 | 0.01072016 | inf | 0.26014521 |
| EFNA4 | Pulmonary | 42144 | 1.2135842 | 0.01266343 | inf | 0.41300144 |
| CCL7 | Pulmonary | 42442 | 0.45139055 | 0.00688781 | inf | 0.29354021 |
| ALCAM | Pulmonary | 43063 | 1.82846739 | 0.02008647 | inf | 0.33553756 |
| ADAM8 | Pulmonary | 42836 | 1.13260666 | 0.01370505 | inf | 0.36218837 |
| WFDC2 | Pulmonary | 40377 | 1.14257259 | 0.00914474 | inf | 0.49531979 |
| PTPRC | Pulmonary | 35700 | 1.37983519 | 0.02132865 | inf | 0.31101235 |
| PRRT3 | Pulmonary | 35637 | 0.52042012 | 0.01316763 | inf | 0.18255788 |
| TPK1 | Pulmonary | 36597 | 1.01577569 | 0.01736136 | inf | 0.29135112 |
| NPC2 | Pulmonary | 35982 | 1.26813808 | 0.01363485 | inf | 0.42483907 |
| OCLN | Pulmonary | 36522 | 0.77148379 | 0.01038678 | inf | 0.35808872 |
| NPL | Pulmonary | 36593 | 0.52818318 | 0.01056223 | inf | 0.25726938 |
| MRPL58 | Pulmonary | 27917 | 0.06620807 | 0.00808386 | 15.5655092 | 0.04947393 |
| CD72 | Pulmonary | 35769 | 0.76479145 | 0.01092597 | inf | 0.3349865 |
| CD80 | Pulmonary | 36597 | 1.44609478 | 0.0126038 | inf | 0.48731422 |
| SHISA5 | Pulmonary | 36589 | 1.41438819 | 0.0176593 | inf | 0.37008579 |
| RBFOX3 | Pulmonary | 36523 | 0.71916239 | 0.01227275 | inf | 0.28924006 |
| AHNAK | Pulmonary | 35637 | 0.86086823 | 0.01474382 | inf | 0.28843754 |
| FBLN2 | Pulmonary | 36596 | 0.83477185 | 0.01541209 | inf | 0.27136678 |
| CHGA | Pulmonary | 36595 | 0.17248172 | 0.00570147 | 197.857874 | 0.15083008 |

| | | | | | | |
|---|---|---|---|---|---|---|
| CSF1R | Pulmonary | 36632 | 0.82744182 | 0.01160392 | inf | 0.3256079 |
| IFI30 | Pulmonary | 35544 | 1.00354852 | 0.01132188 | inf | 0.40977328 |
| LMNB2 | Pulmonary | 36497 | 1.08685214 | 0.01628538 | inf | 0.32227549 |
| RBP2 | Brain | 42323 | 0.17984273 | 0.00548145 | 232.431399 | 0.14497632 |
| TNFRSF21 | Brain | 42909 | 0.72576992 | 0.01515813 | inf | 0.23082718 |
| CTSB | Brain | 42600 | 0.17191362 | 0.00796774 | 101.970906 | 0.10825448 |
| CD74 | Brain | 42909 | 0.59811279 | 0.0117448 | inf | 0.2181516 |
| SOST | Renal | 42501 | 2.16628412 | 0.00223346 | inf | 0.94395053 |
| HSPG2 | Endocrine | 42238 | 1.25264888 | 0.01426913 | inf | 0.34028352 |
| NBL1 | Endocrine | 42671 | 0.94566363 | 0.01495436 | inf | 0.25834324 |
| TNFRSF1A | Endocrine | 42545 | 0.97968264 | 0.01267775 | inf | 0.30536754 |
| DPT | Endocrine | 43063 | 1.0284714 | 0.01405217 | inf | 0.29904131 |
| EFEMP1 | Endocrine | 42660 | 0.76836498 | 0.01269936 | inf | 0.27928773 |
| CCN3 | Endocrine | 42998 | 1.10414037 | 0.01096633 | inf | 0.39275458 |
| SCARB2 | Immune | 41400 | 1.13093486 | 0.01267924 | inf | 0.37378369 |
| COLEC12 | Immune | 42519 | 1.15355382 | 0.01458648 | inf | 0.33674079 |
| CTSV | Immune | 42750 | -1.0446632 | 0.00913292 | inf | -0.4564611 |
| CD1C | Immune | 42671 | -1.0922952 | 0.01698193 | inf | -0.2799883 |
| EDA2R | Immune | 42890 | 0.78701531 | 0.01072019 | inf | 0.26719406 |
| EFEMP1 | Immune | 42660 | 0.80686099 | 0.01302137 | inf | 0.27058966 |
| SEPTIN8 | Immune | 36430 | 0.59488757 | 0.0150228 | inf | 0.18498427 |
| HBEGF | Skin | 42765 | 0.15318758 | 0.00583565 | 149.951626 | 0.12443576 |
| SPINT2 | Skin | 42728 | 0.24509936 | 0.0105615 | 117.676691 | 0.10688443 |
| TGFB1 | Skin | 42924 | 0.39602804 | 0.01092003 | 282.958621 | 0.17007471 |
| CST6 | Skin | 41397 | -0.9233347 | 0.00802483 | inf | -0.479194 |
| FABP9 | Skin | 42453 | -0.6904926 | 0.00705905 | inf | -0.4249063 |
| DSG3 | Skin | 42744 | -0.4763458 | 0.01250944 | inf | -0.1819268 |
| DSG4 | Skin | 42066 | -0.8291196 | 0.00801859 | inf | -0.438686 |
| PPBP | Skin | 36373 | 0.15927538 | 0.00595733 | 155.234544 | 0.14026537 |
| ENDOU | Skin | 33768 | -0.4604101 | 0.00989279 | inf | -0.2332085 |

- **Genetic correlation**

I show significant results after Bonferroni correction based on the unique number of ProtBAGs and proteins (P-value < 0.05/2416/11). I highlight (in bold font) several proteins that exhibited the strongest associations with the ProtBAG (higher genetic correlation) and were also organ-specific for the respective organ. Here, I show only the 132 ProtBAG-protein pairs that are both genetically and phenotypically significant.

| Protein | ProtBAG | gc_mean | gc_std | Z | P |
|---|---|---|---|---|---|
| PDCD1 | Pulmonary | 0.3728 | 0.0631 | 5.9038 | 3.55E-09 |
| PHOSPHO1 | Pulmonary | 0.4362 | 0.0858 | 5.0823 | 3.73E-07 |
| OGFR | Pulmonary | 0.462 | 0.0879 | 5.2543 | 1.49E-07 |
| POLR2F | Pulmonary | 0.4799 | 0.088 | 5.4505 | 5.02E-08 |
| PLAU | Pulmonary | 0.3889 | 0.0653 | 5.956 | 2.58E-09 |
| PLAUR | Pulmonary | 0.5197 | 0.0779 | 6.6698 | 2.56E-11 |
| IL2RA | Pulmonary | 0.3623 | 0.0661 | 5.4781 | 4.30E-08 |
| IL18BP | Pulmonary | 0.4881 | 0.0638 | 7.6559 | 1.92E-14 |
| ITGB2 | Pulmonary | 0.3807 | 0.0736 | 5.1726 | 2.31E-07 |
| HAVCR2 | Pulmonary | 0.4375 | 0.0617 | 7.0943 | 1.30E-12 |
| IFNGR1 | Pulmonary | 0.3789 | 0.0611 | 6.1975 | 5.74E-10 |
| IL10RB | Pulmonary | 0.3966 | 0.0762 | 5.2057 | 1.93E-07 |
| MSR1 | Pulmonary | 0.5516 | 0.0489 | 11.2894 | 1.48E-29 |
| NBL1 | Pulmonary | 0.4284 | 0.0757 | 5.6613 | 1.50E-08 |
| MMP12 | Pulmonary | 0.4281 | 0.0855 | 5.0077 | 5.51E-07 |

| | | | | | |
|---|---|---|---|---|---|
| NPDC1 | Pulmonary | 0.3847 | 0.0794 | 4.8428 | 1.28E-06 |
| LGALS9 | Pulmonary | 0.4809 | 0.0835 | 5.7564 | 8.59E-09 |
| LRRC25 | Pulmonary | 0.3832 | 0.065 | 5.8958 | 3.73E-09 |
| LAMP3 | Pulmonary | 0.6007 | 0.0542 | 11.0743 | 1.67E-28 |
| MANSC1 | Pulmonary | 0.2937 | 0.0574 | 5.1154 | 3.13E-07 |
| SIGLEC10 | Pulmonary | 0.3649 | 0.0663 | 5.503 | 3.73E-08 |
| SIGLEC1 | Pulmonary | 0.4988 | 0.0942 | 5.2942 | 1.20E-07 |
| SFTPD | Pulmonary | 0.3612 | 0.0582 | 6.2053 | 5.46E-10 |
| RELT | Pulmonary | 0.3901 | 0.0773 | 5.0501 | 4.42E-07 |
| PTGDS | Pulmonary | 0.359 | 0.0693 | 5.1813 | 2.20E-07 |
| SCGB1A1 | Pulmonary | 0.576 | 0.0492 | 11.7162 | 1.05E-31 |
| XCL1 | Pulmonary | 0.3907 | 0.0786 | 4.9728 | 6.60E-07 |
| ULBP2 | Pulmonary | 0.3282 | 0.063 | 5.2112 | 1.88E-07 |
| VCAM1 | Pulmonary | 0.292 | 0.0605 | 4.8267 | 1.39E-06 |
| TGFBR2 | Pulmonary | 0.3323 | 0.0627 | 5.3042 | 1.13E-07 |
| THBD | Pulmonary | 0.3655 | 0.0612 | 5.9709 | 2.36E-09 |
| TREML2 | Pulmonary | 0.3317 | 0.0625 | 5.308 | 1.11E-07 |
| TNFRSF9 | Pulmonary | 0.4648 | 0.0597 | 7.7813 | 7.18E-15 |
| TNF | Pulmonary | 0.566 | 0.1029 | 5.4976 | 3.85E-08 |
| TNFRSF8 | Pulmonary | 0.4757 | 0.0679 | 7.007 | 2.44E-12 |
| TNFRSF12A | Pulmonary | 0.383 | 0.0765 | 5.0045 | 5.60E-07 |
| TNFRSF14 | Pulmonary | 0.38 | 0.0692 | 5.4927 | 3.96E-08 |
| TNFRSF19 | Pulmonary | 0.4437 | 0.072 | 6.1648 | 7.06E-10 |
| TNFRSF1A | Pulmonary | 0.437 | 0.0649 | 6.735 | 1.64E-11 |
| TNFRSF1B | Pulmonary | 0.5768 | 0.075 | 7.6893 | 1.48E-14 |
| TNFRSF21 | Pulmonary | 0.3345 | 0.054 | 6.1888 | 6.06E-10 |
| TNFRSF4 | Pulmonary | 0.4665 | 0.06 | 7.7784 | 7.34E-15 |
| COLEC12 | Pulmonary | 0.381 | 0.071 | 5.3638 | 8.15E-08 |
| CLEC7A | Pulmonary | 0.4586 | 0.0674 | 6.8001 | 1.05E-11 |
| CX3CL1 | Pulmonary | 0.3446 | 0.0595 | 5.7925 | 6.93E-09 |
| CXCL10 | Pulmonary | 0.4424 | 0.0792 | 5.5884 | 2.29E-08 |
| CSF1 | Pulmonary | 0.3762 | 0.058 | 6.4838 | 8.94E-11 |
| CD74 | Pulmonary | 0.486 | 0.0665 | 7.3119 | 2.63E-13 |
| CD79B | Pulmonary | 0.3171 | 0.0611 | 5.1899 | 2.10E-07 |
| CD83 | Pulmonary | 0.4415 | 0.0626 | 7.0578 | 1.69E-12 |
| CD93 | Pulmonary | 0.3179 | 0.0653 | 4.8691 | 1.12E-06 |
| CD38 | Pulmonary | 0.362 | 0.0702 | 5.1566 | 2.51E-07 |
| CD163 | Pulmonary | 0.3538 | 0.0677 | 5.2259 | 1.73E-07 |
| CD27 | Pulmonary | 0.3928 | 0.0798 | 4.9204 | 8.64E-07 |
| CD274 | Pulmonary | 0.4821 | 0.0915 | 5.2674 | 1.38E-07 |
| CDCP1 | Pulmonary | 0.4368 | 0.07 | 6.24 | 4.37E-10 |
| CHIT1 | Pulmonary | 0.4368 | 0.07 | 6.24 | 4.37E-10 |
| CKAP4 | Pulmonary | 0.3849 | 0.0736 | 5.2273 | 1.72E-07 |
| CDH3 | Pulmonary | 0.3294 | 0.0605 | 5.4423 | 5.26E-08 |
| CXCL9 | Pulmonary | 0.3751 | 0.0708 | 5.3014 | 1.15E-07 |
| FOLR1 | Pulmonary | 0.4372 | 0.0632 | 6.9178 | 4.59E-12 |
| FOLR2 | Pulmonary | 0.2986 | 0.0571 | 5.232 | 1.68E-07 |
| DLL1 | Pulmonary | 0.4114 | 0.0644 | 6.3878 | 1.68E-10 |
| EPHA2 | Pulmonary | 0.4065 | 0.0772 | 5.263 | 1.42E-07 |
| EDA2R | Pulmonary | 0.3464 | 0.0671 | 5.1583 | 2.49E-07 |
| EFNA4 | Pulmonary | 0.4291 | 0.0667 | 6.4303 | 1.27E-10 |
| CCL7 | Pulmonary | 0.4156 | 0.0861 | 4.83 | 1.37E-06 |
| ALCAM | Pulmonary | 0.3745 | 0.0609 | 6.1522 | 7.64E-10 |
| ADAM8 | Pulmonary | 0.381 | 0.0711 | 5.3553 | 8.54E-08 |
| WFDC2 | Pulmonary | 0.4957 | 0.0669 | 7.414 | 1.23E-13 |
| PTPRC | Pulmonary | 0.4403 | 0.0803 | 5.4827 | 4.19E-08 |

| | | | | | |
|---|---|---|---|---|---|
| PRRT3 | Pulmonary | 0.2977 | 0.0622 | 4.79 | 1.67E-06 |
| TPK1 | Pulmonary | 0.4923 | 0.0754 | 6.5327 | 6.46E-11 |
| NPC2 | Pulmonary | 0.5736 | 0.0696 | 8.2445 | 1.66E-16 |
| OCLN | Pulmonary | 0.5038 | 0.0817 | 6.1688 | 6.88E-10 |
| NPL | Pulmonary | 0.367 | 0.0747 | 4.9135 | 8.94E-07 |
| MRPL58 | Pulmonary | 0.367 | 0.0747 | 4.9135 | 8.94E-07 |
| CD72 | Pulmonary | 0.4608 | 0.065 | 7.0945 | 1.30E-12 |
| CD80 | Pulmonary | 0.5392 | 0.1037 | 5.199 | 2.00E-07 |
| SHISA5 | Pulmonary | 0.4118 | 0.077 | 5.349 | 8.84E-08 |
| RBFOX3 | Pulmonary | 0.5072 | 0.086 | 5.8948 | 3.75E-09 |
| AHNAK | Pulmonary | 0.4205 | 0.0769 | 5.4697 | 4.51E-08 |
| FBLN2 | Pulmonary | 0.3535 | 0.0739 | 4.7836 | 1.72E-06 |
| CHGA | Pulmonary | 0.4281 | 0.087 | 4.9196 | 8.67E-07 |
| CSF1R | Pulmonary | 0.3569 | 0.066 | 5.4098 | 6.31E-08 |
| IFI30 | Pulmonary | 0.4562 | 0.0847 | 5.3865 | 7.18E-08 |
| LMNB2 | Pulmonary | 0.5117 | 0.0912 | 5.6137 | 1.98E-08 |
| HSPG2 | Heart | 0.6616 | 0.0895 | 7.3898 | 1.47E-13 |
| NTproBNP | Heart | 0.6503 | 0.066 | 9.8477 | 7.01E-23 |
| NPPB | Heart | 0.8421 | 0.0725 | 11.6222 | 3.18E-31 |
| LTBP2 | Heart | 0.4727 | 0.0806 | 5.8662 | 4.46E-09 |
| SMOC2 | Heart | 0.4357 | 0.09 | 4.8389 | 1.31E-06 |
| COL6A3 | Heart | 0.5161 | 0.0874 | 5.905 | 3.53E-09 |
| COLEC12 | Heart | 0.5014 | 0.0982 | 5.1056 | 3.30E-07 |
| CX3CL1 | Heart | 0.5047 | 0.0964 | 5.2363 | 1.64E-07 |
| CRIM1 | Heart | 0.5807 | 0.0892 | 6.5077 | 7.63E-11 |
| CD93 | Heart | 0.4481 | 0.0884 | 5.0674 | 4.03E-07 |
| EDA2R | Heart | 0.5067 | 0.0905 | 5.5978 | 2.17E-08 |
| EFEMP1 | Heart | 0.4763 | 0.0877 | 5.4324 | 5.56E-08 |
| AGER | Heart | 0.4253 | 0.0826 | 5.1487 | 2.62E-07 |
| BMP10 | Heart | 0.4837 | 0.0935 | 5.172 | 2.32E-07 |
| RBP2 | Brain | 0.5515 | 0.1088 | 5.0711 | 3.95E-07 |
| TNFRSF21 | Brain | 0.3906 | 0.0768 | 5.0864 | 3.65E-07 |
| CTSB | Brain | 0.2216 | 0.0458 | 4.8376 | 1.31E-06 |
| CD74 | Brain | 0.3629 | 0.0665 | 5.4587 | 4.80E-08 |
| CST3 | Hepatic | 0.424 | 0.0876 | 4.841 | 1.29E-06 |
| AFP | Hepatic | 0.5352 | 0.0806 | 6.6385 | 3.17E-11 |
| AMBP | Hepatic | 0.4024 | 0.0832 | 4.8379 | 1.31E-06 |
| ELN | Hepatic | 0.4341 | 0.085 | 5.1094 | 3.23E-07 |
| SOST | Renal | 1.0061 | 0.0047 | 213.7857 | 0 |
| HSPG2 | Endocrine | 0.5959 | 0.101 | 5.9009 | 3.61E-09 |
| NBL1 | Endocrine | 0.4634 | 0.0851 | 5.4446 | 5.19E-08 |
| TNFRSF1A | Endocrine | 0.4079 | 0.0818 | 4.9849 | 6.20E-07 |
| DPT | Endocrine | 0.4717 | 0.0973 | 4.8478 | 1.25E-06 |
| EFEMP1 | Endocrine | 0.4023 | 0.0799 | 5.0368 | 4.73E-07 |
| CCN3 | Endocrine | 0.5571 | 0.0782 | 7.1209 | 1.07E-12 |
| SCARB2 | Immune | 0.3051 | 0.0593 | 5.1412 | 2.73E-07 |
| COLEC12 | Immune | 0.3346 | 0.0584 | 5.7288 | 1.01E-08 |
| CTSV | Immune | -0.3967 | 0.0774 | -5.1244 | 2.98E-07 |
| CD1C | Immune | -0.3803 | 0.0646 | -5.8845 | 3.99E-09 |
| EDA2R | Immune | 0.2863 | 0.0597 | 4.7988 | 1.60E-06 |
| EFEMP1 | Immune | 0.333 | 0.0558 | 5.9702 | 2.37E-09 |
| SEPTIN8 | Immune | 0.5342 | 0.1109 | 4.8171 | 1.46E-06 |
| HBEGF | Skin | 0.3208 | 0.0631 | 5.0869 | 3.64E-07 |
| SPINT2 | Skin | 0.2623 | 0.0538 | 4.8727 | 1.10E-06 |
| TGFB1 | Skin | 0.3061 | 0.0597 | 5.1246 | 2.98E-07 |
| CST6 | Skin | -0.598 | 0.0743 | -8.0511 | 8.20E-16 |

| | | | | | |
|---|---|---|---|---|---|
| FABP9 | Skin | -0.4498 | 0.094 | -4.7843 | 1.72E-06 |
| DSG3 | Skin | -0.2966 | 0.0603 | -4.9209 | 8.61E-07 |
| DSG4 | Skin | -0.4976 | 0.0825 | -6.035 | 1.59E-09 |
| PPBP | Skin | 0.2759 | 0.0569 | 4.8449 | 1.27E-06 |
| ENDOU | Skin | -0.3195 | 0.0637 | -5.0181 | 5.22E-07 |

**eTable 6: The incremental $R^2$ of the 20 ProtBAG-PRS and PhenoBAG-PRS**

| BAG | BAG_type | R2 | P | BETA | SE |
|---|---|---|---|---|---|
| Brain_age_gap | PhenoBAG | 0.0240832 | 4.59E-81 | 0.50875151 | 0.02651691 |
| Cardiovascular_age_gap | PhenoBAG | 0.02408721 | 8.56E-296 | 0.84344925 | 0.02280597 |
| Eye_age_gap | PhenoBAG | 0.0201707 | 3.14E-81 | 0.46174613 | 0.02406622 |
| Hepatic_age_gap | PhenoBAG | 0.02585121 | 2.40E-317 | 0.95682729 | 0.02496 |
| Immune_age_gap | PhenoBAG | 0.02395875 | 6.27E-294 | 1.01862665 | 0.02763137 |
| Metabolic_age_gap | PhenoBAG | 0.05666742 | 0 | 1.25563439 | 0.02178132 |
| Musculoskeletal_age_gap | PhenoBAG | 0.02213174 | 1.21E-271 | 0.84407316 | 0.02383554 |
| Pulmonary_age_gap | PhenoBAG | 0.03863284 | 0 | 0.92090906 | 0.01951636 |
| Renal_age_gap | PhenoBAG | 0.05917277 | 0 | 1.02108544 | 0.01730457 |
| Brain | ProtBAG | 0.02485321 | 7.16E-111 | 1.22499414 | 0.05440769 |
| Endocrine | ProtBAG | 0.02030364 | 1.00E-90 | 1.24049518 | 0.06109896 |
| Eye | ProtBAG | 0.05322734 | 1.71E-238 | 0.93904501 | 0.0280865 |
| Heart | ProtBAG | 0.26361125 | 0 | 1.42064625 | 0.01683927 |
| Hepatic | ProtBAG | 0.13894129 | 0 | 1.14047556 | 0.02013214 |
| Immune | ProtBAG | 0.0237082 | 8.89E-106 | 0.58780528 | 0.02674779 |
| Pulmonary | ProtBAG | 0.05030382 | 3.10E-225 | 1.50045418 | 0.04622828 |
| Renal | ProtBAG | 0.04768818 | 2.35E-213 | 1.51359025 | 0.04795954 |
| Reproductive_female | ProtBAG | 0.06372904 | 1.01E-286 | 1.5252037 | 0.04145659 |
| Reproductive_male | ProtBAG | 0.1381574 | 0 | 1.25953173 | 0.02230743 |
| Skin | ProtBAG | 0.05085876 | 9.23E-228 | 0.60323649 | 0.01847835 |

**eTable 7: The Mendelian randomization results**

| outcome | exposure | method | nsnp | b | se | pval | lo_ci | up_ci | or | or_lci95 | or_uci95 |
|---|---|---|---|---|---|---|---|---|---|---|---|
| Brain_age_gap | KRA_PSY_ANYMENTAL | MR Egger | 14 | 0.71153893 | 0.13415539 | 0.00018716 | 0.44859436 | 0.97448349 | 2.03712384 | 1.56610926 | 2.64979822 |
| Brain_age_gap | KRA_PSY_ANYMENTAL | Weighted median | 14 | 0.4376985 | 0.07944664 | 3.60E-08 | 0.28198308 | 0.59341392 | 1.54913777 | 1.32575629 | 1.81015761 |
| Brain_age_gap | KRA_PSY_ANYMENTAL | Weighted mode | 14 | 0.46412819 | 0.08211891 | 7.90E-05 | 0.30317512 | 0.62508126 | 1.59062686 | 1.35415158 | 1.86839777 |
| Brain_age_gap | G6_AD_WIDE | Weighted median | 8 | 0.10529143 | 0.0179543 | 4.51E-09 | 0.07010101 | 0.14048185 | 1.11103435 | 1.07261652 | 1.1508282 |
| Brain_age_gap | G6_AD_WIDE | Inverse variance weighted | 8 | 0.09631639 | 0.01683432 | 1.06E-08 | 0.06332112 | 0.12931166 | 1.10110739 | 1.0653689 | 1.13804475 |
| Brain_age_gap | AD | Weighted median | 20 | 0.0721702 | 0.01885535 | 0.00012941 | 0.03521371 | 0.10912669 | 1.07483827 | 1.03584105 | 1.11530364 |
| Brain_age_gap | AD | Inverse variance weighted | 20 | 0.06064019 | 0.01490523 | 4.73E-05 | 0.03142594 | 0.08985443 | 1.06251654 | 1.03192495 | 1.09401502 |
| Cardiovascular_age_gap | RX_ANTIHYP | Weighted median | 87 | 0.2243173 | 0.0215228 | 1.96E-25 | 0.18213261 | 0.26650199 | 1.25146804 | 1.19977328 | 1.30539018 |
| Cardiovascular_age_gap | RX_ANTIHYP | Inverse variance weighted | 87 | 0.2374233 | 0.02388637 | 2.80E-23 | 0.19060602 | 0.28424058 | 1.26797774 | 1.20998265 | 1.32875256 |
| Cardiovascular_age_gap | RX_ANTIHYP | Simple mode | 87 | 0.28170942 | 0.06824557 | 8.44E-05 | 0.1479481 | 0.41547075 | 1.32539354 | 1.15945272 | 1.51508379 |
| Cardiovascular_age_gap | RX_ANTIHYP | Weighted mode | 87 | 0.28170942 | 0.06389492 | 2.99E-05 | 0.15647538 | 0.40694347 | 1.32539354 | 1.16938197 | 1.50221918 |
| Cardiovascular_age_gap | FG_CVD | Weighted median | 34 | 0.20345548 | 0.03518329 | 7.35E-09 | 0.13449623 | 0.27241473 | 1.2256306 | 1.14396035 | 1.31313149 |
| Cardiovascular_age_gap | FG_CVD | Inverse variance weighted | 34 | 0.24965295 | 0.05353616 | 3.11E-06 | 0.14472208 | 0.35458382 | 1.28357987 | 1.15571833 | 1.42558722 |
| Cardiovascular_age_gap | I9_HYPTENS | Weighted median | 110 | 0.18471639 | 0.01661907 | 1.06E-28 | 0.15214302 | 0.21728976 | 1.20287724 | 1.16432675 | 1.24270414 |
| Cardiovascular_age_gap | I9_HYPTENS | Inverse variance weighted | 110 | 0.21027812 | 0.01814394 | 4.66E-31 | 0.174716 | 0.24584024 | 1.23402122 | 1.19090796 | 1.27869527 |
| Cardiovascular_age_gap | I9_HYPTENS | Simple mode | 110 | 0.19869493 | 0.04831567 | 7.62E-05 | 0.10399621 | 0.29339365 | 1.21980979 | 1.10959625 | 1.34097057 |
| Cardiovascular_age_gap | I9_HYPTENS | Weighted mode | 110 | 0.18722837 | 0.04077392 | 1.18E-05 | 0.10731149 | 0.26714525 | 1.20590264 | 1.11328097 | 1.30623016 |
| Cardiovascular_age_gap | I9_HYPTENSESS | Weighted median | 78 | 0.18496506 | 0.01980991 | 9.91E-21 | 0.14613764 | 0.22379249 | 1.2031764 | 1.15735547 | 1.25081143 |
| Cardiovascular_age_gap | I9_HYPTENSESS | Inverse variance weighted | 78 | 0.20426779 | 0.0233082 | 1.89E-18 | 0.15858372 | 0.24995186 | 1.22662659 | 1.17185002 | 1.28396361 |
| Cardiovascular_age_gap | I9_HYPTENSESS | Simple mode | 78 | 0.23369509 | 0.05532016 | 6.51E-05 | 0.12526757 | 0.3421226 | 1.26325925 | 1.13345169 | 1.4079329 |
| Cardiovascular_age_gap | I9_HYPTENSESS | Weighted mode | 78 | 0.18260678 | 0.04432762 | 9.47E-05 | 0.09572465 | 0.26948891 | 1.20034231 | 1.10045601 | 1.30929511 |
| Cardiovascular_age_gap | T2D | Weighted median | 92 | 0.05203129 | 0.01332633 | 9.45E-05 | 0.02591168 | 0.0781509 | 1.05340871 | 1.02625031 | 1.08128582 |

| | | | | | | | | | | | |
|---|---|---|---|---|---|---|---|---|---|---|---|
| Cardiovascular_age_gap | KELA_DIAB_INSUL_EXMORE | Weighted mode | 87 | 0.06774604 | 0.01739172 | 0.0001934 | 0.03365826 | 0.10183382 | 1.07009352 | 1.03423111 | 1.10719946 |
| Cardiovascular_age_gap | I9_AF | Weighted mode | 51 | -0.0480732 | 0.01166726 | 0.00014206 | -0.070941 | -0.0252053 | 0.95306405 | 0.93151685 | 0.97510967 |
| Cardiovascular_age_gap | T2D_WIDE | Weighted median | 62 | 0.05690941 | 0.01563049 | 0.00027166 | 0.02627364 | 0.08754518 | 1.05855991 | 1.02662184 | 1.09149158 |
| Cardiovascular_age_gap | I9_ANGINA | Weighted median | 32 | 0.07446862 | 0.01994856 | 0.00018919 | 0.03536944 | 0.11356781 | 1.07731154 | 1.03600238 | 1.12026785 |
| Cardiovascular_age_gap | I9_HEARTFAIL_AND_ANTIHYPERT | Weighted median | 15 | 0.09949154 | 0.02407362 | 3.58E-05 | 0.05230725 | 0.14667583 | 1.10460913 | 1.05369944 | 1.15797852 |
| Cardiovascular_age_gap | O15_HYPTENSPREG | Weighted median | 7 | 0.20734372 | 0.02924255 | 1.34E-12 | 0.15002831 | 0.26465912 | 1.23040541 | 1.16186714 | 1.30298674 |
| Cardiovascular_age_gap | O15_HYPTENSPREG | Inverse variance weighted | 7 | 0.2126276 | 0.03034027 | 2.42E-12 | 0.15316067 | 0.27209453 | 1.23692393 | 1.16551223 | 1.31271108 |
| Cardiovascular_age_gap | THYROTOXICOSIS | Weighted median | 23 | 0.03427493 | 0.00879288 | 9.70E-05 | 0.01704089 | 0.05150897 | 1.03486908 | 1.01718691 | 1.05285863 |
| Eye_age_gap | H7_AMD | Weighted median | 17 | 0.08895986 | 0.0174166 | 3.26E-07 | 0.05482331 | 0.12309641 | 1.09303678 | 1.05635396 | 1.13099345 |
| Eye_age_gap | H7_AMD | Weighted mode | 17 | 0.0931178 | 0.01843485 | 0.00011805 | 0.05698548 | 0.12925011 | 1.09759102 | 1.05864044 | 1.13797471 |
| Hepatic_age_gap | RX_STATIN | Weighted median | 87 | 0.05992046 | 0.013302 | 6.65E-06 | 0.03384853 | 0.08599239 | 1.06175209 | 1.03442791 | 1.08979803 |
| Hepatic_age_gap | RX_STATIN | Weighted mode | 87 | 0.07245017 | 0.01018406 | 3.15E-10 | 0.05248942 | 0.09241092 | 1.07513923 | 1.05389142 | 1.09681543 |
| Hepatic_age_gap | KRA_PSY_ANYMENTAL | MR Egger | 14 | 0.44100168 | 0.08019809 | 0.0001365 | 0.28381343 | 0.59818994 | 1.55426332 | 1.3281851 | 1.81882364 |
| Hepatic_age_gap | KRA_PSY_ANYMENTAL | Weighted median | 14 | 0.2539425 | 0.05162146 | 8.68E-07 | 0.15276443 | 0.35512057 | 1.28909768 | 1.1650505 | 1.42635261 |
| Hepatic_age_gap | KRA_PSY_ANYMENTAL | Weighted mode | 14 | 0.3096853 | 0.04116258 | 4.35E-06 | 0.22900663 | 0.39036396 | 1.36299611 | 1.25735038 | 1.47751846 |
| Hepatic_age_gap | E4_METABOLIA | Weighted median | 19 | 0.15078735 | 0.02250553 | 2.08E-11 | 0.10667651 | 0.19489819 | 1.16274937 | 1.11257429 | 1.21518727 |
| Hepatic_age_gap | E4_METABOLIA | Weighted mode | 19 | 0.15291628 | 0.02053238 | 6.69E-07 | 0.11267281 | 0.19315974 | 1.16522742 | 1.11926566 | 1.21307656 |
| Hepatic_age_gap | E4_LIPOPROT | Weighted median | 25 | 0.10085643 | 0.01521982 | 3.43E-11 | 0.07102558 | 0.13068727 | 1.10611782 | 1.07360869 | 1.13961134 |
| Hepatic_age_gap | E4_LIPOPROT | Weighted mode | 25 | 0.10006858 | 0.01434636 | 3.27E-07 | 0.07194971 | 0.12818745 | 1.10524671 | 1.0746013 | 1.13676607 |
| Hepatic_age_gap | E4_HYPERCHOL | Weighted median | 21 | 0.09803995 | 0.01441857 | 1.05E-11 | 0.06977954 | 0.12630035 | 1.10300684 | 1.07227177 | 1.1346229 |
| Hepatic_age_gap | E4_HYPERCHOL | Weighted mode | 21 | 0.10146902 | 0.01330182 | 2.41E-07 | 0.07539744 | 0.12754059 | 1.10679563 | 1.07831264 | 1.13603098 |
| Hepatic_age_gap | G6_AD_WIDE | Weighted median | 8 | 0.05242884 | 0.0096022 | 4.76E-08 | 0.03360853 | 0.07124915 | 1.05382757 | 1.03417967 | 1.07384875 |
| Hepatic_age_gap | E4_HYPERLIPNAS | Weighted median | 9 | 0.09166256 | 0.0147351 | 4.95E-10 | 0.06278177 | 0.12054334 | 1.09599492 | 1.06479444 | 1.12810964 |

| | | | | | | | | | | | |
|---|---|---|---|---|---|---|---|---|---|---|---|
| Hepatic_age_gap | E4_HYPERLIPNAS | Weighted mode | 9 | 0.08922017 | 0.01437895 | 0.00025804 | 0.06103744 | 0.11740291 | 1.09332135 | 1.06293871 | 1.12457244 |
| Hepatic_age_gap | DRY_AMD | Weighted median | 14 | 0.02331783 | 0.00610891 | 0.00013507 | 0.01134437 | 0.03529129 | 1.02359182 | 1.01140896 | 1.03592142 |
| Immune_age_gap | E4_LIPOPROT | Weighted median | 25 | -0.0988302 | 0.01953168 | 4.19E-07 | -0.1371123 | -0.0605481 | 0.90589648 | 0.87187227 | 0.94124846 |
| Immune_age_gap | E4_LIPOPROT | Inverse variance weighted | 25 | -0.0721223 | 0.01771944 | 4.70E-05 | -0.1068524 | -0.0373922 | 0.93041706 | 0.89865826 | 0.96329822 |
| Immune_age_gap | G6_AD_WIDE | Weighted median | 8 | -0.0418972 | 0.00938231 | 7.99E-06 | -0.0602866 | -0.0235079 | 0.95896831 | 0.9414947 | 0.97676623 |
| Immune_age_gap | G6_AD_WIDE | Inverse variance weighted | 8 | -0.0366601 | 0.00849411 | 1.59E-05 | -0.0533086 | -0.0200117 | 0.96400372 | 0.94808741 | 0.98018724 |
| Immune_age_gap | E4_HYPERLIPNAS | Weighted median | 9 | -0.0729693 | 0.01916023 | 0.00013988 | -0.1105234 | -0.0354153 | 0.92962934 | 0.89536539 | 0.9652045 |
| Metabolic_age_gap | RX_STATIN | MR Egger | 87 | 0.16764119 | 0.03591366 | 1.13E-05 | 0.09725042 | 0.23803195 | 1.18251223 | 1.10213634 | 1.26874973 |
| Metabolic_age_gap | RX_STATIN | Weighted median | 87 | 0.21593498 | 0.0147815 | 2.48E-48 | 0.18696324 | 0.24490671 | 1.24102168 | 1.20558297 | 1.27750213 |
| Metabolic_age_gap | RX_STATIN | Inverse variance weighted | 87 | 0.19529564 | 0.0247374 | 2.91E-15 | 0.14681034 | 0.24378094 | 1.21567033 | 1.15813429 | 1.27606476 |
| Metabolic_age_gap | RX_STATIN | Weighted mode | 87 | 0.24004312 | 0.01370069 | 4.09E-30 | 0.21318976 | 0.26689648 | 1.27130397 | 1.23761948 | 1.30590525 |
| Metabolic_age_gap | KRA_PSY_ANYMENTAL | MR Egger | 14 | 0.43396835 | 0.08141569 | 0.00017927 | 0.27439359 | 0.59354311 | 1.54337002 | 1.31573256 | 1.81039149 |
| Metabolic_age_gap | KRA_PSY_ANYMENTAL | Weighted mode | 14 | 0.38231956 | 0.04692702 | 1.83E-06 | 0.29034259 | 0.47429652 | 1.46568038 | 1.33688542 | 1.60688339 |
| Metabolic_age_gap | T2D | MR Egger | 92 | 0.19545751 | 0.04848782 | 0.000116 | 0.10042139 | 0.29049363 | 1.21586713 | 1.10563673 | 1.33708735 |
| Metabolic_age_gap | T2D | Weighted median | 92 | 0.16722789 | 0.01590264 | 7.31E-26 | 0.13605871 | 0.19839707 | 1.18202361 | 1.14574916 | 1.2194465 |
| Metabolic_age_gap | T2D | Inverse variance weighted | 92 | 0.13555481 | 0.02240261 | 1.44E-09 | 0.0916457 | 0.17946391 | 1.14517196 | 1.09597645 | 1.19657572 |
| Metabolic_age_gap | T2D | Weighted mode | 92 | 0.19152009 | 0.01862901 | 6.62E-17 | 0.15500722 | 0.22803296 | 1.21108916 | 1.16766639 | 1.25612673 |
| Metabolic_age_gap | KELA_DIAB_INSUL_EXMORE | MR Egger | 87 | 0.18663513 | 0.04618574 | 0.00011639 | 0.09611108 | 0.27715917 | 1.20518746 | 1.10088134 | 1.31937636 |
| Metabolic_age_gap | KELA_DIAB_INSUL_EXMORE | Weighted median | 87 | 0.15134663 | 0.01682323 | 2.33E-19 | 0.11837311 | 0.18432016 | 1.16339986 | 1.12566403 | 1.20240072 |
| Metabolic_age_gap | KELA_DIAB_INSUL_EXMORE | Inverse variance weighted | 87 | 0.13088876 | 0.02090519 | 3.82E-10 | 0.08991458 | 0.17186293 | 1.13984097 | 1.09408083 | 1.18751505 |
| Metabolic_age_gap | KELA_DIAB_INSUL_EXMORE | Weighted mode | 87 | 0.20281363 | 0.02155395 | 7.21E-15 | 0.16056788 | 0.24505937 | 1.22484417 | 1.17417748 | 1.27769717 |
| Metabolic_age_gap | E4_METABOLIA | Weighted median | 19 | 0.36587827 | 0.03437188 | 1.85E-26 | 0.29850939 | 0.43324715 | 1.44177972 | 1.34784819 | 1.54225734 |
| Metabolic_age_gap | E4_METABOLIA | Inverse variance weighted | 19 | 0.39323299 | 0.08533867 | 4.07E-06 | 0.22596921 | 0.56049678 | 1.48176359 | 1.25353707 | 1.75154241 |

| | | | | | | | | | | | |
|---|---|---|---|---|---|---|---|---|---|---|---|
| Metabolic_age_gap | E4_METABOLIA | Simple mode | 19 | 0.36721288 | 0.04527394 | 2.01E-07 | 0.27847595 | 0.4559498 | 1.44370522 | 1.32111484 | 1.57767114 |
| Metabolic_age_gap | E4_METABOLIA | Weighted mode | 19 | 0.3848004 | 0.03593767 | 3.09E-09 | 0.31436257 | 0.45523823 | 1.46932102 | 1.36938614 | 1.57654893 |
| Metabolic_age_gap | E4_DM2NASCOMP | MR Egger | 91 | 0.1759746 | 0.04362546 | 0.00011574 | 0.0904687 | 0.2614805 | 1.19240777 | 1.09468724 | 1.29885161 |
| Metabolic_age_gap | E4_DM2NASCOMP | Weighted median | 91 | 0.16164446 | 0.0140958 | 1.92E-30 | 0.13401669 | 0.18927223 | 1.17544225 | 1.14341191 | 1.20836986 |
| Metabolic_age_gap | E4_DM2NASCOMP | Inverse variance weighted | 91 | 0.1378001 | 0.02015183 | 8.03E-12 | 0.09830252 | 0.17729768 | 1.14774609 | 1.1032965 | 1.19398647 |
| Metabolic_age_gap | E4_DM2NASCOMP | Weighted mode | 91 | 0.17149673 | 0.01630111 | 2.38E-17 | 0.13954655 | 0.2034469 | 1.18708026 | 1.14975233 | 1.22562008 |
| Metabolic_age_gap | T2D_WIDE | Weighted median | 62 | 0.13519703 | 0.0187725 | 5.94E-13 | 0.09840292 | 0.17199114 | 1.14476231 | 1.10340728 | 1.18766731 |
| Metabolic_age_gap | T2D_WIDE | Inverse variance weighted | 62 | 0.13417714 | 0.02729708 | 8.86E-07 | 0.08067487 | 0.18767941 | 1.14359538 | 1.08401839 | 1.20644668 |
| Metabolic_age_gap | T2D_WIDE | Weighted mode | 62 | 0.16217247 | 0.02606194 | 4.92E-08 | 0.11109107 | 0.21325388 | 1.17606306 | 1.11749667 | 1.23769883 |
| Metabolic_age_gap | E4_LIPOPROT | Weighted median | 25 | 0.2933716 | 0.02012303 | 3.83E-48 | 0.25393047 | 0.33281273 | 1.34094099 | 1.28908217 | 1.39488605 |
| Metabolic_age_gap | E4_LIPOPROT | Inverse variance weighted | 25 | 0.28778873 | 0.05137185 | 2.12E-08 | 0.1870999 | 0.38847755 | 1.33347555 | 1.20574773 | 1.47473388 |
| Metabolic_age_gap | E4_LIPOPROT | Simple mode | 25 | 0.30381671 | 0.04260809 | 2.27E-07 | 0.22030485 | 0.38732857 | 1.35502067 | 1.24645665 | 1.47304041 |
| Metabolic_age_gap | E4_LIPOPROT | Weighted mode | 25 | 0.30381671 | 0.02774331 | 8.10E-11 | 0.24943983 | 0.35819359 | 1.35502067 | 1.28330634 | 1.43074257 |
| Metabolic_age_gap | E4_HYPERCHOL | Weighted median | 21 | 0.26899463 | 0.02008061 | 6.40E-41 | 0.22963663 | 0.30835263 | 1.30864811 | 1.25814276 | 1.3611809 |
| Metabolic_age_gap | E4_HYPERCHOL | Inverse variance weighted | 21 | 0.28538989 | 0.04916499 | 6.45E-09 | 0.18902651 | 0.38175326 | 1.33028059 | 1.20807298 | 1.4648506 |
| Metabolic_age_gap | E4_HYPERCHOL | Simple mode | 21 | 0.27788777 | 0.02808998 | 3.79E-09 | 0.2228314 | 0.33294414 | 1.32033801 | 1.24960988 | 1.39506937 |
| Metabolic_age_gap | E4_HYPERCHOL | Weighted mode | 21 | 0.27488701 | 0.02099904 | 2.88E-11 | 0.23372889 | 0.31604513 | 1.31638193 | 1.26330195 | 1.37169216 |
| Metabolic_age_gap | G6_AD_WIDE | Weighted median | 8 | 0.07519516 | 0.00993718 | 3.82E-14 | 0.05571828 | 0.09467205 | 1.07809454 | 1.05729978 | 1.09929828 |
| Metabolic_age_gap | G6_AD_WIDE | Inverse variance weighted | 8 | 0.0667324 | 0.01245897 | 8.50E-08 | 0.04231281 | 0.09115198 | 1.06900937 | 1.04322076 | 1.09543548 |
| Metabolic_age_gap | G6_AD_WIDE | Weighted mode | 8 | 0.07658649 | 0.0094576 | 8.43E-05 | 0.0580496 | 0.09512338 | 1.07959556 | 1.05976756 | 1.09979454 |
| Metabolic_age_gap | L12_PAPULOSQUAMOUS | Weighted median | 22 | 0.04517859 | 0.01195823 | 0.00015807 | 0.02174046 | 0.06861673 | 1.04621469 | 1.02197851 | 1.07102564 |
| Metabolic_age_gap | E4_HYPERLIPNAS | Weighted median | 9 | 0.2777936 | 0.02619762 | 2.86E-26 | 0.22644627 | 0.32914093 | 1.32021368 | 1.25413523 | 1.3897737 |
| Metabolic_age_gap | E4_HYPERLIPNAS | Inverse variance weighted | 9 | 0.33085245 | 0.05794607 | 1.13E-08 | 0.21727814 | 0.44442675 | 1.39215436 | 1.2426897 | 1.5595959 |

| | | | | | | | | | | | |
|---|---|---|---|---|---|---|---|---|---|---|---|
| Metabolic_age_gap | E4_HYPERLIPNAS | Simple mode | 9 | 0.27344273 | 0.03184047 | 2.61E-05 | 0.21103541 | 0.33585004 | 1.31448207 | 1.23495609 | 1.3991292 |
| Metabolic_age_gap | E4_HYPERLIPNAS | Weighted mode | 9 | 0.27622254 | 0.03367315 | 3.64E-05 | 0.21022317 | 0.34222191 | 1.31814117 | 1.23395341 | 1.40807272 |
| Musculoskeletal_age_gap | RX_STATIN | Weighted median | 87 | -0.056889 | 0.01225855 | 3.47E-06 | -0.0809158 | -0.0328623 | 0.94469888 | 0.92227133 | 0.96767182 |
| Musculoskeletal_age_gap | RX_STATIN | Weighted mode | 87 | -0.0505694 | 0.01021008 | 3.61E-06 | -0.0705811 | -0.0305576 | 0.95068799 | 0.93185215 | 0.96990457 |
| Musculoskeletal_age_gap | RX_CROHN_1STLINE | Inverse variance weighted | 22 | 0.098031 | 0.02592611 | 0.00015609 | 0.04721582 | 0.14884618 | 1.10299698 | 1.04834824 | 1.16049447 |
| Musculoskeletal_age_gap | E4_METABOLIA | Weighted median | 19 | -0.0980402 | 0.02252416 | 1.34E-05 | -0.1421876 | -0.0538929 | 0.90661244 | 0.86745852 | 0.94753362 |
| Musculoskeletal_age_gap | E4_METABOLIA | Weighted mode | 19 | -0.0934632 | 0.02005362 | 0.00019452 | -0.1327683 | -0.0541581 | 0.91077151 | 0.87566794 | 0.9472823 |
| Musculoskeletal_age_gap | E4_LIPOPROT | Weighted median | 25 | -0.0635343 | 0.01496802 | 2.19E-05 | -0.0928716 | -0.034197 | 0.93844196 | 0.91131051 | 0.96638115 |
| Musculoskeletal_age_gap | E4_LIPOPROT | Weighted mode | 25 | -0.0686986 | 0.01249969 | 1.19E-05 | -0.093198 | -0.0441992 | 0.93360802 | 0.9110131 | 0.95676334 |
| Musculoskeletal_age_gap | E4_HYPERCHOL | Weighted median | 21 | -0.0619366 | 0.014366 | 1.62E-05 | -0.0900939 | -0.0337792 | 0.93994251 | 0.91384534 | 0.96678495 |
| Musculoskeletal_age_gap | E4_HYPERCHOL | Weighted mode | 21 | -0.064263 | 0.01379557 | 0.00015141 | -0.0913023 | -0.0372236 | 0.93775837 | 0.91274177 | 0.96346063 |
| Musculoskeletal_age_gap | E4_OBESITYCAL | Weighted median | 10 | -0.0732663 | 0.02013262 | 0.0002735 | -0.1127263 | -0.0338064 | 0.92935327 | 0.89339517 | 0.96675863 |
| Musculoskeletal_age_gap | M13_RHEUMA | Weighted median | 18 | 0.04228592 | 0.00831984 | 3.72E-07 | 0.02597904 | 0.05859281 | 1.04319271 | 1.02631943 | 1.06034339 |
| Musculoskeletal_age_gap | M13_RHEUMA | Inverse variance weighted | 18 | 0.03276642 | 0.00769694 | 2.07E-05 | 0.01768042 | 0.04785243 | 1.03330915 | 1.01783764 | 1.04901584 |
| Musculoskeletal_age_gap | M13_RHEUMA | Weighted mode | 18 | 0.04286987 | 0.00932596 | 0.00025679 | 0.02459098 | 0.06114876 | 1.04380206 | 1.02489583 | 1.06305704 |
| Musculoskeletal_age_gap | E4_HYPERLIPNAS | Weighted median | 9 | -0.0574794 | 0.01417412 | 5.01E-05 | -0.0852607 | -0.0296982 | 0.9441413 | 0.91827284 | 0.9707385 |
| Musculoskeletal_age_gap | RHEUMA_SEROPOS_WIDE | Weighted median | 18 | 0.03940785 | 0.00776801 | 3.91E-07 | 0.02418256 | 0.05463315 | 1.04019464 | 1.02447733 | 1.05615309 |
| Musculoskeletal_age_gap | RHEUMA_SEROPOS_WIDE | Weighted mode | 18 | 0.04105778 | 0.00768197 | 5.36E-05 | 0.02600112 | 0.05611444 | 1.0419123 | 1.0263421 | 1.05771872 |
| Musculoskeletal_age_gap | RHEUMA_SEROPOS_OTH | Weighted median | 20 | 0.0363552 | 0.00758702 | 1.65E-06 | 0.02148463 | 0.05122577 | 1.03702413 | 1.02171709 | 1.0525605 |
| Musculoskeletal_age_gap | RHEUMA_SEROPOS_OTH | Weighted mode | 20 | 0.03801391 | 0.00841612 | 0.00023602 | 0.02151831 | 0.05450951 | 1.03874568 | 1.02175149 | 1.05602252 |
| Pulmonary_age_gap | M13_DORSOPATHYOTH | Inverse variance weighted | 16 | 0.15696278 | 0.0430158 | 0.00026331 | 0.07265182 | 0.24127374 | 1.16995207 | 1.07535605 | 1.27286943 |
| Pulmonary_age_gap | RX_CROHN_1STLINE | Inverse variance weighted | 22 | 0.13596369 | 0.02527293 | 7.46E-08 | 0.08642875 | 0.18549863 | 1.1456403 | 1.09027369 | 1.20381855 |
| Pulmonary_age_gap | J10_LOWCHRON | Weighted median | 23 | 0.14722124 | 0.02729573 | 6.91E-08 | 0.0937216 | 0.20072087 | 1.15861026 | 1.09825395 | 1.22228355 |

| | | | | | | | | | | |
|---|---|---|---|---|---|---|---|---|---|---|
| Pulmonary_age _gap | J10_LOWCH RON | Inverse variance weighted | 23 | 0.1576 7524 | 0.034 18421 | 3.98 E-06 | 0.090 67418 | 0.224 6763 | 1.17 0785 91 | 1.094 91221 | 1.25191 741 |
| Pulmonary_age _gap | M13_DORSA LGIA | Weighted median | 11 | 0.2380 3575 | 0.049 62013 | 1.61 E-06 | 0.140 78029 | 0.335 29121 | 1.26 8754 55 | 1.151 17169 | 1.39834 754 |
| Pulmonary_age _gap | M13_DORSA LGIA | Inverse variance weighted | 11 | 0.1981 2544 | 0.050 10987 | 7.69 E-05 | 0.099 9101 | 0.296 34078 | 1.21 9115 31 | 1.105 07156 | 1.34492 84 |
| Pulmonary_age _gap | J10_ASTHM A_EXMORE | Weighted median | 28 | 0.1169 2026 | 0.021 43332 | 4.90 E-08 | 0.074 91095 | 0.158 92957 | 1.12 4029 8 | 1.077 78817 | 1.17225 538 |
| Pulmonary_age _gap | J10_ASTHM A_EXMORE | Inverse variance weighted | 28 | 0.1480 1273 | 0.031 04737 | 1.87 E-06 | 0.087 15988 | 0.208 86558 | 1.15 9527 66 | 1.091 07111 | 1.23227 935 |
| Pulmonary_age _gap | J10_ASTHM A_EXMORE | Weighted mode | 28 | 0.1118 7721 | 0.023 77034 | 6.70 E-05 | 0.065 28736 | 0.158 46707 | 1.11 8375 53 | 1.067 46572 | 1.17171 334 |
| Pulmonary_age _gap | J10_ASTHM ACOPDKEL A | Weighted median | 34 | 0.1117 3176 | 0.018 77349 | 2.66 E-09 | 0.074 93572 | 0.148 5278 | 1.11 8212 87 | 1.077 81487 | 1.16012 505 |
| Pulmonary_age _gap | J10_ASTHM ACOPDKEL A | Inverse variance weighted | 34 | 0.1342 6464 | 0.024 31755 | 3.36 E-08 | 0.086 60225 | 0.181 92703 | 1.14 3695 45 | 1.090 46286 | 1.19952 667 |
| Pulmonary_age _gap | J10_ASTHM ACOPDKEL A | Weighted mode | 34 | 0.1096 0583 | 0.021 25323 | 1.16 E-05 | 0.067 94951 | 0.151 26216 | 1.11 5838 16 | 1.070 31127 | 1.16330 159 |
| Pulmonary_age _gap | M13_SPOND YLOPATHY | Weighted median | 8 | 0.0924 9218 | 0.024 57565 | 0.00 0167 5 | 0.044 32391 | 0.140 66046 | 1.09 6904 57 | 1.045 32089 | 1.15103 376 |
| Pulmonary_age _gap | J10_ASTHM A_MAIN_EX MORE | Weighted median | 23 | 0.1206 2421 | 0.019 74514 | 1.00 E-09 | 0.081 92374 | 0.159 32468 | 1.12 8200 87 | 1.085 37303 | 1.17271 865 |
| Pulmonary_age _gap | J10_ASTHM A_MAIN_EX MORE | Inverse variance weighted | 23 | 0.1286 1423 | 0.018 08628 | 1.15 E-12 | 0.093 16513 | 0.164 06333 | 1.13 7251 32 | 1.097 64297 | 1.17828 893 |
| Pulmonary_age _gap | J10_ASTHM A_MAIN_EX MORE | Weighted mode | 23 | 0.1197 1131 | 0.027 14879 | 0.00 0221 9 | 0.066 49967 | 0.172 92295 | 1.12 7171 4 | 1.068 76061 | 1.18877 451 |
| Pulmonary_age _gap | M13_POLYA RTHROPAT HIES | Weighted median | 15 | 0.0765 4978 | 0.016 86553 | 5.66 E-06 | 0.043 49335 | 0.109 60622 | 1.07 9555 93 | 1.044 45305 | 1.11583 859 |
| Pulmonary_age _gap | M13_POLYA RTHROPAT HIES | Inverse variance weighted | 15 | 0.0560 9206 | 0.015 40888 | 0.00 0272 38 | 0.025 89065 | 0.086 29347 | 1.05 7695 05 | 1.026 22872 | 1.09012 62 |
| Pulmonary_age _gap | H7_CONJUN CTIVITIS | Inverse variance weighted | 8 | 0.1058 6742 | 0.028 5819 | 0.00 0212 22 | 0.049 84688 | 0.161 88795 | 1.11 1674 48 | 1.051 11014 | 1.17572 849 |
| Pulmonary_age _gap | ASTHMA_IN FECTIONS | Weighted median | 13 | 0.1101 6012 | 0.022 38153 | 8.57 E-07 | 0.066 29232 | 0.154 02791 | 1.11 6456 82 | 1.068 53903 | 1.16652 345 |
| Pulmonary_age _gap | ASTHMA_IN FECTIONS | Inverse variance weighted | 13 | 0.1011 8861 | 0.019 48838 | 2.08 E-07 | 0.062 99139 | 0.139 38583 | 1.10 6485 32 | 1.065 01767 | 1.14956 755 |
| Pulmonary_age _gap | K11_REIMB_ 202 | Weighted median | 20 | 0.0596 8709 | 0.012 61779 | 2.24 E-06 | 0.034 95622 | 0.084 41795 | 1.06 1504 34 | 1.035 57438 | 1.08808 356 |
| Pulmonary_age _gap | RX_CROHN_ 2NDLINE | Weighted median | 19 | 0.0622 5319 | 0.015 48879 | 5.84 E-05 | 0.031 89517 | 0.092 61122 | 1.06 4231 77 | 1.032 40927 | 1.09703 514 |
| Pulmonary_age _gap | ASTHMA_A CUTE_RESPI RATORY_IN FECTIONS | Weighted median | 9 | 0.1060 3116 | 0.023 83111 | 8.62 E-06 | 0.059 32219 | 0.152 74013 | 1.11 1856 52 | 1.061 11707 | 1.16502 218 |
| Pulmonary_age _gap | ASTHMA_A CUTE_RESPI RATORY_IN FECTIONS | Inverse variance weighted | 9 | 0.1280 6924 | 0.022 6851 | 1.65 E-08 | 0.083 60645 | 0.172 53203 | 1.13 6631 7 | 1.087 20094 | 1.18830 988 |

| | | | | | | | | | | | |
|---|---|---|---|---|---|---|---|---|---|---|---|
| Pulmonary_age_gap | ALLERG_ASTHMA | Weighted median | 14 | 0.07971783 | 0.01686231 | 2.27E-06 | 0.04666769 | 0.11276796 | 1.08298143 | 1.04777377 | 1.11937216 |
| Pulmonary_age_gap | ALLERG_ASTHMA | Inverse variance weighted | 14 | 0.09248476 | 0.01555921 | 2.78E-09 | 0.06198871 | 0.1229808 | 1.09689642 | 1.06395034 | 1.13086271 |
| Pulmonary_age_gap | ASTHMA_CHILD_EXMORE | Inverse variance weighted | 8 | 0.0606323 | 0.0166522 | 0.00027148 | 0.02799399 | 0.09327062 | 1.06250816 | 1.0283895 | 1.09775877 |
| Renal_age_gap | E4_OBESITY | Weighted median | 19 | 0.10713625 | 0.02230394 | 1.56E-06 | 0.06342053 | 0.15085198 | 1.11308591 | 1.06547481 | 1.16282452 |
| Renal_age_gap | E4_OBESITY | Inverse variance weighted | 19 | 0.10459366 | 0.01868695 | 2.18E-08 | 0.06796724 | 0.14122007 | 1.11025937 | 1.07033024 | 1.15167807 |
| Renal_age_gap | ABDOM_HERNIA | Weighted median | 11 | 0.08978221 | 0.01921997 | 2.99E-06 | 0.05211106 | 0.12745335 | 1.09393601 | 1.05349274 | 1.13593188 |
| Renal_age_gap | ABDOM_HERNIA | Inverse variance weighted | 11 | 0.07995083 | 0.01881688 | 2.15E-05 | 0.04306974 | 0.11683191 | 1.0832338 | 1.04401071 | 1.1239305 |
| Renal_age_gap | H7_MACULADEGEN | Weighted median | 11 | 0.03712115 | 0.00989249 | 0.00017511 | 0.01773187 | 0.05651042 | 1.03781874 | 1.01789001 | 1.05813764 |
| Renal_age_gap | E4_OBESITYCAL | Weighted median | 10 | 0.0875241 | 0.0193079 | 5.81E-06 | 0.04968062 | 0.12536758 | 1.09146857 | 1.0509354 | 1.13356505 |
| Renal_age_gap | E4_OBESITYCAL | Inverse variance weighted | 10 | 0.1116372 | 0.01441947 | 9.78E-15 | 0.08337504 | 0.13989936 | 1.11810714 | 1.08694938 | 1.15015804 |
| Renal_age_gap | G6_AD_WIDE | Weighted median | 8 | -0.0373801 | 0.00892243 | 2.80E-05 | -0.054868 | -0.0198921 | 0.96330995 | 0.94661007 | 0.98030445 |
| Renal_age_gap | G6_AD_WIDE | Inverse variance weighted | 8 | -0.0363863 | 0.00896284 | 4.91E-05 | -0.0539535 | -0.0188191 | 0.96426773 | 0.9474762 | 0.98135684 |
| Renal_age_gap | E4_NONTOXIC_THYROID | Weighted median | 31 | 0.03593613 | 0.0087052 | 3.66E-05 | 0.01887394 | 0.05299832 | 1.03658964 | 1.01905318 | 1.05442788 |
| Renal_age_gap | I9_CAVS_OPERATED | Weighted median | 11 | -0.0712269 | 0.01803861 | 7.86E-05 | -0.1065826 | -0.0358713 | 0.93125053 | 0.89890078 | 0.96476449 |
| Renal_age_gap | THYROTOXICOSIS | Weighted median | 23 | 0.04751959 | 0.01032698 | 4.19E-06 | 0.0272787 | 0.06776048 | 1.04866674 | 1.02765417 | 1.07010896 |
| Renal_age_gap | THYROTOXICOSIS | Inverse variance weighted | 23 | 0.04339125 | 0.01155343 | 0.00017286 | 0.02074653 | 0.06603596 | 1.04434641 | 1.02096323 | 1.06826514 |
| Renal_age_gap | THYROTOXICOSIS | Weighted mode | 23 | 0.05748325 | 0.01052327 | 1.73E-05 | 0.03685765 | 0.07810886 | 1.05916753 | 1.03754531 | 1.08124035 |
| Renal_age_gap | DRY_AMD | Weighted median | 14 | 0.02331544 | 0.00580917 | 5.98E-05 | 0.01192947 | 0.03470141 | 1.02358937 | 1.01200091 | 1.03531053 |
| Renal_age_gap | DRY_AMD | Inverse variance weighted | 14 | 0.0230247 | 0.00611007 | 0.00016435 | 0.01104897 | 0.03500043 | 1.02329181 | 1.01111023 | 1.03562016 |
| RX_ANTIHYP | Cardiovascular_age_gap | Weighted median | 37 | 0.27037705 | 0.06055265 | 8.00E-06 | 0.15169386 | 0.38906025 | 1.31045847 | 1.16380389 | 1.47559345 |
| RX_ANTIHYP | Cardiovascular_age_gap | Inverse variance weighted | 37 | 0.51639968 | 0.10782887 | 1.68E-06 | 0.30505509 | 0.72774426 | 1.67598269 | 1.35669974 | 2.07040505 |
| RX_STATIN | Cardiovascular_age_gap | Weighted median | 37 | 0.30178377 | 0.06118526 | 8.13E-07 | 0.18186066 | 0.42170688 | 1.3522688 | 1.19944705 | 1.52456159 |
| RX_STATIN | Cardiovascular_age_gap | Inverse variance weighted | 37 | 0.29623579 | 0.05791595 | 3.14E-07 | 0.18272053 | 0.40975106 | 1.34478721 | 1.20047886 | 1.50644272 |

| | | | | | | | | | | |
|---|---|---|---|---|---|---|---|---|---|---|
| I9_HYPTENS | Cardiovascular_age_gap | Inverse variance weighted | 37 | 0.54772629 | 0.11707698 | 2.89E-06 | 0.31825541 | 0.77719716 | 1.72931657 | 1.37472733 | 2.17536652 |
| I9_HYPTENSESS | Cardiovascular_age_gap | Inverse variance weighted | 37 | 0.55049816 | 0.11228863 | 9.46E-07 | 0.33041244 | 0.77058388 | 1.73411667 | 1.39154194 | 2.16102767 |
| I9_IHD | Cardiovascular_age_gap | Weighted median | 37 | 0.35806374 | 0.07300346 | 9.35E-07 | 0.21497696 | 0.50115053 | 1.4305568 | 1.23983333 | 1.65061926 |
| I9_IHD | Cardiovascular_age_gap | Inverse variance weighted | 37 | 0.49500709 | 0.09322373 | 1.10E-07 | 0.31228857 | 0.6777256 | 1.64050986 | 1.36654899 | 1.96939344 |
| I9_CVD_HARD | Cardiovascular_age_gap | Weighted median | 37 | 0.35093024 | 0.07166713 | 9.75E-07 | 0.21046266 | 0.49139781 | 1.42038823 | 1.23424897 | 1.63459949 |
| I9_CVD_HARD | Cardiovascular_age_gap | Inverse variance weighted | 37 | 0.38898349 | 0.08524462 | 5.04E-06 | 0.22190405 | 0.55606294 | 1.47548019 | 1.24845158 | 1.74379354 |
| E4_METABOLIA | Cardiovascular_age_gap | Weighted median | 37 | 0.28054232 | 0.0643336 | 1.30E-05 | 0.15444846 | 0.40663617 | 1.32384756 | 1.16701413 | 1.50175763 |
| E4_METABOLIA | Cardiovascular_age_gap | Inverse variance weighted | 37 | 0.23161102 | 0.05719319 | 5.13E-05 | 0.11951237 | 0.34370966 | 1.26062927 | 1.12694719 | 1.41016915 |
| I9_CORATHER | Cardiovascular_age_gap | Weighted median | 37 | 0.4467068 | 0.08494414 | 1.45E-07 | 0.28021628 | 0.61319731 | 1.56315591 | 1.32341601 | 1.84632525 |
| I9_CORATHER | Cardiovascular_age_gap | Inverse variance weighted | 37 | 0.55912952 | 0.11201781 | 5.99E-07 | 0.33957461 | 0.77868443 | 1.74914923 | 1.40435006 | 2.17860426 |
| I9_CHD | Cardiovascular_age_gap | Weighted median | 37 | 0.48939295 | 0.08034507 | 1.12E-09 | 0.33191662 | 0.64686928 | 1.63132562 | 1.39363664 | 1.90955319 |
| I9_CHD | Cardiovascular_age_gap | Inverse variance weighted | 37 | 0.49872723 | 0.11052753 | 6.41E-06 | 0.28209327 | 0.7153612 | 1.64662417 | 1.32590238 | 2.04492517 |
| E4_LIPOPROT | Cardiovascular_age_gap | Weighted median | 37 | 0.28552327 | 0.07141191 | 6.38E-05 | 0.14555594 | 0.42549061 | 1.33045804 | 1.15668244 | 1.53034104 |
| E4_LIPOPROT | Cardiovascular_age_gap | Inverse variance weighted | 37 | 0.31721197 | 0.06874205 | 3.94E-06 | 0.18247756 | 0.45194638 | 1.37329364 | 1.20018721 | 1.57136769 |
| I9_ANGINA | Cardiovascular_age_gap | Weighted median | 37 | 0.50005675 | 0.09265056 | 6.77E-08 | 0.31846165 | 0.68165184 | 1.64881483 | 1.37501089 | 1.97714096 |
| I9_ANGINA | Cardiovascular_age_gap | Inverse variance weighted | 37 | 0.58646241 | 0.12070128 | 1.18E-06 | 0.3498879 | 0.82303692 | 1.79761792 | 1.41890849 | 2.27740564 |
| E4_HYPERCHOL | Cardiovascular_age_gap | Inverse variance weighted | 37 | 0.31987969 | 0.07404539 | 1.56E-05 | 0.17475073 | 0.46500865 | 1.37696209 | 1.19094931 | 1.59202795 |
| I9_HEARTFAIL_AND_ANTIHYPERT | Cardiovascular_age_gap | Weighted median | 37 | 0.52027381 | 0.11782049 | 1.01E-05 | 0.28934565 | 0.75120198 | 1.68248827 | 1.33555328 | 2.11954614 |
| I9_HEARTFAIL_AND_ANTIHYPERT | Cardiovascular_age_gap | Inverse variance weighted | 37 | 0.67734638 | 0.13227482 | 3.04E-07 | 0.41808774 | 0.93660502 | 1.96864675 | 1.51905395 | 2.55130507 |
| I9_MI_STRICT | Cardiovascular_age_gap | Weighted median | 37 | 0.41114314 | 0.10593658 | 0.00010401 | 0.20350745 | 0.61877883 | 1.50854127 | 1.22569428 | 1.85665936 |
| I9_MI_STRICT | Cardiovascular_age_gap | Inverse variance weighted | 37 | 0.53653289 | 0.12363983 | 1.43E-05 | 0.29419882 | 0.77886696 | 1.71006758 | 1.34205071 | 2.17900197 |
| I9_REVASC | Cardiovascular_age_gap | Weighted median | 37 | 0.59516064 | 0.11004549 | 6.36E-08 | 0.37947148 | 0.81084979 | 1.81332221 | 1.46151195 | 2.24981905 |
| I9_REVASC | Cardiovascular_age_gap | Inverse variance weighted | 37 | 0.75939821 | 0.16354994 | 3.43E-06 | 0.43884032 | 1.0799561 | 2.13698981 | 1.55090762 | 2.94455027 |

| | | | | | | | | | | | |
|---|---|---|---|---|---|---|---|---|---|---|---|
| I9_ATHSCLE | Cardiovascula r_age_gap | Inverse variance weighted | 37 | 0.5354 1724 | 0.109 9535 | 1.12 E-06 | 0.319 90837 | 0.750 92611 | 1.70 8160 81 | 1.377 00159 | 2.11896 15 |
| I9_ANGIO | Cardiovascula r_age_gap | Weighted median | 37 | 0.6477 5155 | 0.134 06812 | 1.36 E-06 | 0.384 97804 | 0.910 52506 | 1.91 1238 67 | 1.469 58204 | 2.48562 73 |
| I9_ANGIO | Cardiovascula r_age_gap | Inverse variance weighted | 37 | 0.6919 4446 | 0.149 5554 | 3.72 E-06 | 0.398 81588 | 0.985 07305 | 1.99 7596 01 | 1.490 05924 | 2.67800 751 |
| I9_UAP | Cardiovascula r_age_gap | Weighted median | 37 | 0.5706 1707 | 0.116 59606 | 9.88 E-07 | 0.342 0888 | 0.799 14534 | 1.76 9358 53 | 1.407 88531 | 2.22363 967 |
| I9_UAP | Cardiovascula r_age_gap | Inverse variance weighted | 37 | 0.6073 2893 | 0.137 22557 | 9.61 E-06 | 0.338 36681 | 0.876 29105 | 1.83 5522 04 | 1.402 65492 | 2.40197 436 |
| I9_HEARTFAI L_AND_CHD | Cardiovascula r_age_gap | Inverse variance weighted | 37 | 0.5080 9517 | 0.108 55183 | 2.86 E-06 | 0.295 33359 | 0.720 85676 | 1.66 2122 12 | 1.343 57449 | 2.05619 412 |
| E4_HYPERLIP NAS | Cardiovascula r_age_gap | Weighted median | 37 | 0.4819 3362 | 0.114 13881 | 2.42 E-05 | 0.258 22156 | 0.705 64568 | 1.61 9202 3 | 1.294 62563 | 2.02515 387 |
| I9_CABG | Cardiovascula r_age_gap | Weighted median | 37 | 0.7437 6351 | 0.144 24985 | 2.52 E-07 | 0.461 0338 | 1.026 49323 | 2.10 3838 46 | 1.585 71245 | 2.79126 034 |
| I9_CABG | Cardiovascula r_age_gap | Inverse variance weighted | 37 | 0.8725 9143 | 0.206 06633 | 2.29 E-05 | 0.468 70142 | 1.276 48143 | 2.39 3104 38 | 1.597 91783 | 3.58400 693 |
| O15_PREEC_ OR_FETGRO | Cardiovascula r_age_gap | Inverse variance weighted | 37 | 0.4735 1249 | 0.118 77423 | 6.70 E-05 | 0.240 715 | 0.706 30998 | 1.60 5624 04 | 1.272 15842 | 2.02649 962 |
| RX_STATIN | Metabolic_age _gap | Weighted median | 67 | 0.3437 4014 | 0.046 9019 | 2.32 E-13 | 0.251 81241 | 0.435 66786 | 1.41 0212 12 | 1.286 35471 | 1.54599 522 |
| RX_STATIN | Metabolic_age _gap | Inverse variance weighted | 67 | 0.6298 2143 | 0.133 46238 | 2.37 E-06 | 0.368 23517 | 0.891 4077 | 1.87 7275 33 | 1.445 18186 | 2.43855 999 |
| RX_STATIN | Metabolic_age _gap | Simple mode | 67 | 0.3945 474 | 0.071 83228 | 6.82 E-07 | 0.253 75614 | 0.535 33867 | 1.48 3712 51 | 1.288 85746 | 1.70802 66 |
| RX_STATIN | Metabolic_age _gap | Weighted mode | 67 | 0.3103 5246 | 0.041 05402 | 1.63 E-10 | 0.229 88657 | 0.390 81834 | 1.36 3905 75 | 1.258 45726 | 1.47818 996 |
| E4_METABOL IA | Metabolic_age _gap | Weighted median | 67 | 0.1715 4227 | 0.042 34132 | 5.09 E-05 | 0.088 55329 | 0.254 53125 | 1.18 7134 32 | 1.092 59247 | 1.28985 686 |
| E4_LIPOPROT | Metabolic_age _gap | Weighted median | 67 | 0.2428 2971 | 0.052 66475 | 4.01 E-06 | 0.139 60681 | 0.346 05262 | 1.27 4851 51 | 1.149 82161 | 1.41347 698 |
| E4_LIPOPROT | Metabolic_age _gap | Inverse variance weighted | 67 | 0.4184 7098 | 0.103 11775 | 4.95 E-05 | 0.216 36019 | 0.620 58178 | 1.51 9636 23 | 1.241 54949 | 1.86000 985 |
| E4_HYPERCH OL | Metabolic_age _gap | Weighted median | 67 | 0.3310 0218 | 0.059 07493 | 2.11 E-08 | 0.215 21531 | 0.446 78905 | 1.39 2362 82 | 1.240 12888 | 1.56328 449 |
| E4_HYPERCH OL | Metabolic_age _gap | Inverse variance weighted | 67 | 0.4603 8267 | 0.110 53358 | 3.11 E-05 | 0.243 73685 | 0.677 02848 | 1.58 4680 28 | 1.276 00851 | 1.96802 103 |
| E4_HYPERCH OL | Metabolic_age _gap | Weighted mode | 67 | 0.3362 8527 | 0.072 36639 | 1.66 E-05 | 0.194 44714 | 0.478 1234 | 1.39 9738 28 | 1.214 63928 | 1.61304 453 |
| RX_GLUCOS AMINE | Metabolic_age _gap | Inverse variance weighted | 67 | - 0.1718 615 | 0.042 5704 | 5.41 E-05 | - 0.255 2995 | - 0.088 4235 | 0.84 2095 78 | 0.774 68445 | 0.91537 31 |
| E4_HYPERLIP NAS | Metabolic_age _gap | Inverse variance weighted | 67 | 0.3918 8618 | 0.100 65971 | 9.89 E-05 | 0.194 59314 | 0.589 17922 | 1.47 9769 27 | 1.214 81662 | 1.80250 834 |
| RX_CROHN_1 STLINE | Pulmonary_ag e_gap | Weighted median | 58 | 0.1659 9574 | 0.042 83555 | 0.00 0106 54 | 0.082 03805 | 0.249 95343 | 1.18 0568 07 | 1.085 49712 | 1.28396 562 |

| | | | | | | | | | | | |
|---|---|---|---|---|---|---|---|---|---|---|---|
| RX_CROHN_1STLINE | Pulmonary_age_gap | Inverse variance weighted | 58 | 0.14769433 | 0.0364929 | 5.18E-05 | 0.07616825 | 0.2192204 | 1.15915852 | 1.07914413 | 1.24510567 |
| J10_LOWCHRON | Pulmonary_age_gap | Weighted median | 58 | 0.47200725 | 0.05096977 | 2.03E-20 | 0.3721065 | 0.571908 | 1.60320901 | 1.45078749 | 1.77164413 |
| J10_LOWCHRON | Pulmonary_age_gap | Inverse variance weighted | 58 | 0.46635704 | 0.04432233 | 6.84E-26 | 0.37948528 | 0.5532288 | 1.59417609 | 1.46153212 | 1.73885839 |
| J10_LOWCHRON | Pulmonary_age_gap | Weighted mode | 58 | 0.48283022 | 0.0997834 | 1.03E-05 | 0.28725475 | 0.67840569 | 1.62065473 | 1.33276369 | 1.97073327 |
| J10_ASTHMA_EXMORE | Pulmonary_age_gap | Weighted median | 58 | 0.46877711 | 0.06514315 | 6.20E-13 | 0.34109653 | 0.59645769 | 1.59803877 | 1.40648901 | 1.81567571 |
| J10_ASTHMA_EXMORE | Pulmonary_age_gap | Inverse variance weighted | 58 | 0.50172536 | 0.0545502 | 3.66E-20 | 0.39480697 | 0.60864374 | 1.65156836 | 1.48409769 | 1.83793699 |
| J10_ASTHMACOPDKELA | Pulmonary_age_gap | Weighted median | 58 | 0.55577155 | 0.06338215 | 1.81E-18 | 0.43154254 | 0.68000056 | 1.74328549 | 1.53963063 | 1.97387883 |
| J10_ASTHMACOPDKELA | Pulmonary_age_gap | Inverse variance weighted | 58 | 0.6009413 | 0.06089108 | 5.67E-23 | 0.48159478 | 0.72028783 | 1.82383478 | 1.61865375 | 2.05502462 |
| J10_ASTHMACOPDKELA | Pulmonary_age_gap | Weighted mode | 58 | 0.52463286 | 0.11067058 | 1.46E-05 | 0.30771852 | 0.7415472 | 1.68983833 | 1.36031804 | 2.09918086 |
| J10_ASTHMA_MAIN_EXMORE | Pulmonary_age_gap | Weighted median | 58 | 0.47819315 | 0.07188467 | 2.89E-11 | 0.33729919 | 0.6190871 | 1.61315703 | 1.40115822 | 1.85723181 |
| J10_ASTHMA_MAIN_EXMORE | Pulmonary_age_gap | Inverse variance weighted | 58 | 0.52173876 | 0.06102087 | 1.23E-17 | 0.40213786 | 0.64133967 | 1.68495484 | 1.49501742 | 1.89902324 |
| ASTHMA_INFECTIONS | Pulmonary_age_gap | Weighted median | 58 | 0.51681667 | 0.0732881 | 1.77E-12 | 0.373172 | 0.66046134 | 1.67668171 | 1.45233412 | 1.93568514 |
| ASTHMA_INFECTIONS | Pulmonary_age_gap | Inverse variance weighted | 58 | 0.51565336 | 0.0577494 | 4.29E-19 | 0.40246454 | 0.62884217 | 1.67473234 | 1.49550589 | 1.87543789 |
| ASTHMA_NAS | Pulmonary_age_gap | Weighted median | 58 | 0.53774838 | 0.07661069 | 2.23E-12 | 0.38759143 | 0.68790533 | 1.71214742 | 1.47342766 | 1.98954373 |
| ASTHMA_NAS | Pulmonary_age_gap | Inverse variance weighted | 58 | 0.51651579 | 0.06201958 | 8.20E-17 | 0.39495741 | 0.63807416 | 1.6761773 | 1.48432098 | 1.89283207 |
| J10_COPD | Pulmonary_age_gap | Weighted median | 58 | 0.66896132 | 0.0838726 | 1.51E-15 | 0.50457102 | 0.83335163 | 1.95220856 | 1.65627486 | 2.30101799 |
| J10_COPD | Pulmonary_age_gap | Inverse variance weighted | 58 | 0.58254639 | 0.06439772 | 1.48E-19 | 0.45632686 | 0.70876592 | 1.79059217 | 1.57826613 | 2.0314827 |
| J10_COPD | Pulmonary_age_gap | Weighted mode | 58 | 0.76734711 | 0.17830566 | 6.68E-05 | 0.41786801 | 1.11682621 | 2.15404423 | 1.51872021 | 3.05514243 |
| ASTHMA_ACUTE_RESPIRATORY_INFECTIONS | Pulmonary_age_gap | Weighted median | 58 | 0.50155859 | 0.08866859 | 1.54E-08 | 0.32776814 | 0.67534903 | 1.65129295 | 1.38786715 | 1.9647186 |
| ASTHMA_ACUTE_RESPIRATORY_INFECTIONS | Pulmonary_age_gap | Inverse variance weighted | 58 | 0.52242447 | 0.0698334 | 7.38E-14 | 0.385551 | 0.65929794 | 1.68611062 | 1.47042431 | 1.93343446 |
| ASTHMA_PNEUMONIA | Pulmonary_age_gap | Weighted median | 58 | 0.60861962 | 0.09393832 | 9.24E-11 | 0.42450052 | 0.79273873 | 1.83789267 | 1.52882662 | 2.2094392 |
| ASTHMA_PNEUMONIA | Pulmonary_age_gap | Inverse variance weighted | 58 | 0.51908819 | 0.06810893 | 2.51E-14 | 0.38559469 | 0.6525817 | 1.68049466 | 1.47048854 | 1.92049257 |

| | | | | | | | | | | | |
|---|---|---|---|---|---|---|---|---|---|---|---|
| COPD_LATER | Pulmonary_age_gap | Weighted median | 58 | 0.53749427 | 0.10623879 | 4.21E-07 | 0.32926624 | 0.74572231 | 1.7117124 | 1.38994786 | 2.10796348 |
| COPD_LATER | Pulmonary_age_gap | Inverse variance weighted | 58 | 0.44395253 | 0.07831344 | 1.44E-08 | 0.29045818 | 0.59744688 | 1.55885649 | 1.33703995 | 1.81747264 |
| ASTHMA_OBESITY | Pulmonary_age_gap | Inverse variance weighted | 58 | 0.46726463 | 0.07712933 | 1.38E-09 | 0.31609114 | 0.61843812 | 1.5956236 | 1.37175527 | 1.85602689 |
| ALLERG_ASTHMA | Pulmonary_age_gap | Inverse variance weighted | 58 | 0.45588365 | 0.08582444 | 1.09E-07 | 0.28766775 | 0.62409955 | 1.57756678 | 1.33331424 | 1.86656445 |
| NONALLERG_ASTHMA_EXMORE | Pulmonary_age_gap | Inverse variance weighted | 58 | 0.55333852 | 0.0913676 | 1.39E-09 | 0.37425802 | 0.73241902 | 1.73904919 | 1.45391225 | 2.08010634 |
| COPD_EARLY | Pulmonary_age_gap | Weighted median | 58 | 0.69466083 | 0.12971882 | 8.55E-08 | 0.44041194 | 0.94890972 | 2.00302959 | 1.55334697 | 2.58289205 |
| COPD_EARLY | Pulmonary_age_gap | Inverse variance weighted | 58 | 0.72082126 | 0.10135554 | 1.15E-12 | 0.52216441 | 0.91947811 | 2.05612112 | 1.68567218 | 2.50798115 |
| Brain | Hepatic_age_gap | Inverse variance weighted | 41 | 0.11606646 | 0.03968122 | 0.00344487 | 0.03829126 | 0.19384166 | 1.12307051 | 1.03903382 | 1.21390406 |
| Endocrine | Hepatic_age_gap | MR Egger | 41 | -0.3439261 | 0.1100147 | 0.00333954 | -0.5595549 | -0.1282973 | 0.70898134 | 0.57146338 | 0.87959187 |
| Reproductive_female | Metabolic_age_gap | Inverse variance weighted | 71 | 0.09574456 | 0.03207188 | 0.0028329 | 0.03288367 | 0.15860544 | 1.10047792 | 1.03343031 | 1.17187549 |
| Eye | Metabolic_age_gap | Weighted median | 71 | 0.14879974 | 0.04871247 | 0.0022532 | 0.05332331 | 0.24427618 | 1.16044058 | 1.05477061 | 1.27669688 |
| Eye | Metabolic_age_gap | Inverse variance weighted | 71 | 0.12444807 | 0.03822723 | 0.001132 | 0.04952269 | 0.19937345 | 1.13252321 | 1.05076944 | 1.22063773 |
| Pulmonary | Pulmonary_age_gap | Inverse variance weighted | 61 | -0.1891674 | 0.05589272 | 0.00071315 | -0.2987171 | -0.0796177 | 0.82764794 | 0.74176919 | 0.92346934 |
| Pulmonary | Renal_age_gap | Inverse variance weighted | 46 | 0.15596996 | 0.0508809 | 0.00217384 | 0.0562434 | 0.25569652 | 1.1687911 | 1.05785514 | 1.29136077 |
| Eye | Renal_age_gap | Inverse variance weighted | 46 | 0.13325783 | 0.04273987 | 0.00182156 | 0.04948769 | 0.21702798 | 1.14254455 | 1.05073266 | 1.24237886 |
| Renal | Renal_age_gap | Inverse variance weighted | 46 | 0.16184203 | 0.0564029 | 0.00411262 | 0.05129235 | 0.27239171 | 1.17567451 | 1.05263059 | 1.31310126 |
| Reproductive_male | Renal_age_gap | Inverse variance weighted | 46 | 0.16045392 | 0.04684393 | 0.00061415 | 0.06863981 | 0.25226803 | 1.17404367 | 1.07105036 | 1.28694093 |
| Immune | Renal_age_gap | Inverse variance weighted | 46 | 0.18956828 | 0.06243089 | 0.00239382 | 0.06720374 | 0.31193282 | 1.20872766 | 1.06951336 | 1.36606292 |
| Skin | Renal_age_gap | Weighted median | 46 | -0.1794673 | 0.05827421 | 0.00207206 | -0.2936848 | -0.0652499 | 0.83571527 | 0.74551146 | 0.93683336 |

### eTable 8: The classification results to predict the 14 systemic disease categories

#### a) Cross-omics feature classification results

| Disease category | feature | CV_accuracy | CV_accuracy_std | CV_balanced_accuracy | CV_balanced_accuracy_std | CV_sensitivity | CV_sensitivity_std | CV_specificity | CV_specificity_std | CV_ppv | CV_ppv_std | CV_npv | CV_npv_std | CV_test_N | CV_test_N_CN | CV_test_N_PT |
|---|---|---|---|---|---|---|---|---|---|---|---|---|---|---|---|---|
| infectious_parasitic_disease_diagnosis | phenobagprs_protbagprs | 0.51280728 | 0.01724558 | 0.51310094 | 0.01697084 | 0.55180428 | 0.20330763 | 0.47439759 | 0.20778655 | 0.50169244 | 0.07692356 | 0.47461521 | 0.14498933 | 659 | 332 | 327 |
| neoplasms_diagnosis | phenobagprs_protbagprs | 0.57783636 | 0.13257735 | 0.50113888 | 0.00705805 | 0.69375813 | 0.33234596 | 0.30851964 | 0.3320631 | 0.62997641 | 0.21219868 | 0.18412889 | 0.15252221 | 1100 | 331 | 769 |
| blood_and_immune_system_diagnosis | phenobagprs_protbagprs | 0.3806359 | 0.06647289 | 0.49623163 | 0.00875332 | 0.13614907 | 0.2181398 | 0.8563142 | 0.22949702 | 0.23682554 | 0.32362174 | 0.33624384 | 0.00794824 | 975 | 331 | 644 |
| endocrine_nutritional_metabolic_disease_diagnosis | phenobagprs_protbagprs | 0.50743068 | 0.07865014 | 0.5089179 | 0.01188164 | 0.50508655 | 0.20365676 | 0.51274924 | 0.20741608 | 0.67591374 | 0.13994945 | 0.28866158 | 0.08654394 | 1082 | 331 | 751 |
| mental_behavioural_disorder_diagnosis | phenobagprs_protbagprs | 0.51982659 | 0.01512816 | 0.51548996 | 0.01643414 | 0.62266667 | 0.18887251 | 0.40831325 | 0.20194541 | 0.53518403 | 0.01618362 | 0.43088332 | 0.17663265 | 692 | 332 | 360 |
| nerve_system_diagnosis | phenobagprs_protbagprs | 0.51699605 | 0.03416655 | 0.50992115 | 0.01325877 | 0.56528037 | 0.25322162 | 0.45456193 | 0.25508707 | 0.56805342 | 0.14483488 | 0.40503155 | 0.13770368 | 759 | 331 | 428 |
| eye_diagnosis | phenobagprs_protbagprs | 0.48038251 | 0.03159756 | 0.49840143 | 0.01015854 | 0.30997506 | 0.31587632 | 0.68682779 | 0.31647222 | 0.36518331 | 0.29126671 | 0.41494772 | 0.12407004 | 732 | 331 | 401 |
| ear_diagnosis | phenobagprs_protbagprs | 0.5661435 | 0.16825033 | 0.50451173 | 0.01403992 | 0.37842105 | 0.34892047 | 0.63060241 | 0.34524366 | 0.1625629 | 0.12909971 | 0.67701397 | 0.23129516 | 446 | 332 | 114 |
| circular_system_diagnosis | phenobagprs_protbagprs | 0.50849277 | 0.11325077 | 0.5129111 | 0.01361786 | 0.50479501 | 0.20662969 | 0.52102719 | 0.20581148 | 0.70391326 | 0.23720247 | 0.23058596 | 0.05132001 | 1453 | 331 | 1122 |
| respiratory_system_diagnosis | phenobagprs_protbagprs | 0.5207617 | 0.03734847 | 0.52463587 | 0.0169418 | 0.51078231 | 0.11889809 | 0.53848943 | 0.11785964 | 0.669891 | 0.05026158 | 0.38425323 | 0.01656217 | 919 | 331 | 588 |
| digestive_system_diagnosis | phenobagprs_protbagprs | 0.55160151 | 0.19194473 | 0.50284397 | 0.00932679 | 0.58653386 | 0.32961855 | 0.41915408 | 0.33076598 | 0.69862716 | 0.26072274 | 0.15753554 | 0.09492719 | 1586 | 331 | 1255 |
| skin_system_diagnosis | phenobagprs_protbagprs | 0.49169607 | 0.04187476 | 0.49833978 | 0.00893434 | 0.43305419 | 0.39834514 | 0.56362538 | 0.39748629 | 0.37535155 | 0.25088102 | 0.33101669 | 0.19838498 | 737 | 331 | 406 |
| musculoskeletal_system_diagnosis | phenobagprs_protbagprs | 0.39675507 | 0.13017589 | 0.50395583 | 0.01079007 | 0.28229232 | 0.26637776 | 0.72561934 | 0.26243149 | 0.51976024 | 0.36315223 | 0.2558261 | 0.03782632 | 1282 | 331 | 951 |
| genitourinary_system_diagnosis | phenobagprs_protbagprs | 0.54301656 | 0.15181073 | 0.50236423 | 0.00787978 | 0.5985049 | 0.35848203 | 0.40622356 | 0.35826926 | 0.61389192 | 0.25130097 | 0.19469291 | 0.14203113 | 1147 | 331 | 816 |
| infectious_parasitic_disease_diagnosis | protbag | 0.53241275 | 0.01606799 | 0.53247909 | 0.01606404 | 0.54122324 | 0.03455704 | 0.52373494 | 0.03483319 | 0.52825666 | 0.01580871 | 0.53693699 | 0.01669327 | 659 | 332 | 327 |
| neoplasms_diagnosis | protbag | 0.57890909 | 0.03922637 | 0.52375974 | 0.01671552 | 0.66226268 | 0.09833747 | 0.3852568 | 0.10764102 | 0.71497921 | 0.01136595 | 0.32759942 | 0.05625532 | 1100 | 331 | 769 |
| blood_and_immune_system_diagnosis | protbag | 0.50750769 | 0.09130044 | 0.5132893 | 0.0205482 | 0.4952795 | 0.25757147 | 0.53129909 | 0.23678508 | 0.56887309 | 0.25587071 | 0.35745098 | 0.02468284 | 975 | 331 | 644 |
| endocrine_nutritional_metabolic_disease_diagnosis | protbag | 0.56186691 | 0.01881741 | 0.54167804 | 0.01444034 | 0.59368842 | 0.03620631 | 0.48966767 | 0.03851992 | 0.72532395 | 0.01090767 | 0.34722653 | 0.01495568 | 1082 | 331 | 751 |
| mental_behavioural_disorder_diagnosis | protbag | 0.54132948 | 0.0158409 | 0.54005489 | 0.01577553 | 0.57155556 | 0.03084822 | 0.50855422 | 0.03007348 | 0.55780232 | 0.01456679 | 0.52279771 | 0.0178784 | 692 | 332 | 360 |
| nerve_system_diagnosis | protbag | 0.54221344 | 0.02096614 | 0.53551275 | 0.01986676 | 0.58794393 | 0.04037411 | 0.48308157 | 0.03444774 | 0.59508261 | 0.01726887 | 0.47607938 | 0.02317669 | 759 | 331 | 428 |
| eye_diagnosis | protbag | 0.49797814 | 0.0343207 | 0.50155623 | 0.01843472 | 0.46413965 | 0.24874654 | 0.53897281 | 0.23559974 | 0.46635247 | 0.19228738 | 0.44765759 | 0.06783927 | 732 | 331 | 401 |
| ear_diagnosis | protbag | 0.51587444 | 0.1171396 | 0.52074244 | 0.02864428 | 0.53070175 | 0.25711678 | 0.51078313 | 0.24227276 | 0.2370097 | 0.09270762 | 0.703017 | 0.21081804 | 446 | 332 | 114 |
| circular_system_diagnosis | protbag | 0.56191328 | 0.06162903 | 0.53073894 | 0.01711344 | 0.58800357 | 0.10539843 | 0.47347432 | 0.09322607 | 0.7748224 | 0.11224356 | 0.25484594 | 0.01566864 | 1453 | 331 | 1122 |
| respiratory_system_diagnosis | protbag | 0.52646355 | 0.02172021 | 0.52901242 | 0.01542307 | 0.51989796 | 0.05420359 | 0.53812689 | 0.05208033 | 0.66674642 | 0.01418325 | 0.38797405 | 0.01974486 | 919 | 331 | 588 |
| digestive_system_diagnosis | protbag | 0.61774275 | 0.07403872 | 0.53362634 | 0.01895244 | 0.67800797 | 0.1238589 | 0.38924471 | 0.12261083 | 0.79204947 | 0.11463578 | 0.23488183 | 0.05297096 | 1586 | 331 | 1255 |
| skin_system_diagnosis | protbag | 0.50369064 | 0.02851207 | 0.50962831 | 0.01602872 | 0.45128079 | 0.18462568 | 0.56797583 | 0.17200683 | 0.50189356 | 0.1707055 | 0.45968973 | 0.01646931 | 737 | 331 | 406 |
| musculoskeletal_system_diagnosis | protbag | 0.5274415 | 0.08767969 | 0.5143995 | 0.01574641 | 0.54136698 | 0.17018019 | 0.48743202 | 0.15219206 | 0.71720466 | 0.15039716 | 0.27298914 | 0.01444185 | 1282 | 331 | 951 |
| genitourinary_system_diagnosis | protbag | 0.52092415 | 0.07865359 | 0.51314366 | 0.02303667 | 0.53154412 | 0.16324148 | 0.4947432 | 0.13625968 | 0.69955033 | 0.1072036 | 0.3037624 | 0.02170343 | 1147 | 331 | 816 |
| infectious_parasitic_disease_diagnosis | phenobag | 0.59830046 | 0.01722549 | 0.59739536 | 0.01721343 | 0.47810398 | 0.027041 | 0.71668675 | 0.02886753 | 0.62488161 | 0.02423701 | 0.58238994 | 0.01403214 | 659 | 332 | 327 |
| neoplasms_diagnosis | phenobag | 0.50601818 | 0.01200001 | 0.55616793 | 0.01275583 | 0.43022107 | 0.02064533 | 0.6821148 | 0.03051387 | 0.75903733 | 0.01491263 | 0.34003974 | 0.00864748 | 1100 | 331 | 769 |
| blood_and_immune_system_diagnosis | phenobag | 0.52734359 | 0.01339576 | 0.56874327 | 0.0147189 | 0.43978261 | 0.01875538 | 0.69770393 | 0.02865079 | 0.73923437 | 0.01796192 | 0.39027016 | 0.0106735 | 975 | 331 | 644 |
| endocrine_nutritional_metabolic_disease_diagnosis | phenobag | 0.53700555 | 0.0122525 | 0.58783785 | 0.01315316 | 0.45688415 | 0.01755959 | 0.71879154 | 0.02593753 | 0.78682328 | 0.01472142 | 0.36842894 | 0.00932883 | 1082 | 331 | 751 |
| mental_behavioural_disorder_diagnosis | phenobag | 0.59852601 | 0.01620447 | 0.60339759 | 0.01633374 | 0.483 | 0.02454319 | 0.72379518 | 0.02910922 | 0.65531138 | 0.02396746 | 0.56356573 | 0.01313904 | 692 | 332 | 360 |
| nerve_system_diagnosis | phenobag | 0.57357049 | 0.01403354 | 0.58874622 | 0.01343435 | 0.47 | 0.02589466 | 0.70749245 | 0.02277803 | 0.67519009 | 0.01640744 | 0.50812253 | 0.01176458 | 759 | 331 | 428 |

| | | | | | | | | | | | | | | | | |
|---|---|---|---|---|---|---|---|---|---|---|---|---|---|---|---|---|
| eye_diagnosis | phenobag | 0.56218579 | 0.01391599 | 0.57417227 | 0.01401109 | 0.44882793 | 0.02066564 | 0.69951662 | 0.02317945 | 0.64428161 | 0.01884497 | 0.5116559 | 0.01130263 | 732 | 331 | 401 |
| ear_diagnosis | phenobag | 0.64654709 | 0.02098325 | 0.5854164 | 0.02410505 | 0.46035088 | 0.04466641 | 0.71048193 | 0.0260719 | 0.35351482 | 0.02843045 | 0.79324074 | 0.01393899 | 446 | 332 | 114 |
| circular_system_diagnosis | phenobag | 0.5117137 | 0.01360089 | 0.57271648 | 0.01611152 | 0.46065954 | 0.0146848 | 0.68477341 | 0.02668632 | 0.83205112 | 0.01323296 | 0.27253224 | 0.01005439 | 1453 | 331 | 1122 |
| respiratory_system_diagnosis | phenobag | 0.58707291 | 0.01276239 | 0.62102138 | 0.01262292 | 0.49962585 | 0.01922962 | 0.74241692 | 0.02231638 | 0.77524389 | 0.01498592 | 0.45519762 | 0.01038713 | 919 | 331 | 588 |
| digestive_system_diagnosis | phenobag | 0.49614124 | 0.01344283 | 0.56542622 | 0.01430936 | 0.44650199 | 0.01671545 | 0.68435045 | 0.02618341 | 0.84288381 | 0.01124374 | 0.2459491 | 0.00830764 | 1586 | 331 | 1255 |
| skin_system_diagnosis | phenobag | 0.55199457 | 0.01610051 | 0.56781927 | 0.01624799 | 0.41231527 | 0.02666755 | 0.72332326 | 0.03027978 | 0.64689997 | 0.02442305 | 0.5008955 | 0.01240492 | 737 | 331 | 406 |
| musculoskeletal_system_diagnosis | phenobag | 0.5123869 | 0.01266973 | 0.57472541 | 0.0132243 | 0.44582545 | 0.01745178 | 0.70362538 | 0.02573863 | 0.81221763 | 0.01303142 | 0.30650898 | 0.00851834 | 1282 | 331 | 951 |
| genitourinary_system_diagnosis | phenobag | 0.51954664 | 0.0122841 | 0.57523903 | 0.01186998 | 0.44352941 | 0.01825912 | 0.70694864 | 0.02309726 | 0.78873872 | 0.0124411 | 0.34012467 | 0.00830546 | 1147 | 331 | 816 |
| infectious_parasitic_disease_diagnosis | phenobag_protbag_phenobagprs_protbagprs | 0.59141123 | 0.0195361 | 0.59072519 | 0.01951882 | 0.50030581 | 0.02858759 | 0.68114458 | 0.03142026 | 0.60759263 | 0.02539442 | 0.58057204 | 0.01635487 | 659 | 332 | 327 |
| neoplasms_diagnosis | phenobag_protbag_phenobagprs_protbagprs | 0.54210909 | 0.0154217 | 0.55246968 | 0.01541946 | 0.52644993 | 0.02405755 | 0.57848943 | 0.03208472 | 0.74381583 | 0.01350864 | 0.34466516 | 0.01279349 | 1100 | 331 | 769 |
| blood_and_immune_system_diagnosis | phenobag_protbag_phenobagprs_protbagprs | 0.53876923 | 0.01388196 | 0.55889109 | 0.01467162 | 0.49621118 | 0.01999446 | 0.621571 | 0.02794914 | 0.71855225 | 0.01491312 | 0.38807248 | 0.01204672 | 975 | 331 | 644 |
| endocrine_nutritional_metabolic_disease_diagnosis | phenobag_protbag_phenobagprs_protbagprs | 0.55774492 | 0.01306135 | 0.58866985 | 0.01236008 | 0.50900133 | 0.02115119 | 0.66833837 | 0.02590483 | 0.77703218 | 0.01212055 | 0.3750644 | 0.00991224 | 1082 | 331 | 751 |
| mental_behavioural_disorder_diagnosis | phenobag_protbag_phenobagprs_protbagprs | 0.59254335 | 0.01502334 | 0.59631225 | 0.0149699 | 0.50316667 | 0.02429408 | 0.68945783 | 0.0232059 | 0.63744741 | 0.01880198 | 0.56146769 | 0.01302635 | 692 | 332 | 360 |
| nerve_system_diagnosis | phenobag_protbag_phenobagprs_protbagprs | 0.57172596 | 0.01207063 | 0.5806403 | 0.01280533 | 0.51088785 | 0.02038842 | 0.65039275 | 0.02858981 | 0.65431488 | 0.01575324 | 0.50696273 | 0.01096427 | 759 | 331 | 428 |
| eye_diagnosis | phenobag_protbag_phenobagprs_protbagprs | 0.5589071 | 0.01347209 | 0.56679193 | 0.01375279 | 0.48433915 | 0.02699203 | 0.64924471 | 0.03192764 | 0.6263208 | 0.01765695 | 0.50966147 | 0.01173153 | 732 | 331 | 401 |
| ear_diagnosis | phenobag_protbag_phenobagprs_protbagprs | 0.60331839 | 0.02441424 | 0.57337191 | 0.02402974 | 0.51210526 | 0.04500439 | 0.63463855 | 0.03355317 | 0.32541927 | 0.02415714 | 0.79121319 | 0.01523908 | 446 | 332 | 114 |
| circular_system_diagnosis | phenobag_protbag_phenobagprs_protbagprs | 0.54635926 | 0.01211358 | 0.5776207 | 0.01479388 | 0.52019608 | 0.01499136 | 0.63504532 | 0.02864162 | 0.82861312 | 0.01106923 | 0.28081492 | 0.01004741 | 1453 | 331 | 1122 |
| respiratory_system_diagnosis | phenobag_protbag_phenobagprs_protbagprs | 0.58559304 | 0.01333665 | 0.61287965 | 0.01349509 | 0.51530612 | 0.0199489 | 0.71045317 | 0.02465266 | 0.75990483 | 0.01560012 | 0.45217671 | 0.01119783 | 919 | 331 | 588 |
| digestive_system_diagnosis | phenobag_protbag_phenobagprs_protbagprs | 0.5369483 | 0.01212957 | 0.55946982 | 0.01322098 | 0.52081275 | 0.01569943 | 0.59812689 | 0.02596373 | 0.83094105 | 0.00890798 | 0.24769834 | 0.00870698 | 1586 | 331 | 1255 |
| skin_system_diagnosis | phenobag_protbag_phenobagprs_protbagprs | 0.55826323 | 0.01823122 | 0.56643787 | 0.01889558 | 0.48610837 | 0.0200332 | 0.64676737 | 0.03083903 | 0.62835302 | 0.02287787 | 0.50633041 | 0.01595589 | 737 | 331 | 406 |
| musculoskeletal_system_diagnosis | phenobag_protbag_phenobagprs_protbagprs | 0.52425897 | 0.01344443 | 0.55919052 | 0.01379779 | 0.48696109 | 0.01832319 | 0.63141994 | 0.02617459 | 0.79154203 | 0.01162144 | 0.29994262 | 0.00987989 | 1282 | 331 | 951 |
| genitourinary_system_diagnosis | phenobag_protbag_phenobagprs_protbagprs | 0.53375763 | 0.01259147 | 0.56222439 | 0.01547264 | 0.49490196 | 0.0165056 | 0.62954683 | 0.03136042 | 0.76730693 | 0.01450189 | 0.33574307 | 0.01150159 | 1147 | 331 | 816 |

#### b) Cross-organ feature classification results

| Disease category | feature | CV_accuracy | CV_accuracy_std | CV_balanced_accuracy | CV_balanced_accuracy_std | CV_sensitivity | CV_sensitivity_std | CV_specificity | CV_specificity_std | CV_ppv | CV_ppv_std | CV_npv | CV_npv_std | CV_test_N | CV_test_N_CN | CV_test_N_PT |
|---|---|---|---|---|---|---|---|---|---|---|---|---|---|---|---|---|
| circular_system_diagnosis | cardiovascular | 0.41158981 | 0.01565521 | 0.52394513 | 0.0112913 | 0.31755793 | 0.02853144 | 0.73033233 | 0.03881364 | 0.80034767 | 0.01405295 | 0.23990193 | 0.00567733 | 1453 | 331 | 1122 |
| circular_system_diagnosis | metabolic | 0.41777013 | 0.0513671 | 0.52794691 | 0.01232049 | 0.3255615 | 0.09568625 | 0.73033233 | 0.10305724 | 0.80539095 | 0.01539745 | 0.24314612 | 0.01006181 | 1453 | 331 | 1122 |
| circular_system_diagnosis | immune | 0.42810736 | 0.01074418 | 0.52790989 | 0.01006983 | 0.34458111 | 0.01926327 | 0.71123867 | 0.03003891 | 0.80213928 | 0.01149056 | 0.24245465 | 0.00518177 | 1453 | 331 | 1122 |
| circular_system_diagnosis | brain | 0.43141087 | 0.01158524 | 0.52459643 | 0.01309109 | 0.35342246 | 0.02087581 | 0.69577039 | 0.03697744 | 0.79799411 | 0.01387425 | 0.24086161 | 0.00696616 | 1453 | 331 | 1122 |
| circular_system_diagnosis | eye | 0.42801101 | 0.02213777 | 0.52311916 | 0.01130826 | 0.34841355 | 0.03855525 | 0.69782477 | 0.04257476 | 0.79658604 | 0.01225896 | 0.24010951 | 0.00599254 | 1453 | 331 | 1122 |
| circular_system_diagnosis | pulmonary | 0.47719202 | 0.0125037 | 0.55485759 | 0.01435651 | 0.41219251 | 0.01818389 | 0.69752266 | 0.03208704 | 0.82228709 | 0.01369951 | 0.25926836 | 0.00810396 | 1453 | 331 | 1122 |
| circular_system_diagnosis | musculoskeletal | 0.49179628 | 0.01148529 | 0.55164111 | 0.01137845 | 0.44171123 | 0.0190467 | 0.661571 | 0.03001037 | 0.81586442 | 0.01043608 | 0.25900268 | 0.00665234 | 1453 | 331 | 1122 |
| circular_system_diagnosis | hepatic | 0.49654508 | 0.01342059 | 0.55288428 | 0.0146422 | 0.44939394 | 0.02039587 | 0.65637462 | 0.03426644 | 0.81619054 | 0.01274701 | 0.26014563 | 0.00886097 | 1453 | 331 | 1122 |
| circular_system_diagnosis | renal | 0.53277357 | 0.01229074 | 0.57468108 | 0.01646767 | 0.49770053 | 0.01583948 | 0.65166163 | 0.03421988 | 0.82906882 | 0.01343039 | 0.27674215 | 0.01038306 | 1453 | 331 | 1122 |

| | | | | | | | | | | | | | | | | |
|---|---|---|---|---|---|---|---|---|---|---|---|---|---|---|---|---|
| circular_system_diagnosis | reproductive | 0.53486579 | 0.01320429 | 0.57492827 | 0.01421985 | 0.5013369 | 0.01756997 | 0.64851964 | 0.02806752 | 0.8287077 | 0.01100055 | 0.27731887 | 0.00950272 | 1453 | 331 | 1122 |
| circular_system_diagnosis | Endocrine & skin | 0.54352374 | 0.01402538 | 0.58198265 | 0.01439575 | 0.5113369 | 0.01843428 | 0.6526284 | 0.02723134 | 0.83310482 | 0.01085335 | 0.28270862 | 0.00985378 | 1453 | 331 | 1122 |

**eTable 9: The survival analysis to predict mortality**

a) Results using the full sample sizes of each biomarker. Bold text denotes significant signals after Bonferroni correction (0.05/11).

| BAG/PRS | BAG type | hazard_ratio | CI_lower_bound | CI_upper_bound | p_value | n_case | n_noncase |
|---|---|---|---|---|---|---|---|
| **Cardiovascular_age_gap** | PhenoBAG | 1.06919514 | 1.05101726 | 1.08768742 | **2.05E-14** | 10740 | 131839 |
| **Pulmonary_age_gap** | PhenoBAG | 1.26681616 | 1.24707274 | 1.28687215 | **2.11E-191** | 10740 | 131839 |
| **Musculoskeletal_age_gap** | PhenoBAG | 1.15590917 | 1.13643796 | 1.17571399 | **1.01E-62** | 10740 | 131839 |
| **Immune_age_gap** | PhenoBAG | 1.07771894 | 1.07115129 | 1.08432685 | **2.86E-127** | 10740 | 131839 |
| **Renal_age_gap** | PhenoBAG | 1.22354322 | 1.20936986 | 1.23788269 | **1.85E-252** | 10740 | 131839 |
| **Hepatic_age_gap** | PhenoBAG | 1.17054898 | 1.15156903 | 1.18984176 | **1.66E-79** | 10740 | 131839 |
| **Metabolic_age_gap** | PhenoBAG | 1.12461943 | 1.10574294 | 1.14381817 | **4.08E-42** | 10740 | 131839 |
| **Brain_age_gap** | PhenoBAG | 1.21319889 | 1.13592842 | 1.2957256 | **8.63E-09** | 634 | 35344 |
| **Eye_age_gap** | PhenoBAG | 1.14540864 | 1.10315391 | 1.18928188 | **1.45E-12** | 2657 | 39300 |
| Brain_age_gap_PRS | PhenoBAG-PRS | 1.0029372 | 0.98616395 | 1.01999573 | 0.73322823 | 13057 | 180445 |
| **Cardiovascular_age_gap_PRS** | PhenoBAG-PRS | 1.03785886 | 1.02053585 | 1.05547592 | **1.51E-05** | 13057 | 180445 |
| Eye_age_gap_PRS | PhenoBAG-PRS | 1.0263453 | 1.00843518 | 1.04457352 | 0.00378986 | 13057 | 180445 |
| Hepatic_age_gap_PRS | PhenoBAG-PRS | 1.01341559 | 0.99623604 | 1.0308914 | 0.12659482 | 13057 | 180445 |
| **Immune_age_gap_PRS** | PhenoBAG-PRS | 1.04981868 | 1.03213439 | 1.06780597 | **2.04E-08** | 13057 | 180445 |
| Metabolic_age_gap_PRS | PhenoBAG-PRS | 1.01862164 | 1.00132319 | 1.03621893 | 0.03474858 | 13057 | 180445 |
| **Musculoskeletal_age_gap_PRS** | PhenoBAG-PRS | 1.02416158 | 1.00685575 | 1.04176486 | **0.00603744** | 13057 | 180445 |
| **Pulmonary_age_gap_PRS** | PhenoBAG-PRS | 1.07412169 | 1.05606514 | 1.09248696 | **1.38E-16** | 13057 | 180445 |
| **Renal_age_gap_PRS** | PhenoBAG-PRS | 1.0464256 | 1.02858147 | 1.06457929 | **2.33E-07** | 13057 | 180445 |
| **Reproductive_female** | ProtBAG | 1.32710297 | 1.2913665 | 1.36382839 | **8.66E-92** | 4906 | 38590 |
| **Pulmonary** | ProtBAG | 1.43615007 | 1.39841779 | 1.47490044 | **1.98E-156** | 4906 | 38590 |
| **Heart** | ProtBAG | 1.31202607 | 1.28370895 | 1.34096783 | **1.94E-131** | 4906 | 38590 |
| **Brain** | ProtBAG | 1.58763881 | 1.53757332 | 1.6393345 | **7.09E-176** | 4906 | 38590 |
| Eye | ProtBAG | 1.01629152 | 0.98643121 | 1.04705574 | 0.28819617 | 4906 | 38590 |
| **Hepatic** | ProtBAG | 1.27079152 | 1.23544469 | 1.30714965 | **3.01E-62** | 4906 | 38590 |
| Renal | ProtBAG | 0.98859361 | 0.95963809 | 1.01842282 | 0.4494297 | 4906 | 38590 |
| **Reproductive_male** | ProtBAG | 1.38186743 | 1.34734322 | 1.41727629 | **1.54E-138** | 4906 | 38590 |
| **Endocrine** | ProtBAG | 1.36767162 | 1.32900362 | 1.40746468 | **1.41E-101** | 4906 | 38590 |
| **Immune** | ProtBAG | 1.44013458 | 1.40471538 | 1.47644686 | **3.07E-181** | 4906 | 38590 |
| **Skin** | ProtBAG | 1.14334749 | 1.11109234 | 1.17653901 | **4.52E-20** | 4906 | 38590 |
| Reproductive_female_PRS | ProtBAG-PRS | 1.02313732 | 0.99490046 | 1.05217559 | 0.10917459 | 4901 | 38544 |
| Eye_PRS | ProtBAG-PRS | 1.01298005 | 0.9847122 | 1.04205938 | 0.37180607 | 4901 | 38544 |
| Pulmonary_PRS | ProtBAG-PRS | 1.01418859 | 0.98619135 | 1.04298065 | 0.32392542 | 4901 | 38544 |
| **Heart_PRS** | ProtBAG-PRS | 1.12945326 | 1.09909852 | 1.16064632 | **1.99E-18** | 4901 | 38544 |
| Brain_PRS | ProtBAG-PRS | 1.03757101 | 1.00837779 | 1.0676094 | 0.01131183 | 4901 | 38544 |
| Hepatic_PRS | ProtBAG-PRS | 1.01602216 | 0.9881703 | 1.04465902 | 0.26235969 | 4901 | 38544 |
| Renal_PRS | ProtBAG-PRS | 0.99683265 | 0.96909892 | 1.02536007 | 0.82559041 | 4901 | 38544 |
| Reproductive_male_PRS | ProtBAG-PRS | 1.01071953 | 0.9828988 | 1.03932772 | 0.45402367 | 4901 | 38544 |
| Endocrine_PRS | ProtBAG-PRS | 1.02270445 | 0.99464329 | 1.05155729 | 0.11374478 | 4901 | 38544 |
| **Immune_PRS** | ProtBAG-PRS | 1.09266029 | 1.06236918 | 1.12381509 | **6.50E-10** | 4901 | 38544 |
| Skin_PRS | ProtBAG-PRS | 1.02949082 | 1.00077431 | 1.05903134 | 0.04405286 | 4901 | 38544 |

b) Results using the merged sample sizes of all biomarkers (excluding the brain and eye PhenoBAGs). Bold text denotes significant signals after Bonferroni correction (0.05/11).

| BAG/PRS | BAG type | hazard_ratio | CI_lower_bound | CI_upper_bound | p_value | n_case | n_noncase |
|---|---|---|---|---|---|---|---|

| | | | | | | | |
|---|---|---|---|---|---|---|---|
| Cardiovascular_age_gap | PhenoBAG | 1.07277932 | 1.01883961 | 1.12957473 | 0.00760626 | 1195 | 11498 |
| **Pulmonary_age_gap** | PhenoBAG | 1.26989433 | 1.21119927 | 1.33143376 | **4.34E-23** | 1195 | 11498 |
| **Musculoskeletal_age_gap** | PhenoBAG | 1.16806952 | 1.10540252 | 1.2342892 | **3.36E-08** | 1195 | 11498 |
| **Immune_age_gap** | PhenoBAG | 1.07976149 | 1.05780094 | 1.10217796 | **2.48E-13** | 1195 | 11498 |
| **Renal_age_gap** | PhenoBAG | 1.24721009 | 1.20398176 | 1.2919905 | **1.25E-34** | 1195 | 11498 |
| **Hepatic_age_gap** | PhenoBAG | 1.17203051 | 1.11206928 | 1.23522477 | **3.13E-09** | 1195 | 11498 |
| **Metabolic_age_gap** | PhenoBAG | 1.1060655 | 1.04714332 | 1.16830319 | **0.00030708** | 1195 | 11498 |
| Brain_age_gap_PRS | PhenoBAG-PRS | 1.00869374 | 0.95805838 | 1.06200528 | 0.74184195 | 1195 | 11498 |
| **Cardiovascular_age_gap_PRS** | PhenoBAG-PRS | 1.0686724 | 1.01666952 | 1.12333525 | **0.00906735** | 1195 | 11498 |
| Eye_age_gap_PRS | PhenoBAG-PRS | 1.01861077 | 0.96616942 | 1.0738985 | 0.49412089 | 1195 | 11498 |
| Hepatic_age_gap_PRS | PhenoBAG-PRS | 1.03061062 | 0.97941378 | 1.08448367 | 0.24612198 | 1195 | 11498 |
| **Immune_age_gap_PRS** | PhenoBAG-PRS | 1.08755571 | 1.03459247 | 1.14323026 | **0.00098409** | 1195 | 11498 |
| Metabolic_age_gap_PRS | PhenoBAG-PRS | 1.01914921 | 0.96824591 | 1.07272863 | 0.46809465 | 1195 | 11498 |
| Musculoskeletal_age_gap_PRS | PhenoBAG-PRS | 1.04724455 | 0.99484701 | 1.10240182 | 0.07795265 | 1195 | 11498 |
| **Pulmonary_age_gap_PRS** | PhenoBAG-PRS | 1.1066298 | 1.05229875 | 1.16376599 | **7.99E-05** | 1195 | 11498 |
| **Renal_age_gap_PRS** | PhenoBAG-PRS | 1.11306609 | 1.05684518 | 1.17227777 | **5.11E-05** | 1195 | 11498 |
| **Reproductive_female** | ProtBAG | 1.29632795 | 1.23163473 | 1.36441928 | **2.89E-23** | 1195 | 11498 |
| **Pulmonary** | ProtBAG | 1.34340551 | 1.27896653 | 1.41109117 | **5.52E-32** | 1195 | 11498 |
| **Heart** | ProtBAG | 1.2742243 | 1.22379538 | 1.32673126 | **6.10E-32** | 1195 | 11498 |
| **Brain** | ProtBAG | 1.46268019 | 1.37856407 | 1.55192884 | **2.59E-36** | 1195 | 11498 |
| Eye | ProtBAG | 0.98907416 | 0.93565838 | 1.04553939 | 0.69813902 | 1195 | 11498 |
| **Hepatic** | ProtBAG | 1.21941429 | 1.15834826 | 1.28369961 | **3.80E-14** | 1195 | 11498 |
| Renal | ProtBAG | 0.9703115 | 0.91850055 | 1.025045 | 0.2817274 | 1195 | 11498 |
| **Reproductive_male** | ProtBAG | 1.33006475 | 1.26845064 | 1.39467173 | **4.60E-32** | 1195 | 11498 |
| **Endocrine** | ProtBAG | 1.30754354 | 1.24016455 | 1.37858328 | **2.96E-23** | 1195 | 11498 |
| **Immune** | ProtBAG | 1.35434554 | 1.29268142 | 1.41895119 | **2.83E-37** | 1195 | 11498 |
| **Skin** | ProtBAG | 1.10179765 | 1.0461715 | 1.16038151 | **0.0002448** | 1195 | 11498 |
| Reproductive_female_PRS | ProtBAG-PRS | 0.98495066 | 0.93545767 | 1.03706221 | 0.5642964 | 1195 | 11498 |
| Eye_PRS | ProtBAG-PRS | 1.00831664 | 0.957257 | 1.06209977 | 0.75475351 | 1195 | 11498 |
| Pulmonary_PRS | ProtBAG-PRS | 1.00085256 | 0.95109712 | 1.05321089 | 0.97386925 | 1195 | 11498 |
| **Heart_PRS** | ProtBAG-PRS | 1.14133466 | 1.08589025 | 1.19961 | **1.96E-07** | 1195 | 11498 |
| Brain_PRS | ProtBAG-PRS | 0.99851576 | 0.94867993 | 1.05096954 | 0.95465545 | 1195 | 11498 |
| Hepatic_PRS | ProtBAG-PRS | 0.9808546 | 0.93204818 | 1.03221674 | 0.45788918 | 1195 | 11498 |
| Renal_PRS | ProtBAG-PRS | 0.97035448 | 0.92116025 | 1.02217591 | 0.25692585 | 1195 | 11498 |
| Reproductive_male_PRS | ProtBAG-PRS | 1.00497862 | 0.95502225 | 1.05754816 | 0.84859933 | 1195 | 11498 |
| Endocrine_PRS | ProtBAG-PRS | 1.00650396 | 0.95628338 | 1.05936195 | 0.80394313 | 1195 | 11498 |
| Immune_PRS | ProtBAG-PRS | 1.06097158 | 1.00788316 | 1.11685632 | 0.02383523 | 1195 | 11498 |
| Skin_PRS | ProtBAG-PRS | 1.06918157 | 1.0149746 | 1.12628358 | 0.01173966 | 1195 | 11498 |

**eTable 10: Incremental R2 contributed by the 11 ProtBAGs on top of age, sex, and conventional non-organ specific ProtBAG in Argentieri**

The additional $R^2$ contributed by the 11 multi-organ ProtBAGs (alternative model) was assessed beyond the null model, which included age, sex, and the conventional ProtBAG derived from >2000 proteins and trained on 31,808 mixed-pathology participants following Argentieri et al.'s method. The variable "$R^2$_multiorgan" quantifies the incremental variance explained by the 11 ProtBAGs in a linear regression model. The outcome variables analyzed were 8 cognitive scores (**a**) and age at death (**b**).

**a) 8 cognitive scores**

| Variable | beta_null | se_null | p_null | beta_alt | se_alt | p_alt | Cognitive_Score |
|---|---|---|---|---|---|---|---|
| Age | -0.0911967 | 0.00679673 | 4.87E-40 | -0.0904753 | 0.00684832 | 6.79E-39 | number_of_puzzles_correct_f21004_2_0 |
| Sex | 0.51065055 | 0.10471169 | 1.13E-06 | 0.5296906 | 0.11696425 | 6.14E-06 | |
| Brain | NA | NA | NA | -0.0050001 | 0.01712371 | 0.7703051 | |
| Immune | NA | NA | NA | -0.0088532 | 0.01778133 | 0.61859334 | |
| Pulmonary | NA | NA | NA | -0.0197796 | 0.02804285 | 0.48065089 | |
| Reproductive_male | NA | NA | NA | 0.02629564 | 0.02117719 | 0.21443455 | |
| Endocrine | NA | NA | NA | 0.0079545 | 0.01799685 | 0.6585217 | |
| Reproductive_female | NA | NA | NA | -0.0042906 | 0.02677558 | 0.87269921 | |
| Heart | NA | NA | NA | -0.0171268 | 0.02696831 | 0.52542362 | |
| Hepatic | NA | NA | NA | 0.01607046 | 0.02121438 | 0.44878703 | |
| Renal | NA | NA | NA | -0.0204403 | 0.04744525 | 0.66662736 | |
| Skin | NA | NA | NA | -0.0220017 | 0.02436182 | 0.36652555 | |
| Eye | NA | NA | NA | -0.0508752 | 0.05875011 | 0.38657362 | |
| N | 3407 | NA | NA | NA | NA | NA | |
| R2_null | 0.05368949 | NA | NA | NA | NA | NA | |
| R2_alternative | 0.05527311 | NA | NA | NA | NA | NA | |
| **R2_multiorgan** | 0.00158363 | NA | NA | NA | NA | NA | |
| Age | -0.2931958 | 0.01053215 | 6.36E-154 | -0.292821 | 0.01058979 | 4.90E-152 | number_of_symbol_digit_matches_made_correctly_f23324_2_0 |
| Sex | -0.1308506 | 0.1623099 | 0.42019658 | -0.0432482 | 0.18110202 | 0.81127062 | |
| Brain | NA | NA | NA | -0.0349859 | 0.02650764 | 0.18697671 | |
| Immune | NA | NA | NA | 0.02134688 | 0.02747135 | 0.43717753 | |

| | | | | | | | |
|---|---|---|---|---|---|---|---|
| Pulmonary | NA | NA | NA | -0.0589164 | 0.04332734 | 0.17398404 | |
| Reproductive_male | NA | NA | NA | -0.0153893 | 0.03283074 | 0.63928012 | |
| Endocrine | NA | NA | NA | 0.00533554 | 0.0278455 | 0.84805733 | |
| Reproductive_female | NA | NA | NA | -0.0352149 | 0.04138453 | 0.39487362 | |
| Heart | NA | NA | NA | -0.0405869 | 0.04185017 | 0.33220766 | |
| Hepatic | NA | NA | NA | 0.00143484 | 0.03274014 | 0.96504646 | |
| Renal | NA | NA | NA | -0.1039493 | 0.07338942 | 0.15674771 | |
| Skin | NA | NA | NA | -0.0810095 | 0.03776806 | 0.03202956 | |
| Eye | NA | NA | NA | 0.15537256 | 0.09083767 | 0.08727504 | |
| N | 3428 | NA | NA | NA | NA | NA | |
| R2_null | 0.18737347 | NA | NA | NA | NA | NA | |
| R2_alternative | 0.19197526 | NA | NA | NA | NA | NA | |
| **R2_multiorgan** | 0.00460179 | NA | NA | NA | NA | NA | |
| Age | -0.06462 | 0.00444723 | 1.82E-46 | -0.0650164 | 0.00447309 | 1.73E-46 | |
| Sex | 0.37389704 | 0.06848131 | 5.10E-08 | 0.39877311 | 0.07635656 | 1.87E-07 | |
| Brain | NA | NA | NA | 0.00304577 | 0.01117986 | 0.78530527 | |
| Immune | NA | NA | NA | -0.0113274 | 0.01160099 | 0.3289283 | |
| Pulmonary | NA | NA | NA | -0.0197446 | 0.01831777 | 0.28115701 | |
| Reproductive_male | NA | NA | NA | -0.0282486 | 0.0138528 | 0.04150732 | |
| Endocrine | NA | NA | NA | -0.0123759 | 0.01176564 | 0.29293436 | number_of_puzzles_correctly_solved_f6373_2_0 |
| Reproductive_female | NA | NA | NA | -0.0018002 | 0.01748024 | 0.91798051 | |
| Heart | NA | NA | NA | -0.0116951 | 0.01758481 | 0.50605107 | |
| Hepatic | NA | NA | NA | 0.00425404 | 0.01386085 | 0.75893046 | |
| Renal | NA | NA | NA | -0.03696 | 0.03093961 | 0.23233245 | |
| Skin | NA | NA | NA | -0.0185444 | 0.01590532 | 0.24372553 | |
| Eye | NA | NA | NA | 0.05014101 | 0.03838951 | 0.1916023 | |
| N | 3430 | NA | NA | NA | NA | NA | |
| R2_null | 0.06273198 | NA | NA | NA | NA | NA | |

| | | | | | | | |
|---|---|---|---|---|---|---|---|
| R2_alternative | 0.06776863 | NA | NA | NA | NA | NA | |
| **R2_multiorgan** | 0.00503665 | NA | NA | NA | NA | NA | |
| Age | 3.36027699 | 0.18217197 | 1.55E-72 | 3.40287073 | 0.18326236 | 2.00E-73 | duration_to_complete_numeric_path_trail_1_f6348_2_0 |
| Sex | 6.50882722 | 2.81181278 | 0.02068143 | 5.69881456 | 3.14043377 | 0.06966346 | |
| Brain | NA | NA | NA | 0.17331 | 0.45979936 | 0.70625238 | |
| Immune | NA | NA | NA | 0.32105184 | 0.47656715 | 0.50056258 | |
| Pulmonary | NA | NA | NA | 1.40654624 | 0.7513651 | 0.06129306 | |
| Reproductive_male | NA | NA | NA | 0.57779488 | 0.56897519 | 0.30993846 | |
| Endocrine | NA | NA | NA | -0.0439398 | 0.48339491 | 0.92757872 | |
| Reproductive_female | NA | NA | NA | -0.5751386 | 0.71879843 | 0.42368555 | |
| Heart | NA | NA | NA | 1.15799847 | 0.72396428 | 0.10979595 | |
| Hepatic | NA | NA | NA | 0.76318041 | 0.56877345 | 0.17974928 | |
| Renal | NA | NA | NA | 0.27255463 | 1.27101378 | 0.83021761 | |
| Skin | NA | NA | NA | 0.36189766 | 0.65499993 | 0.58063106 | |
| Eye | NA | NA | NA | -1.9130928 | 1.57750421 | 0.22531483 | |
| N | 3456 | NA | NA | NA | NA | NA | |
| R2_null | 0.0940121 | NA | NA | NA | NA | NA | |
| R2_alternative | 0.09837211 | NA | NA | NA | NA | NA | |
| **R2_multiorgan** | 0.00436001 | NA | NA | NA | NA | NA | |
| Age | 8.48109361 | 0.5851644 | 2.96E-46 | 8.53818869 | 0.58898487 | 2.87E-46 | duration_to_complete_alphanumeric_path_trail_2_f6350_2_0 |
| Sex | 12.4682433 | 9.03197537 | 0.16753571 | 7.57427766 | 10.0930052 | 0.45303612 | |
| Brain | NA | NA | NA | -0.6144117 | 1.47774408 | 0.67759929 | |
| Immune | NA | NA | NA | 3.32174665 | 1.53163386 | 0.03016917 | |
| Pulmonary | NA | NA | NA | 3.88178419 | 2.41480394 | 0.10803767 | |
| Reproductive_male | NA | NA | NA | -0.2856077 | 1.82862306 | 0.87589455 | |
| Endocrine | NA | NA | NA | -1.7552666 | 1.55357755 | 0.25862984 | |
| Reproductive_female | NA | NA | NA | -1.1073334 | 2.31013831 | 0.63172978 | |
| Heart | NA | NA | NA | 3.36742742 | 2.32674077 | 0.14791177 | |
| Hepatic | NA | NA | NA | -0.4689024 | 1.82797467 | 0.79756876 | |
| Renal | NA | NA | NA | -0.2318491 | 4.08489707 | 0.95474156 | |
| Skin | NA | NA | NA | 0.80254643 | 2.10509701 | 0.70304897 | |

| | | | | | | | |
|---|---|---|---|---|---|---|---|
| Eye | NA | NA | NA | -2.0416735 | 5.06992326 | 0.68719172 | |
| N | 3456 | NA | NA | NA | NA | NA | |
| R2_null | 0.05934493 | NA | NA | NA | NA | NA | |
| R2_alternative | 0.06286022 | NA | NA | NA | NA | NA | |
| **R2_multiorgan** | 0.0035153 | NA | NA | NA | NA | NA | |
| Age | -0.0232020 | 0.00336162 | 5.64E-12 | -0.0239417 | 0.00337961 | 1.55E-12 | |
| Sex | 0.22983472 | 0.05161008 | 8.61E-06 | 0.26612488 | 0.05787508 | 4.35E-06 | |
| Brain | NA | NA | NA | 0.00836088 | 0.00823034 | 0.30973511 | |
| Immune | NA | NA | NA | -0.0054866 | 0.00863515 | 0.52520372 | |
| Pulmonary | NA | NA | NA | -0.0414378 | 0.01362788 | 0.00237049 | |
| Reproductive_male | NA | NA | NA | -0.0192334 | 0.0104309 | 0.065247 | |
| Endocrine | NA | NA | NA | 0.00099226 | 0.00886328 | 0.91086556 | |
| Reproductive_female | NA | NA | NA | -0.0124508 | 0.01339001 | 0.352481 | fluid_intelligence_score_f20016_2_0 |
| Heart | NA | NA | NA | -0.0058248 | 0.0133395 | 0.66237522 | |
| Hepatic | NA | NA | NA | 0.01018348 | 0.01020947 | 0.31858302 | |
| Renal | NA | NA | NA | -0.0211197 | 0.02342856 | 0.36563284 | |
| Skin | NA | NA | NA | -0.0029162 | 0.01206735 | 0.80905415 | |
| Eye | NA | NA | NA | 0.0102516 | 0.02973567 | 0.73028842 | |
| N | 6192 | NA | NA | NA | NA | NA | |
| R2_null | 0.00999421 | NA | NA | NA | NA | NA | |
| R2_alternative | 0.01369925 | NA | NA | NA | NA | NA | |
| **R2_multiorgan** | 0.00370505 | NA | NA | NA | NA | NA | |
| Age | -0.0278461 | 0.00302635 | 5.90E-20 | -0.0272791 | 0.00304598 | 5.35E-19 | |
| Sex | 0.19864365 | 0.04670686 | 2.16E-05 | 0.19656963 | 0.05211419 | 0.00016467 | |
| Brain | NA | NA | NA | 0.00332494 | 0.00762637 | 0.66287779 | |
| Immune | NA | NA | NA | -0.0083902 | 0.0078425 | 0.28476887 | maximum_digits_remembered_correctly_f4282_2_0 |
| Pulmonary | NA | NA | NA | -0.0011067 | 0.01239872 | 0.92888001 | |
| Reproductive_male | NA | NA | NA | -0.0060628 | 0.00942528 | 0.52010716 | |
| Endocrine | NA | NA | NA | -0.0098308 | 0.00804343 | 0.22170594 | |

| | | | | | | | |
|---|---|---|---|---|---|---|---|
| Reproductive_female | NA | NA | NA | -0.0053207 | 0.01193707 | 0.65581875 | |
| Heart | NA | NA | NA | 0.02597338 | 0.01202726 | 0.0308753 | |
| Hepatic | NA | NA | NA | -0.0012003 | 0.00945883 | 0.89902512 | |
| Renal | NA | NA | NA | -0.0196356 | 0.02111546 | 0.35247843 | |
| Skin | NA | NA | NA | -0.0161573 | 0.01089088 | 0.13801392 | |
| Eye | NA | NA | NA | -0.0054154 | 0.02628755 | 0.83679823 | |
| N | 3540 | NA | NA | NA | NA | NA | |
| R2_null | 0.02671196 | NA | NA | NA | NA | NA | |
| R2_alternative | 0.03018737 | NA | NA | NA | NA | NA | |
| **R2_multiorgan** | 0.00347541 | NA | NA | NA | NA | NA | |
| Age | 4.15438754 | 0.18575471 | 3.53E-106 | 4.14423088 | 0.18694989 | 2.05E-104 | |
| Sex | -19.511453 | 2.89129395 | 1.65E-11 | -20.161687 | 3.24760147 | 5.75E-10 | |
| Brain | NA | NA | NA | 0.24027347 | 0.46630381 | 0.60638315 | |
| Immune | NA | NA | NA | 0.34223916 | 0.48335583 | 0.47894503 | |
| Pulmonary | NA | NA | NA | -0.1158568 | 0.76841434 | 0.88015961 | |
| Reproductive_male | NA | NA | NA | -0.5894184 | 0.58714559 | 0.31548442 | |
| Endocrine | NA | NA | NA | -0.7837228 | 0.50007754 | 0.11712411 | |
| Reproductive_female | NA | NA | NA | 1.39262524 | 0.75253534 | 0.06428319 | mean_time_to_correctly_identify_matches_f20023_2_0 |
| Heart | NA | NA | NA | 0.52116467 | 0.74424809 | 0.48379628 | |
| Hepatic | NA | NA | NA | -0.3193119 | 0.58118764 | 0.5827442 | |
| Renal | NA | NA | NA | 0.1572647 | 1.31457968 | 0.90477964 | |
| Skin | NA | NA | NA | 0.87771236 | 0.67575537 | 0.19404534 | |
| Eye | NA | NA | NA | -2.7868149 | 1.66091096 | 0.09342594 | |
| N | 5588 | NA | NA | NA | NA | NA | |
| R2_null | 0.08532296 | NA | NA | NA | NA | NA | |
| R2_alternative | 0.08747692 | NA | NA | NA | NA | NA | |
| **R2_multiorgan** | 0.00215396 | NA | NA | NA | NA | NA | |

#### b) Age at death

| Variable | beta_null | se_null | p_null | beta_alt | se_alt | p_alt |
|---|---|---|---|---|---|---|
| Age | 1.01381532 | 0.00869493 | 0 | 1.01064054 | 0.0087474 | 0 |

| | | | | | | |
|---|---|---|---|---|---|---|
| Sex | -0.4400343 | 0.1087178 | 5.26E-05 | -0.5513294 | 0.1264809 | 1.33E-05 |
| Brain | NA | NA | NA | -0.0294661 | 0.01949322 | 0.13069873 |
| Immune | NA | NA | NA | -0.0790833 | 0.01624301 | 1.16E-06 |
| Pulmonary | NA | NA | NA | -0.0319725 | 0.02477314 | 0.19689982 |
| Reproductive_male | NA | NA | NA | -0.0508557 | 0.01945539 | 0.00897721 |
| Endocrine | NA | NA | NA | -0.0009768 | 0.01713276 | 0.95453668 |
| Reproductive_female | NA | NA | NA | -0.0498435 | 0.02637635 | 0.05885666 |
| Heart | NA | NA | NA | -0.0379955 | 0.01972215 | 0.05409509 |
| Hepatic | NA | NA | NA | -0.0017735 | 0.02013137 | 0.92980347 |
| Renal | NA | NA | NA | 0.16888493 | 0.04904536 | 0.00057918 |
| Skin | NA | NA | NA | 0.017905 | 0.02545725 | 0.48187985 |
| Eye | NA | NA | NA | 0.0059692 | 0.06545434 | 0.92734029 |
| N | 4906 | NA | NA | NA | NA | NA |
| R2_null | 0.73874165 | NA | NA | NA | NA | NA |
| R2_alternative | 0.74512148 | NA | NA | NA | NA | NA |
| **R2_multiorgan** | 0.00637983 | NA | NA | NA | NA | NA |

**eTable 11: The statistics of the linear regression between the brain ProtBAG with/without age bias correction and age at death and DSST**

| Variable | Bias correction | Beta±SE | Sample size | -log10(P) |
|---|---|---|---|---|
| Age at death | Yes | -0.082±0.012 | 4906 | 10.68 |
| | No | -0.083±0.012 | 4906 | 10.48 |
| DSST | Yes | -0.026±0.011 | 3428 | 1.61 |
| | No | -0.012±0.013 | 3428 | 0.43 |
